# Impaired glucose tolerance and cardiovascular risk factors in relation to infertility: a Mendelian randomization analysis in the Norwegian Mother, Father and Child Cohort Study

**DOI:** 10.1101/2023.01.26.23285048

**Authors:** Álvaro Hernáez, Yunsung Lee, Christian M. Page, Karoline H. Skåra, Siri E. Håberg, Per Magnus, Pål R. Njølstad, Ole A. Andreassen, Elizabeth C. Corfield, Alexandra Havdahl, Abigail Fraser, Stephen Burgess, Deborah A. Lawlor, Maria C. Magnus

**Affiliations:** Centre for Fertility and Health, Norwegian Institute of Public Health, Oslo, Norway; Blanquerna School of Health Sciences, Universitat Ramon Llull, Barcelona, Spain; Department of Physical Health and Ageing, Division for Mental and Physical Health, Norwegian Institute of Public Health, Oslo, Norway; Department of Community Medicine and Global Health, Institute of Health and Society, University of Oslo, Oslo, Norway; Mohn Center for Diabetes Precision Medicine, Department of Clinical Science, University of Bergen, Bergen, Norway; Children and Youth Clinic, Haukeland University Hospital, Bergen, Norway; Norwegian Centre for Mental Disorders Research, NORMENT, Division of Mental Health and Addiction, Oslo University Hospital, Oslo, Norway; Institute of Clinical Medicine, University of Oslo, Oslo, Norway; Department of Mental Disorders, Norwegian Institute of Public Health, Oslo, Norway; Nic Waals Institute, Lovisenberg Diakonale Hospital, Oslo, Norway; PROMENTA Research Center, Department of Psychology, University of Oslo, Oslo, Norway; Population Health Sciences, Bristol Medical School, University of Bristol, Bristol, UK; MRC Integrative Epidemiology Unit at the University of Bristol, Bristol, UK; MRC Biostatistics Unit, University of Cambridge, Cambridge, UK; Cardiovascular Epidemiology Unit, Department of Public Health and Primary Care, University of Cambridge, Cambridge, UK; NIHR Bristol Biomedical Research Centre, Bristol, UK

**Keywords:** Fasting insulin, glucose metabolism, blood pressure, lipid profile, infertility, Mendelian randomization, MoBa

## Abstract

**Aims/hypothesis:** Observational evidence suggests a shared etiology between impaired glucose tolerance, cardiovascular risk, and fertility problems. We aim to establish whether impaired glucose tolerance (as measured by fasting glucose, glycated hemoglobin, and fasting insulin) and cardiovascular disease risk (as measured by LDL cholesterol, HDL cholesterol, triglycerides, systolic blood pressure, and diastolic blood pressure) are causally related to infertility (having tried to conceive for ≥ 12 months or used assisted reproduction technologies to conceive) by Mendelian randomization (MR) analyses in women and men.

**Methods:** We conducted two-sample MR analyses, in which we used genome-wide association summary data that were publicly available for the cardiometabolic risk factors, and sex specific genome-wide association studies of infertility conducted in the Norwegian Mother, Father, and Child Cohort Study (68,882 women [average age 30, involved in 81,682 pregnancies] and 47,474 of their male partners [average age 33, 55,744 pregnancies]). We applied the inverse variance weighted method with random effects to pool data across variants and a series of sensitivity analyses to explore genetic instrument validity (we checked the robustness of genetic instruments and the lack of unbalanced horizontal pleiotropy, and we used methods that are robust to population stratification). Findings were corrected for multiple comparisons by the Bonferroni method (8 exposures: *p*-value < 0.00625).

**Results:** In women, increases in genetically determined fasting insulin levels were associated with greater odds of infertility (+1 log(pmol/L): OR 1.60, 95% CI 1.17 to 2.18, *p*-value = 0.003). The results were robust in the sensitivity analyses exploring the validity of MR assumptions and the role of pleiotropy of other cardiometabolic risk factors. There was also evidence of higher glucose and glycated hemoglobin in women (and possibly higher fasting insulin in men) causing infertility, but findings were imprecise and did not pass our *p*-value threshold for multiple testing. Results for lipids and blood pressure were close to the null, suggesting that these did not cause infertility.

**Conclusions/interpretation:** Genetic instruments suggest that higher fasting insulin may increase infertility in women.

**TWEET:** Mendelian randomization suggests that higher fasting #insulin levels increase the risk of #infertility in women. @alvaro_hernaez @theCEFH @Folkehelseinst @ERC_Research #MendelianRandomization

**RESEARCH IN CONTEXT:** *What is already known about this subject?:*

- Observational evidence suggests impaired glucose tolerance, higher lipid concentrations, and higher blood pressure is associated with fertility problems.

*What is the key question?:*

- Are fasting glucose, glycated hemoglobin, fasting insulin, LDL cholesterol, HDL cholesterol, triglycerides, systolic blood pressure, and diastolic blood pressure causally related to infertility in women and men?

*What are the new findings?:*

- Two-sample Mendelian randomization suggests that higher fasting insulin levels increase the risk of infertility in women. There was also evidence of higher glucose and glycated hemoglobin in women (and maybe higher fasting insulin in men) causing infertility, but results were imprecise and did not pass our *p*-value threshold for multiple comparisons.
- Findings for lipids and blood pressure were close to the null, suggesting that these did not have a causal role on infertility.

*How might this impact on clinical practice in the foreseeable future?:*

- Treatments to lower fasting insulin levels may reduce the risk of infertility in women.

## INTRODUCTION

Impaired glucose tolerance and other cardiovascular risk factors are related to fertility problems in women [1, 2] and men [3]. However, these associations may be explained by unmeasured or residual confounding (e.g., adiposity) [4]. Mendelian randomization (MR), a method based on genetic instrumental variables, estimates potential causal effects that are unaffected by confounding [4]. Using this methodology, we have previously described that high and low body weight increase the risk of infertility in both sexes [5], while the previously reported associated between smoking and infertility may not be causal [6]. The aim of this study was to investigate whether biomarkers of impaired glucose tolerance (fasting glucose, glycated hemoglobin, fasting insulin) and cardiovascular disease risk factors (LDL cholesterol [LDL-C], HDL cholesterol [HDL-C], triglycerides, systolic and diastolic blood pressure [SBP, DBP]) influence the risk of infertility using a two-sample MR-based approach.

## METHODS

### Population

Our study involved data from the Norwegian Mother, Father and Child Cohort Study (MoBa) [7]. MoBa is a population-based pregnancy cohort study administered by the Norwegian Institute of Public Health. Pregnant women and their partners were recruited at ∼18 gestational weeks across Norway between 1999-2008. The cohort is composed by 114,500 children, 95,200 mothers and 75,200 fathers. Blood samples were obtained from both parents during pregnancy [8]. Details of the MoBa genotyping data and quality control have been previously described [9]. This work is based on a subsample of parents involved in singleton pregnancies with available genotype and infertility information (68,882 women and 47,474 men) from version 12 of the quality-assured data files released in May 11, 2022. Our study followed the STROBE checklists for MR and cohort studies.

### Genetic variants and instrumental variables

We extracted independent SNPs as genetic instruments from the most recent genome-wide association studies (GWASs) on risk factors of impaired glucose metabolism, lipid profile, and blood pressure (see “Data Availability”). Independent SNPs were not in linkage disequilibrium (pairwise *r*^2^ <0.01) and were associated with their phenotypes according to genome-wide significance thresholds. MoBa was not part in any of these GWASs. We used SNPs individually as genetic instruments [10]. SNPs that were used in the analyses are described in **Supplemental Tables 1-8.** The number of SNPs used in each approach is described in **Supplemental Table 9**.

**Table 1.** Population characteristics

|  | Women |  |  | Men |  |  |
| --- | --- | --- | --- | --- | --- | --- |
|  | All<br>(*n* = 68,882) | No infertility<br>reported<br>(*n* = 60,532) | Infertility<br>reported<br>(*n* = 8,350) | All<br>(*n* = 47,474) | No infertility<br>reported<br>(*n* = 41,624) | Infertility<br>reported<br>(*n* = 5,850) |
| Age at delivery,<br>years (mean ± SD) | 30.5 ± 4.61 | 30.3 ± 4.58 | 32.1 ± 4.47 | 32.9 ± 5.27 | 32.6 ± 5.21 | 34.6 ± 5.38 |
| Education years<br>(mean ± SD) | 17.1 ± 3.35 | 17.1 ± 3.34 | 16.9 ± 3.39 | 16.4 ± 3.55 | 16.4 ± 3.54 | 16.1 ± 3.57 |
| BMI, kg/m<sup>2</sup> (median,<br>1<sup>st</sup>-3<sup>rd</sup> quartile) | 23.2<br>(21.2-26.1) | 23.1<br>(21.1-26.0) | 23.9<br>(21.5-27.4) | 25.5<br>(23.7-27.8) | 25.5<br>(23.7-27.7) | 25.9<br>(24.1-28.3) |
| Ever smokers<br>(*n*, %): | 35,632<br>(52.3%) | 31,247<br>(52.3%) | 4,385<br>(53.0%) | 24,299<br>(51.2%) | 21,232<br>(51.0%) | 3,067<br>(52.4%) |
| Trying for a first<br>pregnancy (*n*, %): | 27,030<br>(39.3%) | 23,290<br>(38.6%) | 3,740<br>(44.9%) | 20,026<br>(42.3%) | 17,212<br>(41.4%) | 2,814<br>(48.2%) |

### Infertility

Infertility was defined as trying to conceive for ≥12 months or having used assisted reproductive technologies as previously described [5, 6]. The reference group comprised participants who conceived spontaneously within 12 months. Couples with unplanned pregnancies were excluded.

### Other variables

We obtained information from both parents on the age at delivery (continuous), years of education (continuous), BMI (continuous), tobacco use (having ever smoked or not), and number of previous deliveries (continuous). We report the maximum values for all continuous variables for those parents who participated in more than one pregnancy in MoBa.

### Ethical approval

The MoBa study follows the Declaration of Helsinki for Medical Research involving human subjects. The establishment of MoBa and its initial data collection were based on a license from the Norwegian Data Protection Agency and approved by Regional Committees for Medical and Health Research Ethics. The MoBa cohort is now based on the regulations in the Norwegian Health Registry Act. Participants provided written informed consent before joining the cohort. This project was approved by the Regional Committee for Medical and Health Research Ethics of South/East Norway (reference: 2017/1362).

### Statistical analyses

We described normally distributed continuous variables by means and SDs, non-normally distributed continuous variables by medians and 1^st^-3^rd^ quartiles, and categorical variables by proportions. We assessed differences in baseline characteristics between parents with and without infertility by unpaired t-tests (normally distributed variables), Mann-Whitney U-tests (non-normally distributed variables), and chi-squared tests (categorical variables).

We first performed sex specific GWASs of infertility in MoBa using Scalable and Accurate Implementation of GEneralized mixed models (accounting for kinship and adjusted for the first 20 genetic principal components, genotype batch, and age at reported infertility). We then searched for the SNPs linked to the cardiometabolic risk factors in the summary statistics of the infertility GWASs. Palindromic SNPs with allele frequencies close to 0.5 were excluded. After data harmonization, we estimated the association between the risk factor of interest and infertility using a random effects inverse variance weighted method [10]. We corrected our findings for multiple comparisons (8 exposures) by the Bonferroni method (*p*­value threshold < 0.00625). Finally, the three key MR assumptions that must be met to obtain valid conclusions in MR analyses [10] (genetic instruments are robustly related to the exposure, there is no confounding of the genetic instrument-outcome associations, and genetic instruments are exclusively linked to the outcome through the exposure of interest) were investigated in detail as described in the **Supplemental Methods**.

Analyses were conducted in R Software v. 4.1.2. Code for data management and analysis is available in https://github.com/alvarohernaez/MR_cardiometabolic_infertility_MoBa.

## RESULTS

### Study population

Our study population consisted of 68,882 women (a total of 81,682 pregnancies) and 47,474 men (55,744 pregnancies) (**Supplemental Figure 1**). Infertility was reported in 12% of the couples. Participants with infertility were older, had lower educational attainment, greater mean BMI values, were more likely to have smoked, and were more likely to be trying for a first pregnancy (**Table 1**).

### Cardiometabolic risk factors and infertility

In women, increases in the genetically determined levels of fasting insulin were related to greater odds of infertility (+1 log(pmol/L): OR 1.60, 95% CI 1.17 to 2.18, *p*-value = 0.003)

(**Figure 1**). We found no evidence that this result was affected by weak instrument bias (estimated F-statistic: 52.5) or unbalanced horizontal pleiotropy (MR-Egger intercept test *p*-value = 0.194; no evidence of between-SNP heterogeneity [Cochran’s Q = 55.8, *p*-value = 0.445; Rücker’s Q’ = 54.1, *p*-value = 0.472]; consistency in directionality of estimates in MR sensitivity methods; consistency between main analyses and those only using the genetic variants that were unrelated to other risk factors according to the Steiger filtering) (**Supplemental Table 10**). Associations of higher maternal fasting glucose and glycated hemoglobin in women (and maybe fasting insulin in men) with greater infertility odds were suggested. However, these relationships were imprecise and did not pass our multiple testing *p*-value threshold. Lipid and blood pressure associations with infertility were close to the null (**Figure 1**, **Supplemental Table 10**).

**Figure 1.**
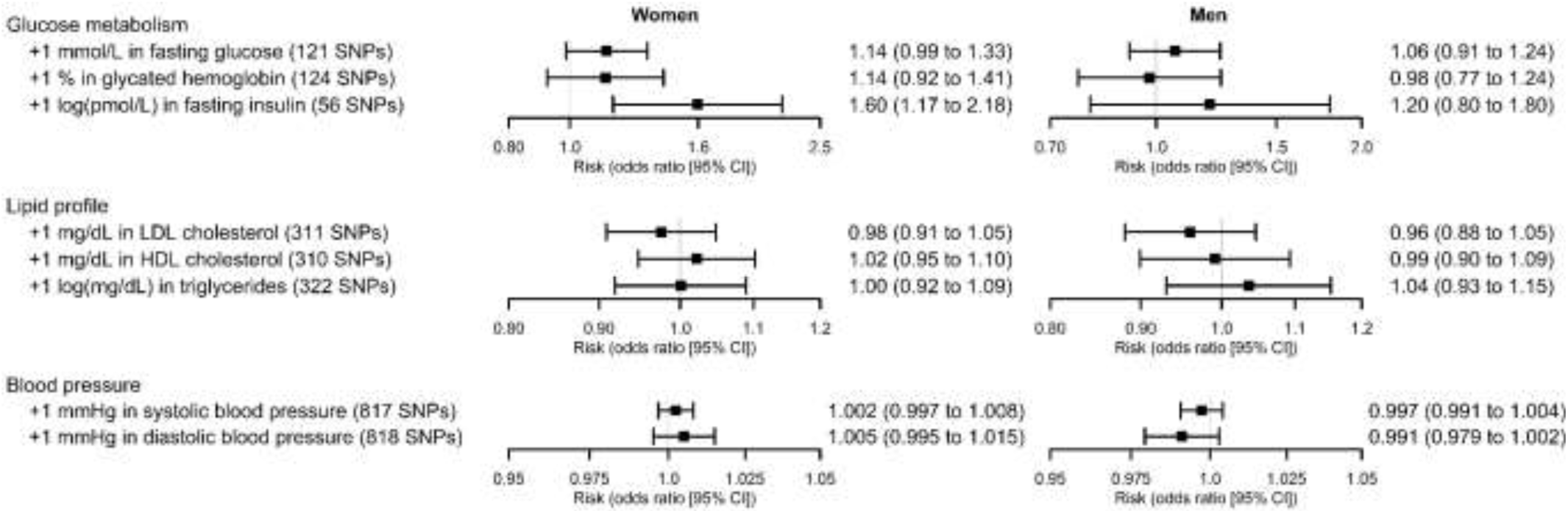
MR analyses on the relationship of cardiometabolic risk factors with infertility in women and men (units differ for each exposure, results are not directly comparable in magnitude).

## DISCUSSION

In this MR analysis, genetic instruments suggest that higher concentrations of fasting insulin increase the risk of infertility in women. An association between higher fasting glucose and glycated hemoglobin in women (and maybe higher fasting insulin in men) and higher risk of infertility is also suggested but analyses in larger numbers of participants are needed to confirm this. Hyperinsulinemia, a marker of insulin resistance, and glucose intolerance are common in women with polycystic ovary syndrome, which is a cause of infertility [11]. Hyperinsulinemia is also linked to follicular dysplasia and impaired synthesis of sexual hormones in the follicle (less progesterone, lower availability of follicle stimulating hormone, and more androgens) [12, 13]. Our findings support hyperinsulinemia as a cause of infertility in women. The lack of robust statistical evidence for an effect of lipids and blood pressure on infertility suggests that previous observational results [1-3] may have been influenced by residual confounding.

Our work has several strengths. MR studies exploring the relationship between cardiometabolic risk factors and a clinical definition of fertility have been lacking. Our findings were based on a relatively homogeneous population, and together with a thorough evaluation of compliance with MR assumptions, our findings appear robust. However, our study has some limitations. First, infertility is a couple-dependent measure. Thus, we could not establish if underlying causes of infertility were in the woman, man, or both. Second, MoBa only recruited couples who had conceived. Therefore, studies with couples who never conceived are warranted. Third, we did not have data on circulating levels of cardiometabolic risk factors in MoBa and there was no GWAS of infertility with available summary data across the genome. Thus, we opted to undertake a GWAS of infertility in MoBa (with no independent replication) and a two-sample MR using summary results from publicly available GWAS of the exposures and those from our MoBa GWAS. Our outcome GWAS is relatively small. Thus, we have highlighted some suggested results that were clinically relevant but did not pass multiple testing. Our findings need replication in two-sample MR using results from a larger GWAS of infertility and/or a large independent sample in which one-sample MR could be undertaken. Fourth, no sex-specific genetic instruments were available in the largest GWASs on cardiometabolic risk factors, and we had to assume that there were no sex differences in the genetic instruments for exposures. However, this assumption may imply a violation of the first MR principle. Finally, the characteristics of our study sample limits the generalizability of our results to other populations, particularly to populations of genetic ancestries other than European.

In conclusion, genetic instruments suggest a role of hyperinsulinemia and possibly glucose intolerance on infertility in women. If further research in larger samples and with sex-specific exposure instruments support our findings, preventing hyperinsulinemia may be a means of reducing fertility problems in women.

## Data Availability

The consent given by the participants does not allow for storage of individual data in repositories or journals. Researchers who want access to data sets for replication should apply through. This process requires approval from the Regional Committee for Medical and Health Research Ethics in Norway and an agreement with MoBa. GWAS summary statistics on fasting glucose, glycated hemoglobin, and fasting insulin have been contributed by the Meta-Analyses of Glucose and Insulin-related traits Consortium investigators and have been downloaded from https://magicinvestigators.org/downloads/ (menu: Trans-ancestry and single-ancestry meta-analysis summary statistics of four glycemic traits). GWAS summary statistics on lipid profile traits have been contributed by the Global Lipids Genetics Consortium investigators and have been downloaded from http://csg.sph.umich.edu/willer/public/glgc-lipids2021/ (menu: Ancestry-specific GWAS summary statistics for HDL-C, LDL-C, nonHDL-C, TC and TG). Finally, GWAS summary statistics on blood pressure have been contributed by Evangelou E et al. and have been downloaded from the Supplemental Tables of the article (https://www.nature.com/articles/s41588-018-0205-x#MOESM3).

## Acknowledgements

The MoBa Cohort Study is supported by the Norwegian Ministry of Health and Care Services and the Ministry of Education and Research. We are grateful to all the families in Norway who take part in this cohort study, those who contributed to the recruitment, and the infrastructure surrounding the MoBa cohort.

We thank Per Magnus, Pål R. Njølstad, Ole Andreassen, and the Norwegian Institute of Public Health for generating high-quality genomic data. This research is part of the HARVEST collaboration, supported by the Research Council of Norway (#229624). We also thank the NORMENT Centre for providing genotype data, funded by the Research Council of Norway (grant #223273), South East Norway Health Authority, Stiftelsen Kristian Gerhard Jebsen, and deCODE Genetics. We further thank the Center for Diabetes Research (University of Bergen) for providing genotype information and performing quality control and imputation of the data, in the context of research projects funded by the European Research Council Advanced Grant SELECTionPREDISPOSED (#293574), Stiftelsen Kristian Gerhard Jebsen (Translational Medical Center), the Trond Mohn Foundation, the Research Council of Norway (FRIPRO grant #240413), the Novo Nordisk Foundation (grant #54741), the Bergen Research Foundation (“Utilizing the Mother and Child Cohort and the Medical Birth Registry for Better Health”), the University of Bergen, the Western Norway Health Authority (Strategic Fund “Personalized Medicine for Children and Adults”), and the Norwegian Diabetes Association.

This work was performed on the TSD (Tjeneste for Sensitive Data) facilities, owned by the University of Oslo, which are operated and developed by the TSD service group at the IT-Department in the University of Oslo.

## Data availability

The consent given by the participants does not allow for storage of individual data in repositories or journals. Researchers who want access to data sets for replication should apply through. This process requires approval from the Regional Committee for Medical and Health Research Ethics in Norway and an agreement with MoBa. GWAS summary statistics on fasting glucose, glycated hemoglobin, and fasting insulin have been contributed by the Meta-Analyses of Glucose and Insulin-related traits Consortium investigators and have been downloaded from https://magicinvestigators.org/downloads/ (menu: “Trans-ancestry and single-ancestry meta-analysis summary statistics of four glycemic traits”). GWAS summary statistics on lipid profile traits have been contributed by the Global Lipids Genetics Consortium investigators and have been downloaded from http://csg.sph.umich.edu/willer/public/glgc-lipids2021/ (menu: “Ancestry-specific GWAS summary statistics for HDL-C, LDL-C, nonHDL-C, TC and TG”). Finally, GWAS summary statistics on blood pressure have been contributed by Evangelou E et al. and have been downloaded from the Supplemental Tables of the article (https://www.nature.com/articles/s41588-018-0205-x#MOESM3).

## Authors’ relationships and activities

D.A.L. receives (or has received in the last 10 years) research support from National and International government and charitable bodies, Roche Diagnostics and Medtronic for research unrelated to the current work. O.A.A. is or has been in the last 10 years consultant to HealthLytix. The rest of the authors declare that no competing interests exist.

## Contribution statement

A.Hernaez was responsible for data curation, formal analyses, data interpretation, and drafting the article. Y.L. performed data analyses (the temporary GWAS on infertility in MoBa) and critically revised the article. C.M.P. provided support in data analysis, software use, and visualization of results, and critically revised the article. K.H.S. provided support in data analysis, software use, and visualization of results, and critically revised the article. S.E.H. contributed to data acquisition, obtained funding, and critically revised the article. P.M. contributed to data acquisition and critically revised the article. P.R.N. contributed to data acquisition and critically revised the article. O.A.A. contributed to data acquisition and critically revised the article. E.C. contributed to data acquisition and critically revised the article. A.Havdahl contributed to data acquisition and critically revised the article. A.F. was involved in the definition of the methodology of the study and the interpretation of data and critically revised the article. S.B. was involved in the definition of the methodology of the study and the interpretation of data and critically revised the article. D.A.L. was involved in the definition of the methodology of the study and the interpretation of data and critically revised the article. M.C.M. conceived and designed the study, obtained funding, coordinated the project, contributed to data interpretation, and critically revised the article. A.Hernaez and M.C.M. are the guarantors of this study, accept full responsibility for the work and the conduct of the study, had access to the data, and controlled the decision to publish.

## ABBREVIATIONS

DBP: diastolic blood pressure
GWAS: genome-wide association study
HDL-C: high-density lipoprotein cholesterol
LDL-C: low-density lipoprotein cholesterol
MoBa: Mother, Father and Child Cohort Study
MR: Mendelian randomization
SBP: systolic blood pressure

## SUPPLEMENTAL MATERIALS

### SUPPLEMENTAL METHODS

#### MR assumptions and sensitivity analyses

There are three key assumptions in MR: 1) genetic instruments are robustly related to the exposure, 2) there is no confounding of the genetic instrument-outcome associations, and 3) genetic instruments are exclusively linked to the outcome through the exposure of interest. If violated, MR conclusions may be invalid [1].

In relation to the first assumption, we estimated the robustness of our genetic instruments as the mean F-statistic (average beta^2^/standard error^2^ across all SNPs; F-statistics < 10 suggest a weak genetic instrument) [1]. Regarding the second assumption, we reduced the potential confounding of the genetic instrument-outcome association due to population stratification by: 1) selecting our genetic instruments from GWASs performed in populations of European ancestry; 2) conducting MR analyses using GWASs adjusted for genetic principal components [2].

Genetic instruments that are related to other risk factors for the outcome different than the exposure of interest (horizontal pleiotropy) violate the third MR assumption [3]. We checked the presence of horizontal pleiotropy by procedures based on sensitivity MR methods (MR-Egger, weighted median, and weighted mode) [4]. It can be identified if: 1) a non-zero intercept is present in the MR-Egger method; 2) there is no concordance among MR estimates in inverse variance weighted and the alternative methods; 3) and between SNP heterogeneity is found according to the Cochran’s Qand the Rücker’s *Q*’ [4]. Some associations were affected by horizontal pleiotropy, possibly due to the high inter-relation among cardiometabolic risk factors. We thus used the Steiger filtering to determine which SNPs explain more variation in our exposure of interest than in the rest of variables related to it (these SNPs are less susceptible of biasing findings due to horizontal pleiotropy) [4]. For example, 71 of the SNPs for fasting glucose explained more variability in fasting glucose than in the other risk factors considered (the rest of risk factors [glycated hemoglobin, fasting insulin, LDL-C, HDL-C, triglycerides, SBP, DBP] plus BMI and ever smoking) and were retained after the filtering. We repeated MR analyses using exclusively the SNPs that passed the filtering process.

### SUPPLEMENTAL TABLES

**Supplemental Table 1.** SNPs included in fasting glucose-related analyses.

| RSID | Chrom. | Position | Effect allele | Other allele | Effect allele freq. | Beta | SE | p-value | Used in MR | Used in MR + Steiger filt. |
| --- | --- | --- | --- | --- | --- | --- | --- | --- | --- | --- |
| rs6662924 | 1 | 100894419 | A | C | 0.19 | 0.014 | 0.002 | 3.34E-10 | Yes | Yes |
| rs78132593 | 1 | 150868102 | A | C | 0.21 | -0.015 | 0.002 | 2.60E-10 | Yes | No |
| rs267738 | 1 | 150940625 | T | G | 0.79 | 0.013 | 0.002 | 3.23E-10 | Yes | No |
| rs2075423 | 1 | 214154719 | T | G | 0.37 | -0.016 | 0.002 | 3.18E-21 | Yes | Yes |
| rs340874 | 1 | 214159256 | T | C | 0.47 | -0.015 | 0.002 | 2.58E-20 | Yes | Yes |
| rs348330 | 1 | 229672955 | A | G | 0.63 | -0.012 | 0.002 | 3.04E-10 | No | No |
| rs877273 | 2 | 27140022 | T | C | 0.38 | 0.013 | 0.002 | 7.22E-15 | Yes | Yes |
| rs1371614 | 2 | 27152874 | T | C | 0.25 | 0.016 | 0.002 | 5.85E-16 | Yes | Yes |
| rs1260326 | 2 | 27730940 | T | C | 0.39 | -0.028 | 0.002 | 4.48E-65 | Yes | Yes |
| rs780093 | 2 | 27742603 | T | C | 0.38 | -0.028 | 0.002 | 1.53E-59 | Yes | Yes |
| rs115640879 | 2 | 28268742 | T | C | 0.08 | -0.024 | 0.004 | 1.30E-11 | Yes | No |
| rs183381538 | 2 | 43775309 | A | C | 0.07 | -0.025 | 0.004 | 7.81E-15 | No | No |
| rs189548 | 2 | 54941112 | A | G | 0.73 | -0.012 | 0.002 | 2.81E-09 | Yes | Yes |
| rs78996387 | 2 | 169238040 | T | C | 0.08 | -0.023 | 0.004 | 1.03E-11 | Yes | No |
| rs72883292 | 2 | 169710158 | T | C | 0.02 | -0.009 | 0.007 | 0.131 | Yes | No |
| rs150171632 | 2 | 169748691 | T | C | 0.01 | -0.046 | 0.011 | 6.63E-05 | No | No |
| rs111485380 | 2 | 169754123 | T | C | 0.83 | 0.07 | 0.003 | 1.02E-137 | No | No |
| rs540524 | 2 | 169756930 | A | G | 0.62 | -0.011 | 0.002 | 1.12E-09 | Yes | Yes |
| rs1402837 | 2 | 169757354 | T | C | 0.22 | 0.056 | 0.002 | 5.38E-162 | Yes | Yes |
| rs573225 | 2 | 169757541 | A | G | 0.68 | 0.069 | 0.002 | 0 | Yes | Yes |
| rs560887 | 2 | 169763148 | T | C | 0.3 | -0.075 | 0.002 | 0 | Yes | Yes |
| rs492594 | 2 | 169764176 | C | G | 0.46 | 0.016 | 0.002 | 7.36E-24 | No | No |
| rs145353824 | 2 | 169765277 | A | C | 0.98 | -0.076 | 0.009 | 7.21E-18 | No | No |
| rs17539351 | 2 | 169766560 | T | C | 0.12 | -0.068 | 0.003 | 5.86E-123 | Yes | Yes |
| rs13430620 | 2 | 169768891 | A | C | 0.96 | 0.073 | 0.006 | 3.43E-40 | No | No |
| rs563694 | 2 | 169774071 | A | C | 0.65 | 0.066 | 0.002 | 0 | Yes | Yes |
| rs114764002 | 2 | 169776141 | A | T | 0.97 | 0.092 | 0.007 | 3.41E-43 | No | No |
| rs191257736 | 2 | 169778471 | A | G | 0.02 | 0.036 | 0.009 | 3.21E-05 | No | No |
| rs56100844 | 2 | 169786707 | T | G | 0.98 | 0.118 | 0.01 | 9.82E-35 | Yes | No |
| rs3755158 | 2 | 169792188 | C | G | 0.89 | -0.04 | 0.003 | 4.83E-49 | Yes | Yes |
| rs508741 | 2 | 169795288 | T | C | 0.64 | -0.013 | 0.002 | 1.91E-12 | Yes | Yes |
| rs199666154 | 2 | 169800434 | G | GT | 0.73 | -0.044 | 0.003 | 1.52E-64 | No | No |
| rs114691375 | 2 | 169813318 | A | T | 0.03 | 0.045 | 0.006 | 7.79E-17 | Yes | No |
| rs6731931 | 2 | 173593726 | T | C | 0.8 | -0.012 | 0.002 | 1.74E-07 | Yes | No |
| rs11708067 | 3 | 123065778 | A | G | 0.77 | 0.028 | 0.002 | 1.63E-43 | Yes | Yes |
| rs75098673 | 3 | 141094338 | T | C | 0.05 | -0.035 | 0.005 | 2.31E-11 | Yes | No |
| rs16851397 | 3 | 141134818 | A | G | 0.95 | 0.033 | 0.004 | 1.26E-12 | Yes | Yes |
| rs17437560 | 3 | 152180329 | T | C | 0.11 | -0.018 | 0.003 | 3.33E-08 | Yes | Yes |
| rs1604038 | 3 | 170709193 | T | C | 0.29 | -0.02 | 0.002 | 4.47E-28 | Yes | Yes |
| rs3215234 | 3 | 170724091 | G | GT | 0.87 | 0.025 | 0.003 | 6.50E-24 | No | No |
| rs7631557 | 3 | 185513646 | C | G | 0.3 | 0.011 | 0.002 | 8.10E-10 | Yes | No |
| rs35188816 | 3 | 185526108 | C | CA | 0.29 | 0.012 | 0.002 | 8.67E-11 | No | No |
| rs6808574 | 3 | 187740523 | T | C | 0.39 | -0.013 | 0.002 | 7.21E-14 | Yes | Yes |
| rs4862423 | 4 | 185726548 | T | C | 0.4 | 0.012 | 0.002 | 4.45E-10 | Yes | Yes |
| rs157512 | 5 | 55809127 | T | C | 0.76 | 0.013 | 0.002 | 5.43E-10 | Yes | No |
| rs7708285 | 5 | 76425867 | A | G | 0.73 | -0.013 | 0.002 | 1.25E-09 | Yes | Yes |
| rs6878122 | 5 | 76427311 | A | G | 0.72 | -0.013 | 0.002 | 2.36E-09 | Yes | No |
| rs1820176 | 5 | 95696585 | T | C | 0.71 | 0.025 | 0.002 | 1.91E-34 | Yes | No |
| rs6876986 | 5 | 95703329 | C | G | 0.29 | -0.024 | 0.002 | 3.07E-33 | Yes | No |
| rs7729395 | 5 | 102100576 | T | C | 0.05 | -0.023 | 0.005 | 3.09E-08 | Yes | Yes |
| rs9379084 | 6 | 7231843 | A | G | 0.13 | -0.012 | 0.003 | 5.25E-06 | Yes | Yes |
| rs3778321 | 6 | 7250270 | A | G | 0.19 | -0.019 | 0.002 | 3.16E-17 | Yes | Yes |
| rs9348441 | 6 | 20680678 | A | T | 0.28 | 0.018 | 0.002 | 4.40E-20 | Yes | No |
| rs10305457 | 6 | 39034095 | T | C | 0.09 | 0.024 | 0.003 | 1.21E-14 | Yes | No |
| rs10305492 | 6 | 39046794 | A | G | 0.014 | -0.076 | 0.01 | 7.50E-16 | Yes | Yes |
| rs12055786 | 6 | 153431125 | T | C | 0.42 | 0.012 | 0.002 | 1.17E-11 | Yes | Yes |
| rs10281892 | 7 | 14919852 | A | G | 0.82 | -0.028 | 0.002 | 2.09E-35 | Yes | No |
| rs2191346 | 7 | 15053878 | C | G | 0.46 | -0.024 | 0.002 | 1.51E-43 | No | No |
| rs10487796 | 7 | 15063430 | A | T | 0.48 | -0.026 | 0.002 | 4.62E-52 | No | No |
| rs10228796 | 7 | 15064190 | C | G | 0.48 | -0.026 | 0.002 | 7.28E-52 | No | No |
| rs55841377 | 7 | 44145178 | C | G | 0.78 | 0.009 | 0.002 | 1.01E-05 | Yes | No |
| rs2971671 | 7 | 44211337 | T | C | 0.77 | -0.043 | 0.002 | 1.24E-104 | Yes | Yes |
| rs2971670 | 7 | 44226101 | T | C | 0.17 | 0.061 | 0.002 | 1.88E-165 | Yes | No |
| rs1799884 | 7 | 44229068 | T | C | 0.17 | 0.062 | 0.002 | 2.51E-167 | Yes | Yes |
| rs3757840 | 7 | 44231216 | T | G | 0.49 | 0.042 | 0.002 | 5.94E-133 | Yes | Yes |
| rs6975024 | 7 | 44231886 | T | C | 0.84 | -0.062 | 0.002 | 4.66E-167 | Yes | Yes |
| rs2908286 | 7 | 44234737 | T | C | 0.17 | 0.061 | 0.002 | 1.88E-165 | Yes | No |
| rs138917529 | 7 | 44235694 | A | T | 0.98 | 0.06 | 0.007 | 4.08E-17 | Yes | No |
| rs878521 | 7 | 44255643 | A | G | 0.24 | 0.055 | 0.002 | 2.65E-174 | Yes | No |
| rs2108349 | 7 | 50786663 | A | G | 0.66 | -0.016 | 0.002 | 1.25E-15 | Yes | Yes |
| rs58925536 | 7 | 75654574 | T | C | 0.03 | 0.031 | 0.005 | 5.82E-09 | Yes | No |
| rs67070387 | 7 | 75836023 | T | C | 0.03 | 0.031 | 0.006 | 1.60E-08 | Yes | No |
| rs13242882 | 7 | 89800053 | A | T | 0.46 | -0.01 | 0.002 | 9.27E-09 | No | No |
| rs194520 | 7 | 89854446 | T | G | 0.45 | -0.009 | 0.002 | 1.14E-08 | Yes | Yes |
| rs7012637 | 8 | 9173209 | A | G | 0.47 | -0.018 | 0.002 | 9.75E-25 | Yes | No |
| rs9987289 | 8 | 9183358 | A | G | 0.1 | 0.028 | 0.003 | 3.57E-23 | Yes | No |
| rs4841132 | 8 | 9183596 | A | G | 0.1 | 0.028 | 0.003 | 1.72E-22 | Yes | No |
| rs12541643 | 8 | 81076874 | T | C | 0.48 | 0.012 | 0.002 | 4.51E-09 | Yes | No |
| rs896854 | 8 | 95960511 | T | C | 0.5 | 0.01 | 0.002 | 5.61E-09 | Yes | Yes |
| rs13266634 | 8 | 118184783 | T | C | 0.32 | -0.029 | 0.002 | 4.81E-57 | Yes | Yes |
| rs9650069 | 8 | 118204020 | T | C | 0.32 | -0.029 | 0.002 | 8.31E-58 | Yes | No |
| rs1574285 | 9 | 4283137 | T | G | 0.59 | -0.019 | 0.002 | 6.85E-30 | Yes | Yes |
| rs57884925 | 9 | 4285119 | C | G | 0.51 | -0.017 | 0.002 | 4.00E-24 | No | No |
| rs10974438 | 9 | 4291928 | A | C | 0.64 | -0.02 | 0.002 | 9.85E-31 | Yes | Yes |
| rs10217762 | 9 | 22133645 | T | C | 0.56 | -0.003 | 0.002 | 0.26 | Yes | No |
| rs10811660 | 9 | 22134068 | A | G | 0.18 | -0.022 | 0.002 | 7.94E-25 | Yes | No |
| rs16913693 | 9 | 111680359 | T | G | 0.97 | 0.039 | 0.005 | 2.82E-16 | Yes | Yes |
| rs507666 | 9 | 136149399 | A | G | 0.2 | 0.016 | 0.002 | 6.99E-17 | Yes | No |
| rs3829109 | 9 | 139256766 | A | G | 0.3 | -0.016 | 0.002 | 1.09E-15 | Yes | Yes |
| rs10781511 | 9 | 139280766 | A | G | 0.27 | -0.011 | 0.002 | 1.33E-06 | Yes | No |
| rs2839671 | 10 | 26505822 | A | G | 0.17 | -0.016 | 0.002 | 8.38E-14 | Yes | Yes |
| rs7095788 | 10 | 95384152 | T | C | 0.35 | -0.011 | 0.002 | 1.98E-09 | Yes | Yes |
| rs35011531 | 10 | 113032095 | T | C | 0.09 | -0.033 | 0.003 | 4.26E-31 | Yes | No |
| rs12784552 | 10 | 113036354 | A | G | 0.91 | 0.033 | 0.003 | 2.86E-31 | Yes | Yes |
| rs11195538 | 10 | 113117650 | T | C | 0.93 | 0.019 | 0.004 | 6.23E-07 | Yes | Yes |
| rs61875120 | 10 | 114753259 | T | C | 0.79 | -0.027 | 0.002 | 1.93E-32 | Yes | No |
| rs34872471 | 10 | 114754071 | T | C | 0.72 | -0.026 | 0.002 | 5.41E-35 | Yes | No |
| rs7903146 | 10 | 114758349 | T | C | 0.27 | 0.026 | 0.002 | 2.00E-35 | Yes | Yes |
| rs3842753 | 11 | 2181060 | T | G | 0.28 | 0.013 | 0.002 | 2.84E-09 | Yes | No |
| rs689 | 11 | 2182224 | A | T | 0.28 | 0.013 | 0.002 | 3.27E-09 | Yes | No |
| rs4930011 | 11 | 2856658 | C | G | 0.59 | -0.009 | 0.002 | 2.84E-08 | Yes | No |
| rs2168101 | 11 | 8255408 | A | C | 0.32 | -0.013 | 0.002 | 3.09E-08 | No | No |
| rs10838524 | 11 | 45870177 | A | G | 0.45 | 0.024 | 0.002 | 1.56E-40 | Yes | Yes |
| rs7115753 | 11 | 45912013 | A | G | 0.46 | 0.024 | 0.002 | 2.83E-39 | Yes | No |
| rs8914 | 11 | 46699124 | A | G | 0.11 | -0.02 | 0.003 | 4.14E-13 | Yes | Yes |
| rs10501320 | 11 | 47293799 | C | G | 0.25 | -0.022 | 0.002 | 1.09E-29 | Yes | Yes |
| rs10717442 | 11 | 47340680 | A | AT | 0.23 | 0.022 | 0.002 | 5.13E-18 | No | No |
| rs34228231 | 11 | 47820241 | A | G | 0.23 | -0.018 | 0.002 | 1.32E-18 | Yes | No |
| rs1483121 | 11 | 48333360 | A | G | 0.14 | -0.016 | 0.003 | 7.99E-10 | Yes | Yes |
| rs10769572 | 11 | 49313235 | A | G | 0.25 | 0.011 | 0.002 | 4.07E-08 | Yes | Yes |
| rs174583 | 11 | 61609750 | T | C | 0.36 | -0.017 | 0.002 | 3.37E-22 | Yes | No |
| rs77464186 | 11 | 72460398 | A | C | 0.82 | 0.023 | 0.002 | 3.50E-25 | Yes | No |
| rs11603349 | 11 | 72460694 | T | C | 0.82 | 0.024 | 0.002 | 3.12E-25 | Yes | No |
| rs7113297 | 11 | 92671744 | T | C | 0.21 | 0.057 | 0.003 | 7.09E-100 | No | No |
| rs11523890 | 11 | 92679778 | T | C | 0.34 | 0.052 | 0.002 | 6.75E-161 | Yes | Yes |
| rs10466351 | 11 | 92697981 | T | C | 0.38 | 0.056 | 0.002 | 1.14E-205 | Yes | No |
| rs10830962 | 11 | 92698427 | C | G | 0.6 | -0.048 | 0.002 | 6.62E-156 | Yes | Yes |
| rs10830963 | 11 | 92708710 | C | G | 0.71 | -0.077 | 0.002 | 0 | Yes | Yes |
| rs79354397 | 11 | 92708961 | T | C | 0.97 | -0.081 | 0.006 | 4.82E-45 | Yes | No |
| rs2657879 | 12 | 56865338 | A | G | 0.82 | -0.012 | 0.002 | 7.33E-09 | Yes | Yes |
| rs12315434 | 12 | 57780936 | A | C | 0.77 | 0.011 | 0.002 | 4.41E-08 | Yes | No |
| rs6538804 | 12 | 97848910 | C | G | 0.61 | 0.014 | 0.002 | 9.41E-14 | Yes | Yes |
| rs6489811 | 12 | 121893626 | A | G | 0.47 | -0.011 | 0.002 | 3.27E-09 | Yes | Yes |
| rs7962128 | 12 | 121907336 | A | G | 0.44 | -0.011 | 0.002 | 1.78E-08 | Yes | No |
| rs11610045 | 12 | 133063768 | A | G | 0.5 | 0.014 | 0.002 | 3.26E-13 | Yes | No |
| rs11619319 | 13 | 28487599 | A | G | 0.78 | -0.017 | 0.002 | 3.41E-20 | Yes | Yes |
| rs576674 | 13 | 33554302 | A | G | 0.84 | -0.018 | 0.002 | 9.70E-13 | Yes | Yes |
| rs8003022 | 14 | 90038821 | A | G | 0.61 | -0.013 | 0.002 | 5.27E-10 | Yes | Yes |
| rs35889227 | 14 | 90055468 | T | G | 0.62 | -0.013 | 0.002 | 3.37E-10 | Yes | No |
| rs12888855 | 14 | 100830818 | A | C | 0.21 | -0.014 | 0.002 | 6.02E-12 | Yes | Yes |
| rs7163757 | 15 | 62391608 | T | C | 0.45 | -0.022 | 0.002 | 2.64E-36 | Yes | Yes |
| rs12594062 | 15 | 75102851 | T | C | 0.36 | 0.01 | 0.002 | 4.64E-09 | Yes | No |
| rs11630478 | 15 | 75102923 | T | G | 0.64 | -0.01 | 0.002 | 4.64E-09 | Yes | No |
| rs7178572 | 15 | 77747190 | A | G | 0.3 | -0.012 | 0.002 | 7.09E-10 | Yes | Yes |
| rs11633054 | 15 | 77747276 | A | G | 0.3 | -0.013 | 0.002 | 3.82E-09 | Yes | Yes |
| rs6598541 | 15 | 99271135 | A | G | 0.36 | 0.011 | 0.002 | 4.12E-12 | Yes | Yes |
| rs2238435 | 16 | 4014282 | C | G | 0.4 | 0.011 | 0.002 | 3.82E-09 | Yes | No |
| rs2302593 | 19 | 46196634 | C | G | 0.5 | 0.011 | 0.002 | 5.67E-10 | No | No |
| rs6113722 | 20 | 22557099 | A | G | 0.04 | -0.042 | 0.004 | 7.66E-25 | Yes | Yes |
| rs3833331 | 20 | 22562326 | A | AG | 0.95 | 0.045 | 0.005 | 1.92E-22 | No | No |
| rs1337918 | 20 | 22567608 | A | C | 0.04 | -0.046 | 0.005 | 2.30E-22 | Yes | No |
| rs17265513 | 20 | 39832628 | T | C | 0.79 | -0.016 | 0.002 | 5.10E-14 | Yes | Yes |
| rs39713 | 22 | 30343186 | T | C | 0.09 | -0.017 | 0.003 | 1.77E-08 | Yes | Yes |

**Supplemental Table 2.** SNPs included in glycated hemoglobin-related analyses.

| RSID | Chrom. | Position | Effect allele | Other allele | Effect allele freq. | Beta | SE | p-value | Used in MR | Used in MR + Steiger filt. |
| --- | --- | --- | --- | --- | --- | --- | --- | --- | --- | --- |
| rs1175549 | 1 | 3691727 | A | C | 0.75 | 0.01 | 0.002 | 7.13E-13 | Yes | Yes |
| rs2375278 | 1 | 25529038 | A | G | 0.17 | 0.011 | 0.002 | 1.05E-11 | Yes | Yes |
| rs644592 | 1 | 25703156 | T | C | 0.15 | 0.013 | 0.002 | 1.58E-11 | Yes | No |
| rs267738 | 1 | 150940625 | T | G | 0.79 | 0.011 | 0.002 | 1.14E-11 | Yes | Yes |
| rs7534795 | 1 | 155275553 | T | C | 0.26 | 0.01 | 0.002 | 2.13E-09 | No | No |
| rs10796941 | 1 | 155284261 | T | C | 0.25 | 0.01 | 0.002 | 9.22E-09 | Yes | No |
| rs857725 | 1 | 158607935 | T | G | 0.73 | -0.021 | 0.001 | 5.43E-55 | Yes | Yes |
| rs857676 | 1 | 158615702 | A | G | 0.57 | 0.01 | 0.001 | 7.04E-16 | Yes | Yes |
| rs7547793 | 1 | 203653544 | A | C | 0.13 | -0.012 | 0.002 | 6.61E-09 | Yes | No |
| rs10900585 | 1 | 203654024 | T | G | 0.88 | 0.012 | 0.002 | 7.97E-09 | Yes | No |
| rs340882 | 1 | 214145731 | C | G | 0.41 | -0.008 | 0.001 | 1.48E-10 | Yes | No |
| rs12612492 | 2 | 24093756 | T | C | 0.12 | 0.019 | 0.002 | 1.88E-26 | Yes | Yes |
| rs6545222 | 2 | 24235704 | A | G | 0.75 | 0.01 | 0.002 | 1.86E-14 | Yes | Yes |
| rs1367173 | 2 | 43449385 | T | C | 0.12 | -0.015 | 0.002 | 1.66E-14 | Yes | Yes |
| rs6723441 | 2 | 48123915 | A | T | 0.81 | -0.009 | 0.002 | 2.26E-08 | Yes | Yes |
| rs17037289 | 2 | 48587198 | A | G | 0.75 | -0.009 | 0.002 | 2.40E-09 | Yes | No |
| rs150171632 | 2 | 169748691 | T | C | 0.01 | -0.033 | 0.009 | 4.48E-05 | No | No |
| rs540524 | 2 | 169756930 | A | G | 0.63 | -0.002 | 0.001 | 0.167 | Yes | No |
| rs560887 | 2 | 169763148 | T | C | 0.3 | -0.031 | 0.001 | 5.55E-122 | Yes | No |
| rs13419326 | 2 | 169771420 | A | C | 0.98 | -0.032 | 0.005 | 5.60E-13 | Yes | No |
| rs557462 | 2 | 169777595 | T | C | 0.65 | 0.027 | 0.001 | 3.94E-102 | Yes | No |
| rs56100844 | 2 | 169786707 | T | G | 0.98 | 0.058 | 0.007 | 4.47E-17 | Yes | No |
| rs17256082 | 2 | 175292364 | T | C | 0.67 | -0.007 | 0.001 | 3.19E-08 | Yes | Yes |
| rs4674280 | 2 | 219141458 | C | G | 0.57 | 0.008 | 0.001 | 5.48E-09 | No | No |
| rs13427681 | 2 | 219167563 | C | G | 0.57 | 0.008 | 0.001 | 1.27E-08 | No | No |
| rs1822534 | 3 | 12266804 | A | G | 0.62 | 0.008 | 0.001 | 3.65E-13 | Yes | No |
| rs12491937 | 3 | 12268244 | A | G | 0.58 | 0.009 | 0.001 | 1.42E-13 | Yes | No |
| rs9818758 | 3 | 49382925 | A | G | 0.18 | 0.013 | 0.002 | 1.49E-13 | Yes | Yes |
| rs66499923 | 3 | 52887861 | A | AT | 0.61 | -0.008 | 0.002 | 9.35E-09 | No | No |
| rs11719201 | 3 | 123068744 | T | C | 0.24 | -0.013 | 0.002 | 2.43E-18 | Yes | No |
| rs1604038 | 3 | 170709193 | T | C | 0.29 | -0.011 | 0.001 | 2.76E-16 | Yes | No |
| rs4894769 | 3 | 171516306 | A | T | 0.43 | -0.007 | 0.001 | 3.61E-09 | No | No |
| rs13089972 | 3 | 171798694 | A | T | 0.58 | 0.011 | 0.001 | 1.87E-15 | No | No |
| rs7632281 | 3 | 171812293 | T | G | 0.43 | -0.011 | 0.001 | 1.54E-14 | Yes | No |
| rs13134327 | 4 | 144659795 | A | G | 0.33 | 0.014 | 0.001 | 2.81E-26 | Yes | Yes |
| rs13129993 | 4 | 144684229 | A | T | 0.32 | 0.015 | 0.002 | 1.61E-24 | Yes | Yes |
| rs112578089 | 4 | 145128105 | A | G | 0.04 | -0.036 | 0.006 | 2.18E-11 | No | No |
| rs77909720 | 4 | 145222284 | T | C | 0.04 | -0.023 | 0.004 | 1.63E-09 | No | No |
| rs6877043 | 5 | 154048367 | T | C | 0.64 | 0.009 | 0.001 | 1.99E-10 | Yes | Yes |
| rs1948759 | 5 | 156442657 | A | G | 0.17 | -0.01 | 0.002 | 2.44E-08 | Yes | No |
| rs3778321 | 6 | 7250270 | A | G | 0.19 | -0.011 | 0.002 | 4.18E-11 | Yes | No |
| rs34499031 | 6 | 20676414 | T | TAA | 0.72 | -0.01 | 0.002 | 2.93E-12 | No | No |
| rs35612982 | 6 | 20682622 | T | C | 0.82 | -0.01 | 0.002 | 2.47E-10 | Yes | No |
| rs6931514 | 6 | 20703952 | A | G | 0.73 | -0.01 | 0.001 | 1.18E-13 | Yes | No |
| rs12193223 | 6 | 24978511 | C | G | 0.94 | 0.02 | 0.003 | 5.87E-09 | Yes | Yes |
| rs75580845 | 6 | 25578433 | T | C | 0.92 | 0.02 | 0.002 | 1.23E-20 | Yes | No |
| rs1799945 | 6 | 26091179 | C | G | 0.86 | 0.025 | 0.002 | 3.32E-47 | Yes | Yes |
| rs1800562 | 6 | 26093141 | A | G | 0.06 | -0.038 | 0.003 | 2.33E-50 | Yes | Yes |
| rs13194491 | 6 | 27037080 | T | C | 0.07 | -0.024 | 0.003 | 1.20E-19 | No | No |
| rs13214703 | 6 | 27941387 | T | C | 0.93 | 0.02 | 0.003 | 2.49E-15 | Yes | Yes |
| rs34979126 | 6 | 28449380 | A | G | 0.07 | -0.018 | 0.003 | 5.60E-13 | Yes | No |
| rs116735744 | 6 | 28984755 | T | G | 0.24 | -0.011 | 0.002 | 2.66E-10 | No | No |
| rs6929796 | 6 | 31522669 | A | G | 0.17 | -0.009 | 0.002 | 3.26E-08 | Yes | Yes |
| rs9376090 | 6 | 135411228 | T | C | 0.73 | 0.025 | 0.001 | 1.90E-62 | Yes | Yes |
| rs9389268 | 6 | 135419631 | A | G | 0.73 | 0.023 | 0.002 | 1.62E-51 | Yes | Yes |
| rs10231021 | 7 | 15060429 | A | T | 0.51 | 0.009 | 0.001 | 8.69E-14 | No | No |
| rs2191349 | 7 | 15064309 | T | G | 0.53 | 0.008 | 0.001 | 1.87E-12 | Yes | No |
| rs10259649 | 7 | 44219705 | T | C | 0.77 | -0.024 | 0.002 | 4.02E-55 | Yes | No |
| rs2971670 | 7 | 44226101 | T | C | 0.17 | 0.032 | 0.002 | 5.10E-88 | Yes | No |
| rs1799884 | 7 | 44229068 | T | C | 0.17 | 0.032 | 0.002 | 1.62E-87 | Yes | No |
| rs3757840 | 7 | 44231216 | T | G | 0.5 | 0.022 | 0.001 | 4.25E-71 | Yes | No |
| rs2908286 | 7 | 44234737 | T | C | 0.17 | 0.032 | 0.002 | 1.62E-87 | Yes | No |
| rs35332062 | 7 | 73012042 | A | G | 0.12 | 0.011 | 0.002 | 4.66E-09 | Yes | No |
| rs13234131 | 7 | 73025975 | A | G | 0.87 | -0.011 | 0.002 | 2.06E-09 | Yes | No |
| rs4731113 | 7 | 123283949 | T | C | 0.96 | 0.02 | 0.004 | 4.90E-08 | Yes | No |
| rs4317621 | 8 | 41516581 | A | G | 0.42 | -0.004 | 0.001 | 0.00784 | Yes | No |
| rs34664882 | 8 | 41543675 | A | G | 0.03 | -0.049 | 0.004 | 4.82E-37 | Yes | No |
| rs4737009 | 8 | 41630405 | A | G | 0.24 | 0.023 | 0.002 | 8.29E-56 | Yes | Yes |
| rs6980507 | 8 | 42383084 | A | G | 0.4 | 0.011 | 0.001 | 8.15E-20 | Yes | Yes |
| rs11558471 | 8 | 118185733 | A | G | 0.68 | 0.015 | 0.001 | 3.38E-25 | Yes | No |
| rs35859536 | 8 | 118191475 | T | C | 0.32 | -0.015 | 0.001 | 1.15E-24 | Yes | No |
| rs2954021 | 8 | 126482077 | A | G | 0.49 | -0.007 | 0.001 | 1.92E-10 | Yes | No |
| rs10811661 | 9 | 22134094 | T | C | 0.82 | 0.013 | 0.002 | 1.74E-14 | Yes | No |
| rs10811662 | 9 | 22134253 | A | G | 0.18 | -0.013 | 0.002 | 3.17E-14 | Yes | No |
| rs7847351 | 9 | 79977312 | A | G | 0.81 | -0.013 | 0.002 | 4.95E-14 | Yes | No |
| rs12351997 | 9 | 80015424 | T | C | 0.8 | -0.013 | 0.002 | 4.50E-14 | Yes | Yes |
| rs61750929 | 9 | 91495135 | T | C | 0.06 | -0.028 | 0.003 | 9.49E-24 | Yes | No |
| rs7042939 | 9 | 110511408 | A | G | 0.4 | 0.01 | 0.001 | 1.50E-15 | Yes | Yes |
| rs1467311 | 9 | 110536932 | A | G | 0.66 | -0.011 | 0.001 | 2.26E-15 | Yes | Yes |
| rs649129 | 9 | 136154304 | T | C | 0.22 | 0.011 | 0.002 | 3.28E-15 | Yes | No |
| rs3829109 | 9 | 139256766 | A | G | 0.3 | -0.009 | 0.002 | 2.68E-08 | Yes | No |
| rs11257655 | 10 | 12307894 | T | C | 0.22 | 0.011 | 0.002 | 1.91E-13 | Yes | Yes |
| rs5785903 | 10 | 71002040 | T | TG | 0.43 | 0.017 | 0.002 | 4.39E-29 | No | No |
| rs4745982 | 10 | 71089843 | T | G | 0.92 | 0.074 | 0.003 | 8.36E-143 | Yes | Yes |
| rs150705486 | 10 | 71093216 | A | G | 0.02 | -0.114 | 0.006 | 4.24E-72 | Yes | No |
| rs17476364 | 10 | 71094504 | T | C | 0.9 | 0.086 | 0.002 | 1.33E-314 | Yes | No |
| rs72805692 | 10 | 71099109 | A | G | 0.89 | 0.079 | 0.002 | 8.52E-294 | Yes | No |
| rs200572185 | 10 | 71118821 | A | AAG | 0.04 | -0.094 | 0.004 | 1.53E-140 | No | No |
| rs7903146 | 10 | 114758349 | T | C | 0.28 | 0.013 | 0.001 | 1.04E-22 | Yes | No |
| rs4980325 | 11 | 234451 | T | G | 0.53 | 0.011 | 0.001 | 4.70E-14 | Yes | No |
| rs3842753 | 11 | 2181060 | T | G | 0.28 | 0.008 | 0.002 | 3.93E-08 | Yes | No |
| rs373894 | 11 | 9763094 | A | C | 0.72 | 0.008 | 0.002 | 5.28E-09 | Yes | Yes |
| rs360140 | 11 | 9776567 | A | C | 0.66 | -0.008 | 0.001 | 9.62E-13 | Yes | Yes |
| rs117706710 | 11 | 10508903 | T | G | 0.009 | 0.046 | 0.009 | 4.80E-08 | Yes | No |
| rs11039154 | 11 | 47278502 | T | C | 0.28 | -0.009 | 0.001 | 3.11E-09 | Yes | No |
| rs10838696 | 11 | 47363285 | A | G | 0.35 | -0.007 | 0.001 | 6.53E-09 | Yes | No |
| rs174559 | 11 | 61581656 | A | G | 0.27 | -0.011 | 0.001 | 3.31E-13 | Yes | No |
| rs174584 | 11 | 61610750 | A | G | 0.35 | -0.009 | 0.001 | 2.60E-10 | Yes | No |
| rs10466351 | 11 | 92697981 | T | C | 0.38 | 0.015 | 0.001 | 5.02E-26 | Yes | No |
| rs10830963 | 11 | 92708710 | C | G | 0.72 | -0.02 | 0.002 | 1.54E-36 | Yes | No |
| rs11224302 | 11 | 100456604 | T | C | 0.1 | -0.016 | 0.002 | 4.63E-14 | Yes | Yes |
| rs117233107 | 12 | 4328521 | A | G | 0.02 | -0.047 | 0.007 | 8.45E-11 | Yes | No |
| rs2110073 | 12 | 7075882 | T | C | 0.1 | 0.012 | 0.002 | 5.79E-09 | Yes | Yes |
| rs76261711 | 12 | 48486696 | A | T | 0.9 | 0.014 | 0.002 | 4.35E-10 | Yes | No |
| rs4760682 | 12 | 48512285 | A | C | 0.8 | 0.016 | 0.002 | 3.20E-20 | Yes | No |
| rs10774624 | 12 | 111833788 | A | G | 0.52 | 0.009 | 0.001 | 4.17E-14 | Yes | No |
| rs17696736 | 12 | 112486818 | A | G | 0.57 | 0.009 | 0.001 | 4.73E-12 | Yes | No |
| rs4766971 | 12 | 113018479 | A | G | 0.09 | 0.01 | 0.002 | 4.73E-08 | Yes | No |
| rs76533333 | 13 | 113352916 | A | G | 0.92 | -0.027 | 0.003 | 2.81E-29 | Yes | No |
| rs1278769 | 13 | 113536627 | A | G | 0.24 | -0.009 | 0.002 | 5.52E-12 | Yes | Yes |
| rs7994900 | 13 | 114553134 | T | C | 0.27 | 0.011 | 0.002 | 6.65E-14 | Yes | No |
| rs2273475 | 14 | 65268605 | A | G | 0.9 | -0.013 | 0.002 | 1.99E-09 | Yes | Yes |
| rs147727276 | 14 | 73601025 | A | G | 0.1 | -0.013 | 0.002 | 9.16E-10 | Yes | No |
| rs10151436 | 14 | 73616095 | A | T | 0.89 | 0.013 | 0.002 | 3.85E-11 | Yes | Yes |
| rs1535464 | 14 | 100793431 | A | G | 0.23 | -0.009 | 0.002 | 1.11E-08 | Yes | No |
| rs33952550 | 14 | 100798141 | C | CCTCA | 0.73 | 0.009 | 0.002 | 9.02E-09 | No | No |
| rs452306 | 15 | 65822777 | T | C | 0.59 | -0.01 | 0.001 | 5.51E-13 | Yes | No |
| rs368777 | 15 | 65849552 | A | G | 0.57 | -0.009 | 0.001 | 1.69E-12 | Yes | No |
| rs11248914 | 16 | 293562 | T | C | 0.66 | 0.011 | 0.001 | 1.42E-14 | Yes | Yes |
| rs9926463 | 16 | 11437607 | A | C | 0.31 | 0.008 | 0.002 | 4.01E-09 | Yes | No |
| rs11643024 | 16 | 11443183 | A | G | 0.31 | 0.008 | 0.002 | 7.98E-10 | Yes | No |
| rs4787458 | 16 | 28531287 | A | G | 0.62 | -0.008 | 0.001 | 1.16E-10 | Yes | No |
| rs7190771 | 16 | 28590030 | A | G | 0.38 | 0.009 | 0.001 | 6.02E-11 | No | No |
| rs7198799 | 16 | 68818390 | T | C | 0.29 | 0.008 | 0.001 | 4.76E-09 | Yes | Yes |
| rs1862748 | 16 | 68832943 | T | C | 0.3 | 0.008 | 0.001 | 8.35E-09 | Yes | Yes |
| rs4238685 | 16 | 88775220 | T | C | 0.1 | 0.006 | 0.002 | 0.00101 | Yes | Yes |
| rs837763 | 16 | 88853729 | T | C | 0.56 | 0.018 | 0.001 | 5.20E-38 | Yes | Yes |
| rs8075153 | 17 | 17622666 | T | C | 0.43 | -0.007 | 0.001 | 5.19E-08 | Yes | Yes |
| rs9914988 | 17 | 27183104 | A | G | 0.8 | 0.013 | 0.002 | 4.66E-17 | Yes | Yes |
| rs2748427 | 17 | 76121864 | A | G | 0.78 | -0.031 | 0.002 | 9.82E-49 | No | No |
| rs2748424 | 17 | 76124865 | C | G | 0.81 | -0.031 | 0.002 | 4.13E-39 | No | No |
| rs200350733 | 17 | 76125194 | A | AGC | 0.2 | 0.033 | 0.003 | 1.48E-40 | No | No |
| rs72634341 | 17 | 80682778 | A | G | 0.72 | -0.031 | 0.002 | 5.59E-97 | Yes | No |
| rs9909940 | 17 | 80689036 | T | C | 0.31 | 0.032 | 0.001 | 1.43E-116 | Yes | No |
| rs62076520 | 17 | 80695406 | A | G | 0.37 | -0.026 | 0.001 | 3.06E-82 | Yes | No |
| rs113373052 | 17 | 80697458 | T | C | 0.3 | 0.033 | 0.001 | 7.09E-113 | Yes | No |
| rs12452572 | 17 | 80702963 | A | G | 0.47 | 0.023 | 0.001 | 9.53E-67 | No | No |
| rs191734192 | 17 | 80778724 | A | G | 0.13 | 0.042 | 0.003 | 3.75E-66 | No | No |
| rs141786856 | 17 | 80950648 | C | CAAT | 0.04 | 0.021 | 0.005 | 8.52E-07 | No | No |
| rs28671200 | 18 | 43774444 | T | G | 0.68 | 0.009 | 0.002 | 1.56E-08 | No | No |
| rs12982956 | 19 | 17238413 | A | G | 0.72 | 0.007 | 0.002 | 3.26E-08 | Yes | No |
| rs17533945 | 19 | 17257802 | T | C | 0.6 | -0.013 | 0.001 | 1.62E-23 | Yes | Yes |
| rs12978547 | 19 | 33037212 | C | G | 0.97 | 0.028 | 0.004 | 3.96E-12 | Yes | Yes |
| rs10405535 | 19 | 33072085 | A | G | 0.3 | 0.012 | 0.002 | 6.47E-14 | Yes | No |
| rs4499344 | 19 | 33073431 | A | G | 0.31 | -0.011 | 0.002 | 1.35E-12 | Yes | Yes |
| rs737092 | 20 | 55990405 | T | C | 0.51 | -0.007 | 0.001 | 7.57E-09 | Yes | Yes |
| rs855791 | 22 | 37462936 | A | G | 0.43 | 0.019 | 0.001 | 1.34E-56 | Yes | Yes |
| rs2143923 | 22 | 43141907 | A | G | 0.48 | 0.007 | 0.001 | 3.97E-08 | Yes | No |
| rs184506746 | X | 150104927 | T | C | 0.013 | -0.171 | 0.028 | 2.46E-08 | No | No |
| rs182859029 | X | 152051223 | T | C | 0.99 | 0.246 | 0.038 | 8.03E-10 | No | No |
| rs138245006 | X | 153018275 | T | C | 0.97 | 0.251 | 0.015 | 1.98E-58 | No | No |
| rs140801471 | X | 153542572 | A | G | 0.05 | -0.082 | 0.007 | 6.42E-31 | No | No |
| rs149309160 | X | 154212603 | T | G | 0.95 | 0.125 | 0.008 | 2.36E-47 | No | No |

**Supplemental Table 3.** SNPs included in fasting insulin-related analyses.

| RSID | Chrom. | Position | Effect allele | Other allele | Effect allele freq. | Beta | SE | p-value | Used in MR | Used in MR + Steiger filt. |
| --- | --- | --- | --- | --- | --- | --- | --- | --- | --- | --- |
| rs6674544 | 1 | 219628973 | A | G | 0.57 | 0.018 | 0.002 | 6.97E-21 | No | No |
| rs77935490 | 2 | 630902 | A | T | 0.21 | 0.014 | 0.003 | 1.07E-08 | No | No |
| rs1260326 | 2 | 27730940 | T | C | 0.39 | -0.023 | 0.002 | 8.42E-38 | Yes | No |
| rs6713419 | 2 | 165508300 | T | C | 0.63 | 0.019 | 0.002 | 1.44E-24 | Yes | Yes |
| rs75265117 | 2 | 165518799 | C | G | 0.88 | 0.028 | 0.003 | 1.54E-24 | Yes | No |
| rs1128249 | 2 | 165528624 | T | G | 0.39 | -0.02 | 0.002 | 7.71E-28 | Yes | Yes |
| rs13389219 | 2 | 165528876 | T | C | 0.39 | -0.02 | 0.002 | 5.84E-28 | Yes | Yes |
| rs2943646 | 2 | 227099534 | A | G | 0.37 | -0.025 | 0.002 | 8.47E-39 | Yes | Yes |
| rs2972145 | 2 | 227101309 | T | C | 0.37 | -0.025 | 0.002 | 6.08E-38 | Yes | No |
| rs308971 | 3 | 12116620 | A | G | 0.87 | -0.022 | 0.003 | 3.91E-13 | Yes | Yes |
| rs11712037 | 3 | 12344730 | C | G | 0.87 | 0.028 | 0.003 | 2.10E-21 | Yes | No |
| rs35000407 | 3 | 12351521 | T | G | 0.86 | 0.026 | 0.003 | 1.50E-21 | Yes | No |
| rs9819511 | 3 | 49884261 | T | C | 0.3 | 0.014 | 0.002 | 2.26E-08 | Yes | No |
| rs17331151 | 3 | 52844534 | T | C | 0.11 | -0.016 | 0.003 | 1.52E-08 | Yes | Yes |
| rs11708067 | 3 | 123065778 | A | G | 0.78 | -0.014 | 0.002 | 1.30E-09 | Yes | No |
| rs62271373 | 3 | 150066540 | A | T | 0.06 | 0.026 | 0.005 | 1.60E-08 | Yes | No |
| rs2276936 | 4 | 89726283 | A | C | 0.5 | -0.012 | 0.002 | 4.25E-11 | Yes | Yes |
| rs3775380 | 4 | 89739808 | A | G | 0.5 | -0.012 | 0.002 | 1.48E-11 | Yes | Yes |
| rs9884482 | 4 | 106081636 | T | C | 0.61 | -0.013 | 0.002 | 2.88E-11 | Yes | Yes |
| rs10010325 | 4 | 106106353 | A | C | 0.48 | 0.012 | 0.002 | 4.05E-11 | Yes | Yes |
| rs11727676 | 4 | 145659064 | T | C | 0.91 | -0.02 | 0.004 | 2.90E-08 | Yes | Yes |
| rs6855363 | 4 | 157670537 | T | C | 0.68 | 0.013 | 0.002 | 4.04E-08 | Yes | Yes |
| rs4865796 | 5 | 53272664 | A | G | 0.68 | 0.017 | 0.002 | 7.33E-17 | Yes | Yes |
| rs459193 | 5 | 55806751 | A | G | 0.27 | -0.018 | 0.002 | 1.12E-18 | Yes | Yes |
| rs3936511 | 5 | 55860781 | A | G | 0.82 | -0.019 | 0.003 | 2.81E-14 | Yes | Yes |
| rs116141873 | 6 | 34222201 | T | G | 0.04 | 0.043 | 0.006 | 1.42E-11 | Yes | No |
| rs2780215 | 6 | 34236973 | A | G | 0.96 | 0.039 | 0.006 | 1.06E-09 | No | No |
| rs998584 | 6 | 43757896 | A | C | 0.49 | 0.012 | 0.002 | 2.31E-10 | Yes | No |
| rs9472135 | 6 | 43809802 | T | C | 0.7 | 0.011 | 0.002 | 4.21E-08 | Yes | Yes |
| rs1474696 | 6 | 127449246 | A | G | 0.49 | -0.015 | 0.002 | 3.02E-16 | Yes | No |
| rs2745353 | 6 | 127452935 | T | C | 0.52 | 0.015 | 0.002 | 4.63E-16 | Yes | Yes |
| rs73013411 | 6 | 164126233 | A | C | 0.13 | -0.018 | 0.003 | 2.08E-08 | Yes | No |
| rs4709746 | 6 | 164133001 | T | C | 0.13 | -0.018 | 0.003 | 2.97E-08 | Yes | No |
| rs2108349 | 7 | 50786663 | A | G | 0.66 | -0.012 | 0.002 | 1.13E-08 | Yes | No |
| rs13234269 | 7 | 130429186 | A | T | 0.48 | -0.011 | 0.002 | 1.27E-08 | No | No |
| rs972283 | 7 | 130466854 | A | G | 0.47 | -0.011 | 0.002 | 1.09E-08 | Yes | No |
| rs330945 | 8 | 9021933 | T | C | 0.63 | 0.014 | 0.002 | 1.82E-11 | Yes | No |
| rs7012637 | 8 | 9173209 | A | G | 0.48 | -0.022 | 0.002 | 1.48E-29 | Yes | No |
| rs7012814 | 8 | 9173358 | A | G | 0.48 | -0.022 | 0.002 | 8.34E-30 | Yes | No |
| rs4841132 | 8 | 9183596 | A | G | 0.11 | 0.026 | 0.003 | 3.83E-20 | Yes | No |
| rs13258890 | 8 | 23615445 | T | C | 0.77 | 0.013 | 0.003 | 2.77E-08 | Yes | Yes |
| rs75179845 | 9 | 136132954 | T | C | 0.91 | -0.022 | 0.004 | 6.05E-11 | Yes | No |
| rs8176693 | 9 | 136137657 | T | C | 0.1 | 0.02 | 0.003 | 1.10E-10 | Yes | Yes |
| rs118164457 | 10 | 89680631 | T | C | 0.96 | -0.035 | 0.006 | 3.86E-10 | Yes | No |
| rs12769346 | 10 | 89764490 | T | G | 0.86 | -0.015 | 0.003 | 1.20E-08 | Yes | Yes |
| rs7903146 | 10 | 114758349 | T | C | 0.27 | -0.012 | 0.002 | 1.24E-09 | Yes | No |
| rs2845885 | 11 | 63869062 | T | C | 0.93 | -0.02 | 0.004 | 1.18E-08 | Yes | Yes |
| rs2054435 | 12 | 21696684 | A | G | 0.22 | -0.015 | 0.003 | 9.50E-09 | Yes | Yes |
| rs6487237 | 12 | 21699928 | A | C | 0.78 | 0.015 | 0.003 | 4.68E-09 | Yes | Yes |
| rs111264094 | 12 | 48202696 | C | G | 0.97 | 0.057 | 0.009 | 1.64E-09 | Yes | No |
| rs1351394 | 12 | 66351826 | T | C | 0.49 | -0.011 | 0.002 | 2.71E-09 | Yes | Yes |
| rs7968682 | 12 | 66371880 | T | G | 0.51 | 0.011 | 0.002 | 3.17E-09 | Yes | Yes |
| rs1402013 | 12 | 102740822 | A | G | 0.35 | -0.009 | 0.002 | 1.31E-06 | Yes | Yes |
| rs860598 | 12 | 102898446 | A | G | 0.82 | 0.018 | 0.003 | 6.88E-12 | Yes | Yes |
| rs35747 | 12 | 102912558 | A | G | 0.82 | 0.017 | 0.003 | 2.68E-11 | Yes | Yes |
| rs7133378 | 12 | 124409502 | A | G | 0.32 | -0.013 | 0.002 | 6.00E-11 | Yes | No |
| rs7975482 | 12 | 124481690 | A | G | 0.67 | 0.012 | 0.002 | 1.97E-10 | Yes | No |
| rs12454712 | 18 | 60845884 | T | C | 0.58 | 0.014 | 0.003 | 1.78E-09 | No | No |
| rs10422861 | 19 | 33894846 | T | C | 0.67 | -0.013 | 0.002 | 6.65E-11 | Yes | No |
| rs731839 | 19 | 33899065 | A | G | 0.66 | -0.012 | 0.002 | 3.87E-11 | Yes | Yes |
| rs1999536 | 20 | 45581777 | C | G | 0.55 | -0.012 | 0.002 | 1.32E-09 | No | No |
| rs1206760 | 20 | 45582472 | A | G | 0.54 | -0.011 | 0.002 | 8.82E-10 | Yes | Yes |

**Supplemental Table 4.**
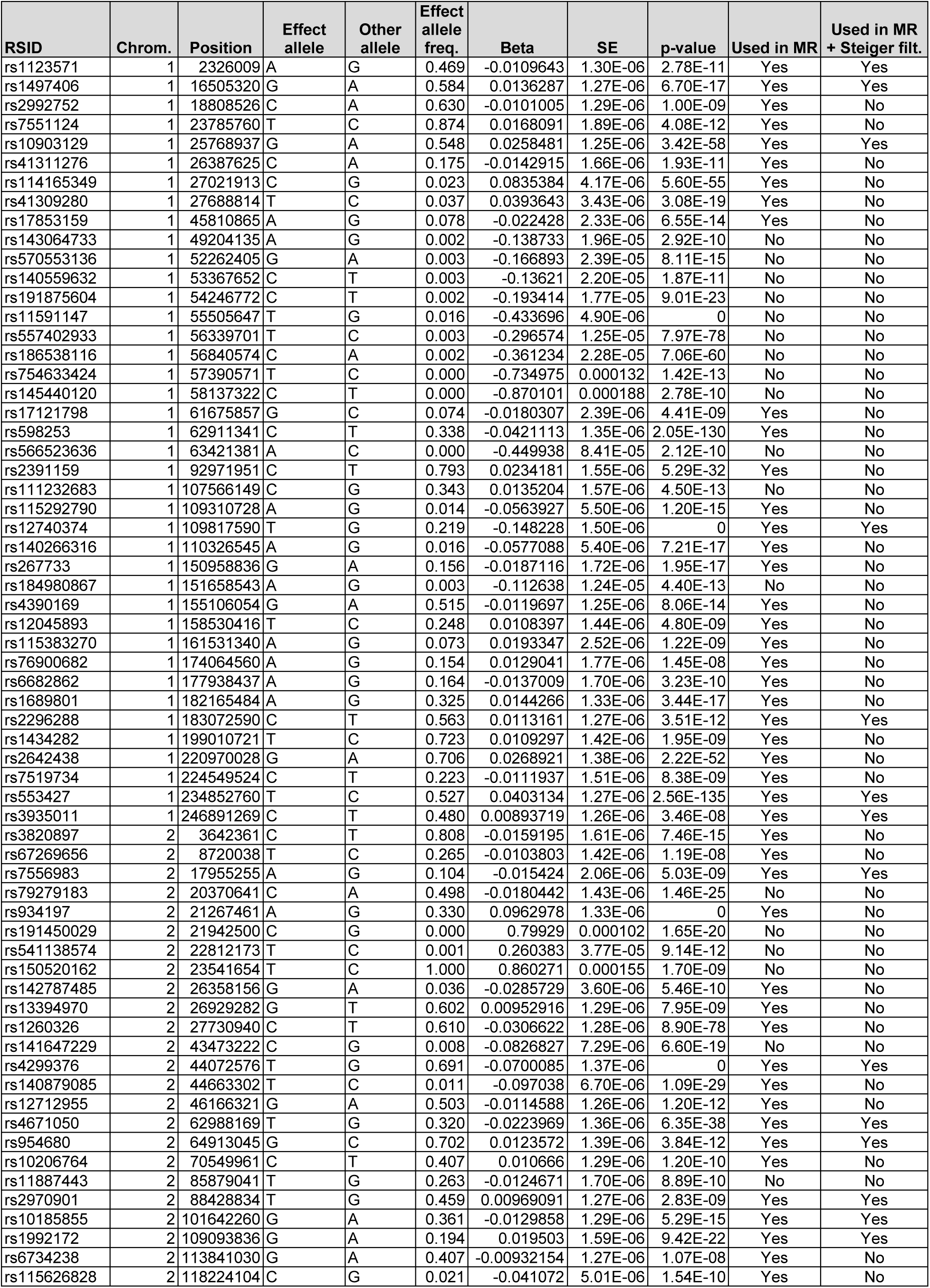
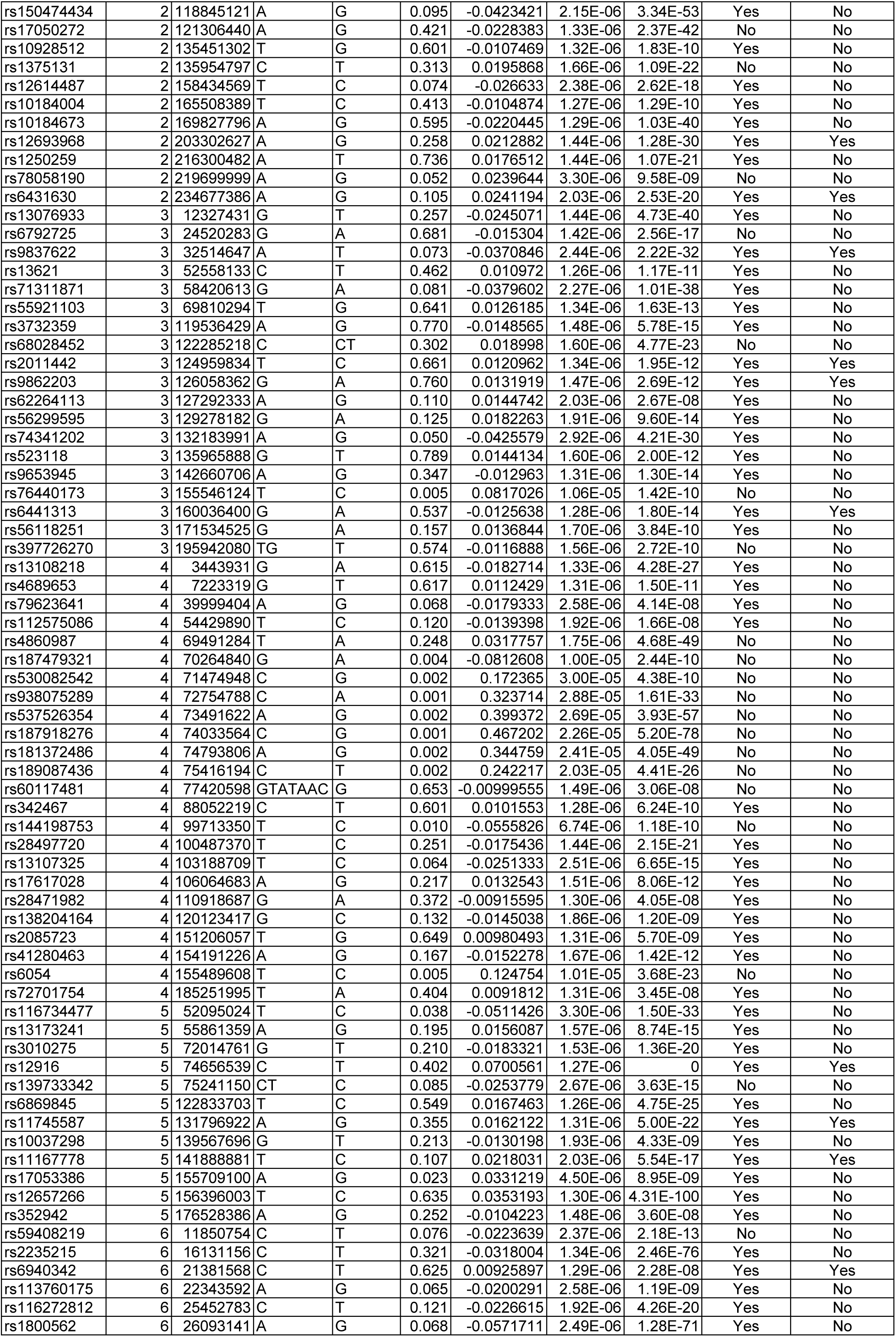
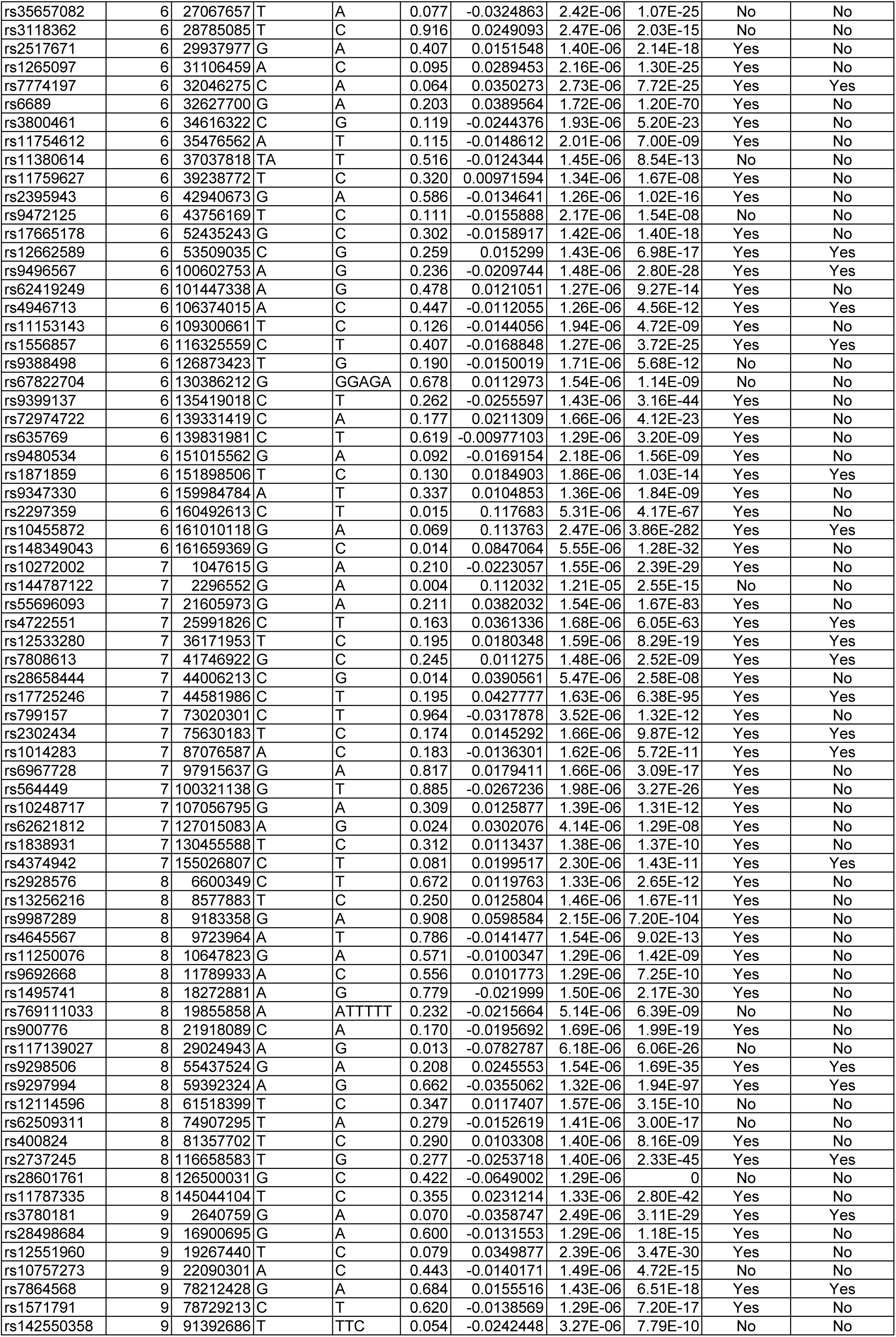
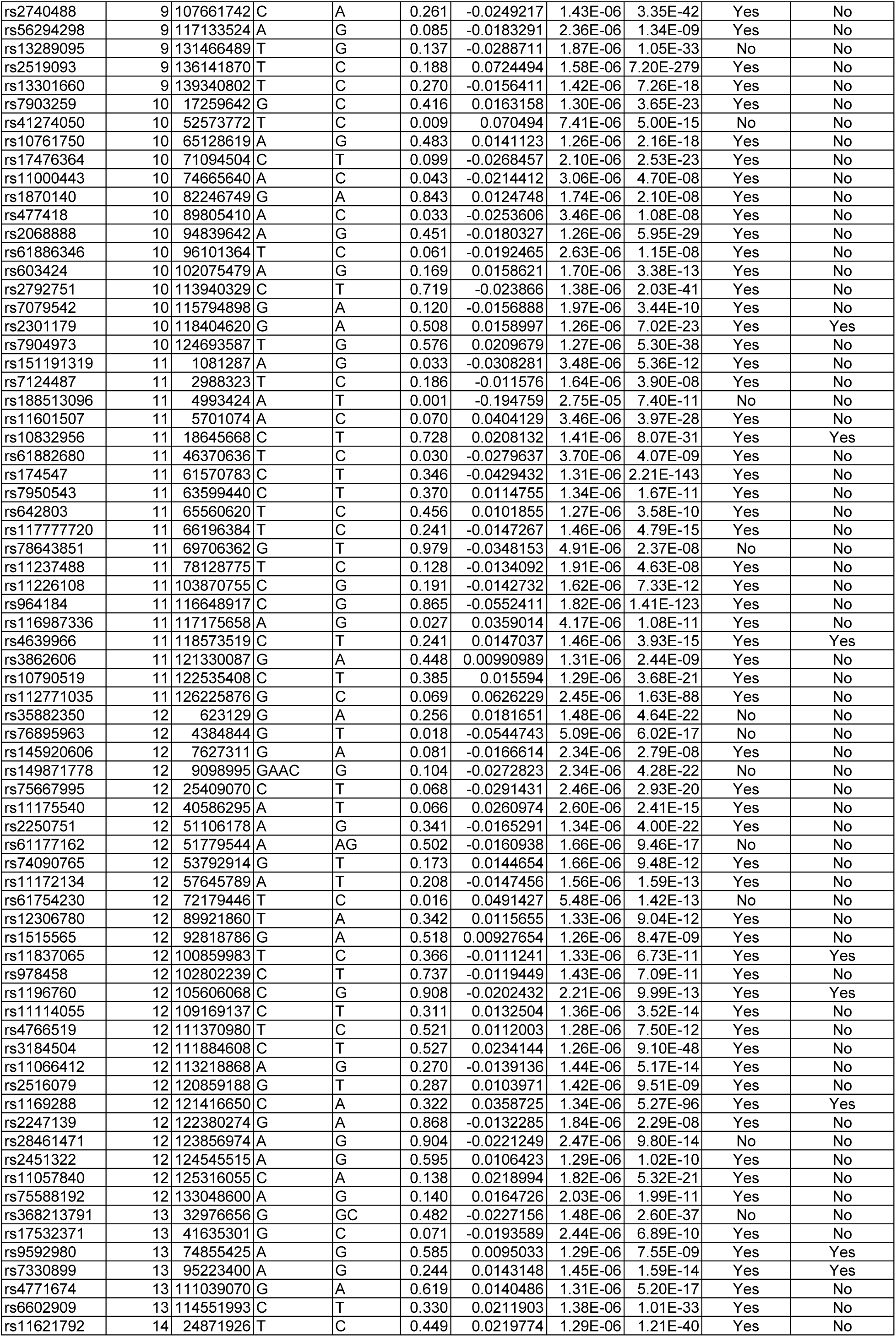
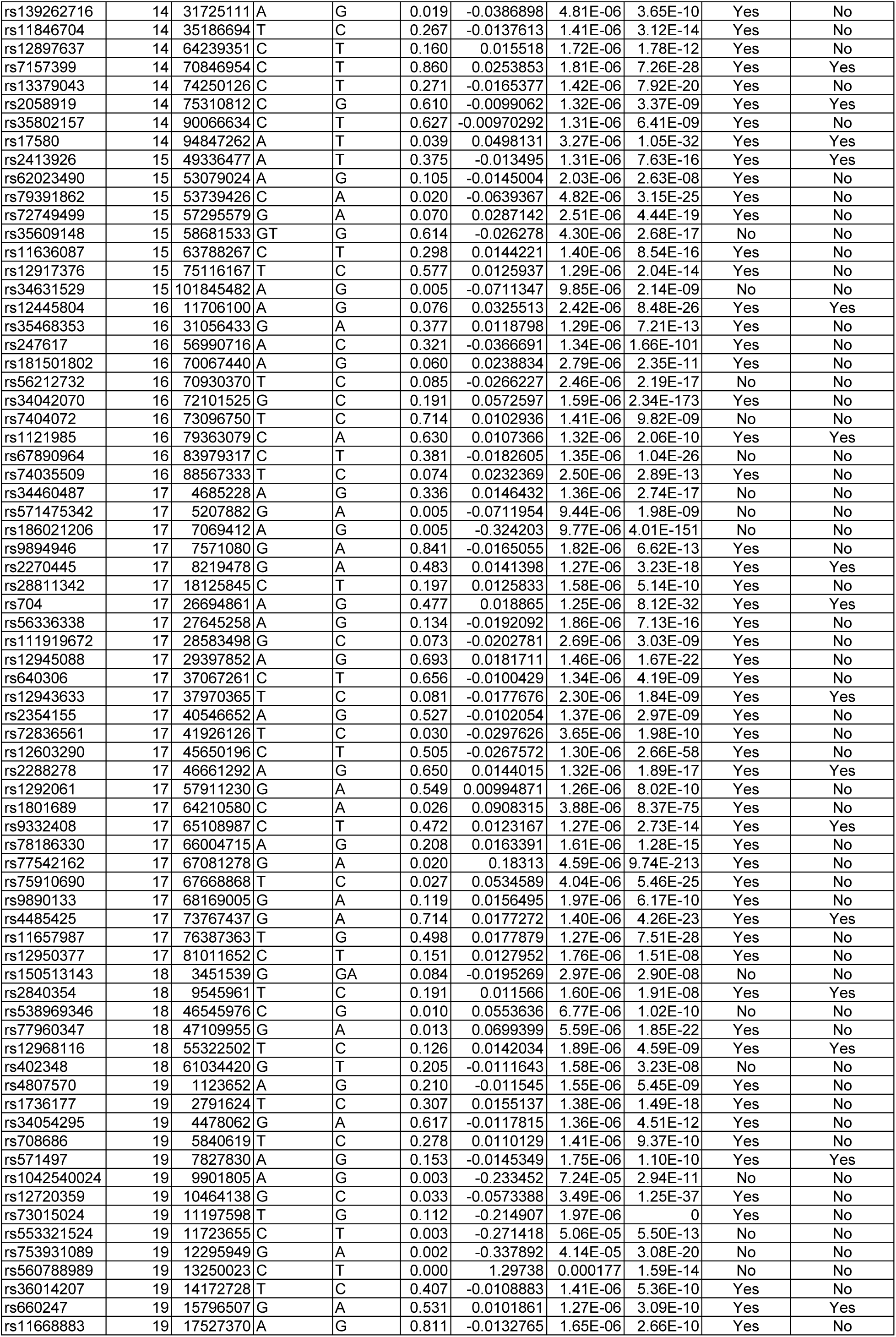
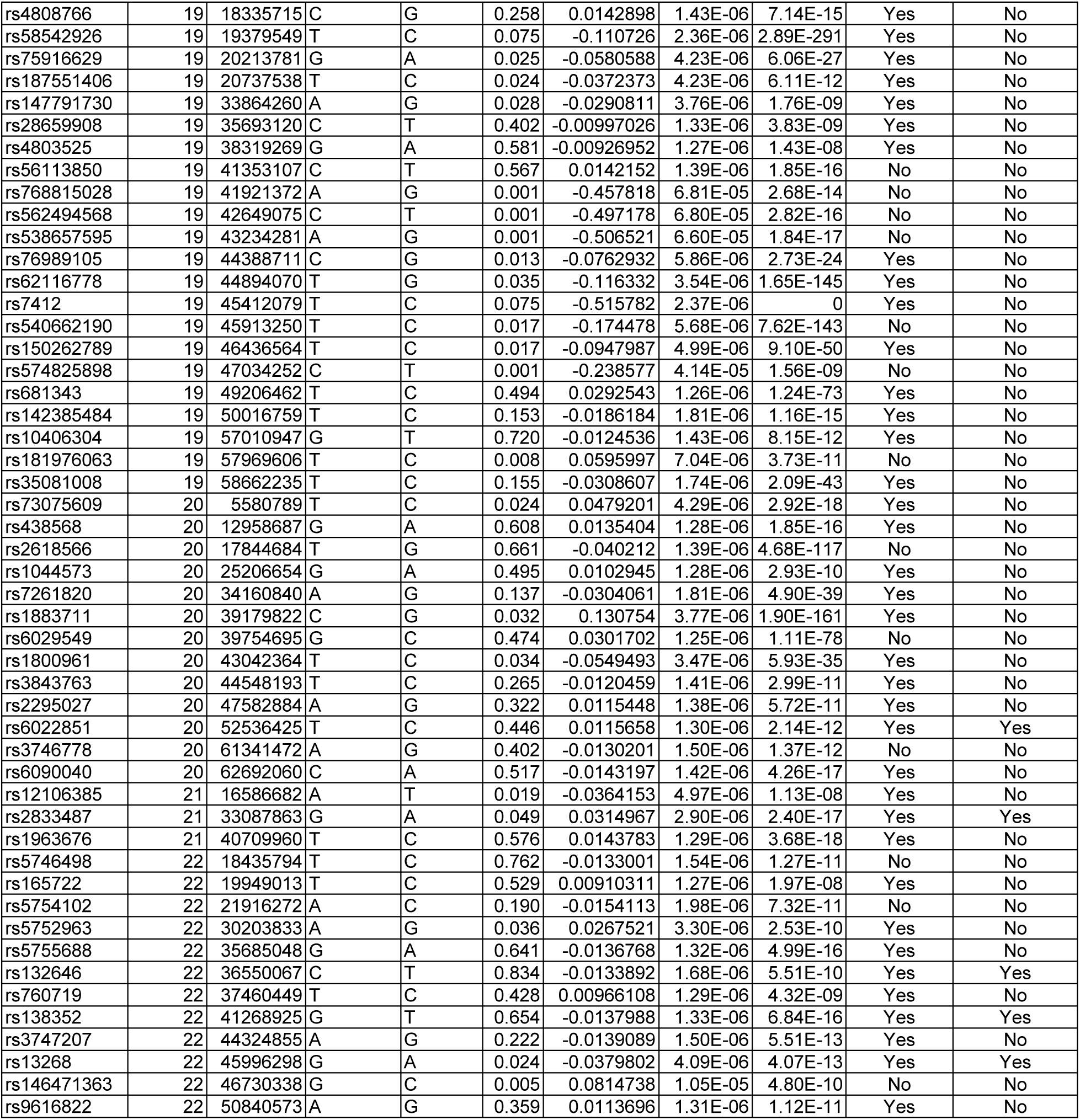
SNPs included in LDL cholesterol-related analyses.

**Supplemental Table 5.**
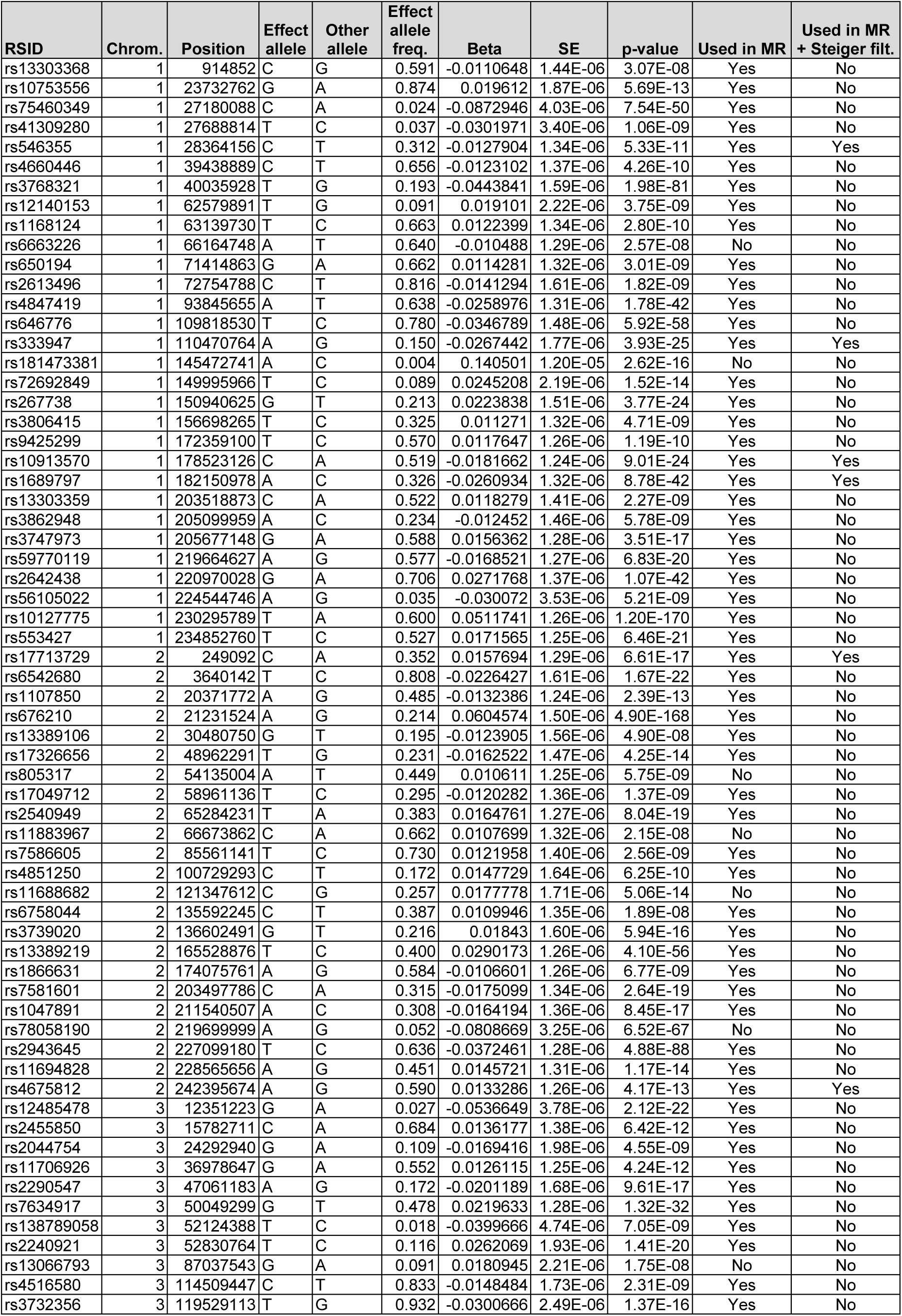
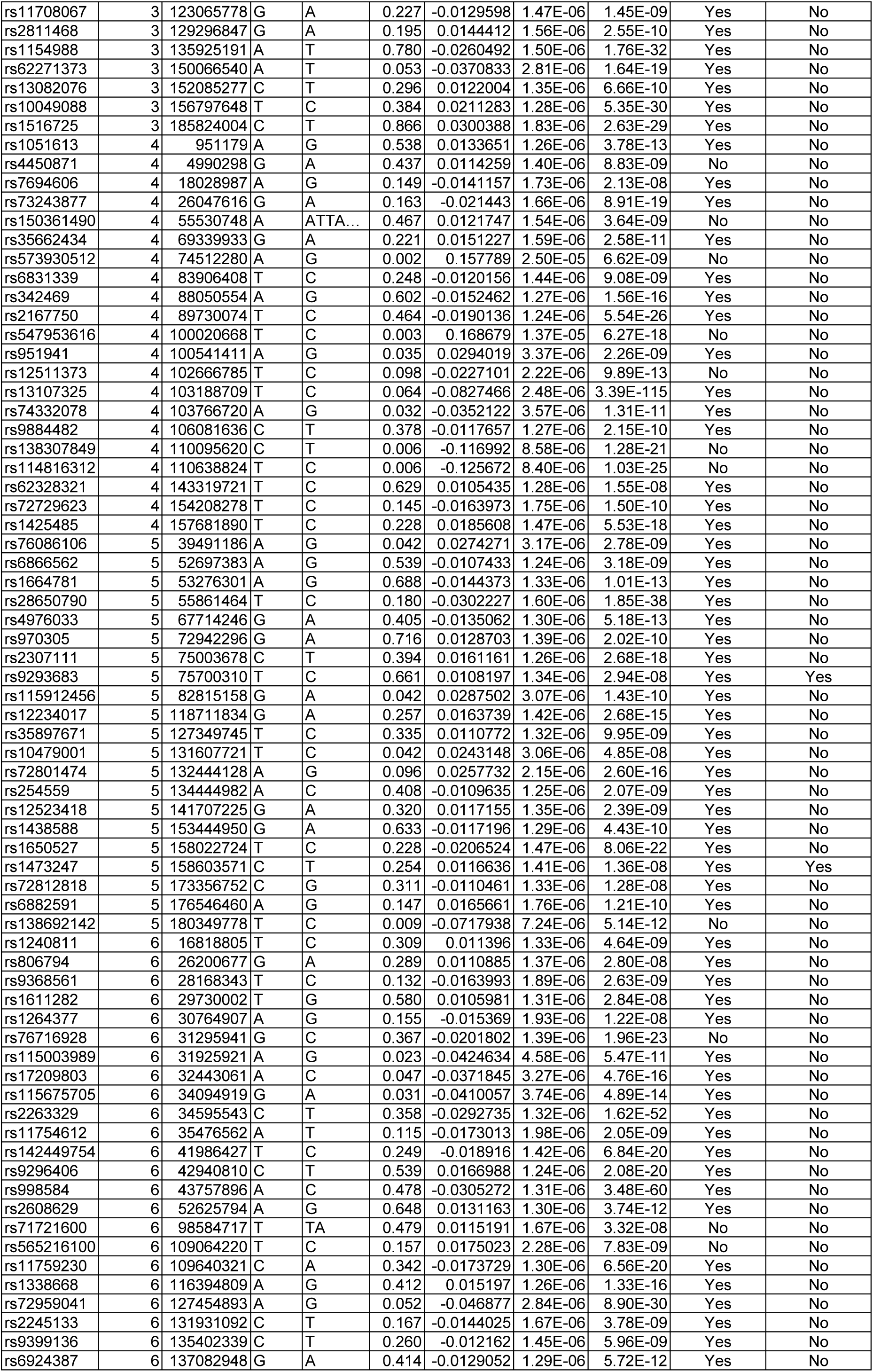
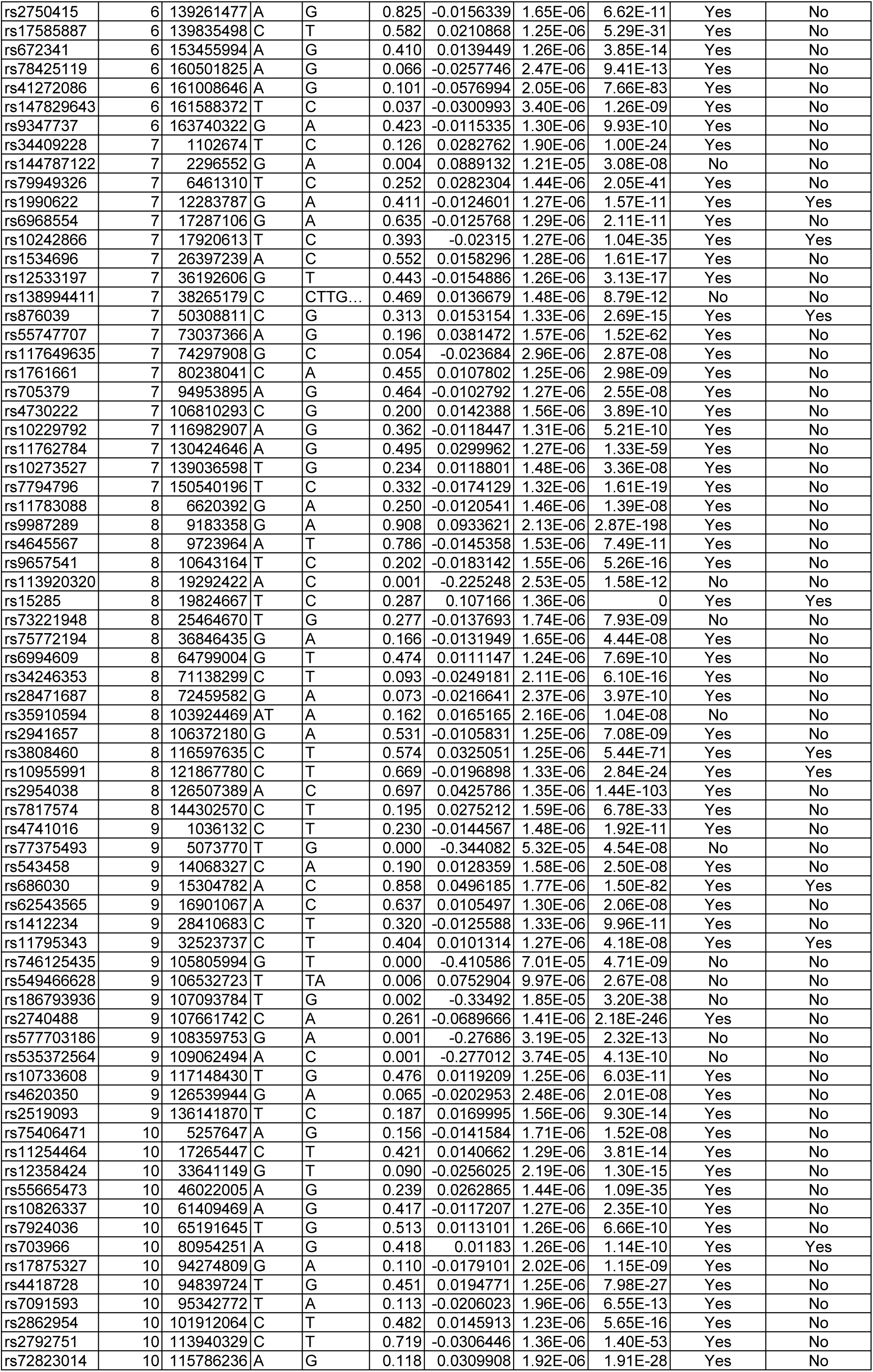
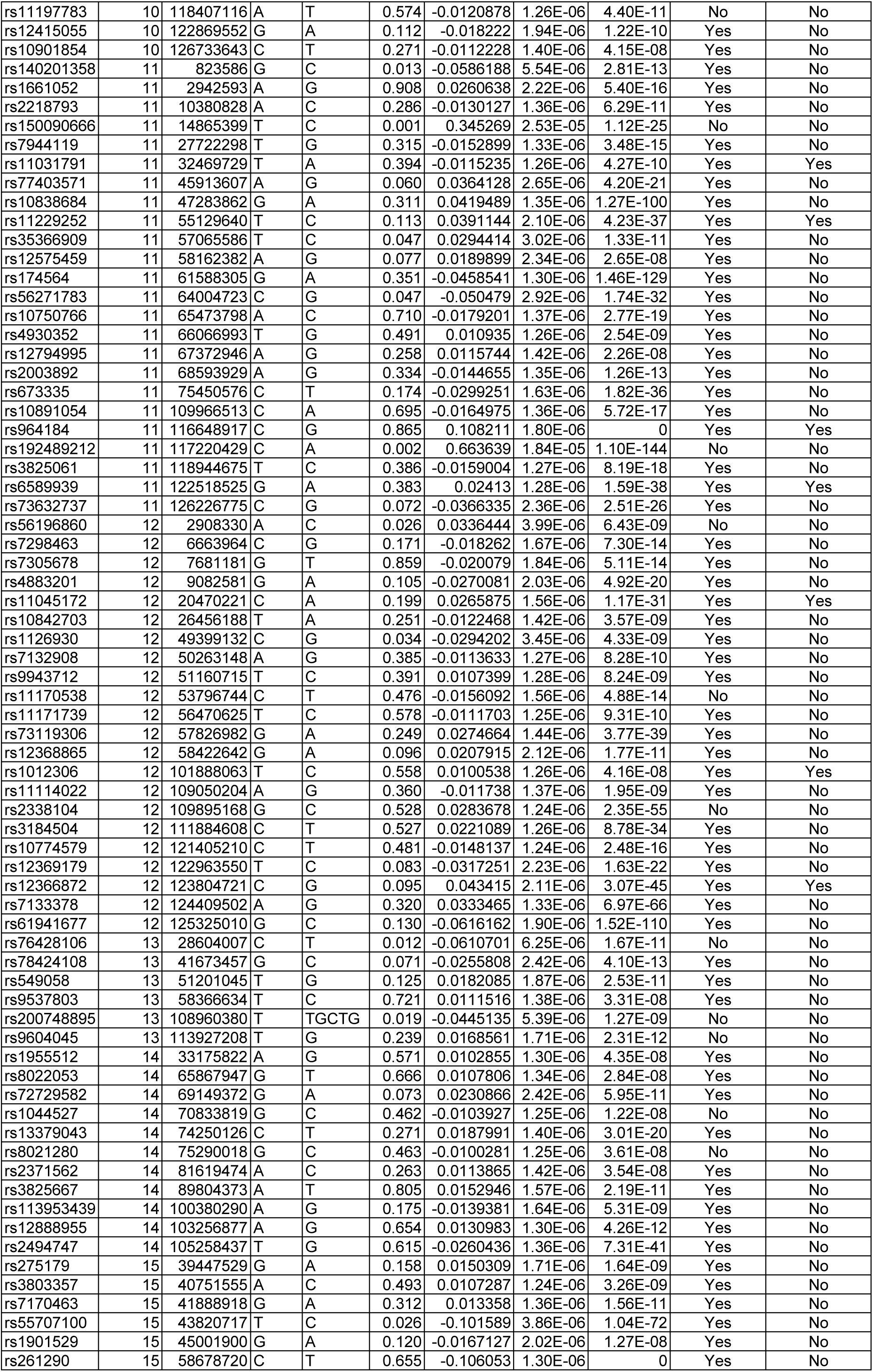
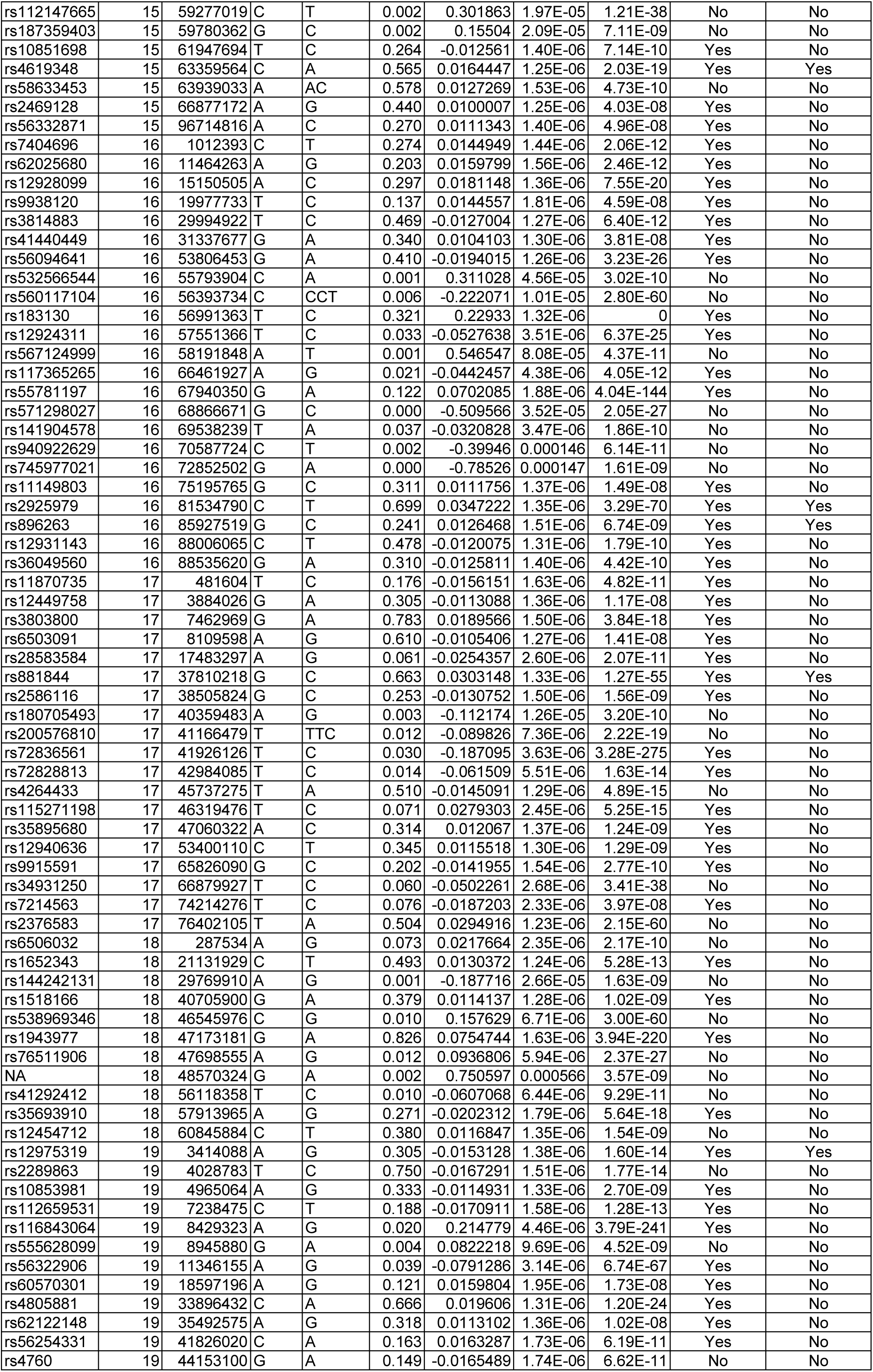
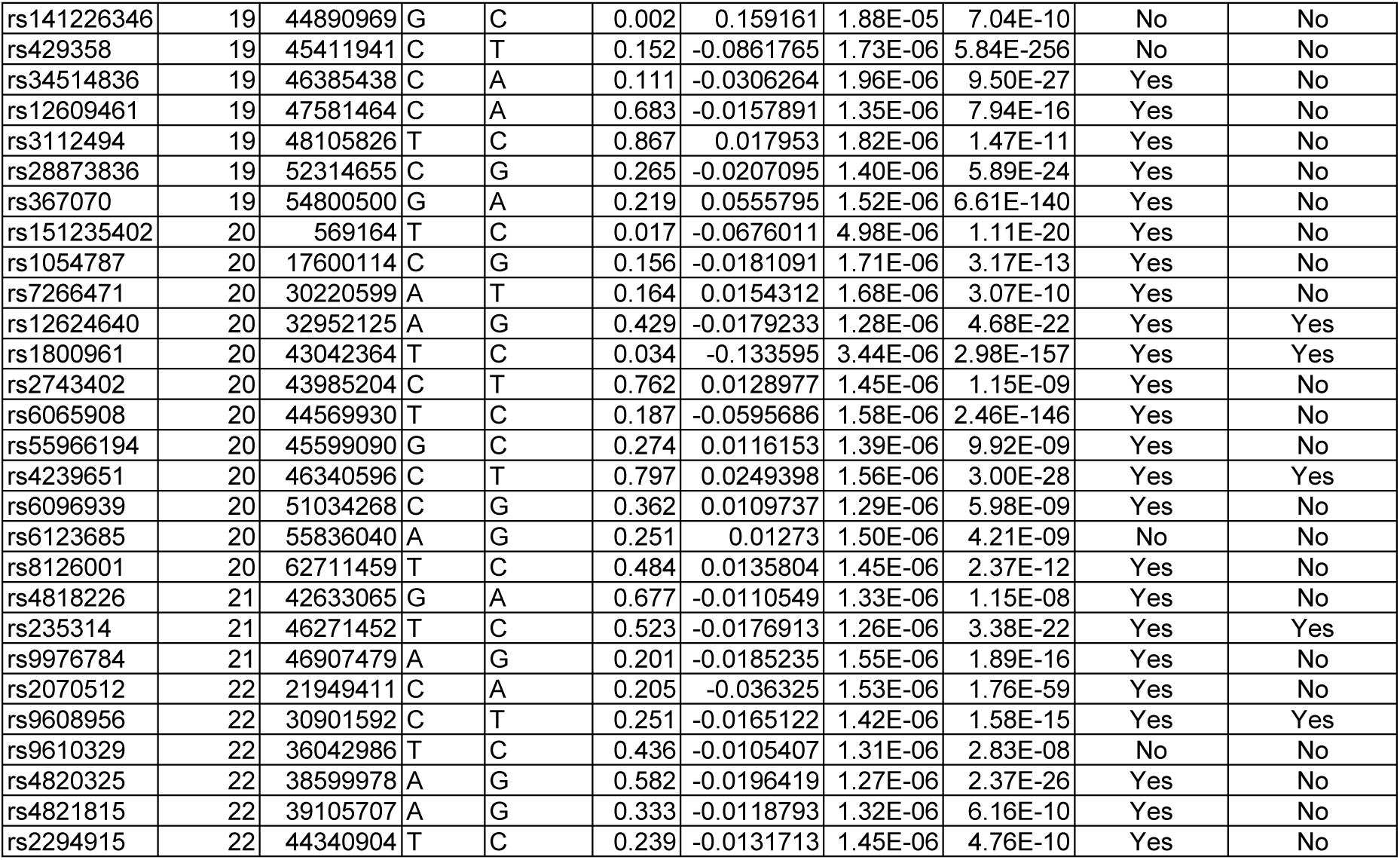
SNPs included in HDL cholesterol-related analyses.

**Supplemental Table 6.** SNPs included in triglyceride-related analyses.

| RSID | Chrom. | Position | Effect allele | Other allele | Effect allele freq. | Beta | SE | p-value | Used in MR | Used in MR + Steiger filt. |
| --- | --- | --- | --- | --- | --- | --- | --- | --- | --- | --- |
| rs6605083 | 1 | 2147162 | C | T | 0.233 | 0.0119684 | 1.49E-06 | 6.45E-09 | Yes | No |
| rs7538833 | 1 | 16504381 | C | T | 0.628 | 0.0111822 | 1.27E-06 | 2.58E-10 | Yes | No |
| rs7551124 | 1 | 23785760 | T | C | 0.874 | 0.0174888 | 1.85E-06 | 1.09E-11 | Yes | No |
| rs213641 | 1 | 26232356 | A | C | 0.593 | 0.00960533 | 1.27E-06 | 4.97E-08 | Yes | No |
| rs114165349 | 1 | 27021913 | C | G | 0.023 | 0.0684903 | 4.08E-06 | 1.81E-33 | Yes | No |
| rs4660690 | 1 | 39819503 | A | G | 0.215 | 0.0206177 | 1.49E-06 | 2.92E-23 | Yes | No |
| rs61781290 | 1 | 40393160 | G | A | 0.262 | 0.0132882 | 1.39E-06 | 5.97E-12 | Yes | No |
| rs12078980 | 1 | 51508526 | C | T | 0.092 | -0.0222738 | 2.13E-06 | 5.86E-14 | Yes | No |
| rs1883783 | 1 | 54890956 | G | T | 0.581 | 0.0137463 | 1.26E-06 | 2.83E-15 | Yes | Yes |
| rs188503412 | 1 | 61680217 | G | A | 0.001 | -0.362757 | 4.98E-05 | 1.98E-14 | No | No |
| rs10889333 | 1 | 62957030 | A | G | 0.334 | -0.0713472 | 1.31E-06 | 0 | Yes | Yes |
| rs574313788 | 1 | 63679932 | A | C | 1.000 | 0.493268 | 7.95E-05 | 3.71E-11 | No | No |
| rs2613505 | 1 | 72835410 | T | C | 0.811 | 0.0139303 | 1.58E-06 | 2.54E-10 | Yes | No |
| rs165316 | 1 | 91533297 | G | A | 0.200 | -0.0132725 | 1.53E-06 | 4.26E-10 | Yes | No |
| rs12133907 | 1 | 93850283 | A | C | 0.609 | 0.0118584 | 1.28E-06 | 1.85E-11 | Yes | No |
| rs4378243 | 1 | 98395881 | T | G | 0.826 | -0.0147155 | 1.64E-06 | 9.63E-11 | Yes | No |
| rs646776 | 1 | 109818530 | T | C | 0.780 | 0.0113777 | 1.47E-06 | 2.53E-08 | Yes | No |
| rs10494363 | 1 | 149909495 | A | G | 0.075 | -0.0269128 | 2.33E-06 | 9.74E-17 | Yes | No |
| rs12750321 | 1 | 154251626 | G | A | 0.292 | -0.0148669 | 1.37E-06 | 4.59E-15 | Yes | No |
| rs7539464 | 1 | 172359576 | A | T | 0.570 | -0.0131357 | 1.24E-06 | 2.69E-14 | No | No |
| rs115276619 | 1 | 184865132 | A | T | 0.016 | -0.0497207 | 5.06E-06 | 1.61E-12 | Yes | No |
| rs2821228 | 1 | 203517723 | G | C | 0.537 | -0.0133718 | 1.35E-06 | 3.12E-13 | No | No |
| rs11240358 | 1 | 205070573 | A | G | 0.397 | 0.013375 | 1.26E-06 | 2.03E-14 | Yes | Yes |
| rs1337101 | 1 | 219726100 | T | G | 0.313 | -0.020048 | 1.35E-06 | 5.19E-27 | Yes | No |
| rs2642438 | 1 | 220970028 | G | A | 0.706 | -0.0157453 | 1.35E-06 | 4.56E-17 | Yes | No |
| rs2281721 | 1 | 230297136 | T | C | 0.610 | -0.0444727 | 1.25E-06 | 3.36E-144 | Yes | No |
| rs75054794 | 1 | 230823363 | G | A | 0.052 | 0.0232243 | 2.81E-06 | 2.78E-09 | Yes | No |
| rs17713879 | 2 | 254215 | A | G | 0.368 | -0.0111716 | 1.28E-06 | 3.04E-10 | Yes | No |
| rs3820897 | 2 | 3642361 | C | T | 0.807 | 0.0161779 | 1.57E-06 | 1.03E-13 | Yes | No |
| rs12478327 | 2 | 20374249 | C | A | 0.487 | -0.0171517 | 1.23E-06 | 1.00E-23 | Yes | No |
| rs676210 | 2 | 21231524 | A | G | 0.214 | -0.0704109 | 1.48E-06 | 1.16E-254 | Yes | No |
| rs62128360 | 2 | 26309420 | G | T | 0.070 | 0.0205154 | 2.40E-06 | 8.38E-10 | Yes | No |
| rs57035494 | 2 | 26924018 | G | C | 0.038 | -0.0383752 | 3.23E-06 | 1.15E-17 | Yes | No |
| rs1260326 | 2 | 27730940 | C | T | 0.609 | -0.110708 | 1.25E-06 | 0 | Yes | No |
| rs72780006 | 2 | 28599749 | A | G | 0.099 | 0.0343261 | 2.27E-06 | 1.09E-27 | No | No |
| rs72616903 | 2 | 43743012 | C | T | 0.165 | 0.0146626 | 1.67E-06 | 2.74E-10 | Yes | No |
| rs17326656 | 2 | 48962291 | T | G | 0.231 | 0.0126477 | 1.46E-06 | 5.06E-10 | Yes | No |
| rs1861410 | 2 | 58933591 | T | C | 0.551 | -0.0108161 | 1.24E-06 | 3.52E-10 | Yes | No |
| rs3087898 | 2 | 61765074 | A | G | 0.424 | -0.00993216 | 1.24E-06 | 9.21E-09 | Yes | No |
| rs2540949 | 2 | 65284231 | T | A | 0.383 | -0.019884 | 1.26E-06 | 1.01E-29 | Yes | No |
| rs939136 | 2 | 66677531 | C | G | 0.662 | -0.0100461 | 1.31E-06 | 3.41E-08 | No | No |
| rs6708784 | 2 | 111927379 | G | A | 0.496 | -0.0131009 | 1.24E-06 | 2.38E-14 | Yes | Yes |
| rs13409360 | 2 | 113838102 | A | G | 0.398 | -0.0102795 | 1.26E-06 | 4.10E-09 | Yes | No |
| rs78198921 | 2 | 119749146 | G | C | 0.014 | 0.0436598 | 5.22E-06 | 1.79E-09 | Yes | No |
| rs7578604 | 2 | 121308660 | T | G | 0.248 | 0.0161417 | 1.43E-06 | 4.29E-16 | Yes | No |
| rs6430090 | 2 | 146347459 | G | A | 0.598 | -0.0104499 | 1.26E-06 | 2.33E-09 | Yes | No |
| rs6743795 | 2 | 161122134 | G | A | 0.723 | 0.0112246 | 1.38E-06 | 5.16E-09 | Yes | No |
| rs13389219 | 2 | 165528876 | T | C | 0.400 | -0.0335082 | 1.25E-06 | 5.27E-83 | Yes | No |
| rs12472667 | 2 | 171629063 | G | C | 0.361 | 0.0100003 | 1.31E-06 | 2.90E-08 | Yes | No |
| rs148566631 | 2 | 203447875 | A | G | 0.117 | 0.0177558 | 1.91E-06 | 2.26E-11 | Yes | No |
| rs78058190 | 2 | 219699999 | A | G | 0.052 | 0.0766795 | 3.20E-06 | 9.79E-68 | No | No |
| rs1024137 | 2 | 227095876 | T | G | 0.637 | 0.0342883 | 1.27E-06 | 7.77E-84 | Yes | No |
| rs79639690 | 2 | 227605895 | C | T | 0.046 | 0.0236798 | 2.97E-06 | 1.00E-08 | Yes | No |
| rs9812100 | 3 | 4763301 | A | G | 0.471 | -0.0117735 | 1.24E-06 | 6.21E-12 | Yes | No |
| rs62246311 | 3 | 9498143 | A | G | 0.102 | 0.0162634 | 2.06E-06 | 9.90E-09 | Yes | No |
| rs1801282 | 3 | 12393125 | G | C | 0.125 | -0.0270379 | 1.84E-06 | 4.85E-26 | Yes | No |
| rs6792725 | 3 | 24520283 | G | A | 0.681 | -0.0159417 | 1.39E-06 | 1.09E-16 | No | No |
| rs144323903 | 3 | 36981990 | CA | C | 0.508 | -0.0132743 | 1.62E-06 | 6.36E-11 | No | No |
| rs4955411 | 3 | 49145304 | G | A | 0.776 | 0.0137496 | 1.51E-06 | 3.28E-11 | No | No |
| rs17052061 | 3 | 52346240 | G | T | 0.169 | -0.0200737 | 1.64E-06 | 1.19E-18 | Yes | No |
| rs12486424 | 3 | 66874481 | C | A | 0.289 | 0.0118231 | 1.35E-06 | 3.51E-10 | Yes | No |
| rs13066793 | 3 | 87037543 | G | A | 0.091 | -0.0175987 | 2.18E-06 | 6.29E-09 | No | No |
| rs684773 | 3 | 135956305 | C | A | 0.791 | 0.0253918 | 1.56E-06 | 1.79E-31 | Yes | No |
| rs111834982 | 3 | 142019751 | G | A | 0.129 | -0.0142007 | 1.84E-06 | 2.83E-08 | Yes | No |
| rs9653945 | 3 | 142660706 | A | G | 0.347 | -0.0131346 | 1.28E-06 | 1.97E-13 | Yes | No |
| rs62271373 | 3 | 150066540 | A | T | 0.053 | 0.035005 | 2.80E-06 | 2.19E-19 | Yes | No |
| rs6787781 | 3 | 155535351 | C | G | 0.270 | 0.0127098 | 1.42E-06 | 6.65E-11 | Yes | No |
| rs9817452 | 3 | 156795414 | T | G | 0.383 | -0.0178823 | 1.28E-06 | 7.61E-24 | Yes | No |
| rs11924648 | 3 | 170717996 | G | A | 0.128 | 0.020145 | 1.81E-06 | 1.49E-15 | Yes | No |
| rs79287178 | 3 | 172294500 | A | G | 0.029 | 0.0601879 | 3.77E-06 | 1.77E-30 | Yes | No |
| rs34311866 | 4 | 951947 | C | T | 0.182 | 0.017284 | 1.59E-06 | 5.68E-15 | Yes | No |
| rs3775067 | 4 | 2888622 | A | G | 0.386 | 0.0136097 | 1.28E-06 | 1.86E-14 | Yes | Yes |
| rs13108218 | 4 | 3443931 | G | A | 0.614 | -0.0286405 | 1.31E-06 | 6.60E-57 | Yes | No |
| rs4450871 | 4 | 4990298 | G | A | 0.439 | -0.0146138 | 1.41E-06 | 1.13E-14 | No | No |
| rs9942171 | 4 | 18032487 | A | G | 0.130 | 0.0208147 | 1.82E-06 | 1.87E-16 | Yes | No |
| rs6448429 | 4 | 26066863 | T | C | 0.164 | 0.0216715 | 1.66E-06 | 6.40E-21 | Yes | No |
| rs35484700 | 4 | 55498781 | C | CG | 0.480 | -0.0105291 | 1.42E-06 | 1.34E-08 | No | No |
| rs9884390 | 4 | 69373407 | C | T | 0.223 | 0.0168636 | 1.55E-06 | 2.46E-15 | Yes | No |
| rs573930512 | 4 | 74512280 | A | G | 0.002 | 0.156249 | 2.48E-05 | 1.56E-09 | No | No |
| rs10008637 | 4 | 77414144 | C | T | 0.456 | 0.0104937 | 1.23E-06 | 8.17E-10 | Yes | No |
| rs17449582 | 4 | 87240157 | T | C | 0.358 | 0.0162529 | 1.28E-06 | 6.22E-20 | Yes | No |
| rs17605615 | 4 | 87996745 | A | G | 0.394 | 0.0272349 | 1.26E-06 | 1.36E-54 | Yes | Yes |
| rs2167750 | 4 | 89730074 | T | C | 0.465 | 0.0121113 | 1.23E-06 | 1.22E-12 | Yes | No |
| rs1126673 | 4 | 100045616 | T | C | 0.694 | 0.0142118 | 1.33E-06 | 1.54E-14 | Yes | No |
| rs13107325 | 4 | 103188709 | T | C | 0.064 | 0.0372865 | 2.47E-06 | 1.76E-27 | Yes | No |
| rs10017313 | 4 | 103861380 | G | A | 0.486 | -0.00997113 | 1.24E-06 | 7.60E-09 | Yes | No |
| rs138307849 | 4 | 110095620 | C | T | 0.006 | 0.121263 | 8.47E-06 | 7.10E-26 | No | No |
| rs114816312 | 4 | 110638824 | T | C | 0.006 | 0.134231 | 8.26E-06 | 9.11E-33 | No | No |
| rs192929239 | 4 | 124641827 | G | A | 0.013 | 0.0462829 | 5.58E-06 | 2.43E-09 | Yes | No |
| rs7696969 | 4 | 143326714 | G | T | 0.632 | -0.0118988 | 1.27E-06 | 1.89E-11 | Yes | No |
| rs6054 | 4 | 155489608 | T | C | 0.005 | 0.118104 | 9.92E-06 | 9.38E-19 | No | No |
| rs6822892 | 4 | 157734675 | G | A | 0.330 | -0.0140865 | 1.31E-06 | 8.82E-15 | Yes | No |
| rs79760705 | 5 | 53298716 | T | G | 0.110 | 0.0258967 | 1.95E-06 | 1.71E-21 | Yes | No |
| rs28650790 | 5 | 55861464 | T | C | 0.181 | 0.0382226 | 1.58E-06 | 1.94E-67 | Yes | No |
| rs151912 | 5 | 57607142 | T | A | 0.589 | -0.0119062 | 1.27E-06 | 1.32E-11 | No | No |
| rs4976033 | 5 | 67714246 | G | A | 0.406 | 0.0176192 | 1.28E-06 | 1.57E-23 | Yes | No |
| rs10052346 | 5 | 78472599 | T | G | 0.390 | -0.0112414 | 1.29E-06 | 2.47E-10 | Yes | No |
| rs115912456 | 5 | 82815158 | G | A | 0.042 | -0.0273591 | 3.05E-06 | 1.09E-10 | Yes | No |
| rs6595187 | 5 | 118723609 | G | A | 0.283 | -0.0192878 | 1.36E-06 | 2.89E-24 | Yes | No |
| rs17764730 | 5 | 127357526 | T | C | 0.232 | -0.0112479 | 1.47E-06 | 3.61E-08 | Yes | No |
| rs253942 | 5 | 131320462 | T | C | 0.062 | 0.0247988 | 2.53E-06 | 1.93E-12 | Yes | No |
| rs72801474 | 5 | 132444128 | A | G | 0.095 | -0.0301264 | 2.13E-06 | 3.22E-24 | Yes | No |
| rs111998037 | 5 | 139798007 | C | A | 0.257 | -0.0110462 | 1.42E-06 | 2.02E-08 | Yes | No |
| rs12523418 | 5 | 141707225 | G | A | 0.320 | -0.0103615 | 1.34E-06 | 2.29E-08 | Yes | No |
| rs245078 | 5 | 149353598 | G | A | 0.406 | -0.00975476 | 1.26E-06 | 2.36E-08 | Yes | No |
| rs6882076 | 5 | 156390297 | C | T | 0.635 | 0.0344004 | 1.27E-06 | 1.23E-84 | Yes | No |
| rs2914231 | 5 | 158011435 | C | G | 0.230 | 0.021926 | 1.46E-06 | 2.72E-27 | Yes | No |
| rs72812818 | 5 | 173356752 | C | G | 0.311 | 0.0107924 | 1.32E-06 | 4.06E-09 | Yes | No |
| rs62397245 | 5 | 176750688 | G | C | 0.231 | 0.0145686 | 1.46E-06 | 8.47E-13 | Yes | No |
| rs6924805 | 6 | 18747705 | T | G | 0.592 | -0.0100688 | 1.25E-06 | 7.49E-09 | Yes | No |
| rs7451008 | 6 | 20673880 | C | T | 0.267 | 0.0118633 | 1.38E-06 | 6.46E-10 | Yes | No |
| rs9379828 | 6 | 26167951 | G | C | 0.355 | -0.00997724 | 1.30E-06 | 2.86E-08 | Yes | No |
| rs56114371 | 6 | 27274834 | T | C | 0.089 | -0.0169634 | 2.22E-06 | 3.68E-08 | No | No |
| rs3131337 | 6 | 28796071 | C | G | 0.106 | -0.0188271 | 2.11E-06 | 1.29E-10 | No | No |
| rs9258375 | 6 | 29752808 | G | A | 0.102 | -0.0217211 | 2.30E-06 | 2.39E-12 | Yes | No |
| rs3094034 | 6 | 30363351 | T | A | 0.097 | -0.0329575 | 3.46E-06 | 2.16E-18 | No | No |
| rs9468937 | 6 | 31270118 | A | C | 0.435 | 0.0280444 | 1.28E-06 | 6.90E-56 | Yes | No |
| rs622871 | 6 | 31878495 | G | A | 0.703 | 0.0281582 | 1.46E-06 | 3.15E-46 | Yes | No |
| rs9461755 | 6 | 32441046 | A | G | 0.046 | 0.0755246 | 3.23E-06 | 9.14E-69 | Yes | No |
| rs144607208 | 6 | 32947516 | G | A | 0.023 | 0.0651614 | 4.46E-06 | 2.68E-27 | Yes | No |
| rs75507056 | 6 | 33457187 | T | C | 0.016 | 0.0562096 | 5.39E-06 | 5.77E-14 | Yes | No |
| rs6934083 | 6 | 35237153 | T | C | 0.034 | 0.0263927 | 3.35E-06 | 1.44E-08 | Yes | No |
| rs3176320 | 6 | 36646788 | G | A | 0.339 | -0.0100726 | 1.30E-06 | 2.17E-08 | Yes | No |
| rs742493 | 6 | 40998167 | C | T | 0.116 | -0.0167097 | 1.90E-06 | 2.68E-10 | Yes | No |
| rs7757363 | 6 | 42876386 | T | C | 0.309 | 0.0103216 | 1.32E-06 | 2.09E-08 | Yes | Yes |
| rs998584 | 6 | 43757896 | A | C | 0.479 | 0.0360954 | 1.29E-06 | 5.54E-94 | Yes | No |
| rs2749005 | 6 | 52621433 | G | T | 0.648 | -0.0186302 | 1.32E-06 | 9.25E-25 | Yes | No |
| rs398085093 | 6 | 86296925 | C | CA | 0.398 | 0.0121528 | 1.50E-06 | 3.60E-10 | No | No |
| rs9496567 | 6 | 100602753 | A | G | 0.236 | -0.0120741 | 1.45E-06 | 2.23E-09 | Yes | No |
| rs6913325 | 6 | 106378009 | T | G | 0.441 | -0.00969798 | 1.23E-06 | 1.61E-08 | Yes | No |
| rs35045014 | 6 | 107432157 | A | C | 0.462 | 0.0135114 | 1.32E-06 | 3.45E-14 | Yes | No |
| rs9480889 | 6 | 109189021 | G | C | 0.788 | 0.0151355 | 1.52E-06 | 7.10E-13 | Yes | No |
| rs577721086 | 6 | 127440047 | C | T | 0.052 | 0.0528261 | 2.82E-06 | 1.61E-41 | Yes | No |
| rs7740188 | 6 | 130346105 | A | G | 0.705 | 0.0143399 | 1.35E-06 | 2.00E-14 | Yes | No |
| rs672457 | 6 | 139836562 | A | G | 0.582 | -0.0262392 | 1.24E-06 | 1.98E-52 | Yes | No |
| rs9321765 | 6 | 140595744 | C | T | 0.208 | -0.0125852 | 1.52E-06 | 2.78E-09 | No | No |
| rs607335 | 6 | 153465230 | A | C | 0.412 | -0.00981496 | 1.25E-06 | 1.68E-08 | Yes | No |
| rs78425119 | 6 | 160501825 | A | G | 0.066 | 0.0345754 | 2.45E-06 | 4.38E-24 | Yes | No |
| rs41272086 | 6 | 161008646 | A | G | 0.102 | 0.0386596 | 2.03E-06 | 1.45E-42 | Yes | No |
| rs1464780 | 6 | 161520837 | A | G | 0.945 | 0.0225946 | 2.74E-06 | 3.01E-09 | Yes | No |
| rs73029263 | 6 | 164113762 | G | A | 0.130 | -0.0188921 | 1.82E-06 | 9.09E-14 | Yes | No |
| rs5011439 | 7 | 12268811 | C | G | 0.403 | 0.0101804 | 1.41E-06 | 4.30E-08 | No | No |
| rs38205 | 7 | 15913588 | C | A | 0.624 | -0.0126731 | 1.29E-06 | 1.23E-12 | Yes | Yes |
| rs4410790 | 7 | 17284577 | C | T | 0.632 | 0.0131573 | 1.27E-06 | 8.31E-14 | Yes | No |
| rs4142995 | 7 | 17919258 | T | G | 0.392 | 0.0129524 | 1.26E-06 | 1.26E-13 | Yes | No |
| rs55696093 | 7 | 21605973 | G | A | 0.210 | 0.0118305 | 1.51E-06 | 1.64E-08 | Yes | No |
| rs4722551 | 7 | 25991826 | C | T | 0.163 | -0.0279113 | 1.65E-06 | 4.02E-34 | Yes | No |
| rs849336 | 7 | 28224053 | G | A | 0.649 | -0.0102383 | 1.28E-06 | 9.27E-09 | Yes | No |
| rs2070971 | 7 | 44197583 | T | G | 0.140 | 0.0291243 | 1.78E-06 | 3.44E-32 | Yes | Yes |
| rs71551223 | 7 | 71256489 | C | A | 0.030 | -0.0366084 | 3.72E-06 | 1.31E-12 | Yes | No |
| rs36104871 | 7 | 71808047 | T | G | 0.029 | -0.0579807 | 3.89E-06 | 1.59E-27 | Yes | No |
| rs35753501 | 7 | 72330303 | T | C | 0.026 | -0.0942105 | 4.15E-06 | 3.40E-60 | Yes | No |
| rs3812316 | 7 | 73020337 | G | C | 0.124 | -0.121587 | 1.85E-06 | 0 | Yes | No |
| rs3135688 | 7 | 73651743 | C | T | 0.040 | -0.0304901 | 3.17E-06 | 4.57E-12 | Yes | Yes |
| rs1057868 | 7 | 75615006 | T | C | 0.288 | 0.0118739 | 1.35E-06 | 2.51E-10 | Yes | No |
| rs1229499 | 7 | 81573045 | A | T | 0.706 | -0.0110413 | 1.36E-06 | 5.65E-09 | Yes | Yes |
| rs2283038 | 7 | 106835410 | T | C | 0.235 | -0.0112869 | 1.48E-06 | 4.29E-08 | Yes | No |
| rs10215153 | 7 | 116399131 | A | G | 0.311 | 0.0120159 | 1.34E-06 | 8.81E-11 | Yes | Yes |
| rs12534129 | 7 | 117018135 | C | T | 0.392 | 0.0120072 | 1.30E-06 | 1.61E-11 | Yes | No |
| rs4731702 | 7 | 130433384 | T | C | 0.495 | -0.0248647 | 1.22E-06 | 1.11E-48 | Yes | No |
| rs7808446 | 7 | 150214465 | G | T | 0.754 | -0.0160134 | 1.43E-06 | 9.38E-16 | Yes | No |
| rs2921060 | 8 | 8317817 | C | A | 0.458 | -0.0159013 | 1.27E-06 | 2.55E-19 | Yes | No |
| rs17149279 | 8 | 9195638 | T | C | 0.231 | -0.0186103 | 1.47E-06 | 9.58E-20 | Yes | No |
| rs615632 | 8 | 9796321 | T | C | 0.525 | 0.0150368 | 1.29E-06 | 3.14E-17 | Yes | No |
| rs7821812 | 8 | 10644101 | C | G | 0.205 | 0.0281306 | 1.52E-06 | 3.39E-40 | Yes | No |
| rs4320509 | 8 | 11528339 | C | G | 0.127 | 0.0319964 | 1.89E-06 | 4.36E-34 | Yes | No |
| rs28415552 | 8 | 13511219 | A | C | 0.631 | -0.0100524 | 1.27E-06 | 1.34E-08 | Yes | No |
| rs1968041 | 8 | 17616562 | T | C | 0.358 | -0.0100629 | 1.29E-06 | 2.19E-08 | Yes | No |
| rs35246381 | 8 | 18272535 | T | C | 0.778 | -0.0342271 | 1.46E-06 | 2.50E-63 | Yes | No |
| rs2958557 | 8 | 19251679 | C | T | 0.992 | -0.0762122 | 7.60E-06 | 3.52E-13 | No | No |
| rs328 | 8 | 19819724 | G | C | 0.100 | -0.179736 | 2.03E-06 | 0 | Yes | No |
| rs2616213 | 8 | 20610866 | C | T | 0.544 | -0.0103828 | 1.25E-06 | 2.12E-09 | Yes | No |
| rs56193509 | 8 | 23525075 | A | T | 0.244 | 0.0109869 | 1.42E-06 | 2.80E-08 | Yes | No |
| rs73221948 | 8 | 25464670 | T | G | 0.278 | 0.0134884 | 1.68E-06 | 1.21E-09 | No | No |
| rs7007256 | 8 | 26203081 | A | G | 0.712 | 0.0102662 | 1.35E-06 | 4.53E-08 | Yes | No |
| rs7816345 | 8 | 36846109 | T | C | 0.169 | 0.014053 | 1.62E-06 | 4.34E-10 | Yes | No |
| rs4647906 | 8 | 38325411 | A | G | 0.402 | 0.0125267 | 1.26E-06 | 7.16E-13 | Yes | No |
| rs2081687 | 8 | 59388565 | C | T | 0.659 | -0.0231942 | 1.29E-06 | 2.01E-38 | Yes | No |
| rs745578 | 8 | 72466324 | A | G | 0.245 | 0.0169094 | 1.43E-06 | 2.15E-17 | Yes | Yes |
| rs7830852 | 8 | 106415088 | G | A | 0.226 | 0.0115959 | 1.47E-06 | 1.32E-08 | Yes | No |
| rs2737246 | 8 | 116659578 | C | G | 0.277 | -0.0124772 | 1.37E-06 | 5.48E-11 | Yes | No |
| rs28601761 | 8 | 126500031 | G | C | 0.421 | -0.0893169 | 1.26E-06 | 0 | No | No |
| rs1561929 | 8 | 129567373 | T | C | 0.883 | 0.0158895 | 1.93E-06 | 2.74E-09 | Yes | Yes |
| rs7832515 | 8 | 144306379 | G | A | 0.197 | -0.0141355 | 1.58E-06 | 1.04E-10 | Yes | No |
| rs7856817 | 9 | 1054587 | A | G | 0.173 | 0.0143397 | 1.62E-06 | 2.02E-10 | No | No |
| rs1658970 | 9 | 6664853 | A | C | 0.139 | 0.0139805 | 1.76E-06 | 1.07E-08 | Yes | Yes |
| rs7041117 | 9 | 13679856 | T | A | 0.583 | -0.0105964 | 1.24E-06 | 9.07E-10 | Yes | No |
| rs686030 | 9 | 15304782 | A | C | 0.859 | 0.0167448 | 1.75E-06 | 6.58E-12 | Yes | No |
| rs62543565 | 9 | 16901067 | A | C | 0.637 | -0.0158249 | 1.28E-06 | 5.48E-19 | Yes | No |
| rs2383766 | 9 | 28412500 | T | G | 0.319 | 0.0100595 | 1.31E-06 | 3.68E-08 | Yes | No |
| rs10971957 | 9 | 34151463 | T | C | 0.598 | -0.0111533 | 1.27E-06 | 2.37E-10 | Yes | No |
| rs296883 | 9 | 86578925 | T | A | 0.264 | -0.0112733 | 1.40E-06 | 6.38E-09 | Yes | No |
| rs17055001 | 9 | 92195246 | A | G | 0.311 | 0.0115782 | 1.33E-06 | 3.76E-10 | Yes | No |
| rs1800978 | 9 | 107665978 | G | C | 0.123 | -0.0252201 | 1.89E-06 | 7.58E-22 | Yes | No |
| rs11789974 | 9 | 110488875 | A | C | 0.032 | -0.0294302 | 3.52E-06 | 1.78E-09 | Yes | No |
| rs75113691 | 9 | 112245773 | C | G | 0.036 | -0.0305867 | 3.30E-06 | 2.57E-11 | No | No |
| rs150611042 | 9 | 117083803 | A | C | 0.069 | -0.0198344 | 2.54E-06 | 1.80E-08 | Yes | No |
| rs2416797 | 9 | 123480600 | A | C | 0.699 | 0.0113578 | 1.34E-06 | 1.14E-09 | Yes | No |
| rs142122182 | 10 | 5263341 | G | A | 0.155 | -0.0213308 | 1.74E-06 | 4.34E-19 | Yes | No |
| rs4934927 | 10 | 33628062 | C | G | 0.090 | 0.0180806 | 2.21E-06 | 2.39E-09 | Yes | No |
| rs41274050 | 10 | 52573772 | T | C | 0.009 | 0.0860984 | 7.30E-06 | 3.19E-19 | No | No |
| rs1171616 | 10 | 61468589 | T | G | 0.775 | 0.0156446 | 1.48E-06 | 2.60E-14 | Yes | Yes |
| rs35706909 | 10 | 63922233 | C | T | 0.099 | -0.0178206 | 2.06E-06 | 4.84E-10 | Yes | No |
| rs7924036 | 10 | 65191645 | T | G | 0.512 | -0.0292802 | 1.24E-06 | 1.73E-64 | Yes | No |
| rs7904738 | 10 | 74710484 | C | G | 0.086 | 0.0239034 | 2.20E-06 | 6.29E-15 | Yes | No |
| rs149590572 | 10 | 93575831 | A | G | 0.064 | 0.0247658 | 2.49E-06 | 9.53E-13 | Yes | No |
| rs17875327 | 10 | 94274809 | G | A | 0.111 | 0.0240046 | 2.00E-06 | 4.88E-18 | Yes | No |
| rs2068888 | 10 | 94839642 | A | G | 0.451 | -0.0291415 | 1.24E-06 | 8.25E-65 | Yes | Yes |
| rs7091593 | 10 | 95342772 | T | A | 0.113 | 0.0177942 | 1.95E-06 | 4.96E-11 | Yes | No |
| rs3891783 | 10 | 96015793 | G | C | 0.434 | -0.0100667 | 1.24E-06 | 5.26E-09 | No | No |
| rs1408579 | 10 | 101912194 | T | C | 0.482 | 0.00997137 | 1.22E-06 | 4.51E-09 | Yes | No |
| rs75398587 | 10 | 103946480 | G | C | 0.062 | -0.0265685 | 2.53E-06 | 4.98E-14 | Yes | No |
| rs2803619 | 10 | 113934384 | C | G | 0.718 | 0.0179132 | 1.35E-06 | 1.56E-21 | Yes | No |
| rs2484294 | 10 | 115792062 | A | G | 0.737 | -0.0105345 | 1.39E-06 | 4.68E-08 | Yes | No |
| rs12772930 | 10 | 120493295 | A | G | 0.353 | 0.0103304 | 1.29E-06 | 7.58E-09 | Yes | No |
| rs1873449 | 10 | 122931652 | T | G | 0.022 | -0.0367787 | 4.23E-06 | 3.73E-10 | Yes | No |
| rs1133400 | 10 | 134459388 | G | A | 0.213 | 0.0134112 | 1.52E-06 | 1.71E-10 | Yes | No |
| rs537132753 | 10 | 135440683 | G | T | 0.034 | 0.0355115 | 4.62E-06 | 2.60E-09 | No | No |
| rs117739035 | 11 | 408174 | T | G | 0.036 | 0.0253911 | 3.39E-06 | 4.75E-08 | Yes | No |
| rs4909945 | 11 | 10673739 | C | T | 0.680 | 0.0134149 | 1.30E-06 | 1.45E-13 | Yes | No |
| rs1037169 | 11 | 13361005 | C | T | 0.683 | 0.0180982 | 1.36E-06 | 2.60E-22 | Yes | No |
| rs150090666 | 11 | 14865399 | T | C | 0.001 | -0.302766 | 2.52E-05 | 4.07E-22 | No | No |
| rs142108391 | 11 | 26216166 | A | G | 0.063 | 0.0208814 | 2.52E-06 | 2.50E-09 | Yes | No |
| rs1519480 | 11 | 27675712 | T | C | 0.679 | -0.01485 | 1.31E-06 | 3.40E-16 | Yes | No |
| rs75160368 | 11 | 30402714 | A | G | 0.088 | -0.01732 | 2.22E-06 | 2.09E-08 | Yes | No |
| rs117847116 | 11 | 46048240 | C | T | 0.040 | -0.0269749 | 3.17E-06 | 1.02E-09 | Yes | No |
| rs3758669 | 11 | 47272579 | G | A | 0.310 | -0.0230282 | 1.33E-06 | 2.16E-35 | Yes | No |
| rs541958245 | 11 | 56385221 | T | A | 0.002 | -0.158508 | 2.52E-05 | 3.03E-08 | No | No |
| rs174562 | 11 | 61585144 | G | A | 0.346 | 0.0522293 | 1.28E-06 | 1.91E-187 | Yes | No |
| rs11231161 | 11 | 62378221 | G | A | 0.358 | 0.0111123 | 1.29E-06 | 5.86E-10 | Yes | Yes |
| rs56271783 | 11 | 64004723 | C | G | 0.047 | 0.0530991 | 2.89E-06 | 7.35E-40 | Yes | No |
| rs10750766 | 11 | 65473798 | A | C | 0.710 | 0.017147 | 1.36E-06 | 1.05E-19 | Yes | No |
| rs10896373 | 11 | 68614810 | T | A | 0.514 | -0.0141036 | 1.24E-06 | 2.44E-16 | No | No |
| rs10796821 | 11 | 69305417 | G | A | 0.513 | 0.0096435 | 1.26E-06 | 2.42E-08 | Yes | No |
| rs2063724 | 11 | 78133077 | C | T | 0.173 | -0.0144717 | 1.64E-06 | 2.12E-10 | Yes | No |
| rs759831202 | 11 | 115554537 | A | T | 0.000 | -0.621662 | 7.47E-05 | 3.89E-17 | No | No |
| rs117288140 | 11 | 116139965 | T | C | 0.017 | 0.0395228 | 5.04E-06 | 1.59E-08 | Yes | No |
| rs964184 | 11 | 116648917 | C | G | 0.865 | -0.246288 | 1.78E-06 | 0 | Yes | No |
| rs192489212 | 11 | 117220429 | C | A | 0.002 | -0.893298 | 1.81E-05 | 1.04E-292 | No | No |
| rs747437837 | 11 | 117849058 | G | C | 0.001 | -0.413548 | 6.08E-05 | 8.34E-11 | No | No |
| rs749804194 | 11 | 118574327 | C | T | 0.000 | -0.827719 | 0.000106 | 2.31E-16 | No | No |
| rs76895963 | 12 | 4384844 | G | T | 0.018 | -0.10398 | 4.98E-06 | 5.11E-51 | No | No |
| rs73047887 | 12 | 6736843 | C | T | 0.255 | 0.0113359 | 1.48E-06 | 3.26E-08 | Yes | No |
| rs11045171 | 12 | 20470199 | G | A | 0.200 | -0.0201072 | 1.55E-06 | 7.51E-21 | Yes | No |
| rs4149056 | 12 | 21331549 | C | T | 0.158 | 0.0270573 | 1.67E-06 | 2.25E-31 | Yes | Yes |
| rs11046481 | 12 | 22747967 | G | T | 0.235 | -0.0154524 | 1.49E-06 | 9.88E-14 | Yes | Yes |
| rs17389465 | 12 | 25414408 | A | T | 0.069 | -0.0199909 | 2.42E-06 | 2.82E-09 | Yes | No |
| rs4963975 | 12 | 26443030 | A | G | 0.250 | 0.0150408 | 1.42E-06 | 2.55E-14 | Yes | No |
| rs10082956 | 12 | 29504556 | G | A | 0.290 | -0.0127726 | 1.35E-06 | 1.15E-11 | Yes | No |
| rs11183212 | 12 | 46213867 | G | A | 0.202 | 0.0157919 | 1.55E-06 | 2.33E-13 | Yes | Yes |
| rs145878042 | 12 | 48143315 | G | A | 0.011 | 0.0464549 | 5.81E-06 | 8.89E-09 | Yes | No |
| rs1126930 | 12 | 49399132 | C | G | 0.034 | 0.0279141 | 3.39E-06 | 3.05E-09 | Yes | No |
| rs7312441 | 12 | 56941146 | A | C | 0.345 | -0.0114274 | 1.33E-06 | 4.27E-10 | Yes | No |
| rs79395356 | 12 | 57738600 | G | T | 0.248 | -0.0248319 | 1.46E-06 | 4.52E-35 | Yes | No |
| rs8756 | 12 | 66359752 | A | C | 0.509 | 0.00972395 | 1.22E-06 | 9.37E-09 | Yes | No |
| rs11615712 | 12 | 69619736 | T | A | 0.448 | -0.00958192 | 1.27E-06 | 4.70E-08 | No | No |
| rs11113118 | 12 | 107199142 | A | G | 0.235 | 0.0166424 | 1.45E-06 | 1.25E-16 | Yes | No |
| rs4964736 | 12 | 109100503 | T | C | 0.359 | 0.0101781 | 1.28E-06 | 1.16E-08 | Yes | No |
| rs149793040 | 12 | 109661672 | G | A | 0.001 | -0.215222 | 2.05E-05 | 2.91E-17 | No | No |
| rs2009170 | 12 | 111703313 | C | T | 0.255 | -0.0127277 | 1.43E-06 | 1.20E-10 | Yes | No |
| rs580063 | 12 | 123206340 | C | T | 0.212 | -0.01712 | 1.51E-06 | 2.66E-16 | Yes | No |
| rs12303671 | 12 | 124492610 | G | T | 0.337 | -0.025901 | 1.31E-06 | 5.37E-46 | Yes | No |
| rs11057840 | 12 | 125316055 | C | A | 0.138 | 0.0190088 | 1.78E-06 | 1.36E-14 | Yes | No |
| rs4770433 | 13 | 23903791 | G | A | 0.449 | -0.0105335 | 1.22E-06 | 6.03E-10 | Yes | Yes |
| rs1340819 | 13 | 29145323 | C | A | 0.337 | -0.00996674 | 1.31E-06 | 3.69E-08 | Yes | No |
| rs398098735 | 13 | 31001091 | A | AT | 0.389 | -0.0125411 | 1.46E-06 | 4.15E-11 | No | No |
| rs138358301 | 13 | 45970147 | G | A | 0.004 | 0.0711919 | 9.48E-06 | 4.23E-08 | No | No |
| rs2812208 | 13 | 50707087 | C | G | 0.023 | -0.0480352 | 4.19E-06 | 1.55E-16 | Yes | No |
| rs797482 | 13 | 51214613 | G | A | 0.875 | 0.0149092 | 1.85E-06 | 6.72E-09 | Yes | No |
| rs2298058 | 13 | 95248566 | T | C | 0.316 | 0.0183947 | 1.33E-06 | 2.00E-23 | Yes | Yes |
| rs6602909 | 13 | 114551993 | C | T | 0.330 | 0.0263997 | 1.35E-06 | 6.66E-46 | Yes | No |
| rs72681869 | 14 | 50655357 | C | G | 0.009 | -0.0542227 | 6.96E-06 | 1.57E-08 | No | No |
| rs12878001 | 14 | 64239629 | G | T | 0.161 | 0.0207307 | 1.67E-06 | 4.82E-19 | Yes | No |
| rs3825669 | 14 | 89804276 | G | A | 0.805 | -0.0122844 | 1.56E-06 | 1.34E-08 | Yes | No |
| rs61993685 | 14 | 100765823 | C | T | 0.071 | -0.0194213 | 2.41E-06 | 6.12E-09 | Yes | No |
| rs12891399 | 14 | 104293533 | C | T | 0.339 | 0.0112518 | 1.30E-06 | 5.14E-10 | Yes | No |
| rs28624578 | 15 | 31637666 | C | T | 0.166 | 0.0134612 | 1.69E-06 | 7.93E-09 | Yes | No |
| rs275179 | 15 | 39447529 | G | A | 0.158 | -0.0154206 | 1.70E-06 | 6.88E-11 | Yes | No |
| rs34245505 | 15 | 40397191 | G | C | 0.188 | 0.0153228 | 1.66E-06 | 1.37E-11 | Yes | No |
| rs72735627 | 15 | 41057507 | T | C | 0.088 | -0.0206445 | 2.27E-06 | 5.46E-11 | Yes | No |
| rs8025212 | 15 | 41880619 | G | A | 0.324 | -0.01134 | 1.33E-06 | 8.38E-10 | Yes | No |
| rs139661283 | 15 | 42384229 | A | G | 0.014 | 0.0603118 | 5.20E-06 | 7.84E-17 | Yes | No |
| rs139974673 | 15 | 44027885 | C | T | 0.026 | 0.13397 | 3.83E-06 | 9.32E-140 | Yes | No |
| rs1901529 | 15 | 45001900 | G | A | 0.121 | 0.0181697 | 2.01E-06 | 6.30E-11 | Yes | No |
| rs72749502 | 15 | 57297078 | T | A | 0.070 | 0.0296693 | 2.45E-06 | 3.46E-18 | Yes | No |
| rs1077835 | 15 | 58723426 | G | A | 0.216 | 0.0447226 | 1.49E-06 | 1.14E-103 | Yes | No |
| rs112147665 | 15 | 59277019 | C | T | 0.002 | 0.183501 | 2.00E-05 | 2.71E-16 | No | No |
| rs12438742 | 15 | 61947280 | G | C | 0.560 | -0.0103506 | 1.24E-06 | 1.69E-09 | No | No |
| rs11635675 | 15 | 63793238 | G | T | 0.352 | 0.0235284 | 1.28E-06 | 6.14E-40 | Yes | Yes |
| rs12442852 | 15 | 64677413 | T | C | 0.881 | 0.0149168 | 1.94E-06 | 2.79E-08 | Yes | No |
| rs2218181 | 15 | 66872325 | C | T | 0.346 | 0.0140202 | 1.28E-06 | 3.40E-15 | Yes | Yes |
| rs11072332 | 15 | 72108307 | T | G | 0.214 | -0.0122426 | 1.51E-06 | 5.05E-09 | Yes | No |
| rs7164727 | 15 | 73093991 | T | C | 0.681 | 0.0109811 | 1.32E-06 | 2.19E-09 | Yes | No |
| rs1037117 | 15 | 102068658 | A | G | 0.253 | 0.0140873 | 1.44E-06 | 1.40E-12 | Yes | Yes |
| rs12600110 | 16 | 962154 | C | T | 0.375 | -0.0150678 | 1.29E-06 | 3.90E-17 | Yes | No |
| rs12921195 | 16 | 4677604 | A | C | 0.129 | 0.0170527 | 2.15E-06 | 1.62E-09 | Yes | No |
| rs12928099 | 16 | 15150505 | A | C | 0.297 | -0.025582 | 1.35E-06 | 2.24E-42 | Yes | No |
| rs7196161 | 16 | 31110981 | A | G | 0.624 | 0.0149822 | 1.27E-06 | 1.90E-17 | Yes | No |
| rs62033400 | 16 | 53811788 | G | A | 0.401 | 0.0156307 | 1.25E-06 | 2.08E-19 | Yes | No |
| rs17231506 | 16 | 56994528 | T | C | 0.320 | -0.0329787 | 1.31E-06 | 4.65E-73 | Yes | No |
| rs73597575 | 16 | 67677001 | T | C | 0.085 | -0.0174762 | 2.18E-06 | 8.62E-09 | Yes | No |
| rs6499240 | 16 | 69686912 | G | A | 0.582 | 0.0162186 | 1.27E-06 | 2.87E-20 | Yes | No |
| rs12445401 | 16 | 72148419 | G | A | 0.192 | 0.0221914 | 1.59E-06 | 3.47E-24 | Yes | No |
| rs76862947 | 16 | 79746461 | C | T | 0.289 | 0.0120231 | 1.36E-06 | 2.25E-10 | Yes | No |
| rs2925979 | 16 | 81534790 | C | T | 0.699 | -0.0257081 | 1.33E-06 | 9.10E-44 | Yes | No |
| rs11641586 | 16 | 85270707 | T | G | 0.198 | -0.013998 | 1.57E-06 | 1.52E-10 | Yes | No |
| rs8054124 | 16 | 86434553 | C | T | 0.231 | 0.0121188 | 1.46E-06 | 2.52E-09 | Yes | No |
| rs11870735 | 17 | 481604 | T | C | 0.176 | 0.0133745 | 1.61E-06 | 2.52E-09 | Yes | No |
| rs11078597 | 17 | 1618363 | C | T | 0.187 | 0.0174414 | 1.59E-06 | 2.71E-15 | Yes | Yes |
| rs79202680 | 17 | 4692640 | T | G | 0.006 | -0.149708 | 8.06E-06 | 3.55E-41 | No | No |
| rs571475342 | 17 | 5207882 | G | A | 0.005 | -0.106433 | 9.23E-06 | 2.71E-17 | No | No |
| rs200489612 | 17 | 7106378 | A | G | 0.004 | -0.118553 | 1.13E-05 | 7.32E-15 | No | No |
| rs9905032 | 17 | 17457768 | A | G | 0.063 | 0.0287209 | 2.53E-06 | 3.14E-16 | Yes | No |
| rs704 | 17 | 26694861 | A | G | 0.477 | -0.0111031 | 1.23E-06 | 7.48E-11 | Yes | No |
| rs17767256 | 17 | 28581043 | G | T | 0.337 | -0.0106848 | 1.30E-06 | 3.31E-09 | Yes | No |
| rs650558 | 17 | 40721042 | T | C | 0.242 | 0.0153589 | 1.44E-06 | 1.47E-14 | Yes | No |
| rs186398347 | 17 | 41335716 | G | A | 0.005 | 0.0933955 | 9.80E-06 | 4.90E-12 | No | No |
| rs72836561 | 17 | 41926126 | T | C | 0.030 | 0.149617 | 3.58E-06 | 2.98E-199 | Yes | No |
| rs72824773 | 17 | 42626279 | A | G | 0.006 | 0.0739476 | 9.06E-06 | 2.88E-09 | Yes | No |
| rs7207542 | 17 | 45697549 | G | C | 0.495 | 0.0104051 | 1.27E-06 | 2.10E-09 | No | No |
| rs10775406 | 17 | 46197755 | G | A | 0.752 | 0.0151746 | 1.43E-06 | 1.90E-14 | Yes | No |
| rs28394864 | 17 | 47450775 | A | G | 0.462 | 0.015812 | 1.25E-06 | 5.50E-20 | Yes | No |
| rs1292072 | 17 | 57925649 | G | A | 0.211 | 0.0164089 | 1.50E-06 | 4.31E-15 | Yes | No |
| rs1801689 | 17 | 64210580 | C | A | 0.026 | -0.0573699 | 3.80E-06 | 1.83E-27 | Yes | No |
| rs11656743 | 17 | 65980289 | A | G | 0.788 | -0.0244634 | 1.52E-06 | 2.86E-31 | Yes | No |
| rs77542162 | 17 | 67081278 | G | A | 0.020 | -0.0487845 | 4.51E-06 | 6.50E-15 | Yes | No |
| rs717118 | 17 | 68464817 | A | T | 0.506 | -0.0102742 | 1.24E-06 | 2.30E-09 | No | No |
| rs3744231 | 17 | 73268801 | T | C | 0.701 | 0.0103018 | 1.36E-06 | 4.76E-08 | Yes | No |
| rs193220 | 17 | 74268619 | T | C | 0.309 | -0.0117996 | 1.34E-06 | 1.94E-10 | Yes | No |
| rs7212201 | 17 | 76393896 | A | C | 0.611 | -0.0195429 | 1.25E-06 | 3.17E-29 | Yes | No |
| rs62078746 | 17 | 80053590 | A | G | 0.553 | -0.0106501 | 1.27E-06 | 1.24E-09 | Yes | No |
| rs8083730 | 18 | 293519 | A | G | 0.082 | -0.0185654 | 2.22E-06 | 1.73E-09 | No | No |
| rs11664106 | 18 | 2846812 | T | A | 0.364 | -0.0108268 | 1.45E-06 | 2.76E-08 | No | No |
| rs35321365 | 18 | 19905141 | T | C | 0.516 | -0.0114514 | 1.25E-06 | 3.66E-11 | Yes | No |
| rs1623060 | 18 | 21143183 | T | C | 0.493 | -0.0145633 | 1.23E-06 | 1.57E-17 | Yes | No |
| rs150261265 | 18 | 29772887 | C | T | 0.001 | 0.191355 | 2.63E-05 | 1.35E-10 | No | No |
| rs143393480 | 18 | 56089858 | T | G | 0.009 | 0.0561342 | 6.53E-06 | 5.86E-10 | No | No |
| rs1457489 | 18 | 57861961 | A | G | 0.270 | 0.0132806 | 1.39E-06 | 6.08E-12 | Yes | No |
| rs12454712 | 18 | 60845884 | C | T | 0.380 | -0.0150757 | 1.33E-06 | 1.55E-16 | No | No |
| rs1135908 | 19 | 808742 | T | G | 0.146 | 0.0134369 | 1.80E-06 | 4.95E-08 | Yes | No |
| rs350885 | 19 | 4147445 | C | A | 0.742 | 0.0168072 | 1.43E-06 | 1.82E-17 | Yes | Yes |
| rs35502362 | 19 | 4966041 | T | C | 0.338 | -0.0151418 | 1.31E-06 | 6.07E-17 | Yes | No |
| rs4804414 | 19 | 7223785 | T | C | 0.427 | 0.0203605 | 1.24E-06 | 2.42E-32 | Yes | No |
| rs11672003 | 19 | 7909765 | A | G | 0.276 | 0.0138575 | 1.41E-06 | 1.35E-12 | Yes | No |
| rs116843064 | 19 | 8429323 | A | G | 0.020 | -0.23635 | 4.40E-06 | 0 | Yes | No |
| rs555628099 | 19 | 8945880 | G | A | 0.004 | -0.0900764 | 9.63E-06 | 1.29E-11 | No | No |
| rs145464906 | 19 | 11350874 | T | C | 0.001 | -0.361925 | 3.47E-05 | 2.22E-19 | No | No |
| rs1966500 | 19 | 18735666 | C | T | 0.452 | -0.0122074 | 1.27E-06 | 3.33E-12 | Yes | No |
| rs58542926 | 19 | 19379549 | T | C | 0.075 | -0.108022 | 2.32E-06 | 6.77E-246 | Yes | No |
| rs75916629 | 19 | 20213781 | G | A | 0.025 | -0.0565652 | 4.13E-06 | 5.54E-23 | Yes | No |
| rs7256564 | 19 | 33889593 | G | A | 0.689 | -0.0146903 | 1.33E-06 | 1.37E-15 | Yes | No |
| rs58895965 | 19 | 35551428 | A | C | 0.165 | 0.0238047 | 1.64E-06 | 2.11E-25 | Yes | No |
| rs35538872 | 19 | 41754430 | A | G | 0.006 | -0.0889336 | 8.34E-06 | 1.55E-14 | No | No |
| rs141226346 | 19 | 44890969 | G | C | 0.002 | -0.212674 | 1.83E-05 | 6.24E-19 | No | No |
| rs483082 | 19 | 45416178 | T | G | 0.229 | 0.0960549 | 1.48E-06 | 0 | Yes | No |
| rs12459222 | 19 | 46859301 | C | G | 0.301 | -0.0129416 | 1.35E-06 | 4.05E-12 | Yes | No |
| rs10408163 | 19 | 47597102 | C | T | 0.709 | 0.0118257 | 1.35E-06 | 2.55E-10 | Yes | No |
| rs62129968 | 19 | 48383542 | A | C | 0.162 | -0.0164856 | 1.69E-06 | 1.52E-12 | Yes | No |
| rs838133 | 19 | 49259529 | G | A | 0.556 | -0.019842 | 1.35E-06 | 3.98E-27 | No | No |
| rs113886122 | 19 | 50044741 | G | C | 0.151 | 0.0198719 | 1.73E-06 | 1.41E-16 | Yes | No |
| rs798889 | 19 | 54793250 | T | G | 0.220 | -0.0130809 | 1.50E-06 | 3.63E-10 | Yes | No |
| rs7251733 | 19 | 56099951 | A | G | 0.161 | 0.0209954 | 1.68E-06 | 2.13E-19 | Yes | No |
| rs8102873 | 19 | 57488423 | T | C | 0.581 | 0.0104809 | 1.24E-06 | 1.27E-09 | Yes | No |
| rs151235402 | 20 | 569164 | T | C | 0.017 | 0.0475823 | 4.93E-06 | 3.56E-12 | Yes | No |
| rs6120879 | 20 | 30202961 | C | T | 0.164 | -0.012668 | 1.67E-06 | 4.49E-08 | Yes | No |
| rs67611724 | 20 | 32308275 | T | C | 0.147 | 0.0155546 | 1.74E-06 | 1.42E-10 | Yes | No |
| rs78185617 | 20 | 34124603 | C | G | 0.132 | -0.0182643 | 1.84E-06 | 5.41E-13 | Yes | No |
| rs1883711 | 20 | 39179822 | C | G | 0.032 | 0.0590939 | 3.68E-06 | 7.09E-31 | Yes | No |
| rs6102322 | 20 | 39872768 | T | C | 0.298 | 0.0157916 | 1.35E-06 | 2.95E-17 | Yes | No |
| rs6073958 | 20 | 44551855 | C | T | 0.202 | 0.0487116 | 1.52E-06 | 7.47E-117 | Yes | No |
| rs6066138 | 20 | 45594711 | A | G | 0.274 | -0.0171088 | 1.37E-06 | 3.31E-19 | Yes | No |
| rs6068174 | 20 | 51046309 | T | C | 0.637 | 0.0112436 | 1.29E-06 | 3.34E-10 | Yes | No |
| rs6070138 | 20 | 56118558 | A | C | 0.594 | 0.0146908 | 1.28E-06 | 5.72E-17 | Yes | Yes |
| rs8126001 | 20 | 62711459 | T | C | 0.484 | -0.0158162 | 1.43E-06 | 6.08E-18 | Yes | No |
| rs8134638 | 21 | 40644170 | C | T | 0.384 | 0.0122478 | 1.29E-06 | 6.24E-12 | Yes | No |
| rs960230 | 21 | 42622479 | G | A | 0.786 | 0.0117786 | 1.51E-06 | 1.88E-08 | Yes | No |
| rs200559406 | 21 | 46875817 | A | G | 0.001 | 0.23301 | 3.39E-05 | 3.34E-08 | No | No |
| rs35665085 | 22 | 17625915 | A | G | 0.056 | 0.0230441 | 2.70E-06 | 8.51E-10 | Yes | No |
| rs4822458 | 22 | 24265659 | T | C | 0.546 | -0.0110083 | 1.26E-06 | 2.45E-10 | Yes | No |
| rs4352308 | 22 | 29131788 | T | C | 0.708 | 0.0115682 | 1.36E-06 | 9.30E-10 | Yes | No |
| rs2530679 | 22 | 30082569 | G | A | 0.548 | 0.0108578 | 1.25E-06 | 4.04E-10 | No | No |
| rs9610329 | 22 | 36042986 | T | C | 0.437 | 0.0124373 | 1.30E-06 | 3.94E-12 | No | No |
| rs4820325 | 22 | 38599978 | A | G | 0.582 | 0.0209178 | 1.26E-06 | 3.70E-33 | Yes | No |
| rs4821815 | 22 | 39105707 | A | G | 0.334 | 0.0151832 | 1.30E-06 | 4.60E-17 | Yes | Yes |
| rs3747207 | 22 | 44324855 | A | G | 0.222 | -0.0121357 | 1.47E-06 | 3.01E-09 | Yes | No |
| rs4253766 | 22 | 46623905 | T | C | 0.107 | 0.0225365 | 1.98E-06 | 2.91E-16 | Yes | Yes |
| rs140947018 | 22 | 50165082 | A | G | 0.029 | -0.0307324 | 3.73E-06 | 3.22E-09 | Yes | No |
| rs73188911 | 22 | 50746706 | T | C | 0.082 | -0.0182969 | 2.30E-06 | 9.16E-09 | Yes | No |

**Supplemental Table 7.** SNPs included in systolic blood pressure-related analyses.

| RSID | Chrom. | Position | Effect allele | Other allele | Effect allele freq. | Beta | SE | p-value | Used in MR | Used in MR + Steiger filt. |
| --- | --- | --- | --- | --- | --- | --- | --- | --- | --- | --- |
| rs2076328 | 1 | 1687482 | T | G | 0.4892 | -0.334 | 0.0311 | 7.96E-27 | Yes | No |
| rs260508 | 1 | 2187085 | T | G | 0.6151 | 0.1578 | 0.0311 | 3.85E-07 | Yes | No |
| rs2493292 | 1 | 3328659 | T | C | 0.1446 | 0.4078 | 0.0438 | 1.35E-20 | Yes | No |
| rs709209 | 1 | 6278414 | A | G | 0.6582 | 0.0899 | 0.0329 | 0.006243 | Yes | No |
| rs4908678 | 1 | 7739250 | T | C | 0.6253 | -0.1343 | 0.0315 | 1.98E-05 | Yes | No |
| rs2252865 | 1 | 8422676 | T | C | 0.3485 | 0.2022 | 0.0317 | 1.75E-10 | Yes | No |
| rs9662255 | 1 | 9441949 | A | C | 0.4271 | -0.2291 | 0.031 | 1.40E-13 | Yes | No |
| rs880315 | 1 | 10796866 | T | C | 0.6565 | -0.5076 | 0.0323 | 9.59E-56 | Yes | Yes |
| rs17367504 | 1 | 11862778 | A | G | 0.8409 | 0.8968 | 0.0413 | 1.58E-104 | Yes | Yes |
| rs3820068 | 1 | 15798197 | A | G | 0.8038 | 0.2935 | 0.0387 | 3.31E-14 | Yes | No |
| rs2807337 | 1 | 22577371 | T | C | 0.3655 | 0.1864 | 0.0314 | 2.78E-09 | Yes | Yes |
| rs150266910 | 1 | 23442265 | T | C | 0.1766 | 0.1152 | 0.0394 | 0.003509 | Yes | No |
| rs6686889 | 1 | 25030470 | T | C | 0.2531 | 0.205 | 0.0348 | 3.89E-09 | Yes | No |
| rs79598313 | 1 | 27284913 | T | C | 0.0252 | 0.6651 | 0.0996 | 2.40E-11 | Yes | No |
| rs3737801 | 1 | 27960832 | C | G | 0.9224 | 0.3886 | 0.0601 | 1.02E-10 | Yes | No |
| rs143167197 | 1 | 28734372 | A | G | 0.9286 | -0.4337 | 0.0618 | 2.31E-12 | Yes | No |
| rs1565716 | 1 | 29549216 | A | G | 0.0697 | 0.247 | 0.0594 | 3.23E-05 | Yes | No |
| rs4652875 | 1 | 33868469 | C | G | 0.4224 | 0.1396 | 0.0305 | 4.81E-06 | No | No |
| rs9729719 | 1 | 38298207 | A | G | 0.2932 | 0.1529 | 0.0338 | 5.95E-06 | Yes | No |
| rs11210029 | 1 | 41865293 | A | G | 0.6322 | -0.203 | 0.0313 | 8.92E-11 | Yes | No |
| rs7515635 | 1 | 42408070 | T | C | 0.4619 | 0.2509 | 0.0303 | 1.25E-16 | Yes | No |
| rs72659998 | 1 | 43037556 | T | C | 0.1508 | -0.1811 | 0.0427 | 2.22E-05 | Yes | No |
| rs839755 | 1 | 43856410 | A | C | 0.6149 | -0.2665 | 0.0308 | 5.41E-18 | Yes | No |
| rs512083 | 1 | 46027355 | T | C | 0.5349 | -0.1111 | 0.0303 | 0.0002425 | Yes | No |
| rs12142296 | 1 | 46541679 | T | G | 0.8639 | -0.112 | 0.0442 | 0.01134 | Yes | No |
| rs4926923 | 1 | 48109225 | T | C | 0.9114 | 0.3199 | 0.0537 | 2.56E-09 | Yes | No |
| rs11579440 | 1 | 49052423 | T | C | 0.8483 | 0.2674 | 0.0425 | 3.24E-10 | Yes | Yes |
| rs147696085 | 1 | 51021867 | A | G | 0.094 | -0.0841 | 0.0527 | 0.1103 | Yes | No |
| rs6681713 | 1 | 51527684 | T | G | 0.9811 | 0.3175 | 0.1138 | 0.005268 | Yes | No |
| rs112557609 | 1 | 56576924 | A | G | 0.3438 | 0.2838 | 0.0319 | 5.87E-19 | Yes | No |
| rs2404715 | 1 | 57008778 | T | C | 0.0925 | -0.305 | 0.0523 | 5.49E-09 | Yes | No |
| rs60199046 | 1 | 59663341 | A | G | 0.711 | 0.2722 | 0.0332 | 2.41E-16 | Yes | No |
| rs20354 | 1 | 67071356 | T | G | 0.1335 | 0.2014 | 0.0442 | 5.31E-06 | Yes | No |
| rs34517439 | 1 | 78450517 | A | C | 0.1191 | -0.2446 | 0.049 | 5.95E-07 | No | No |
| rs12034319 | 1 | 86040107 | A | G | 0.2197 | 0.1267 | 0.0365 | 0.000519 | Yes | No |
| rs385437 | 1 | 86822231 | A | G | 0.8598 | 0.1831 | 0.0438 | 2.89E-05 | Yes | No |
| rs10923038 | 1 | 88651771 | A | C | 0.6179 | 0.2008 | 0.0313 | 1.36E-10 | Yes | Yes |
| rs10922502 | 1 | 89360158 | A | G | 0.628 | -0.2863 | 0.0313 | 6.14E-20 | Yes | No |
| rs2065152 | 1 | 90228519 | T | C | 0.3574 | 0.155 | 0.0314 | 8.12E-07 | Yes | No |
| rs17516329 | 1 | 92319781 | A | T | 0.6855 | 0.0971 | 0.0327 | 0.002961 | Yes | No |
| rs7514579 | 1 | 94051350 | A | C | 0.7712 | 0.2243 | 0.0361 | 5.45E-10 | Yes | No |
| rs17396055 | 1 | 94730954 | A | G | 0.3317 | -0.1762 | 0.0321 | 3.96E-08 | Yes | No |
| rs17030613 | 1 | 113190807 | A | C | 0.7898 | -0.4053 | 0.0371 | 8.36E-28 | Yes | No |
| rs2932538 | 1 | 113216543 | A | G | 0.2569 | -0.3389 | 0.0345 | 8.48E-23 | Yes | No |
| rs12078697 | 1 | 117015118 | C | G | 0.2113 | -0.1462 | 0.0371 | 8.06E-05 | Yes | No |
| rs7553422 | 1 | 119540719 | T | C | 0.4117 | -0.1007 | 0.0306 | 0.001004 | Yes | No |
| rs72704264 | 1 | 145713305 | C | G | 0.2163 | 0.0926 | 0.0372 | 0.01273 | Yes | No |
| rs11585169 | 1 | 150572037 | A | T | 0.5773 | 0.1796 | 0.0308 | 5.34E-09 | No | No |
| rs13796 | 1 | 154245917 | T | C | 0.8643 | -0.053 | 0.0449 | 0.2377 | Yes | No |
| rs76719272 | 1 | 156129796 | T | C | 0.1312 | -0.2738 | 0.0461 | 2.97E-09 | Yes | No |
| rs2171690 | 1 | 164740099 | T | C | 0.5352 | 0.1527 | 0.0303 | 4.76E-07 | Yes | No |
| rs7524019 | 1 | 167367193 | T | C | 0.4909 | 0.1308 | 0.0303 | 1.60E-05 | Yes | No |
| rs2157597 | 1 | 169201567 | T | C | 0.3546 | -0.1432 | 0.0317 | 6.18E-06 | Yes | No |
| rs12405515 | 1 | 172357441 | T | G | 0.5709 | -0.1411 | 0.0304 | 3.33E-06 | Yes | No |
| rs12118102 | 1 | 176634724 | A | G | 0.947 | 0.1916 | 0.0676 | 0.004579 | Yes | No |
| rs150816167 | 1 | 179571862 | T | C | 0.955 | -0.2367 | 0.0782 | 0.002465 | Yes | No |
| rs10913934 | 1 | 180131640 | T | G | 0.5929 | 0.1021 | 0.0307 | 0.0008957 | Yes | No |
| rs1043069 | 1 | 180859368 | T | G | 0.6156 | 0.234 | 0.0311 | 5.26E-14 | Yes | Yes |
| rs41475048 | 1 | 183058452 | A | G | 0.745 | -0.1233 | 0.0352 | 0.0004657 | Yes | No |
| rs4651224 | 1 | 184585182 | T | C | 0.4474 | 0.1986 | 0.0306 | 9.00E-11 | Yes | No |
| rs12042924 | 1 | 197297417 | T | C | 0.5284 | -0.1807 | 0.0303 | 2.62E-09 | Yes | No |
| rs882624 | 1 | 201735913 | T | C | 0.332 | -0.0178 | 0.0324 | 0.5825 | No | No |
| rs33996239 | 1 | 203109801 | T | C | 0.0601 | -0.3655 | 0.0662 | 3.39E-08 | Yes | No |
| rs4245739 | 1 | 204518842 | A | C | 0.7284 | 0.0949 | 0.034 | 0.005305 | Yes | No |
| rs2629665 | 1 | 207220800 | A | C | 0.4101 | -0.1795 | 0.0309 | 6.03E-09 | Yes | No |
| rs2761436 | 1 | 207919748 | T | C | 0.5368 | 0.1839 | 0.0302 | 1.15E-09 | Yes | No |
| rs7555285 | 1 | 209970355 | C | G | 0.8011 | 0.2294 | 0.0376 | 1.05E-09 | Yes | No |
| rs12408022 | 1 | 217718789 | T | C | 0.2587 | 0.1939 | 0.0347 | 2.41E-08 | Yes | No |
| rs35981664 | 1 | 218549354 | A | T | 0.6879 | -0.1561 | 0.033 | 2.27E-06 | Yes | No |
| rs2820443 | 1 | 219753509 | T | C | 0.7089 | 0.1596 | 0.0331 | 1.40E-06 | Yes | No |
| rs9431431 | 1 | 221358796 | A | G | 0.294 | -0.2107 | 0.0331 | 1.97E-10 | Yes | No |
| rs73091767 | 1 | 227250775 | T | C | 0.7343 | -0.179 | 0.0339 | 1.35E-07 | Yes | No |
| rs2004776 | 1 | 230848702 | T | C | 0.2397 | 0.3789 | 0.0354 | 8.37E-27 | Yes | No |
| rs6429422 | 1 | 243472801 | T | G | 0.6769 | -0.1532 | 0.0323 | 2.10E-06 | Yes | No |
| rs4926499 | 1 | 249155909 | C | G | 0.8263 | 0.2965 | 0.0438 | 1.33E-11 | No | No |
| rs4850047 | 2 | 3634753 | T | C | 0.1468 | -0.132 | 0.0446 | 0.003067 | Yes | No |
| rs2175337 | 2 | 9298590 | A | C | 0.6104 | 0.146 | 0.031 | 2.53E-06 | Yes | No |
| rs67720684 | 2 | 18975439 | A | C | 0.2385 | 0.1943 | 0.0353 | 3.85E-08 | Yes | No |
| rs1344653 | 2 | 19730845 | A | G | 0.499 | -0.2534 | 0.03 | 3.09E-17 | Yes | No |
| rs7255 | 2 | 20878820 | T | C | 0.454 | -0.1518 | 0.0305 | 6.42E-07 | Yes | No |
| rs66774912 | 2 | 21423532 | A | G | 0.1371 | -0.1645 | 0.0441 | 0.0001928 | Yes | No |
| rs10779936 | 2 | 23950200 | A | G | 0.7129 | -0.1413 | 0.0332 | 2.15E-05 | Yes | No |
| rs55701159 | 2 | 25139596 | T | G | 0.8867 | 0.3959 | 0.0481 | 1.73E-16 | Yes | No |
| rs1275988 | 2 | 26914364 | T | C | 0.6112 | -0.541 | 0.0308 | 4.42E-69 | Yes | Yes |
| rs9678851 | 2 | 27887034 | A | C | 0.5675 | -0.1722 | 0.0307 | 1.99E-08 | Yes | No |
| rs7562 | 2 | 28635740 | T | C | 0.5207 | 0.2313 | 0.0305 | 3.26E-14 | Yes | Yes |
| rs1607644 | 2 | 34679626 | A | G | 0.3664 | -0.1274 | 0.0313 | 4.61E-05 | Yes | No |
| rs13420463 | 2 | 37517566 | A | G | 0.7734 | 0.3143 | 0.036 | 2.72E-18 | Yes | No |
| rs2707238 | 2 | 38094149 | C | G | 0.284 | 0.0927 | 0.0335 | 0.005663 | Yes | No |
| rs4952611 | 2 | 40567743 | T | C | 0.5795 | -0.211 | 0.0315 | 1.97E-11 | Yes | No |
| rs11681462 | 2 | 42352567 | A | C | 0.7891 | -0.1776 | 0.0374 | 2.00E-06 | Yes | No |
| rs76326501 | 2 | 43167878 | A | C | 0.9092 | 0.5681 | 0.0533 | 1.63E-26 | Yes | No |
| rs35590893 | 2 | 43716933 | A | G | 0.2773 | -0.2374 | 0.0336 | 1.66E-12 | Yes | No |
| rs11690961 | 2 | 46363336 | A | C | 0.8833 | 0.1388 | 0.047 | 0.003158 | Yes | No |
| rs6545155 | 2 | 50429861 | T | C | 0.7817 | 0.1821 | 0.0364 | 5.78E-07 | Yes | Yes |
| rs10189186 | 2 | 53025757 | A | G | 0.53 | 0.1893 | 0.0302 | 3.91E-10 | Yes | No |
| rs2920899 | 2 | 55279681 | T | G | 0.7864 | 0.1986 | 0.0372 | 9.45E-08 | Yes | No |
| rs1975487 | 2 | 55809054 | A | G | 0.4812 | -0.1986 | 0.0307 | 9.62E-11 | Yes | No |
| rs6730325 | 2 | 59315828 | A | G | 0.6053 | -0.1268 | 0.0308 | 3.81E-05 | Yes | No |
| rs72816333 | 2 | 60096560 | A | T | 0.8298 | 0.2336 | 0.04 | 5.45E-09 | Yes | No |
| rs925484 | 2 | 60611437 | C | G | 0.5998 | -0.1696 | 0.0308 | 3.66E-08 | Yes | No |
| rs7608483 | 2 | 61836235 | A | C | 0.4176 | 0.1383 | 0.0307 | 6.56E-06 | Yes | No |
| rs13014371 | 2 | 64217786 | T | C | 0.5705 | -0.1709 | 0.0305 | 1.99E-08 | Yes | No |
| rs2631669 | 2 | 66104881 | T | C | 0.471 | 0.0429 | 0.0302 | 0.1558 | Yes | No |
| rs2300481 | 2 | 66782467 | T | C | 0.3865 | 0.1981 | 0.031 | 1.56E-10 | Yes | Yes |
| rs6731373 | 2 | 68503044 | A | G | 0.3492 | 0.1913 | 0.0326 | 4.18E-09 | Yes | No |
| rs12052761 | 2 | 69065841 | A | G | 0.3936 | -0.1336 | 0.031 | 1.60E-05 | Yes | No |
| rs3771371 | 2 | 71627539 | T | C | 0.5692 | -0.1489 | 0.0304 | 9.72E-07 | Yes | No |
| rs10193543 | 2 | 72483329 | T | G | 0.8367 | 0.2023 | 0.0407 | 6.73E-07 | Yes | No |
| rs1876487 | 2 | 73114352 | A | C | 0.2939 | -0.1277 | 0.0342 | 0.000191 | Yes | No |
| rs11689667 | 2 | 85491365 | T | C | 0.5447 | 0.1983 | 0.0304 | 6.66E-11 | Yes | No |
| rs72847885 | 2 | 86326717 | A | G | 0.663 | 0.2413 | 0.0318 | 3.08E-14 | Yes | No |
| rs2579519 | 2 | 96675166 | T | C | 0.6161 | -0.0094 | 0.0314 | 0.7639 | Yes | No |
| rs4851462 | 2 | 98357163 | T | C | 0.6236 | -0.0706 | 0.0317 | 0.02578 | No | No |
| rs150194832 | 2 | 106126880 | C | G | 0.0934 | -0.2901 | 0.0523 | 2.88E-08 | Yes | No |
| rs28377357 | 2 | 112769721 | A | G | 0.2938 | -0.2142 | 0.0331 | 9.60E-11 | No | No |
| rs62158170 | 2 | 114082175 | A | G | 0.7834 | 0.2369 | 0.0368 | 1.21E-10 | Yes | No |
| rs10864859 | 2 | 121440018 | T | G | 0.9155 | 0.3089 | 0.0567 | 5.07E-08 | Yes | No |
| rs6723509 | 2 | 122000745 | T | C | 0.8609 | 0.2531 | 0.0438 | 7.61E-09 | Yes | No |
| rs13001283 | 2 | 127183454 | A | G | 0.1595 | 0.1816 | 0.0417 | 1.32E-05 | Yes | No |
| rs4954192 | 2 | 135632981 | T | C | 0.391 | -0.1796 | 0.0312 | 8.62E-09 | Yes | No |
| rs72844590 | 2 | 138421227 | T | G | 0.1497 | 0.2173 | 0.0432 | 4.95E-07 | Yes | No |
| rs7606205 | 2 | 144146311 | A | C | 0.7023 | -0.1843 | 0.0333 | 3.06E-08 | Yes | No |
| rs1438896 | 2 | 145646072 | T | C | 0.2982 | 0.208 | 0.0329 | 2.48E-10 | Yes | No |
| rs34570306 | 2 | 146272860 | T | C | 0.5278 | -0.1515 | 0.0309 | 9.08E-07 | Yes | No |
| rs62169544 | 2 | 146950908 | A | G | 0.4421 | -0.1885 | 0.0305 | 6.21E-10 | Yes | No |
| rs12990959 | 2 | 148572160 | T | C | 0.6866 | -0.097 | 0.0326 | 0.002953 | Yes | No |
| rs4664080 | 2 | 152978341 | A | G | 0.3975 | -0.1451 | 0.0308 | 2.44E-06 | Yes | No |
| rs3175 | 2 | 153618773 | A | G | 0.6519 | 0.16 | 0.0324 | 7.70E-07 | Yes | No |
| rs1220128 | 2 | 158499902 | C | G | 0.8519 | 0.2034 | 0.0429 | 2.12E-06 | Yes | No |
| rs79523138 | 2 | 161368213 | A | G | 0.8807 | -0.2469 | 0.0474 | 1.85E-07 | Yes | No |
| rs55732192 | 2 | 162278233 | T | G | 0.0947 | -0.3358 | 0.0521 | 1.15E-10 | Yes | No |
| rs1446468 | 2 | 164963486 | T | C | 0.4538 | -0.5063 | 0.0304 | 4.38E-62 | Yes | Yes |
| rs6712203 | 2 | 165557318 | T | C | 0.3723 | -0.2092 | 0.0313 | 2.41E-11 | Yes | No |
| rs2390258 | 2 | 166250129 | A | G | 0.3074 | -0.1309 | 0.0328 | 6.55E-05 | Yes | No |
| rs560887 | 2 | 169763148 | T | C | 0.299 | -0.1572 | 0.0329 | 1.79E-06 | Yes | No |
| rs151054210 | 2 | 172381487 | A | G | 0.1825 | 0.1823 | 0.039 | 3.00E-06 | Yes | No |
| rs6758859 | 2 | 173965056 | T | C | 0.6357 | 0.0898 | 0.0313 | 0.0041 | Yes | No |
| rs11694601 | 2 | 174949358 | A | G | 0.5968 | -0.1909 | 0.0309 | 6.41E-10 | Yes | No |
| rs72914576 | 2 | 175529967 | C | G | 0.8099 | -0.292 | 0.0388 | 5.38E-14 | Yes | No |
| rs60148403 | 2 | 177989414 | A | T | 0.1955 | -0.2043 | 0.0385 | 1.12E-07 | Yes | No |
| rs1837164 | 2 | 178716601 | A | T | 0.3697 | 0.1824 | 0.0311 | 4.66E-09 | Yes | Yes |
| rs79146658 | 2 | 179786068 | T | C | 0.9147 | -0.074 | 0.0545 | 0.1742 | Yes | No |
| rs1486236 | 2 | 180739450 | A | C | 0.3702 | -0.2339 | 0.0321 | 2.96E-13 | Yes | Yes |
| rs10184839 | 2 | 181946115 | A | T | 0.2914 | -0.1853 | 0.0332 | 2.40E-08 | Yes | No |
| rs16823124 | 2 | 183224127 | A | G | 0.3066 | 0.236 | 0.0326 | 4.59E-13 | Yes | No |
| rs12473688 | 2 | 185033470 | A | G | 0.2844 | 0.1805 | 0.0334 | 6.47E-08 | Yes | No |
| rs28558491 | 2 | 187816321 | T | C | 0.7336 | -0.2114 | 0.0343 | 7.54E-10 | Yes | No |
| rs11901929 | 2 | 189643316 | A | G | 0.3462 | 0.0908 | 0.0318 | 0.004324 | Yes | No |
| rs7592578 | 2 | 191439591 | T | G | 0.1947 | -0.2857 | 0.0391 | 2.79E-13 | Yes | No |
| rs296797 | 2 | 201102905 | T | C | 0.405 | 0.2161 | 0.0308 | 2.16E-12 | Yes | Yes |
| rs1469760 | 2 | 204125426 | T | C | 0.5835 | -0.2485 | 0.0307 | 5.87E-16 | Yes | No |
| rs2162003 | 2 | 205077128 | T | C | 0.6042 | 0.1985 | 0.0321 | 6.21E-10 | No | No |
| rs1263671 | 2 | 207996447 | T | C | 0.8367 | -0.1298 | 0.0415 | 0.001754 | Yes | No |
| rs55780018 | 2 | 208526140 | T | C | 0.5434 | -0.2784 | 0.031 | 2.73E-19 | Yes | No |
| rs1047891 | 2 | 211540507 | A | C | 0.3193 | -0.2528 | 0.0328 | 1.37E-14 | Yes | No |
| rs12694277 | 2 | 213188795 | T | C | 0.2946 | -0.2018 | 0.0335 | 1.80E-09 | Yes | No |
| rs4674114 | 2 | 217659266 | A | G | 0.2006 | -0.1773 | 0.0377 | 2.53E-06 | Yes | No |
| rs1063281 | 2 | 218668732 | T | C | 0.6031 | -0.2585 | 0.0313 | 1.40E-16 | Yes | No |
| rs1996992 | 2 | 219651349 | T | G | 0.0514 | -0.1248 | 0.0687 | 0.06902 | Yes | No |
| rs12474050 | 2 | 220362557 | T | C | 0.3433 | 0.0647 | 0.0318 | 0.04184 | Yes | No |
| rs2972146 | 2 | 227100698 | T | G | 0.6425 | 0.2532 | 0.0313 | 6.53E-16 | Yes | No |
| rs1044822 | 2 | 230629138 | T | C | 0.1482 | -0.248 | 0.0424 | 5.16E-09 | Yes | No |
| rs12052878 | 2 | 238227594 | A | G | 0.32 | -0.1163 | 0.0324 | 0.0003356 | Yes | No |
| rs4507125 | 2 | 239864732 | A | C | 0.7858 | -0.121 | 0.0368 | 0.0009911 | Yes | No |
| rs139354822 | 2 | 242344695 | T | C | 0.9704 | 0.6115 | 0.0975 | 3.51E-10 | Yes | No |
| rs9865843 | 3 | 7489993 | A | G | 0.518 | -0.1368 | 0.0308 | 8.82E-06 | Yes | No |
| rs347591 | 3 | 11290122 | T | G | 0.6625 | 0.3181 | 0.032 | 2.87E-23 | Yes | Yes |
| rs729639 | 3 | 13826854 | T | C | 0.3453 | -0.113 | 0.0319 | 0.0003941 | Yes | No |
| rs11128722 | 3 | 14958126 | A | G | 0.5696 | -0.2865 | 0.0309 | 1.93E-20 | Yes | No |
| rs189267552 | 3 | 20073193 | A | T | 0.0132 | -0.8664 | 0.139 | 4.55E-10 | Yes | No |
| rs4634143 | 3 | 23163749 | T | C | 0.3002 | 0.1482 | 0.0329 | 6.79E-06 | Yes | No |
| rs13082711 | 3 | 27537909 | T | C | 0.7617 | -0.3087 | 0.0354 | 2.78E-18 | Yes | No |
| rs72851229 | 3 | 29374219 | C | G | 0.1746 | -0.1793 | 0.0403 | 8.79E-06 | Yes | No |
| rs12638085 | 3 | 30405936 | A | T | 0.3559 | 0.2189 | 0.0318 | 5.61E-12 | Yes | Yes |
| rs4678915 | 3 | 36964583 | A | G | 0.4315 | -0.1377 | 0.0311 | 9.44E-06 | Yes | No |
| rs267517 | 3 | 37539090 | A | G | 0.5916 | -0.261 | 0.0313 | 7.18E-17 | Yes | No |
| rs6801957 | 3 | 38767315 | T | C | 0.4101 | 0.0868 | 0.0306 | 0.004561 | Yes | No |
| rs6788984 | 3 | 41107173 | A | G | 0.8563 | 0.2999 | 0.0432 | 3.81E-12 | Yes | Yes |
| rs9815354 | 3 | 41912651 | A | G | 0.1641 | -0.2166 | 0.0413 | 1.52E-07 | Yes | No |
| rs141979279 | 3 | 44858131 | T | C | 0.9509 | 0.2883 | 0.0699 | 3.75E-05 | Yes | No |
| rs113134141 | 3 | 46861939 | A | G | 0.8983 | -0.2732 | 0.0504 | 5.75E-08 | Yes | No |
| rs6797587 | 3 | 48197614 | A | G | 0.3279 | -0.3271 | 0.0322 | 3.40E-24 | Yes | No |
| rs73082337 | 3 | 49009570 | C | G | 0.8791 | 0.2795 | 0.0481 | 6.40E-09 | Yes | No |
| rs36022378 | 3 | 49913705 | T | C | 0.8004 | -0.1925 | 0.0383 | 4.99E-07 | Yes | No |
| rs13303 | 3 | 52558008 | T | C | 0.4365 | -0.0957 | 0.0307 | 0.001803 | Yes | No |
| rs9810888 | 3 | 53635595 | T | G | 0.4984 | -0.172 | 0.0304 | 1.60E-08 | Yes | No |
| rs9827472 | 3 | 56726646 | T | C | 0.3635 | -0.169 | 0.0315 | 8.10E-08 | No | No |
| rs12486605 | 3 | 57706503 | T | C | 0.5726 | -0.0832 | 0.0307 | 0.006754 | Yes | No |
| rs3774702 | 3 | 63856870 | A | G | 0.1766 | 0.0514 | 0.0398 | 0.197 | Yes | No |
| rs918466 | 3 | 64710253 | A | G | 0.4096 | -0.067 | 0.0309 | 0.03036 | Yes | No |
| rs7630745 | 3 | 66427029 | T | C | 0.6588 | 0.1692 | 0.0317 | 9.36E-08 | Yes | No |
| rs4499560 | 3 | 70920485 | A | T | 0.3171 | -0.2199 | 0.0326 | 1.46E-11 | Yes | No |
| rs729448 | 3 | 73260545 | A | G | 0.5472 | -0.1213 | 0.0304 | 6.62E-05 | Yes | No |
| rs9857362 | 3 | 74710462 | A | C | 0.5291 | 0.1727 | 0.0306 | 1.62E-08 | Yes | No |
| rs1375564 | 3 | 85656311 | T | C | 0.6395 | 0.2579 | 0.0315 | 2.84E-16 | Yes | No |
| rs9860290 | 3 | 99839106 | A | G | 0.2093 | -0.1386 | 0.0371 | 0.0001862 | Yes | No |
| rs11923667 | 3 | 101268080 | A | T | 0.407 | 0.0914 | 0.0309 | 0.003052 | No | No |
| rs28675079 | 3 | 111500002 | A | G | 0.1876 | -0.1762 | 0.0387 | 5.32E-06 | Yes | No |
| rs1882289 | 3 | 114461208 | A | G | 0.8846 | -0.2089 | 0.0471 | 9.25E-06 | Yes | No |
| rs6806529 | 3 | 123049938 | A | C | 0.4338 | 0.1768 | 0.0309 | 1.03E-08 | Yes | No |
| rs6438857 | 3 | 124557643 | T | C | 0.5774 | 0.2736 | 0.0305 | 3.13E-19 | Yes | No |
| rs62270945 | 3 | 128201889 | T | C | 0.0289 | 0.4362 | 0.096 | 5.51E-06 | Yes | No |
| rs9875380 | 3 | 132780356 | T | C | 0.4638 | -0.1752 | 0.0302 | 6.53E-09 | Yes | No |
| rs6783086 | 3 | 133959552 | T | C | 0.3954 | 0.3048 | 0.0308 | 5.13E-23 | Yes | No |
| rs590198 | 3 | 135953729 | A | G | 0.5286 | 0.1871 | 0.0301 | 5.11E-10 | Yes | No |
| rs2306374 | 3 | 138119952 | T | C | 0.838 | -0.2825 | 0.0411 | 6.11E-12 | Yes | No |
| rs16851397 | 3 | 141134818 | A | G | 0.953 | -0.5294 | 0.0726 | 2.96E-13 | Yes | No |
| rs62278541 | 3 | 142631909 | A | G | 0.6473 | 0.137 | 0.0316 | 1.43E-05 | Yes | No |
| rs6772704 | 3 | 149524692 | A | C | 0.6842 | -0.1237 | 0.0325 | 0.0001394 | Yes | No |
| rs73158427 | 3 | 153721493 | A | T | 0.1618 | 0.2804 | 0.0409 | 7.24E-12 | Yes | No |
| rs143112823 | 3 | 154707967 | A | G | 0.0876 | -0.4171 | 0.0557 | 7.16E-14 | Yes | No |
| rs9833313 | 3 | 157576791 | A | T | 0.2418 | -0.1907 | 0.0356 | 8.43E-08 | Yes | Yes |
| rs78151625 | 3 | 158316726 | T | C | 0.8344 | -0.2453 | 0.0407 | 1.61E-09 | Yes | No |
| rs419076 | 3 | 169100886 | T | C | 0.4732 | 0.4049 | 0.0301 | 3.00E-41 | Yes | No |
| rs4894535 | 3 | 171995605 | T | C | 0.1601 | 0.1883 | 0.0414 | 5.42E-06 | Yes | No |
| rs73171158 | 3 | 176927949 | T | C | 0.4672 | -0.1763 | 0.032 | 3.51E-08 | No | No |
| rs7611674 | 3 | 179169230 | T | G | 0.803 | 0.1184 | 0.0389 | 0.002348 | Yes | No |
| rs262986 | 3 | 183435713 | A | G | 0.4704 | -0.2371 | 0.0305 | 7.67E-15 | Yes | No |
| rs12374077 | 3 | 185317674 | C | G | 0.3443 | 0.2143 | 0.0317 | 1.48E-11 | Yes | No |
| rs1706003 | 3 | 194299967 | T | G | 0.4682 | 0.0276 | 0.0315 | 0.3807 | No | No |
| rs6777317 | 3 | 197070959 | A | G | 0.2911 | 0.1843 | 0.034 | 5.86E-08 | No | No |
| rs1250129 | 4 | 1254930 | A | G | 0.1149 | -0.2057 | 0.0474 | 1.46E-05 | Yes | No |
| rs55829085 | 4 | 2165493 | A | C | 0.9546 | -0.4309 | 0.074 | 5.73E-09 | Yes | No |
| rs231708 | 4 | 2694773 | C | G | 0.6921 | -0.2824 | 0.0326 | 4.74E-18 | Yes | No |
| rs2498323 | 4 | 3451109 | A | G | 0.098 | 0.3171 | 0.0517 | 8.52E-10 | Yes | No |
| rs3822239 | 4 | 10095539 | A | T | 0.6971 | 0.1473 | 0.0328 | 7.04E-06 | Yes | No |
| rs13122790 | 4 | 15356795 | A | G | 0.7316 | 0.1161 | 0.0341 | 0.0006688 | Yes | No |
| rs11730129 | 4 | 16032948 | T | C | 0.2183 | -0.0824 | 0.0367 | 0.02474 | Yes | No |
| rs2610990 | 4 | 18008232 | A | G | 0.2641 | -0.2903 | 0.0343 | 2.86E-17 | Yes | No |
| rs28667801 | 4 | 26785356 | A | T | 0.5923 | -0.2627 | 0.0314 | 5.80E-17 | No | No |
| rs1878825 | 4 | 36091370 | C | G | 0.6416 | -0.1215 | 0.0319 | 0.0001366 | Yes | No |
| rs2291435 | 4 | 38387395 | T | C | 0.5335 | -0.262 | 0.0303 | 5.31E-18 | Yes | Yes |
| rs12511987 | 4 | 46595623 | T | G | 0.8226 | -0.2329 | 0.0399 | 5.39E-09 | Yes | No |
| rs871606 | 4 | 54799245 | T | C | 0.895 | 0.3705 | 0.0495 | 7.25E-14 | Yes | No |
| rs1718845 | 4 | 57943153 | A | G | 0.2978 | -0.0938 | 0.0331 | 0.00457 | Yes | No |
| rs6551716 | 4 | 63575696 | A | T | 0.856 | 0.2137 | 0.0438 | 1.05E-06 | Yes | No |
| rs10008637 | 4 | 77414144 | T | C | 0.5405 | 0.2157 | 0.0302 | 9.24E-13 | Yes | No |
| rs1458038 | 4 | 81164723 | T | C | 0.294 | 0.8233 | 0.0333 | 3.82E-135 | Yes | No |
| rs6823199 | 4 | 83925895 | T | C | 0.7438 | 0.2094 | 0.0348 | 1.72E-09 | Yes | No |
| rs2014912 | 4 | 86715670 | T | C | 0.1528 | 0.4772 | 0.042 | 6.97E-30 | Yes | Yes |
| rs13149209 | 4 | 89750668 | T | C | 0.7773 | 0.281 | 0.0367 | 1.97E-14 | Yes | No |
| rs7694000 | 4 | 95324968 | A | T | 0.5394 | -0.0636 | 0.0305 | 0.0368 | No | No |
| rs1347345 | 4 | 95938386 | A | G | 0.6179 | -0.1806 | 0.0312 | 6.92E-09 | Yes | Yes |
| rs17248480 | 4 | 102435265 | A | G | 0.0283 | -0.8442 | 0.0966 | 2.37E-18 | Yes | No |
| rs13107325 | 4 | 103188709 | T | C | 0.0739 | -0.9086 | 0.0592 | 4.22E-53 | Yes | No |
| rs223361 | 4 | 103769304 | T | C | 0.6651 | 0.0477 | 0.0321 | 0.1368 | Yes | No |
| rs4699165 | 4 | 106109381 | A | G | 0.3565 | 0.1069 | 0.0315 | 0.0006875 | Yes | No |
| rs13112725 | 4 | 106911742 | C | G | 0.7631 | 0.4137 | 0.0358 | 6.81E-31 | Yes | No |
| rs7694643 | 4 | 109017528 | A | G | 0.6455 | -0.1953 | 0.0315 | 5.77E-10 | Yes | No |
| rs6825911 | 4 | 111381638 | T | C | 0.7915 | -0.3066 | 0.0375 | 3.05E-16 | Yes | No |
| rs4834735 | 4 | 119958809 | T | C | 0.1366 | 0.1843 | 0.0443 | 3.16E-05 | Yes | No |
| rs66887589 | 4 | 120509279 | T | C | 0.5216 | -0.1951 | 0.0303 | 1.20E-10 | Yes | No |
| rs3097937 | 4 | 124794644 | A | T | 0.8026 | 0.2226 | 0.0381 | 4.95E-09 | Yes | No |
| rs7439567 | 4 | 138464842 | T | C | 0.4106 | 0.2537 | 0.0309 | 2.31E-16 | Yes | Yes |
| rs72719160 | 4 | 144051276 | A | T | 0.6829 | -0.2243 | 0.0324 | 4.34E-12 | Yes | No |
| rs4292285 | 4 | 145271954 | A | T | 0.402 | -0.1397 | 0.0308 | 5.63E-06 | Yes | No |
| rs4835266 | 4 | 146821725 | T | C | 0.5217 | 0.2155 | 0.0309 | 3.07E-12 | Yes | No |
| rs10305838 | 4 | 148400256 | T | C | 0.8592 | -0.2659 | 0.0433 | 7.94E-10 | Yes | No |
| rs6823767 | 4 | 151295085 | T | C | 0.7226 | -0.2129 | 0.0341 | 4.40E-10 | Yes | No |
| rs13139571 | 4 | 156645513 | A | C | 0.2366 | -0.3299 | 0.0355 | 1.33E-20 | Yes | No |
| rs17035181 | 4 | 157678511 | T | G | 0.8552 | 0.3074 | 0.0429 | 7.61E-13 | Yes | No |
| rs869396 | 4 | 169688000 | A | C | 0.4659 | -0.2115 | 0.0305 | 4.12E-12 | Yes | No |
| rs4957026 | 5 | 361148 | A | G | 0.3399 | 0.1982 | 0.0323 | 8.12E-10 | Yes | No |
| rs10069690 | 5 | 1279790 | T | C | 0.2582 | 0.3098 | 0.0369 | 4.47E-17 | Yes | Yes |
| rs954767 | 5 | 3706050 | A | C | 0.7413 | -0.2127 | 0.0346 | 7.53E-10 | Yes | No |
| rs1173771 | 5 | 32815028 | A | G | 0.3987 | -0.6321 | 0.0307 | 5.92E-94 | Yes | Yes |
| rs74774746 | 5 | 33411769 | C | G | 0.2609 | -0.1902 | 0.035 | 5.60E-08 | Yes | No |
| rs7710854 | 5 | 43824677 | A | G | 0.8834 | 0.2244 | 0.0479 | 2.76E-06 | Yes | No |
| rs6867399 | 5 | 52135543 | A | C | 0.2671 | 0.1576 | 0.0357 | 1.02E-05 | Yes | No |
| rs1694068 | 5 | 53283630 | A | T | 0.6139 | 0.2657 | 0.0311 | 1.18E-17 | Yes | No |
| rs13179413 | 5 | 55868097 | T | C | 0.2819 | 0.2238 | 0.0347 | 1.08E-10 | Yes | No |
| rs12515541 | 5 | 57095011 | T | G | 0.6081 | 0.1582 | 0.0308 | 2.89E-07 | Yes | No |
| rs1848510 | 5 | 57754005 | A | G | 0.3621 | 0.1839 | 0.0316 | 5.72E-09 | Yes | No |
| rs10062049 | 5 | 61553881 | T | C | 0.1358 | 0.2732 | 0.0445 | 8.02E-10 | Yes | No |
| rs6875372 | 5 | 64079015 | A | T | 0.5151 | 0.1886 | 0.0303 | 4.80E-10 | No | No |
| rs3121685 | 5 | 65662133 | T | C | 0.4838 | -0.1563 | 0.0302 | 2.28E-07 | Yes | Yes |
| rs4286632 | 5 | 66291370 | A | G | 0.7306 | 0.211 | 0.0343 | 7.64E-10 | Yes | No |
| rs246973 | 5 | 68007803 | T | C | 0.2882 | 0.2479 | 0.0335 | 1.45E-13 | Yes | Yes |
| rs72761109 | 5 | 71506529 | T | C | 0.3026 | 0.1501 | 0.0329 | 4.93E-06 | Yes | No |
| rs4443403 | 5 | 72654304 | T | C | 0.9031 | -0.2496 | 0.0513 | 1.13E-06 | Yes | No |
| rs10078021 | 5 | 75038431 | T | G | 0.6274 | -0.0476 | 0.0317 | 0.1328 | Yes | No |
| rs10057188 | 5 | 77837789 | A | G | 0.4584 | -0.2275 | 0.0307 | 1.19E-13 | Yes | No |
| rs10059921 | 5 | 87514515 | T | G | 0.0825 | -0.4248 | 0.0587 | 4.39E-13 | Yes | No |
| rs62380354 | 5 | 89484911 | A | C | 0.8916 | 0.2161 | 0.0511 | 2.36E-05 | Yes | No |
| rs709668 | 5 | 96174186 | A | G | 0.2003 | -0.2943 | 0.0377 | 5.96E-15 | Yes | Yes |
| rs1871190 | 5 | 97953719 | T | G | 0.3349 | 0.1954 | 0.0324 | 1.66E-09 | Yes | Yes |
| rs286809 | 5 | 107458637 | A | G | 0.1725 | -0.1748 | 0.0399 | 1.20E-05 | Yes | No |
| rs79409628 | 5 | 108113740 | T | G | 0.0845 | -0.2578 | 0.0543 | 2.04E-06 | Yes | No |
| rs10077885 | 5 | 114390121 | A | C | 0.5084 | -0.259 | 0.0308 | 4.43E-17 | Yes | Yes |
| rs1432457 | 5 | 119781569 | A | G | 0.7152 | -0.1561 | 0.0334 | 2.94E-06 | Yes | No |
| rs9885577 | 5 | 121194226 | T | C | 0.3709 | 0.2309 | 0.0321 | 6.53E-13 | Yes | No |
| rs13359291 | 5 | 122476457 | A | G | 0.1573 | 0.4333 | 0.0415 | 1.64E-25 | Yes | Yes |
| rs6891344 | 5 | 123136656 | A | G | 0.8179 | 0.2706 | 0.0395 | 6.96E-12 | Yes | No |
| rs62373688 | 5 | 127352807 | A | T | 0.1306 | 0.2742 | 0.0454 | 1.58E-09 | Yes | No |
| rs6595838 | 5 | 127868199 | A | G | 0.2985 | 0.3229 | 0.0331 | 1.54E-22 | Yes | No |
| rs12521868 | 5 | 131784393 | T | G | 0.4223 | -0.0872 | 0.0308 | 0.004612 | Yes | No |
| rs55747751 | 5 | 132397351 | A | G | 0.0807 | -0.2776 | 0.0582 | 1.82E-06 | Yes | No |
| rs702395 | 5 | 140086677 | T | C | 0.4369 | 0.2318 | 0.0305 | 3.24E-14 | Yes | No |
| rs1650911 | 5 | 141740620 | C | G | 0.7647 | 0.2549 | 0.0368 | 4.34E-12 | Yes | No |
| rs2400509 | 5 | 147696018 | A | G | 0.2583 | -0.1671 | 0.0344 | 1.21E-06 | Yes | No |
| rs9687065 | 5 | 148391140 | A | G | 0.8099 | 0.3356 | 0.0387 | 4.62E-18 | Yes | No |
| rs157678 | 5 | 156145654 | A | T | 0.6689 | -0.183 | 0.0329 | 2.66E-08 | Yes | No |
| rs11953630 | 5 | 157845402 | T | C | 0.3659 | -0.4529 | 0.032 | 1.55E-45 | Yes | No |
| rs62385385 | 5 | 158367249 | A | T | 0.3829 | 0.2834 | 0.031 | 6.30E-20 | Yes | No |
| rs114503346 | 5 | 172192350 | T | C | 0.0458 | -0.1801 | 0.0746 | 0.01576 | Yes | No |
| rs72812846 | 5 | 173377636 | A | T | 0.278 | -0.2579 | 0.0344 | 6.91E-14 | Yes | No |
| rs28362590 | 5 | 176731452 | T | G | 0.7547 | 0.1302 | 0.0354 | 0.0002299 | Yes | No |
| rs12153395 | 5 | 179411477 | A | G | 0.1147 | -0.3303 | 0.0486 | 1.07E-11 | Yes | No |
| rs2745599 | 6 | 1613686 | A | G | 0.552 | 0.2164 | 0.0317 | 8.96E-12 | Yes | No |
| rs1334576 | 6 | 7211818 | A | G | 0.4198 | -0.0706 | 0.0306 | 0.02107 | Yes | No |
| rs9392172 | 6 | 7723962 | C | G | 0.5357 | -0.1969 | 0.0302 | 7.31E-11 | No | No |
| rs114275780 | 6 | 8224648 | A | T | 0.0592 | 0.3039 | 0.0675 | 6.78E-06 | Yes | No |
| rs1630736 | 6 | 12295987 | T | C | 0.465 | -0.1706 | 0.0309 | 3.52E-08 | No | No |
| rs9349379 | 6 | 12903957 | A | G | 0.593 | 0.2664 | 0.0312 | 1.31E-17 | Yes | No |
| rs12216497 | 6 | 19028623 | T | C | 0.5622 | 0.1551 | 0.0304 | 3.28E-07 | Yes | No |
| rs9368222 | 6 | 20686996 | A | C | 0.2688 | 0.2281 | 0.0339 | 1.84E-11 | Yes | No |
| rs6911827 | 6 | 22130601 | T | C | 0.4577 | 0.2378 | 0.0306 | 7.96E-15 | Yes | Yes |
| rs1799945 | 6 | 26091179 | C | G | 0.8505 | -0.5445 | 0.0426 | 1.83E-37 | Yes | No |
| rs409558 | 6 | 31708147 | T | C | 0.8487 | 0.3844 | 0.0431 | 4.66E-19 | Yes | No |
| rs115245297 | 6 | 34244132 | T | C | 0.9555 | -0.4453 | 0.0769 | 7.06E-09 | Yes | No |
| rs3176336 | 6 | 36648816 | A | T | 0.6092 | 0.1083 | 0.0314 | 0.0005583 | Yes | No |
| rs4714224 | 6 | 39186743 | C | G | 0.2779 | -0.0809 | 0.0341 | 0.01757 | Yes | No |
| rs1563788 | 6 | 43308363 | T | C | 0.2867 | 0.3385 | 0.0332 | 2.36E-24 | Yes | No |
| rs9472135 | 6 | 43809802 | T | C | 0.6986 | 0.0467 | 0.0332 | 0.1595 | Yes | No |
| rs78648104 | 6 | 50683009 | T | C | 0.9075 | -0.4287 | 0.0541 | 2.37E-15 | Yes | No |
| rs13205180 | 6 | 51832494 | T | C | 0.4877 | 0.1241 | 0.0303 | 4.31E-05 | Yes | No |
| rs631441 | 6 | 53994626 | T | G | 0.6949 | -0.1686 | 0.0327 | 2.48E-07 | Yes | No |
| rs1925153 | 6 | 56102780 | T | C | 0.4455 | -0.1193 | 0.0313 | 0.0001377 | Yes | No |
| rs504691 | 6 | 72206620 | A | C | 0.4005 | -0.0362 | 0.0309 | 0.2407 | Yes | No |
| rs12195276 | 6 | 73657714 | T | C | 0.7246 | -0.1672 | 0.034 | 9.05E-07 | Yes | No |
| rs10943605 | 6 | 79655477 | A | G | 0.4887 | 0.239 | 0.0302 | 2.72E-15 | Yes | No |
| rs7753695 | 6 | 80818531 | T | C | 0.4374 | 0.0979 | 0.0307 | 0.001441 | Yes | No |
| rs9449350 | 6 | 82281417 | T | C | 0.6795 | -0.2189 | 0.0323 | 1.19E-11 | Yes | No |
| rs60255247 | 6 | 85283253 | A | C | 0.8875 | 0.2191 | 0.0486 | 6.66E-06 | Yes | No |
| rs35410524 | 6 | 96885405 | T | C | 0.1885 | 0.3368 | 0.0387 | 3.19E-18 | Yes | No |
| rs9486916 | 6 | 109013930 | T | C | 0.1979 | 0.2657 | 0.0385 | 5.42E-12 | Yes | No |
| rs3822857 | 6 | 116313931 | C | G | 0.3664 | -0.0837 | 0.0314 | 0.007753 | Yes | No |
| rs2693560 | 6 | 117523671 | A | G | 0.366 | -0.206 | 0.0316 | 6.99E-11 | Yes | No |
| rs2498586 | 6 | 118026126 | T | C | 0.1652 | -0.2073 | 0.0412 | 5.02E-07 | Yes | Yes |
| rs9372498 | 6 | 118572486 | A | T | 0.0812 | 0.3967 | 0.0556 | 9.43E-13 | Yes | No |
| rs9401090 | 6 | 119113317 | T | C | 0.7457 | 0.1575 | 0.0349 | 6.29E-06 | Yes | No |
| rs11154027 | 6 | 121781390 | T | C | 0.4606 | 0.063 | 0.0307 | 0.03993 | Yes | No |
| rs6925750 | 6 | 122287990 | T | C | 0.8858 | 0.0896 | 0.0475 | 0.05928 | Yes | No |
| rs10782230 | 6 | 126228512 | A | G | 0.4845 | 0.2106 | 0.0302 | 2.91E-12 | Yes | Yes |
| rs13209747 | 6 | 127115454 | T | C | 0.4413 | 0.5117 | 0.0305 | 2.49E-63 | Yes | No |
| rs9885632 | 6 | 131311909 | T | C | 0.7309 | 0.2358 | 0.0341 | 4.37E-12 | Yes | Yes |
| rs4896104 | 6 | 135119089 | T | C | 0.5595 | -0.0703 | 0.0308 | 0.02258 | Yes | No |
| rs668459 | 6 | 139835689 | T | C | 0.5859 | -0.1292 | 0.0305 | 2.27E-05 | Yes | No |
| rs7763294 | 6 | 140383733 | T | G | 0.3163 | -0.2004 | 0.0324 | 6.39E-10 | Yes | No |
| rs6941056 | 6 | 143591821 | C | G | 0.5582 | 0.1476 | 0.0306 | 1.37E-06 | No | No |
| rs7765526 | 6 | 147713764 | A | G | 0.4633 | 0.201 | 0.0307 | 5.88E-11 | Yes | Yes |
| rs17080102 | 6 | 151004770 | C | G | 0.0694 | -0.8085 | 0.0594 | 3.52E-42 | Yes | No |
| rs9479200 | 6 | 152398505 | A | G | 0.8856 | -0.2562 | 0.0478 | 8.28E-08 | Yes | No |
| rs9479509 | 6 | 153427265 | A | G | 0.2959 | -0.1448 | 0.033 | 1.15E-05 | Yes | No |
| rs598682 | 6 | 154418759 | A | G | 0.2506 | -0.1566 | 0.0347 | 6.56E-06 | Yes | No |
| rs449789 | 6 | 159699125 | C | G | 0.139 | 0.3233 | 0.0438 | 1.50E-13 | Yes | No |
| rs555754 | 6 | 160769423 | A | G | 0.4749 | -0.1559 | 0.0302 | 2.41E-07 | Yes | No |
| rs9456648 | 6 | 161712235 | T | C | 0.3227 | -0.1771 | 0.0322 | 3.76E-08 | Yes | No |
| rs73030266 | 6 | 166179459 | A | T | 0.9331 | 0.4393 | 0.062 | 1.42E-12 | Yes | No |
| rs4342401 | 6 | 169016615 | A | T | 0.5414 | 0.0639 | 0.0304 | 0.03573 | No | No |
| rs1322639 | 6 | 169587103 | A | G | 0.7761 | 0.1535 | 0.0365 | 2.59E-05 | Yes | No |
| rs6959688 | 7 | 1966831 | A | G | 0.5981 | -0.2344 | 0.031 | 4.22E-14 | Yes | No |
| rs2969070 | 7 | 2512545 | A | G | 0.6313 | -0.2726 | 0.0313 | 2.86E-18 | Yes | No |
| rs73049928 | 7 | 4669949 | A | G | 0.8061 | -0.2382 | 0.0392 | 1.20E-09 | Yes | No |
| rs1468520 | 7 | 7290732 | A | G | 0.8321 | -0.1874 | 0.0407 | 4.21E-06 | Yes | No |
| rs13240040 | 7 | 14375977 | A | G | 0.6841 | 0.1753 | 0.0331 | 1.18E-07 | Yes | No |
| rs2107595 | 7 | 19049388 | A | G | 0.159 | 0.4176 | 0.0415 | 7.37E-24 | Yes | No |
| rs4507656 | 7 | 22156538 | C | G | 0.6934 | -0.2167 | 0.0351 | 6.78E-10 | No | No |
| rs2069833 | 7 | 22767664 | T | C | 0.5746 | -0.1988 | 0.0306 | 8.34E-11 | Yes | No |
| rs12979 | 7 | 24738164 | C | G | 0.8698 | 0.2739 | 0.0449 | 1.09E-09 | Yes | Yes |
| rs1055144 | 7 | 25871109 | T | C | 0.1927 | 0.11 | 0.0382 | 0.003985 | Yes | No |
| rs6969780 | 7 | 27159136 | C | G | 0.0918 | 0.2957 | 0.0526 | 1.88E-08 | Yes | No |
| rs7777128 | 7 | 27337113 | C | G | 0.0798 | 0.5158 | 0.0557 | 2.06E-20 | Yes | No |
| rs10274928 | 7 | 28142088 | A | G | 0.4845 | 0.1641 | 0.0306 | 8.15E-08 | Yes | No |
| rs917275 | 7 | 28658522 | A | G | 0.6099 | -0.1726 | 0.0312 | 3.26E-08 | Yes | No |
| rs10233127 | 7 | 30933453 | A | T | 0.1076 | 0.2555 | 0.0507 | 4.74E-07 | Yes | No |
| rs342989 | 7 | 35467896 | A | G | 0.2282 | 0.0914 | 0.036 | 0.0112 | Yes | No |
| rs2052263 | 7 | 36225818 | A | G | 0.1177 | 0.2543 | 0.0483 | 1.40E-07 | Yes | No |
| rs76206723 | 7 | 40447971 | A | G | 0.107 | -0.4037 | 0.0491 | 2.02E-16 | Yes | No |
| rs12701929 | 7 | 41722494 | T | C | 0.2498 | 0.1075 | 0.035 | 0.002107 | Yes | No |
| rs1004558 | 7 | 44240407 | T | C | 0.1782 | 0.1639 | 0.0398 | 3.87E-05 | Yes | No |
| rs73105827 | 7 | 45036785 | T | G | 0.0903 | -0.2232 | 0.0547 | 4.51E-05 | Yes | No |
| rs11977526 | 7 | 46008110 | A | G | 0.4009 | -0.3213 | 0.0312 | 6.62E-25 | Yes | No |
| rs71543920 | 7 | 46554358 | T | G | 0.9398 | -0.2142 | 0.0642 | 0.0008562 | Yes | No |
| rs12668436 | 7 | 47548893 | T | C | 0.7541 | -0.2151 | 0.035 | 7.88E-10 | Yes | No |
| rs17454517 | 7 | 50915776 | A | G | 0.4926 | 0.0276 | 0.0303 | 0.3629 | Yes | No |
| rs6593297 | 7 | 56122058 | A | T | 0.3029 | 0.1674 | 0.0337 | 6.55E-07 | Yes | No |
| rs2222544 | 7 | 69769369 | T | C | 0.737 | -0.1419 | 0.0342 | 3.36E-05 | Yes | No |
| rs1091811 | 7 | 73491212 | A | G | 0.1688 | -0.1554 | 0.0405 | 0.0001262 | Yes | No |
| rs34324971 | 7 | 74107374 | A | G | 0.1846 | 0.1515 | 0.0408 | 0.0002019 | Yes | No |
| rs6963105 | 7 | 75097488 | A | G | 0.4388 | -0.1892 | 0.0321 | 3.83E-09 | Yes | No |
| rs848445 | 7 | 77572461 | T | C | 0.2851 | -0.2025 | 0.0339 | 2.28E-09 | Yes | No |
| rs11770630 | 7 | 89805241 | T | C | 0.5355 | 0.0386 | 0.0302 | 0.2014 | Yes | No |
| rs10245696 | 7 | 90449362 | A | C | 0.4003 | 0.1695 | 0.0308 | 3.59E-08 | Yes | No |
| rs2282978 | 7 | 92264410 | T | C | 0.6677 | 0.2802 | 0.0321 | 2.51E-18 | Yes | No |
| rs1947228 | 7 | 96461649 | T | C | 0.418 | -0.1756 | 0.0308 | 1.19E-08 | Yes | No |
| rs12705090 | 7 | 100467700 | T | C | 0.1896 | -0.0696 | 0.0386 | 0.07122 | Yes | No |
| rs17477177 | 7 | 106411858 | T | C | 0.796 | -0.7351 | 0.0375 | 9.01E-86 | Yes | Yes |
| rs115172170 | 7 | 107019947 | T | C | 0.9441 | -0.3136 | 0.0662 | 2.14E-06 | Yes | No |
| rs1997571 | 7 | 116198621 | A | G | 0.5911 | -0.1553 | 0.0307 | 4.09E-07 | Yes | No |
| rs4728142 | 7 | 128573967 | A | G | 0.4439 | -0.1814 | 0.0305 | 2.59E-09 | Yes | Yes |
| rs11556924 | 7 | 129663496 | T | C | 0.3818 | -0.2097 | 0.0317 | 3.43E-11 | Yes | No |
| rs34072724 | 7 | 130432469 | A | G | 0.4889 | -0.2422 | 0.0303 | 1.37E-15 | Yes | No |
| rs13238550 | 7 | 131059056 | A | G | 0.3964 | 0.2572 | 0.0309 | 7.80E-17 | Yes | No |
| rs1722886 | 7 | 134215259 | A | T | 0.5666 | 0.2072 | 0.0305 | 1.09E-11 | No | No |
| rs10267979 | 7 | 136618188 | A | T | 0.3245 | -0.1197 | 0.0322 | 0.000198 | Yes | No |
| rs7810028 | 7 | 139461616 | C | G | 0.8068 | 0.2835 | 0.0382 | 1.24E-13 | Yes | No |
| rs12703989 | 7 | 140238048 | A | G | 0.4991 | 0.1531 | 0.0306 | 5.82E-07 | Yes | No |
| rs73727605 | 7 | 149474622 | A | G | 0.0663 | 0.3616 | 0.0623 | 6.60E-09 | Yes | No |
| rs11771693 | 7 | 150050111 | A | G | 0.6755 | 0.183 | 0.0326 | 1.90E-08 | Yes | No |
| rs3918226 | 7 | 150690176 | T | C | 0.0811 | 0.664 | 0.0575 | 8.46E-31 | Yes | No |
| rs891511 | 7 | 150704843 | A | G | 0.3335 | -0.3506 | 0.0333 | 6.13E-26 | Yes | No |
| rs10224002 | 7 | 151415041 | A | G | 0.7141 | -0.3672 | 0.0337 | 1.31E-27 | Yes | Yes |
| rs1870735 | 7 | 155744303 | C | G | 0.4531 | 0.206 | 0.0311 | 3.61E-11 | No | No |
| rs9638084 | 7 | 156311745 | A | G | 0.3979 | 0.1578 | 0.031 | 3.68E-07 | Yes | No |
| rs4875958 | 8 | 1721090 | A | G | 0.7119 | 0.2256 | 0.0336 | 1.85E-11 | Yes | No |
| rs2922895 | 8 | 6379932 | C | G | 0.43 | 0.1396 | 0.0306 | 4.94E-06 | No | No |
| rs61040371 | 8 | 8503700 | T | C | 0.6287 | 0.1836 | 0.0314 | 4.77E-09 | Yes | No |
| rs62491354 | 8 | 9730663 | A | G | 0.1342 | 0.3087 | 0.0443 | 3.25E-12 | Yes | No |
| rs1986971 | 8 | 10268736 | A | G | 0.7064 | 0.2565 | 0.0334 | 1.61E-14 | Yes | Yes |
| rs2898290 | 8 | 11433909 | T | C | 0.4762 | 0.3123 | 0.0304 | 1.08E-24 | Yes | No |
| rs75902664 | 8 | 17427186 | A | G | 0.9742 | -0.2514 | 0.1002 | 0.01208 | Yes | No |
| rs1047030 | 8 | 22428708 | A | G | 0.8045 | 0.2073 | 0.0408 | 3.71E-07 | Yes | No |
| rs62503324 | 8 | 23400615 | T | C | 0.2393 | 0.2638 | 0.0356 | 1.26E-13 | Yes | No |
| rs6557876 | 8 | 25900675 | T | C | 0.2503 | -0.4156 | 0.0349 | 1.11E-32 | Yes | No |
| rs17321041 | 8 | 26445194 | T | C | 0.0631 | 0.2943 | 0.0633 | 3.36E-06 | Yes | No |
| rs2979470 | 8 | 30288272 | T | C | 0.49 | 0.1991 | 0.0302 | 4.62E-11 | Yes | Yes |
| rs11991469 | 8 | 32413280 | C | G | 0.5392 | -0.1118 | 0.0303 | 0.0002221 | No | No |
| rs7845722 | 8 | 33309993 | A | G | 0.6005 | -0.1269 | 0.0308 | 3.72E-05 | Yes | No |
| rs1906672 | 8 | 38130025 | A | G | 0.2319 | 0.2966 | 0.0358 | 1.20E-16 | Yes | Yes |
| rs2978456 | 8 | 42324765 | T | C | 0.5517 | -0.1798 | 0.0313 | 8.95E-09 | No | No |
| rs4873492 | 8 | 51947549 | T | C | 0.1724 | 0.3431 | 0.0403 | 1.61E-17 | Yes | Yes |
| rs6996733 | 8 | 60535824 | T | C | 0.8488 | 0.2106 | 0.0426 | 7.79E-07 | Yes | No |
| rs2354862 | 8 | 64501744 | A | C | 0.6407 | 0.2507 | 0.0317 | 2.42E-15 | Yes | No |
| rs13253358 | 8 | 68920135 | T | C | 0.2979 | 0.2127 | 0.033 | 1.13E-10 | Yes | No |
| rs1350100 | 8 | 76054904 | A | G | 0.4446 | 0.1028 | 0.0306 | 0.000781 | Yes | No |
| rs1449544 | 8 | 76591880 | A | C | 0.5436 | 0.2117 | 0.0303 | 2.57E-12 | Yes | No |
| rs72688070 | 8 | 81393697 | T | C | 0.1686 | -0.2705 | 0.0406 | 2.82E-11 | Yes | No |
| rs56345595 | 8 | 82814156 | A | G | 0.5853 | 0.2066 | 0.0308 | 1.91E-11 | Yes | No |
| rs2142141 | 8 | 90940205 | C | G | 0.5325 | -0.1476 | 0.0312 | 2.26E-06 | No | No |
| rs7009170 | 8 | 92149429 | T | C | 0.3189 | -0.2151 | 0.0324 | 3.01E-11 | Yes | No |
| rs62526122 | 8 | 92769569 | A | G | 0.2899 | 0.2108 | 0.0345 | 1.02E-09 | Yes | No |
| rs4582532 | 8 | 95969257 | A | G | 0.5059 | -0.2003 | 0.0303 | 3.64E-11 | Yes | No |
| rs2978098 | 8 | 101676675 | A | C | 0.5468 | 0.2227 | 0.0307 | 3.78E-13 | Yes | No |
| rs142449193 | 8 | 102750597 | T | C | 0.0464 | -0.4549 | 0.074 | 7.86E-10 | Yes | No |
| rs2513877 | 8 | 103883630 | A | G | 0.1924 | -0.1586 | 0.0383 | 3.52E-05 | Yes | No |
| rs35783704 | 8 | 105966258 | A | G | 0.1042 | -0.4619 | 0.0507 | 8.81E-20 | Yes | No |
| rs28499085 | 8 | 110107161 | A | G | 0.7253 | 0.2096 | 0.0339 | 6.42E-10 | Yes | No |
| rs2205260 | 8 | 116959837 | A | C | 0.1677 | 0.0666 | 0.0405 | 0.1002 | Yes | No |
| rs2071518 | 8 | 120435812 | T | C | 0.2611 | 0.2757 | 0.0343 | 8.76E-16 | Yes | No |
| rs62523863 | 8 | 126520544 | A | G | 0.2197 | 0.2689 | 0.0368 | 2.87E-13 | Yes | No |
| rs4598218 | 8 | 129483956 | T | C | 0.6158 | 0.1911 | 0.0313 | 1.00E-09 | Yes | Yes |
| rs894344 | 8 | 135612745 | A | G | 0.5926 | -0.2136 | 0.0306 | 3.11E-12 | Yes | No |
| rs4454254 | 8 | 141060027 | A | G | 0.6312 | -0.2177 | 0.0312 | 3.05E-12 | Yes | No |
| rs10087782 | 8 | 141858620 | T | C | 0.4466 | 0.2254 | 0.0303 | 9.86E-14 | Yes | No |
| rs34591516 | 8 | 142367087 | T | C | 0.0494 | 0.4862 | 0.071 | 7.65E-12 | Yes | No |
| rs4129585 | 8 | 143312933 | A | C | 0.4413 | 0.1862 | 0.0305 | 1.03E-09 | Yes | Yes |
| rs62524579 | 8 | 144060955 | A | G | 0.533 | -0.2676 | 0.0321 | 6.87E-17 | Yes | No |
| rs56233017 | 8 | 144981488 | A | G | 0.0458 | -0.1583 | 0.077 | 0.03988 | Yes | No |
| rs520015 | 9 | 211762 | C | G | 0.513 | 0.2003 | 0.0301 | 2.84E-11 | No | No |
| rs60191654 | 9 | 753648 | A | G | 0.8118 | -0.2382 | 0.0385 | 5.88E-10 | Yes | No |
| rs12216886 | 9 | 2493751 | T | G | 0.8073 | 0.1902 | 0.0382 | 6.30E-07 | Yes | No |
| rs28558845 | 9 | 4334791 | C | G | 0.1553 | -0.2569 | 0.0422 | 1.16E-09 | Yes | No |
| rs1332813 | 9 | 9350706 | T | C | 0.3514 | 0.2203 | 0.0314 | 2.32E-12 | Yes | No |
| rs35287509 | 9 | 10594635 | T | C | 0.6627 | -0.1645 | 0.0319 | 2.57E-07 | Yes | No |
| rs11789875 | 9 | 16872323 | A | G | 0.1519 | 0.1609 | 0.042 | 0.0001271 | Yes | No |
| rs4977492 | 9 | 19057551 | T | C | 0.6624 | -0.124 | 0.0317 | 9.07E-05 | Yes | No |
| rs4364717 | 9 | 21801530 | A | G | 0.5476 | -0.0993 | 0.0302 | 0.000995 | Yes | No |
| rs9886665 | 9 | 22942770 | T | C | 0.2671 | 0.2048 | 0.0343 | 2.47E-09 | Yes | Yes |
| rs4553000 | 9 | 34223553 | T | C | 0.5141 | -0.2035 | 0.03 | 1.09E-11 | Yes | No |
| rs76452347 | 9 | 35906471 | T | C | 0.205 | -0.2974 | 0.0397 | 7.13E-14 | No | No |
| rs11139596 | 9 | 85128518 | T | G | 0.7759 | -0.1167 | 0.037 | 0.001616 | Yes | No |
| rs11141731 | 9 | 89888472 | T | C | 0.2272 | -0.1945 | 0.0359 | 6.19E-08 | Yes | No |
| rs7045409 | 9 | 95201540 | A | T | 0.3669 | -0.1862 | 0.0313 | 2.55E-09 | Yes | No |
| rs10988442 | 9 | 101739709 | A | G | 0.6193 | 0.1477 | 0.0311 | 2.04E-06 | Yes | No |
| rs7020564 | 9 | 109670016 | A | T | 0.7007 | -0.0739 | 0.0332 | 0.02594 | Yes | No |
| rs7043304 | 9 | 112358150 | T | C | 0.8545 | 0.1985 | 0.0432 | 4.31E-06 | Yes | No |
| rs111245230 | 9 | 113169775 | T | C | 0.9654 | -0.7486 | 0.0834 | 2.81E-19 | Yes | No |
| rs13290326 | 9 | 116696625 | T | C | 0.5014 | -0.2122 | 0.03 | 1.52E-12 | Yes | No |
| rs10982910 | 9 | 118534500 | T | G | 0.9039 | -0.1196 | 0.0517 | 0.02066 | Yes | No |
| rs1861881 | 9 | 119312256 | T | G | 0.316 | 0.1276 | 0.0323 | 7.63E-05 | Yes | No |
| rs1953126 | 9 | 123640500 | T | C | 0.3536 | 0.1581 | 0.0315 | 5.20E-07 | Yes | No |
| rs10818775 | 9 | 125755571 | T | C | 0.1213 | -0.302 | 0.0462 | 6.23E-11 | Yes | No |
| rs7861040 | 9 | 127044135 | C | G | 0.367 | -0.1888 | 0.031 | 1.21E-09 | Yes | No |
| rs72765298 | 9 | 127900996 | T | C | 0.8751 | -0.3675 | 0.0462 | 1.75E-15 | Yes | No |
| rs7023828 | 9 | 128498594 | T | C | 0.418 | -0.2595 | 0.0306 | 2.17E-17 | Yes | No |
| rs1891730 | 9 | 130309028 | T | C | 0.6166 | -0.1812 | 0.0314 | 7.74E-09 | Yes | Yes |
| rs7869756 | 9 | 131210410 | A | G | 0.8099 | -0.0724 | 0.039 | 0.06328 | Yes | No |
| rs184457 | 9 | 131940019 | A | G | 0.308 | -0.1716 | 0.0326 | 1.43E-07 | Yes | No |
| rs687621 | 9 | 136137065 | A | G | 0.6717 | 0.1026 | 0.0321 | 0.001399 | Yes | No |
| rs6271 | 9 | 136522274 | T | C | 0.0735 | -0.5547 | 0.0611 | 1.18E-19 | No | No |
| rs11145807 | 9 | 139520789 | A | G | 0.4057 | 0.2135 | 0.0322 | 3.54E-11 | No | No |
| rs56352451 | 10 | 5804865 | T | C | 0.1324 | 0.2224 | 0.0444 | 5.51E-07 | Yes | No |
| rs36006409 | 10 | 10876943 | T | G | 0.7937 | -0.159 | 0.0376 | 2.35E-05 | Yes | No |
| rs10906391 | 10 | 13523937 | T | C | 0.3191 | 0.1254 | 0.0327 | 0.0001261 | Yes | No |
| rs4373814 | 10 | 18419972 | C | G | 0.4277 | 0.3266 | 0.0306 | 1.20E-26 | No | No |
| rs1813353 | 10 | 18707448 | T | C | 0.6625 | 0.5198 | 0.0319 | 8.15E-60 | Yes | No |
| rs11010905 | 10 | 19934813 | A | T | 0.4854 | 0.0774 | 0.0302 | 0.01035 | No | No |
| rs72795925 | 10 | 20531420 | T | C | 0.2208 | 0.1246 | 0.0364 | 0.0006166 | Yes | No |
| rs10732433 | 10 | 21037294 | T | C | 0.4206 | 0.1553 | 0.0306 | 3.79E-07 | Yes | No |
| rs3802517 | 10 | 28233469 | A | T | 0.4618 | 0.2527 | 0.0301 | 4.65E-17 | No | No |
| rs1265842 | 10 | 28924901 | T | C | 0.4836 | 0.128 | 0.0304 | 2.53E-05 | Yes | No |
| rs9337951 | 10 | 30317073 | A | G | 0.3414 | 0.1505 | 0.0334 | 6.46E-06 | No | No |
| rs11008355 | 10 | 31412561 | C | G | 0.2388 | -0.0559 | 0.0354 | 0.1143 | Yes | No |
| rs10826995 | 10 | 32082658 | T | C | 0.7161 | -0.1936 | 0.0335 | 7.55E-09 | Yes | No |
| rs76164690 | 10 | 32590362 | T | G | 0.8597 | -0.1967 | 0.0438 | 6.94E-06 | Yes | No |
| rs2246438 | 10 | 45273079 | A | G | 0.2762 | -0.0453 | 0.0338 | 0.18 | Yes | No |
| rs34130368 | 10 | 48411796 | T | G | 0.117 | -0.3016 | 0.0497 | 1.28E-09 | No | No |
| rs10761530 | 10 | 62390726 | T | C | 0.4989 | 0.2004 | 0.03 | 2.54E-11 | Yes | No |
| rs1530440 | 10 | 63524591 | T | C | 0.1875 | -0.5669 | 0.0386 | 7.36E-49 | Yes | No |
| rs7090758 | 10 | 65335315 | T | C | 0.5284 | -0.1607 | 0.0301 | 9.66E-08 | Yes | No |
| rs7914287 | 10 | 69350563 | T | C | 0.7794 | -0.1889 | 0.0368 | 2.86E-07 | Yes | No |
| rs10823136 | 10 | 69855363 | T | C | 0.9215 | -0.3501 | 0.0583 | 1.91E-09 | Yes | No |
| rs10998362 | 10 | 70404159 | T | C | 0.3172 | 0.1072 | 0.0336 | 0.00141 | Yes | No |
| rs12572586 | 10 | 74751579 | T | C | 0.9377 | -0.3899 | 0.0642 | 1.23E-09 | Yes | No |
| rs10887914 | 10 | 82215288 | T | C | 0.4625 | 0.2007 | 0.0302 | 3.18E-11 | Yes | No |
| rs77413490 | 10 | 89681688 | T | G | 0.0424 | 0.4489 | 0.0764 | 4.27E-09 | Yes | No |
| rs11187142 | 10 | 94468685 | T | C | 0.1047 | 0.3312 | 0.0496 | 2.53E-11 | Yes | No |
| rs932764 | 10 | 95895940 | A | G | 0.5689 | -0.4303 | 0.0306 | 5.43E-45 | Yes | Yes |
| rs4494250 | 10 | 96563757 | A | G | 0.3625 | 0.3038 | 0.0316 | 6.05E-22 | Yes | No |
| rs603424 | 10 | 102075479 | A | G | 0.1784 | 0.233 | 0.0402 | 6.67E-09 | Yes | No |
| rs112184198 | 10 | 102604514 | A | G | 0.1047 | -0.6626 | 0.0498 | 1.94E-40 | Yes | No |
| rs72847884 | 10 | 103115345 | A | G | 0.9534 | 0.1035 | 0.0741 | 0.1626 | Yes | No |
| rs11191156 | 10 | 103702763 | A | G | 0.6484 | -0.1553 | 0.0317 | 9.57E-07 | Yes | No |
| rs11191548 | 10 | 104846178 | T | C | 0.9188 | 1.0983 | 0.0553 | 1.16E-87 | Yes | Yes |
| rs4387287 | 10 | 105677897 | A | C | 0.165 | 0.2282 | 0.0408 | 2.26E-08 | Yes | No |
| rs191784289 | 10 | 106894942 | T | C | 0.0121 | 1.2287 | 0.1432 | 9.24E-18 | Yes | No |
| rs111777102 | 10 | 111965826 | T | C | 0.0661 | 0.2908 | 0.0617 | 2.39E-06 | Yes | No |
| rs34872471 | 10 | 114754071 | T | C | 0.7076 | -0.2201 | 0.0333 | 3.83E-11 | Yes | No |
| rs2782980 | 10 | 115781527 | T | C | 0.2841 | -0.4249 | 0.0338 | 3.58E-36 | Yes | No |
| rs11197813 | 10 | 118523933 | A | G | 0.6992 | -0.182 | 0.033 | 3.53E-08 | Yes | Yes |
| rs72842207 | 10 | 121433675 | T | C | 0.2144 | -0.203 | 0.0367 | 3.14E-08 | Yes | No |
| rs11592107 | 10 | 122968964 | A | G | 0.3096 | 0.3024 | 0.0326 | 1.55E-20 | Yes | Yes |
| rs72834453 | 10 | 124235226 | T | G | 0.8756 | -0.3246 | 0.0465 | 2.95E-12 | Yes | No |
| rs4411245 | 10 | 126712781 | A | G | 0.2934 | 0.1254 | 0.0331 | 0.0001512 | Yes | No |
| rs7096563 | 10 | 133770229 | A | G | 0.3548 | 0.21 | 0.0321 | 6.39E-11 | Yes | No |
| rs1133400 | 10 | 134459388 | A | G | 0.786 | -0.2975 | 0.0376 | 2.53E-15 | Yes | Yes |
| rs7126805 | 11 | 828916 | A | G | 0.7277 | 0.1599 | 0.0349 | 4.46E-06 | Yes | No |
| rs661348 | 11 | 1905292 | T | C | 0.5747 | -0.4451 | 0.0316 | 5.23E-45 | Yes | Yes |
| rs17224476 | 11 | 4673788 | A | G | 0.1113 | 0.2078 | 0.0482 | 1.61E-05 | Yes | No |
| rs2929184 | 11 | 6289118 | A | G | 0.771 | 0.1294 | 0.0372 | 0.0005068 | Yes | No |
| rs110419 | 11 | 8252853 | A | G | 0.4785 | 0.1133 | 0.0299 | 0.0001519 | Yes | No |
| rs10743086 | 11 | 8774923 | A | G | 0.2072 | -0.2053 | 0.0373 | 3.60E-08 | Yes | No |
| rs360153 | 11 | 9762274 | T | C | 0.4166 | -0.3445 | 0.0306 | 1.73E-29 | Yes | No |
| rs7129220 | 11 | 10350538 | A | G | 0.1192 | 0.5002 | 0.0472 | 2.96E-26 | Yes | Yes |
| rs900145 | 11 | 13293905 | T | C | 0.7043 | 0.1786 | 0.0329 | 5.63E-08 | Yes | No |
| rs4757391 | 11 | 16302939 | T | C | 0.798 | -0.5112 | 0.0375 | 2.07E-42 | Yes | No |
| rs381815 | 11 | 16902268 | T | C | 0.2746 | 0.3009 | 0.0338 | 5.62E-19 | Yes | No |
| rs5219 | 11 | 17409572 | T | C | 0.3657 | 0.3474 | 0.0312 | 9.18E-29 | Yes | No |
| rs10766533 | 11 | 19224677 | A | T | 0.7108 | 0.2099 | 0.0337 | 4.69E-10 | Yes | Yes |
| rs11026586 | 11 | 22515533 | A | G | 0.0705 | 0.4341 | 0.0597 | 3.66E-13 | Yes | No |
| rs11030119 | 11 | 27728102 | A | G | 0.3013 | -0.2218 | 0.033 | 1.79E-11 | Yes | No |
| rs871004 | 11 | 28512458 | A | G | 0.3481 | 0.2336 | 0.0317 | 1.65E-13 | Yes | Yes |
| rs11031051 | 11 | 30355707 | A | C | 0.6901 | -0.2238 | 0.0327 | 7.73E-12 | Yes | No |
| rs919045 | 11 | 31111810 | T | C | 0.6281 | 0.1704 | 0.0312 | 4.64E-08 | Yes | No |
| rs4922591 | 11 | 32374199 | T | C | 0.3869 | -0.1892 | 0.0314 | 1.76E-09 | Yes | No |
| rs190194639 | 11 | 34068037 | T | C | 0.0766 | 0.3023 | 0.0577 | 1.61E-07 | Yes | No |
| rs7480089 | 11 | 45207851 | A | G | 0.118 | -0.2614 | 0.0471 | 2.94E-08 | Yes | No |
| rs1585453 | 11 | 46884713 | A | T | 0.8868 | -0.3825 | 0.049 | 6.15E-15 | Yes | No |
| rs7103648 | 11 | 47461783 | A | G | 0.6129 | -0.3862 | 0.031 | 1.27E-35 | Yes | No |
| rs11537751 | 11 | 47587452 | T | C | 0.0548 | 0.4404 | 0.0674 | 6.30E-11 | Yes | No |
| rs2688716 | 11 | 54835623 | T | G | 0.2894 | -0.2479 | 0.035 | 1.37E-12 | No | No |
| rs75905900 | 11 | 55113534 | A | C | 0.8656 | 0.4174 | 0.0449 | 1.36E-20 | Yes | No |
| rs11607056 | 11 | 57496820 | T | C | 0.3276 | -0.2511 | 0.0321 | 4.79E-15 | Yes | No |
| rs11229457 | 11 | 58207203 | T | C | 0.213 | -0.317 | 0.0369 | 8.45E-18 | Yes | Yes |
| rs751984 | 11 | 61278246 | T | C | 0.8824 | 0.4408 | 0.0479 | 3.60E-20 | Yes | No |
| rs4980515 | 11 | 63744609 | T | C | 0.4993 | 0.2253 | 0.0303 | 9.73E-14 | Yes | No |
| rs3741378 | 11 | 65408937 | T | C | 0.1352 | -0.4087 | 0.0446 | 4.83E-20 | Yes | Yes |
| rs67976715 | 11 | 68023742 | C | G | 0.2342 | 0.2082 | 0.0359 | 6.80E-09 | Yes | No |
| rs67330701 | 11 | 69079707 | T | C | 0.0936 | -0.3913 | 0.0563 | 3.63E-12 | Yes | No |
| rs875106 | 11 | 70005641 | A | G | 0.5222 | -0.0824 | 0.0302 | 0.006299 | Yes | No |
| rs504217 | 11 | 72006086 | T | C | 0.0738 | 0.3855 | 0.0583 | 3.90E-11 | Yes | No |
| rs2298807 | 11 | 73068571 | T | C | 0.784 | 0.1501 | 0.0368 | 4.49E-05 | Yes | No |
| rs4420291 | 11 | 74374950 | A | G | 0.5057 | 0.0745 | 0.0303 | 0.01385 | Yes | No |
| rs7927515 | 11 | 76125330 | A | C | 0.3459 | 0.2271 | 0.0319 | 1.05E-12 | Yes | Yes |
| rs59986178 | 11 | 77359909 | C | G | 0.1043 | 0.1284 | 0.0516 | 0.01278 | Yes | No |
| rs2450128 | 11 | 77940075 | A | G | 0.1544 | -0.1728 | 0.0419 | 3.65E-05 | Yes | No |
| rs2289125 | 11 | 89224453 | A | C | 0.2205 | -0.2592 | 0.0375 | 4.90E-12 | Yes | No |
| rs10830963 | 11 | 92708710 | C | G | 0.7235 | -0.1324 | 0.0347 | 0.0001389 | Yes | No |
| rs11021221 | 11 | 95308854 | A | T | 0.1663 | 0.0019 | 0.0406 | 0.9631 | Yes | No |
| rs633185 | 11 | 100593538 | C | G | 0.7127 | 0.6373 | 0.0334 | 5.60E-81 | Yes | No |
| rs61892344 | 11 | 101100768 | T | C | 0.1738 | -0.0948 | 0.0399 | 0.01738 | Yes | No |
| rs12807220 | 11 | 102077200 | A | G | 0.3593 | -0.0718 | 0.0317 | 0.02337 | Yes | No |
| rs4754196 | 11 | 107096777 | A | G | 0.5216 | -0.3486 | 0.0303 | 1.46E-30 | Yes | No |
| rs12362593 | 11 | 111586091 | C | G | 0.729 | -0.2004 | 0.0347 | 7.91E-09 | Yes | No |
| rs17119370 | 11 | 116097136 | A | T | 0.6926 | 0.2207 | 0.033 | 2.16E-11 | Yes | Yes |
| rs1076485 | 11 | 116772441 | T | C | 0.1304 | 0.3388 | 0.0457 | 1.19E-13 | Yes | No |
| rs8258 | 11 | 117283676 | T | C | 0.3739 | 0.1648 | 0.0312 | 1.29E-07 | Yes | No |
| rs12574332 | 11 | 122521123 | T | C | 0.1231 | 0.2452 | 0.0463 | 1.20E-07 | Yes | No |
| rs11222386 | 11 | 130779068 | C | G | 0.1973 | 0.153 | 0.038 | 5.57E-05 | Yes | No |
| rs78998485 | 12 | 434755 | C | G | 0.7443 | -0.2449 | 0.0346 | 1.48E-12 | Yes | No |
| rs11571376 | 12 | 1059556 | C | G | 0.7057 | -0.1803 | 0.0332 | 5.70E-08 | Yes | No |
| rs55935819 | 12 | 2521579 | A | G | 0.3634 | 0.1745 | 0.0313 | 2.43E-08 | Yes | No |
| rs117233107 | 12 | 4328521 | A | G | 0.0158 | -0.6366 | 0.1368 | 3.27E-06 | Yes | No |
| rs75507123 | 12 | 5417856 | T | G | 0.1276 | -0.2319 | 0.0452 | 2.86E-07 | Yes | No |
| rs7132012 | 12 | 8832203 | A | G | 0.6748 | 0.2 | 0.0321 | 4.76E-10 | Yes | No |
| rs2024385 | 12 | 12888438 | A | T | 0.424 | -0.2642 | 0.0306 | 5.88E-18 | No | No |
| rs28621435 | 12 | 13860990 | A | G | 0.1151 | -0.2976 | 0.0482 | 6.47E-10 | Yes | No |
| rs7313556 | 12 | 15297359 | A | G | 0.3484 | -0.048 | 0.0313 | 0.1254 | Yes | No |
| rs61912333 | 12 | 19554817 | C | G | 0.4968 | 0.1837 | 0.0304 | 1.57E-09 | No | No |
| rs12579720 | 12 | 20173764 | C | G | 0.2434 | -0.258 | 0.0352 | 2.46E-13 | Yes | No |
| rs73080726 | 12 | 20754154 | T | C | 0.102 | -0.2444 | 0.0499 | 9.69E-07 | Yes | No |
| rs704191 | 12 | 22015022 | T | C | 0.4629 | 0.1526 | 0.0302 | 4.46E-07 | Yes | No |
| rs7976167 | 12 | 24210599 | T | C | 0.6889 | 0.1784 | 0.0324 | 3.81E-08 | Yes | Yes |
| rs17287293 | 12 | 24770878 | A | G | 0.8508 | -0.0703 | 0.042 | 0.09424 | Yes | No |
| rs6487543 | 12 | 26438189 | A | G | 0.7699 | 0.1904 | 0.0368 | 2.29E-07 | Yes | No |
| rs1098708 | 12 | 27321112 | A | G | 0.5423 | -0.1969 | 0.0303 | 8.21E-11 | Yes | No |
| rs10842991 | 12 | 27962103 | T | C | 0.1975 | -0.0908 | 0.0381 | 0.01731 | Yes | No |
| rs7965392 | 12 | 42540280 | A | G | 0.3866 | 0.0935 | 0.031 | 0.00257 | Yes | No |
| rs11168245 | 12 | 48204499 | C | G | 0.7655 | 0.2784 | 0.0357 | 5.83E-15 | No | No |
| rs2261608 | 12 | 48721634 | A | T | 0.3511 | 0.1723 | 0.0313 | 3.65E-08 | Yes | No |
| rs1126930 | 12 | 49399132 | C | G | 0.035 | 0.6378 | 0.0856 | 9.48E-14 | Yes | No |
| rs7977389 | 12 | 49981722 | T | C | 0.8938 | 0.2575 | 0.049 | 1.46E-07 | Yes | No |
| rs7302981 | 12 | 50537815 | A | G | 0.3788 | 0.3755 | 0.0308 | 2.77E-34 | Yes | No |
| rs61926181 | 12 | 50767037 | A | G | 0.0384 | -0.6472 | 0.0846 | 2.02E-14 | Yes | No |
| rs73099903 | 12 | 53440779 | T | C | 0.0824 | 0.4694 | 0.0557 | 3.43E-17 | Yes | No |
| rs7297416 | 12 | 54443090 | A | C | 0.6955 | 0.3764 | 0.0329 | 2.90E-30 | Yes | Yes |
| rs7137749 | 12 | 57098040 | T | C | 0.3675 | 0.0775 | 0.0314 | 0.0136 | Yes | No |
| rs10437954 | 12 | 58003922 | A | G | 0.9069 | -0.4101 | 0.0534 | 1.60E-14 | Yes | No |
| rs4143175 | 12 | 67782397 | T | C | 0.2409 | 0.2187 | 0.0352 | 5.10E-10 | Yes | Yes |
| rs513177 | 12 | 69950545 | T | C | 0.1405 | 0.2386 | 0.0431 | 3.06E-08 | No | No |
| rs7963801 | 12 | 79685226 | T | C | 0.4221 | -0.2362 | 0.0311 | 2.87E-14 | No | No |
| rs17249754 | 12 | 90060586 | A | G | 0.1683 | -0.8446 | 0.0403 | 1.25E-97 | Yes | Yes |
| rs10858966 | 12 | 90567026 | C | G | 0.2989 | 0.2565 | 0.0331 | 9.27E-15 | Yes | No |
| rs76785029 | 12 | 94882905 | T | C | 0.0787 | -0.1901 | 0.0582 | 0.001084 | Yes | No |
| rs7977311 | 12 | 95487226 | T | C | 0.1154 | -0.2296 | 0.0469 | 1.00E-06 | Yes | No |
| rs11108209 | 12 | 96109855 | T | C | 0.9069 | -0.1849 | 0.0521 | 0.0003823 | Yes | No |
| rs7134060 | 12 | 96717095 | A | G | 0.4465 | -0.1109 | 0.0301 | 0.0002284 | Yes | No |
| rs10778174 | 12 | 102838996 | A | G | 0.2479 | -0.2209 | 0.0348 | 2.08E-10 | Yes | No |
| rs11112548 | 12 | 105871914 | A | T | 0.9556 | 0.5092 | 0.0768 | 3.34E-11 | Yes | Yes |
| rs12184466 | 12 | 111281636 | T | C | 0.1991 | 0.2992 | 0.0434 | 5.30E-12 | No | No |
| rs3184504 | 12 | 111884608 | T | C | 0.48 | 0.5779 | 0.0303 | 6.54E-81 | Yes | No |
| rs10850411 | 12 | 115387796 | T | C | 0.6969 | 0.2908 | 0.0326 | 4.58E-19 | Yes | No |
| rs35444 | 12 | 115552437 | A | G | 0.6138 | 0.4368 | 0.031 | 3.47E-45 | Yes | No |
| rs11067763 | 12 | 116198341 | A | G | 0.8983 | 0.3069 | 0.05 | 8.10E-10 | Yes | No |
| rs11615689 | 12 | 116699675 | T | C | 0.8196 | -0.1511 | 0.039 | 0.0001057 | Yes | No |
| rs3898618 | 12 | 120813921 | T | C | 0.9445 | -0.3089 | 0.0664 | 3.33E-06 | Yes | No |
| rs28498002 | 12 | 122599796 | T | C | 0.4751 | -0.1604 | 0.031 | 2.19E-07 | Yes | No |
| rs1060105 | 12 | 123806219 | T | C | 0.203 | -0.1025 | 0.0375 | 0.006263 | Yes | No |
| rs1271309 | 12 | 124820705 | A | G | 0.162 | -0.1607 | 0.0416 | 0.0001106 | Yes | No |
| rs117206641 | 12 | 133086888 | T | C | 0.1108 | 0.3154 | 0.0499 | 2.66E-10 | Yes | No |
| rs2480171 | 13 | 21559858 | T | C | 0.1221 | 0.2909 | 0.0467 | 4.69E-10 | Yes | No |
| rs606950 | 13 | 22298923 | A | G | 0.6163 | 0.2704 | 0.0311 | 3.23E-18 | Yes | No |
| rs55641580 | 13 | 25257917 | T | C | 0.1262 | 0.2685 | 0.046 | 5.29E-09 | Yes | No |
| rs1331012 | 13 | 27115424 | T | G | 0.2703 | 0.2043 | 0.0338 | 1.49E-09 | Yes | No |
| rs63418562 | 13 | 30146201 | T | C | 0.7565 | -0.3557 | 0.0353 | 6.34E-24 | No | No |
| rs9532243 | 13 | 32191408 | A | C | 0.4839 | 0.2241 | 0.03 | 8.17E-14 | Yes | No |
| rs9549297 | 13 | 41397482 | A | G | 0.8207 | -0.1101 | 0.0397 | 0.005558 | Yes | No |
| rs4274337 | 13 | 41967193 | A | G | 0.1697 | -0.2968 | 0.0406 | 2.48E-13 | Yes | No |
| rs73187288 | 13 | 42738672 | A | C | 0.8909 | -0.2766 | 0.0483 | 1.04E-08 | Yes | No |
| rs912434 | 13 | 47189928 | T | G | 0.7594 | 0.234 | 0.0351 | 2.47E-11 | Yes | Yes |
| rs12583615 | 13 | 50564085 | A | G | 0.1321 | 0.1457 | 0.0446 | 0.00109 | Yes | No |
| rs9526707 | 13 | 51489186 | A | G | 0.3216 | -0.2039 | 0.0323 | 2.77E-10 | Yes | No |
| rs75961402 | 13 | 56398286 | A | G | 0.1534 | 0.2659 | 0.0418 | 1.95E-10 | Yes | No |
| rs9563529 | 13 | 58316637 | T | G | 0.204 | 0.1859 | 0.0373 | 6.30E-07 | Yes | No |
| rs3861113 | 13 | 72364382 | A | C | 0.0838 | 0.2231 | 0.0555 | 5.92E-05 | Yes | No |
| rs78474310 | 13 | 73826901 | A | G | 0.9552 | -0.4699 | 0.0734 | 1.51E-10 | Yes | No |
| rs4304924 | 13 | 79238925 | A | G | 0.5671 | -0.0927 | 0.0306 | 0.002421 | Yes | No |
| rs7988232 | 13 | 79808655 | A | G | 0.4113 | 0.1525 | 0.0307 | 6.91E-07 | Yes | No |
| rs1215469 | 13 | 80707408 | A | C | 0.2297 | -0.2124 | 0.0365 | 6.16E-09 | Yes | No |
| rs55684003 | 13 | 97988689 | A | G | 0.6964 | 0.1172 | 0.0327 | 0.0003369 | Yes | No |
| rs3742182 | 13 | 111375132 | T | C | 0.8098 | -0.2058 | 0.0382 | 7.30E-08 | Yes | No |
| rs9549328 | 13 | 113636156 | T | C | 0.2305 | 0.2915 | 0.0362 | 7.91E-16 | Yes | No |
| rs7331680 | 13 | 115000650 | T | G | 0.1491 | 0.4101 | 0.0423 | 3.35E-22 | Yes | No |
| rs17880989 | 14 | 23313633 | A | G | 0.0259 | 0.5248 | 0.1027 | 3.27E-07 | No | No |
| rs452036 | 14 | 23865885 | A | G | 0.3546 | -0.2218 | 0.0315 | 2.02E-12 | Yes | No |
| rs17115145 | 14 | 30122409 | T | C | 0.3967 | 0.1775 | 0.0307 | 7.39E-09 | Yes | Yes |
| rs4424827 | 14 | 35110857 | T | C | 0.567 | -0.1269 | 0.0303 | 2.79E-05 | Yes | No |
| rs8904 | 14 | 35871217 | A | G | 0.3678 | 0.3061 | 0.0314 | 1.71E-22 | Yes | No |
| rs34983854 | 14 | 39858442 | A | G | 0.6028 | -0.2056 | 0.0307 | 2.06E-11 | Yes | No |
| rs72683923 | 14 | 50735947 | T | C | 0.9788 | 0.9587 | 0.1101 | 3.08E-18 | No | No |
| rs9888615 | 14 | 53377540 | T | C | 0.291 | -0.274 | 0.0332 | 1.46E-16 | Yes | Yes |
| rs210381 | 14 | 54107791 | A | G | 0.5651 | -0.1446 | 0.0303 | 1.88E-06 | Yes | No |
| rs7144602 | 14 | 55285588 | T | G | 0.6488 | -0.1575 | 0.0317 | 6.52E-07 | Yes | No |
| rs11628933 | 14 | 60700903 | C | G | 0.2335 | -0.1025 | 0.0357 | 0.00409 | Yes | No |
| rs731681 | 14 | 68010224 | C | G | 0.5585 | -0.154 | 0.0302 | 3.40E-07 | No | No |
| rs57786342 | 14 | 69260028 | A | G | 0.2059 | 0.2317 | 0.0374 | 5.63E-10 | Yes | No |
| rs11623535 | 14 | 72462381 | A | G | 0.7439 | 0.2095 | 0.0343 | 1.02E-09 | Yes | Yes |
| rs4903064 | 14 | 73279420 | T | C | 0.7643 | -0.0277 | 0.0357 | 0.4381 | Yes | No |
| rs11159091 | 14 | 75074316 | A | G | 0.4615 | 0.1978 | 0.0303 | 6.79E-11 | Yes | Yes |
| rs11627326 | 14 | 85785251 | C | G | 0.2869 | 0.0937 | 0.0333 | 0.004916 | Yes | No |
| rs4904503 | 14 | 89565130 | T | C | 0.2976 | 0.0875 | 0.033 | 0.00802 | Yes | No |
| rs11160085 | 14 | 93112102 | T | C | 0.7039 | 0.1565 | 0.0335 | 3.00E-06 | Yes | No |
| rs8013933 | 14 | 94465789 | T | C | 0.6913 | 0.2027 | 0.0327 | 5.48E-10 | Yes | No |
| rs9323988 | 14 | 98587630 | T | C | 0.6152 | -0.2491 | 0.0309 | 6.83E-16 | Yes | No |
| rs1475130 | 14 | 100225144 | T | C | 0.346 | -0.1977 | 0.0318 | 4.83E-10 | Yes | No |
| rs28470843 | 14 | 100742658 | T | C | 0.5971 | 0.1185 | 0.0309 | 0.0001262 | Yes | No |
| rs11626434 | 14 | 101998443 | C | G | 0.3615 | -0.1094 | 0.0321 | 0.0006515 | Yes | No |
| rs8014182 | 14 | 103859962 | T | C | 0.1324 | -0.3339 | 0.0444 | 5.23E-14 | Yes | No |
| rs34161718 | 14 | 104620193 | T | C | 0.242 | -0.1482 | 0.0376 | 8.04E-05 | No | No |
| rs10873612 | 15 | 26105602 | T | C | 0.5957 | -0.1053 | 0.031 | 0.0006929 | Yes | No |
| rs11629850 | 15 | 40317075 | A | G | 0.5287 | 0.2297 | 0.0301 | 2.26E-14 | Yes | No |
| rs2925345 | 15 | 41311799 | T | C | 0.4678 | 0.274 | 0.0301 | 9.14E-20 | Yes | No |
| rs4924570 | 15 | 41974660 | T | C | 0.6287 | -0.211 | 0.0314 | 1.80E-11 | Yes | No |
| rs1036477 | 15 | 48914926 | A | G | 0.8971 | 0.3843 | 0.0494 | 6.88E-15 | Yes | No |
| rs3098186 | 15 | 50810621 | T | C | 0.5156 | -0.2422 | 0.0303 | 1.41E-15 | Yes | No |
| rs3191402 | 15 | 59429160 | A | G | 0.4068 | -0.1217 | 0.0311 | 8.94E-05 | No | No |
| rs956006 | 15 | 62808539 | T | C | 0.3309 | -0.0963 | 0.0323 | 0.002828 | Yes | No |
| rs832890 | 15 | 65166309 | T | C | 0.465 | 0.0943 | 0.0302 | 0.001806 | Yes | No |
| rs7178615 | 15 | 66869072 | A | G | 0.3768 | -0.1647 | 0.0312 | 1.32E-07 | Yes | No |
| rs2289261 | 15 | 67457485 | C | G | 0.6523 | -0.11 | 0.0316 | 0.0005069 | Yes | No |
| rs62004794 | 15 | 68454523 | A | G | 0.4393 | -0.0971 | 0.0302 | 0.001301 | Yes | No |
| rs11853359 | 15 | 71621524 | A | G | 0.3321 | -0.0241 | 0.0317 | 0.4466 | Yes | No |
| rs61653296 | 15 | 74557817 | A | G | 0.8008 | -0.1836 | 0.0378 | 1.17E-06 | Yes | No |
| rs1378942 | 15 | 75077367 | A | C | 0.6676 | -0.5076 | 0.032 | 9.37E-57 | Yes | No |
| rs11634028 | 15 | 76276150 | A | T | 0.2143 | 0.1907 | 0.0376 | 3.84E-07 | Yes | No |
| rs62011052 | 15 | 79156983 | T | C | 0.8493 | -0.1105 | 0.0422 | 0.00888 | Yes | No |
| rs2759308 | 15 | 81016227 | A | G | 0.4749 | 0.3155 | 0.0303 | 1.89E-25 | Yes | Yes |
| rs2034618 | 15 | 83799632 | T | C | 0.2225 | 0.0099 | 0.0363 | 0.7857 | Yes | No |
| rs899927 | 15 | 84561879 | T | C | 0.3177 | 0.0836 | 0.0326 | 0.01027 | Yes | No |
| rs7180952 | 15 | 85162551 | T | C | 0.5385 | -0.0719 | 0.0305 | 0.01827 | Yes | No |
| rs3743157 | 15 | 85680532 | A | C | 0.1669 | 0.293 | 0.0404 | 4.20E-13 | Yes | Yes |
| rs11632436 | 15 | 86295286 | C | G | 0.5035 | 0.222 | 0.0302 | 1.96E-13 | No | No |
| rs28611491 | 15 | 90641809 | T | C | 0.0814 | 0.3343 | 0.0576 | 6.54E-09 | Yes | No |
| rs2521501 | 15 | 91437388 | A | T | 0.6745 | -0.6314 | 0.0331 | 6.40E-81 | No | No |
| rs873122 | 15 | 92702020 | C | G | 0.7197 | 0.1373 | 0.0339 | 5.04E-05 | Yes | No |
| rs11632112 | 15 | 93468276 | C | G | 0.2405 | -0.1771 | 0.0353 | 5.39E-07 | Yes | No |
| rs12906962 | 15 | 95312071 | T | C | 0.676 | -0.2653 | 0.0325 | 3.28E-16 | Yes | No |
| rs4984496 | 15 | 96635898 | T | G | 0.3359 | 0.0744 | 0.0325 | 0.02192 | Yes | No |
| rs34756251 | 15 | 100192540 | T | C | 0.171 | -0.2616 | 0.0399 | 5.44E-11 | Yes | No |
| rs9932866 | 16 | 706067 | A | G | 0.3668 | 0.1596 | 0.0318 | 5.14E-07 | Yes | No |
| rs11248862 | 16 | 1344291 | A | G | 0.1252 | 0.2796 | 0.0463 | 1.53E-09 | Yes | No |
| rs28590346 | 16 | 2080653 | A | T | 0.6597 | -0.2709 | 0.0332 | 3.37E-16 | Yes | No |
| rs2379829 | 16 | 3538873 | C | G | 0.7304 | -0.2678 | 0.0342 | 4.48E-15 | Yes | Yes |
| rs4785955 | 16 | 4297651 | T | G | 0.2181 | 0.1382 | 0.0375 | 0.0002264 | Yes | No |
| rs12921187 | 16 | 4943019 | T | G | 0.4283 | -0.2925 | 0.0303 | 5.19E-22 | Yes | No |
| rs35450617 | 16 | 6889675 | T | G | 0.6993 | -0.1773 | 0.0333 | 1.05E-07 | Yes | No |
| rs11642631 | 16 | 11198835 | T | C | 0.5621 | -0.0936 | 0.0303 | 0.002003 | Yes | No |
| rs57327054 | 16 | 14487036 | T | C | 0.3082 | -0.0757 | 0.0331 | 0.02208 | Yes | No |
| rs3915425 | 16 | 15912544 | T | C | 0.6819 | 0.206 | 0.0323 | 1.86E-10 | Yes | No |
| rs4782211 | 16 | 19152219 | A | G | 0.2712 | 0.1775 | 0.0363 | 9.92E-07 | No | No |
| rs13333226 | 16 | 20365654 | A | G | 0.8156 | 0.377 | 0.0386 | 1.70E-22 | Yes | No |
| rs11639856 | 16 | 24788645 | A | T | 0.1934 | -0.3197 | 0.0379 | 3.53E-17 | Yes | No |
| rs6565174 | 16 | 30111904 | A | C | 0.1121 | -0.2882 | 0.0484 | 2.57E-09 | Yes | No |
| rs72799341 | 16 | 30936743 | A | G | 0.2399 | 0.057 | 0.0354 | 0.1072 | Yes | No |
| rs10468291 | 16 | 49768046 | A | C | 0.5688 | -0.1868 | 0.0305 | 9.38E-10 | Yes | No |
| rs34941092 | 16 | 50550137 | A | G | 0.1498 | -0.3225 | 0.0425 | 3.23E-14 | Yes | No |
| rs9932220 | 16 | 51758116 | A | G | 0.2177 | -0.229 | 0.0364 | 3.18E-10 | Yes | No |
| rs37060 | 16 | 58566304 | A | G | 0.2467 | 0.1103 | 0.0347 | 0.0015 | Yes | No |
| rs28633979 | 16 | 65282820 | A | C | 0.4303 | -0.1794 | 0.0302 | 2.97E-09 | Yes | No |
| rs45474499 | 16 | 66914492 | T | C | 0.0474 | 0.4432 | 0.0717 | 6.41E-10 | Yes | No |
| rs7185555 | 16 | 69131281 | C | G | 0.1526 | -0.0131 | 0.0422 | 0.7557 | Yes | No |
| rs33063 | 16 | 69640217 | A | G | 0.1482 | 0.2244 | 0.0424 | 1.20E-07 | Yes | No |
| rs62053102 | 16 | 71654365 | A | T | 0.953 | -0.5691 | 0.0751 | 3.41E-14 | Yes | No |
| rs1012089 | 16 | 74171973 | C | G | 0.4752 | -0.192 | 0.0302 | 1.95E-10 | No | No |
| rs35261357 | 16 | 75444572 | T | C | 0.585 | 0.3502 | 0.0307 | 4.43E-30 | Yes | No |
| rs56844452 | 16 | 80864776 | T | C | 0.0708 | -0.4506 | 0.0596 | 3.97E-14 | Yes | No |
| rs8059962 | 16 | 81574197 | T | C | 0.4214 | -0.197 | 0.0307 | 1.46E-10 | Yes | No |
| rs7500448 | 16 | 83045790 | A | G | 0.7467 | 0.2227 | 0.035 | 1.98E-10 | Yes | No |
| rs7187540 | 16 | 85318302 | A | C | 0.3351 | -0.1951 | 0.0341 | 1.02E-08 | No | No |
| rs3851018 | 16 | 86437811 | C | G | 0.5693 | 0.1918 | 0.0309 | 5.40E-10 | No | No |
| rs6540125 | 16 | 87993889 | T | G | 0.3411 | 0.2042 | 0.0317 | 1.21E-10 | Yes | No |
| rs1126464 | 16 | 89704365 | C | G | 0.2428 | 0.1285 | 0.036 | 0.0003654 | Yes | No |
| rs12941318 | 17 | 1333598 | T | C | 0.4955 | -0.2351 | 0.0309 | 2.74E-14 | Yes | No |
| rs4480845 | 17 | 1958609 | T | C | 0.3652 | 0.3156 | 0.0316 | 1.85E-23 | Yes | No |
| rs7215084 | 17 | 3880148 | T | C | 0.5127 | 0.095 | 0.03 | 0.001537 | Yes | No |
| rs28427409 | 17 | 6473882 | T | C | 0.4169 | 0.2187 | 0.0306 | 8.84E-13 | Yes | No |
| rs78378222 | 17 | 7571752 | T | G | 0.9859 | 0.4549 | 0.1384 | 0.001017 | Yes | No |
| rs8069739 | 17 | 8078765 | T | C | 0.3217 | -0.1594 | 0.0321 | 6.80E-07 | Yes | No |
| rs4925159 | 17 | 18185510 | A | G | 0.4246 | 0.2174 | 0.0305 | 9.66E-13 | Yes | Yes |
| rs7502046 | 17 | 19196440 | T | C | 0.8103 | 0.1769 | 0.0389 | 5.46E-06 | Yes | No |
| rs138285687 | 17 | 27195674 | T | C | 0.0431 | -0.3474 | 0.076 | 4.81E-06 | No | No |
| rs11080134 | 17 | 29161503 | A | G | 0.6432 | -0.1682 | 0.032 | 1.46E-07 | Yes | Yes |
| rs1551355 | 17 | 30032420 | T | C | 0.2334 | 0.2098 | 0.0356 | 3.89E-09 | Yes | Yes |
| rs9899540 | 17 | 30777924 | A | T | 0.3999 | 0.2011 | 0.0316 | 1.87E-10 | Yes | No |
| rs3135967 | 17 | 33313729 | A | G | 0.5238 | 0.0642 | 0.03 | 0.03248 | Yes | No |
| rs79089478 | 17 | 40317241 | T | C | 0.9726 | 0.3327 | 0.0946 | 0.0004378 | Yes | No |
| rs56228409 | 17 | 40919596 | A | C | 0.8461 | -0.1854 | 0.0438 | 2.36E-05 | Yes | No |
| rs9904409 | 17 | 42680402 | A | G | 0.0974 | 0.3682 | 0.0508 | 4.20E-13 | Yes | No |
| rs12946454 | 17 | 43208121 | A | T | 0.7348 | -0.4125 | 0.0341 | 1.01E-33 | Yes | Yes |
| rs115231027 | 17 | 44199290 | T | C | 0.8216 | -0.2968 | 0.0418 | 1.23E-12 | No | No |
| rs17608766 | 17 | 45013271 | T | C | 0.8555 | -0.6903 | 0.0433 | 2.48E-57 | Yes | No |
| rs62076103 | 17 | 45888374 | A | G | 0.9312 | -0.5803 | 0.0604 | 7.91E-22 | Yes | No |
| rs7406910 | 17 | 46688256 | T | C | 0.0884 | -0.4347 | 0.0533 | 3.39E-16 | Yes | Yes |
| rs12940887 | 17 | 47402807 | T | C | 0.3671 | 0.2597 | 0.0312 | 8.03E-17 | Yes | No |
| rs12325702 | 17 | 55446364 | A | T | 0.4433 | 0.1263 | 0.0307 | 3.78E-05 | No | No |
| rs34430710 | 17 | 56876627 | A | T | 0.6763 | -0.2116 | 0.0322 | 5.02E-11 | Yes | No |
| rs2645466 | 17 | 57853214 | A | C | 0.7045 | -0.1603 | 0.0329 | 1.08E-06 | Yes | No |
| rs1036902 | 17 | 58950791 | T | C | 0.8469 | -0.2543 | 0.0422 | 1.70E-09 | Yes | No |
| rs2240736 | 17 | 59485393 | T | C | 0.7344 | 0.4088 | 0.0343 | 1.17E-32 | Yes | Yes |
| rs740698 | 17 | 60767151 | T | C | 0.5651 | -0.2104 | 0.0308 | 8.48E-12 | Yes | No |
| rs4308 | 17 | 61559625 | A | G | 0.3777 | 0.2731 | 0.0314 | 3.11E-18 | Yes | No |
| rs6504213 | 17 | 62381714 | T | C | 0.4182 | -0.2982 | 0.0312 | 1.25E-21 | Yes | No |
| rs112260610 | 17 | 64252393 | T | C | 0.1403 | 0.2588 | 0.0435 | 2.69E-09 | Yes | No |
| rs2467099 | 17 | 73949045 | T | C | 0.2232 | -0.2462 | 0.036 | 8.26E-12 | Yes | No |
| rs35504735 | 17 | 74686809 | A | G | 0.5576 | 0.024 | 0.0302 | 0.4261 | Yes | No |
| rs57927100 | 17 | 75317300 | C | G | 0.7371 | 0.3149 | 0.0347 | 1.13E-19 | Yes | No |
| rs9302885 | 17 | 76799898 | A | G | 0.4452 | 0.2242 | 0.0302 | 1.03E-13 | Yes | Yes |
| rs112280096 | 17 | 79367409 | A | C | 0.3624 | -0.203 | 0.0335 | 1.33E-09 | No | No |
| rs34413141 | 18 | 777282 | A | T | 0.1822 | -0.3531 | 0.0393 | 2.47E-19 | Yes | No |
| rs11665020 | 18 | 10879503 | C | G | 0.3211 | -0.1304 | 0.0324 | 5.87E-05 | Yes | No |
| rs963920 | 18 | 12711052 | T | G | 0.675 | -0.1022 | 0.0321 | 0.001435 | Yes | No |
| rs4800420 | 18 | 20158965 | A | G | 0.2923 | 0.0936 | 0.0332 | 0.0048 | Yes | No |
| rs1154214 | 18 | 24546824 | T | G | 0.3963 | -0.2031 | 0.0306 | 3.27E-11 | Yes | No |
| rs10164193 | 18 | 31161426 | T | G | 0.9221 | -0.2782 | 0.0566 | 8.93E-07 | Yes | No |
| rs61735998 | 18 | 34289285 | T | G | 0.0241 | 0.5567 | 0.1033 | 7.01E-08 | Yes | No |
| rs12958173 | 18 | 42141977 | A | C | 0.297 | 0.295 | 0.0329 | 2.97E-19 | Yes | Yes |
| rs7236548 | 18 | 43097750 | A | C | 0.1848 | 0.3431 | 0.0388 | 8.51E-19 | Yes | No |
| rs745821 | 18 | 48142854 | T | G | 0.7548 | 0.2214 | 0.0351 | 2.80E-10 | Yes | No |
| rs36010659 | 18 | 48283949 | T | C | 0.8586 | 0.2912 | 0.0432 | 1.55E-11 | Yes | No |
| rs11876341 | 18 | 48799991 | A | G | 0.6919 | -0.213 | 0.0334 | 1.82E-10 | Yes | No |
| rs34163044 | 18 | 51851616 | A | C | 0.4199 | 0.1753 | 0.031 | 1.59E-08 | Yes | No |
| rs72930904 | 18 | 52607301 | T | C | 0.1597 | -0.2316 | 0.041 | 1.57E-08 | Yes | No |
| rs12605156 | 18 | 53498114 | A | T | 0.8081 | 0.2044 | 0.0383 | 9.60E-08 | Yes | No |
| rs10048404 | 18 | 54578482 | T | C | 0.3701 | -0.2607 | 0.0317 | 1.91E-16 | No | No |
| rs7235890 | 18 | 55732115 | T | G | 0.8957 | -0.2125 | 0.0498 | 2.01E-05 | Yes | No |
| rs6567160 | 18 | 57829135 | T | C | 0.7661 | 0.2242 | 0.0357 | 3.33E-10 | Yes | No |
| rs12172847 | 18 | 60223017 | A | G | 0.3229 | -0.1436 | 0.032 | 7.22E-06 | Yes | No |
| rs12454712 | 18 | 60845884 | T | C | 0.6229 | 0.1908 | 0.0328 | 5.82E-09 | No | No |
| rs10460108 | 18 | 73034151 | A | G | 0.4801 | 0.2141 | 0.0301 | 1.12E-12 | Yes | No |
| rs1047922 | 18 | 74070562 | T | C | 0.8486 | -0.1923 | 0.0437 | 1.10E-05 | No | No |
| rs7250835 | 19 | 670234 | T | C | 0.1648 | 0.2067 | 0.0428 | 1.40E-06 | Yes | No |
| rs3760994 | 19 | 1435771 | A | G | 0.4915 | -0.1787 | 0.032 | 2.29E-08 | No | No |
| rs740406 | 19 | 2232221 | A | G | 0.9401 | -0.5905 | 0.0659 | 3.25E-19 | Yes | No |
| rs2656523 | 19 | 4085896 | C | G | 0.7735 | 0.1398 | 0.037 | 0.0001607 | Yes | No |
| rs2613765 | 19 | 5066330 | A | G | 0.4747 | -0.2353 | 0.0301 | 5.32E-15 | Yes | No |
| rs7248104 | 19 | 7224431 | A | G | 0.4124 | -0.2326 | 0.0306 | 2.74E-14 | Yes | No |
| rs4247374 | 19 | 7252756 | T | C | 0.1394 | -0.6354 | 0.0456 | 4.52E-44 | No | No |
| rs2009733 | 19 | 8398714 | A | G | 0.4999 | 0.1148 | 0.0306 | 0.0001758 | Yes | No |
| rs10409243 | 19 | 10332988 | T | C | 0.5904 | -0.3077 | 0.0313 | 8.10E-23 | Yes | No |
| rs1529744 | 19 | 10841472 | T | C | 0.3183 | -0.1542 | 0.0323 | 1.82E-06 | Yes | No |
| rs167479 | 19 | 11526765 | T | G | 0.4726 | -0.5642 | 0.0327 | 7.21E-67 | No | No |
| rs17638167 | 19 | 11584818 | T | C | 0.0439 | -0.4519 | 0.0741 | 1.07E-09 | Yes | No |
| rs10418305 | 19 | 15278808 | C | G | 0.0991 | -0.1619 | 0.0505 | 0.00135 | Yes | No |
| rs3745318 | 19 | 16436262 | T | C | 0.2494 | 0.1637 | 0.0359 | 5.06E-06 | No | No |
| rs1077795 | 19 | 17222584 | A | G | 0.7393 | 0.2507 | 0.0344 | 3.33E-13 | Yes | No |
| rs8111708 | 19 | 18558876 | A | G | 0.653 | 0.0644 | 0.0316 | 0.04185 | Yes | No |
| rs2304130 | 19 | 19789528 | A | G | 0.9157 | -0.3387 | 0.0552 | 8.22E-10 | Yes | No |
| rs6511291 | 19 | 21950402 | T | C | 0.4361 | -0.1692 | 0.0308 | 3.95E-08 | Yes | No |
| rs62104477 | 19 | 30294991 | T | G | 0.3302 | 0.1078 | 0.0321 | 0.0007895 | Yes | No |
| rs8105753 | 19 | 31927547 | A | C | 0.6315 | 0.2178 | 0.0318 | 6.84E-12 | No | No |
| rs1821295 | 19 | 32590773 | T | C | 0.6987 | -0.2253 | 0.0328 | 6.75E-12 | Yes | No |
| rs7256564 | 19 | 33889593 | A | G | 0.3122 | 0.1955 | 0.0324 | 1.53E-09 | Yes | No |
| rs12983238 | 19 | 39438532 | A | G | 0.3064 | -0.1293 | 0.0351 | 0.0002276 | Yes | No |
| rs1800470 | 19 | 41858921 | A | G | 0.6236 | -0.1011 | 0.0313 | 0.00122 | Yes | No |
| rs7412 | 19 | 45412079 | T | C | 0.0819 | -0.4339 | 0.0574 | 4.04E-14 | Yes | No |
| rs34783010 | 19 | 46180414 | T | G | 0.1996 | 0.2208 | 0.0381 | 6.79E-09 | Yes | No |
| rs73046792 | 19 | 49605705 | A | G | 0.1588 | -0.3554 | 0.0426 | 7.23E-17 | Yes | No |
| rs138877676 | 19 | 50935809 | T | G | 0.0193 | -0.66 | 0.1237 | 9.53E-08 | Yes | No |
| rs2143635 | 20 | 2793063 | T | C | 0.8996 | -0.1676 | 0.0514 | 0.001096 | Yes | No |
| rs1764975 | 20 | 4101290 | A | T | 0.7954 | 0.2819 | 0.0379 | 1.08E-13 | Yes | No |
| rs11087740 | 20 | 6657554 | T | C | 0.5116 | -0.1627 | 0.0303 | 7.91E-08 | Yes | No |
| rs6108168 | 20 | 8626271 | A | C | 0.2542 | -0.2957 | 0.0344 | 8.71E-18 | Yes | No |
| rs680515 | 20 | 10458688 | A | G | 0.3893 | -0.0876 | 0.031 | 0.004681 | Yes | No |
| rs1327235 | 20 | 10969030 | A | G | 0.5295 | -0.426 | 0.03 | 7.88E-46 | Yes | No |
| rs1232482 | 20 | 11886643 | T | C | 0.4015 | -0.1379 | 0.0306 | 6.42E-06 | Yes | No |
| rs2618647 | 20 | 17882452 | A | G | 0.5055 | -0.1884 | 0.0301 | 4.01E-10 | Yes | No |
| rs6081613 | 20 | 19465907 | A | G | 0.2744 | 0.1815 | 0.0336 | 6.58E-08 | Yes | No |
| rs6060114 | 20 | 30169673 | T | C | 0.8416 | 0.2731 | 0.0413 | 3.79E-11 | Yes | No |
| rs6141767 | 20 | 31225069 | C | G | 0.157 | 0.3107 | 0.0419 | 1.17E-13 | Yes | No |
| rs13042148 | 20 | 32298286 | T | C | 0.1534 | -0.1344 | 0.0424 | 0.001506 | Yes | No |
| rs6141479 | 20 | 33121942 | C | G | 0.1968 | 0.0997 | 0.0392 | 0.011 | Yes | No |
| rs4811601 | 20 | 36849007 | T | C | 0.4588 | 0.0642 | 0.0306 | 0.03582 | Yes | No |
| rs4810332 | 20 | 40268334 | A | T | 0.3792 | -0.2608 | 0.0317 | 1.92E-16 | Yes | No |
| rs6031435 | 20 | 42797358 | A | G | 0.5398 | -0.2592 | 0.0303 | 1.09E-17 | Yes | No |
| rs6095241 | 20 | 47308798 | A | G | 0.4395 | -0.1718 | 0.0302 | 1.26E-08 | Yes | No |
| rs237485 | 20 | 48004238 | A | G | 0.698 | 0.1101 | 0.0331 | 0.0008706 | Yes | No |
| rs6021247 | 20 | 50108980 | A | G | 0.529 | 0.2328 | 0.0301 | 9.80E-15 | Yes | Yes |
| rs6015450 | 20 | 57751117 | A | G | 0.8761 | -0.7003 | 0.0461 | 3.95E-52 | Yes | No |
| rs35213536 | 20 | 62694319 | T | G | 0.2466 | 0.2937 | 0.0357 | 1.94E-16 | Yes | No |
| rs1882961 | 21 | 16556367 | T | C | 0.3087 | 0.2443 | 0.0326 | 6.69E-14 | Yes | Yes |
| rs11909120 | 21 | 30131872 | A | T | 0.857 | -0.0916 | 0.043 | 0.03324 | Yes | No |
| rs11701033 | 21 | 33788341 | C | G | 0.8178 | -0.2187 | 0.0392 | 2.52E-08 | Yes | No |
| rs9976596 | 21 | 35596842 | T | C | 0.843 | 0.097 | 0.0424 | 0.02209 | Yes | No |
| rs62229372 | 21 | 37692507 | T | C | 0.1275 | 0.2408 | 0.047 | 2.95E-07 | Yes | No |
| rs2277788 | 21 | 40817702 | C | G | 0.1054 | 0.2295 | 0.0489 | 2.67E-06 | Yes | No |
| rs12627651 | 21 | 44760603 | A | G | 0.2872 | 0.3498 | 0.0341 | 1.02E-24 | Yes | No |
| rs9306160 | 21 | 45107562 | T | C | 0.4107 | -0.2123 | 0.0309 | 6.35E-12 | Yes | No |
| rs35796750 | 21 | 47422412 | T | C | 0.5502 | -0.1337 | 0.0305 | 1.19E-05 | Yes | No |
| rs11701512 | 21 | 47962811 | A | G | 0.1809 | 0.1958 | 0.039 | 5.24E-07 | Yes | No |
| rs12628032 | 22 | 19967980 | T | C | 0.3094 | 0.2445 | 0.0328 | 9.81E-14 | Yes | No |
| rs134041 | 22 | 28056338 | T | C | 0.4358 | 0.0193 | 0.0303 | 0.5252 | Yes | No |
| rs9608690 | 22 | 28921347 | A | G | 0.0682 | -0.3711 | 0.06 | 6.13E-10 | Yes | No |
| rs4823006 | 22 | 29451671 | A | G | 0.5553 | 0.2172 | 0.0302 | 6.23E-13 | Yes | No |
| rs737721 | 22 | 30172254 | C | G | 0.9478 | -0.4074 | 0.0684 | 2.62E-09 | Yes | No |
| rs5753103 | 22 | 30768777 | A | G | 0.4519 | 0.1303 | 0.0303 | 1.74E-05 | Yes | No |
| rs9609429 | 22 | 32517431 | T | C | 0.7243 | 0.1304 | 0.0337 | 0.0001087 | Yes | No |
| rs5750482 | 22 | 38117943 | T | C | 0.3813 | 0.1578 | 0.031 | 3.67E-07 | Yes | No |
| rs470113 | 22 | 40729614 | A | G | 0.8182 | -0.1849 | 0.039 | 2.07E-06 | Yes | No |
| rs73161324 | 22 | 42038786 | T | C | 0.0562 | 0.203 | 0.0702 | 0.003837 | Yes | No |
| rs77692990 | 22 | 50219952 | T | C | 0.0797 | -0.3178 | 0.0566 | 1.95E-08 | Yes | No |
| rs28578714 | 22 | 50727921 | T | C | 0.6062 | 0.2066 | 0.0327 | 2.53E-10 | Yes | No |

**Supplemental Table 8.** SNPs included in diastolic blood pressure-related analyses.

| RSID | Chrom. | Position | Effect allele | Other allele | Effect allele freq. | Beta | SE | p-value | Used in MR | Used in MR + Steiger filt. |
| --- | --- | --- | --- | --- | --- | --- | --- | --- | --- | --- |
| rs2076328 | 1 | 1687482 | T | G | 0.49 | -0.1318 | 0.0178 | 1.33E-13 | Yes | No |
| rs260508 | 1 | 2187085 | T | G | 0.6148 | 0.0548 | 0.0178 | 0.002125 | Yes | No |
| rs2493292 | 1 | 3328659 | T | C | 0.144 | 0.2481 | 0.0251 | 5.54E-23 | Yes | Yes |
| rs709209 | 1 | 6278414 | A | G | 0.6584 | -0.0357 | 0.0188 | 0.05666 | Yes | No |
| rs4908678 | 1 | 7739250 | T | C | 0.626 | -0.1124 | 0.018 | 4.59E-10 | Yes | Yes |
| rs2252865 | 1 | 8422676 | T | C | 0.3499 | 0.1189 | 0.0181 | 5.51E-11 | Yes | Yes |
| rs9662255 | 1 | 9441949 | A | C | 0.4273 | -0.0041 | 0.0177 | 0.8184 | Yes | No |
| rs880315 | 1 | 10796866 | T | C | 0.6567 | -0.2583 | 0.0185 | 1.99E-44 | Yes | No |
| rs17367504 | 1 | 11862778 | A | G | 0.8406 | 0.511 | 0.0237 | 3.55E-103 | Yes | No |
| rs3820068 | 1 | 15798197 | A | G | 0.8042 | 0.0953 | 0.0221 | 1.65E-05 | Yes | No |
| rs2807337 | 1 | 22577371 | T | C | 0.3642 | 0.0901 | 0.018 | 5.54E-07 | Yes | No |
| rs150266910 | 1 | 23442265 | T | C | 0.1767 | -0.0651 | 0.0226 | 0.003997 | Yes | No |
| rs6686889 | 1 | 25030470 | T | C | 0.2533 | 0.1918 | 0.0199 | 6.95E-22 | Yes | No |
| rs79598313 | 1 | 27284913 | T | C | 0.0251 | 0.3286 | 0.057 | 8.13E-09 | Yes | No |
| rs3737801 | 1 | 27960832 | C | G | 0.9231 | 0.1855 | 0.0344 | 7.08E-08 | Yes | No |
| rs143167197 | 1 | 28734372 | A | G | 0.9284 | -0.1242 | 0.0352 | 0.0004161 | Yes | No |
| rs1565716 | 1 | 29549216 | A | G | 0.0698 | 0.2139 | 0.0341 | 3.45E-10 | Yes | No |
| rs4652875 | 1 | 33868469 | C | G | 0.4207 | 0.0319 | 0.0175 | 0.06864 | No | No |
| rs9729719 | 1 | 38298207 | A | G | 0.2929 | -0.0503 | 0.0193 | 0.009217 | Yes | No |
| rs11210029 | 1 | 41865293 | A | G | 0.6325 | -0.0917 | 0.018 | 3.39E-07 | Yes | No |
| rs7515635 | 1 | 42408070 | T | C | 0.4612 | 0.0979 | 0.0174 | 1.82E-08 | Yes | No |
| rs72659998 | 1 | 43037556 | T | C | 0.1513 | 0.0322 | 0.0244 | 0.1872 | Yes | No |
| rs839755 | 1 | 43856410 | A | C | 0.6146 | -0.1483 | 0.0177 | 5.22E-17 | Yes | No |
| rs512083 | 1 | 46027355 | T | C | 0.5349 | 0.0161 | 0.0173 | 0.3525 | Yes | No |
| rs12142296 | 1 | 46541679 | T | G | 0.8639 | -0.1643 | 0.0253 | 8.89E-11 | Yes | No |
| rs4926923 | 1 | 48109225 | T | C | 0.9117 | 0.1918 | 0.0308 | 4.75E-10 | Yes | Yes |
| rs11579440 | 1 | 49052423 | T | C | 0.8482 | 0.1303 | 0.0243 | 8.57E-08 | Yes | No |
| rs147696085 | 1 | 51021867 | A | G | 0.0934 | 0.1533 | 0.0302 | 3.98E-07 | Yes | No |
| rs6681713 | 1 | 51527684 | T | G | 0.9813 | 0.4146 | 0.0656 | 2.66E-10 | Yes | No |
| rs112557609 | 1 | 56576924 | A | G | 0.3441 | 0.0685 | 0.0183 | 0.0001774 | Yes | No |
| rs2404715 | 1 | 57008778 | T | C | 0.0926 | -0.0282 | 0.03 | 0.3469 | Yes | No |
| rs60199046 | 1 | 59663341 | A | G | 0.7102 | -0.0884 | 0.019 | 3.28E-06 | Yes | No |
| rs20354 | 1 | 67071356 | T | G | 0.1333 | -0.0072 | 0.0254 | 0.7764 | Yes | No |
| rs34517439 | 1 | 78450517 | A | C | 0.1199 | -0.2514 | 0.0279 | 2.02E-19 | No | No |
| rs12034319 | 1 | 86040107 | A | G | 0.2202 | -0.0095 | 0.0209 | 0.6492 | Yes | No |
| rs385437 | 1 | 86822231 | A | G | 0.8607 | 0.0187 | 0.0251 | 0.4557 | Yes | No |
| rs10923038 | 1 | 88651771 | A | C | 0.6175 | 0.0847 | 0.0179 | 2.32E-06 | Yes | No |
| rs10922502 | 1 | 89360158 | A | G | 0.6269 | -0.1094 | 0.0179 | 1.10E-09 | Yes | No |
| rs2065152 | 1 | 90228519 | T | C | 0.3574 | 0.1103 | 0.018 | 8.95E-10 | Yes | Yes |
| rs17516329 | 1 | 92319781 | A | T | 0.6861 | -0.0376 | 0.0188 | 0.04519 | Yes | No |
| rs7514579 | 1 | 94051350 | A | C | 0.7706 | 0.1121 | 0.0207 | 6.16E-08 | Yes | No |
| rs17396055 | 1 | 94730954 | A | G | 0.3324 | -0.115 | 0.0184 | 4.13E-10 | Yes | Yes |
| rs17030613 | 1 | 113190807 | A | C | 0.7905 | -0.2847 | 0.0213 | 8.22E-41 | Yes | Yes |
| rs2932538 | 1 | 113216543 | A | G | 0.2566 | -0.2435 | 0.0198 | 8.85E-35 | Yes | No |
| rs12078697 | 1 | 117015118 | C | G | 0.2113 | -0.1077 | 0.0213 | 4.07E-07 | Yes | Yes |
| rs7553422 | 1 | 119540719 | T | C | 0.4118 | -0.1097 | 0.0176 | 4.17E-10 | Yes | No |
| rs72704264 | 1 | 145713305 | C | G | 0.2172 | 0.117 | 0.0212 | 3.60E-08 | Yes | No |
| rs11585169 | 1 | 150572037 | A | T | 0.5774 | 0.0274 | 0.0176 | 0.1202 | No | No |
| rs13796 | 1 | 154245917 | T | C | 0.8641 | -0.1626 | 0.0257 | 2.44E-10 | Yes | No |
| rs76719272 | 1 | 156129796 | T | C | 0.1315 | -0.1438 | 0.0264 | 4.86E-08 | Yes | No |
| rs2171690 | 1 | 164740099 | T | C | 0.5352 | 0.1181 | 0.0174 | 1.19E-11 | Yes | Yes |
| rs7524019 | 1 | 167367193 | T | C | 0.492 | 0.1036 | 0.0174 | 2.60E-09 | Yes | No |
| rs2157597 | 1 | 169201567 | T | C | 0.3558 | 0.034 | 0.0181 | 0.06064 | Yes | No |
| rs12405515 | 1 | 172357441 | T | G | 0.5702 | -0.1698 | 0.0174 | 1.92E-22 | Yes | No |
| rs12118102 | 1 | 176634724 | A | G | 0.947 | -0.1766 | 0.0387 | 5.16E-06 | Yes | No |
| rs150816167 | 1 | 179571862 | T | C | 0.9549 | -0.2873 | 0.0446 | 1.17E-10 | Yes | No |
| rs10913934 | 1 | 180131640 | T | G | 0.5935 | -0.0254 | 0.0176 | 0.1503 | Yes | No |
| rs1043069 | 1 | 180859368 | T | G | 0.6148 | 0.1123 | 0.0178 | 2.93E-10 | Yes | No |
| rs41475048 | 1 | 183058452 | A | G | 0.7451 | -0.1233 | 0.0202 | 9.93E-10 | Yes | No |
| rs4651224 | 1 | 184585182 | T | C | 0.4469 | 0.1102 | 0.0175 | 3.39E-10 | Yes | No |
| rs12042924 | 1 | 197297417 | T | C | 0.5287 | -0.0604 | 0.0174 | 0.0005231 | Yes | No |
| rs882624 | 1 | 201735913 | T | C | 0.3325 | -0.1571 | 0.0185 | 2.33E-17 | No | No |
| rs33996239 | 1 | 203109801 | T | C | 0.0607 | -0.2513 | 0.0378 | 2.86E-11 | Yes | No |
| rs4245739 | 1 | 204518842 | A | C | 0.7279 | 0.1591 | 0.0195 | 3.44E-16 | Yes | No |
| rs2629665 | 1 | 207220800 | A | C | 0.4096 | -0.1193 | 0.0177 | 1.52E-11 | Yes | No |
| rs2761436 | 1 | 207919748 | T | C | 0.5379 | 0.0093 | 0.0173 | 0.5929 | Yes | No |
| rs7555285 | 1 | 209970355 | C | G | 0.8012 | 0.1005 | 0.0216 | 3.21E-06 | Yes | No |
| rs12408022 | 1 | 217718789 | T | C | 0.2588 | 0.1483 | 0.0199 | 9.60E-14 | Yes | Yes |
| rs35981664 | 1 | 218549354 | A | T | 0.6881 | -0.1606 | 0.0189 | 2.01E-17 | Yes | No |
| rs2820443 | 1 | 219753509 | T | C | 0.7078 | -0.0308 | 0.0189 | 0.104 | Yes | No |
| rs9431431 | 1 | 221358796 | A | G | 0.294 | -0.134 | 0.019 | 1.71E-12 | Yes | No |
| rs73091767 | 1 | 227250775 | T | C | 0.7345 | -0.1346 | 0.0195 | 4.77E-12 | Yes | No |
| rs2004776 | 1 | 230848702 | T | C | 0.2403 | 0.2513 | 0.0203 | 2.55E-35 | Yes | Yes |
| rs6429422 | 1 | 243472801 | T | G | 0.678 | -0.246 | 0.0185 | 3.29E-40 | Yes | Yes |
| rs4926499 | 1 | 249155909 | C | G | 0.826 | 0.1694 | 0.0248 | 9.37E-12 | No | No |
| rs4850047 | 2 | 3634753 | T | C | 0.1455 | -0.1244 | 0.0255 | 1.10E-06 | Yes | No |
| rs2175337 | 2 | 9298590 | A | C | 0.6108 | -0.024 | 0.0178 | 0.1765 | Yes | No |
| rs67720684 | 2 | 18975439 | A | C | 0.239 | 0.1014 | 0.0203 | 5.73E-07 | Yes | No |
| rs1344653 | 2 | 19730845 | A | G | 0.4996 | 0.055 | 0.0172 | 0.001397 | Yes | No |
| rs7255 | 2 | 20878820 | T | C | 0.453 | 0.0345 | 0.0175 | 0.04818 | Yes | No |
| rs66774912 | 2 | 21423532 | A | G | 0.1368 | 0.0249 | 0.0253 | 0.3263 | Yes | No |
| rs10779936 | 2 | 23950200 | A | G | 0.7134 | 0.0099 | 0.0191 | 0.6054 | Yes | No |
| rs55701159 | 2 | 25139596 | T | G | 0.8864 | 0.2254 | 0.0275 | 2.47E-16 | Yes | No |
| rs1275988 | 2 | 26914364 | T | C | 0.6111 | -0.2945 | 0.0177 | 1.92E-62 | Yes | No |
| rs9678851 | 2 | 27887034 | A | C | 0.568 | -0.076 | 0.0176 | 1.49E-05 | Yes | No |
| rs7562 | 2 | 28635740 | T | C | 0.5192 | 0.1059 | 0.0175 | 1.33E-09 | Yes | No |
| rs1607644 | 2 | 34679626 | A | G | 0.3658 | -0.0934 | 0.0179 | 1.95E-07 | Yes | Yes |
| rs13420463 | 2 | 37517566 | A | G | 0.7725 | 0.1645 | 0.0206 | 1.60E-15 | Yes | No |
| rs2707238 | 2 | 38094149 | C | G | 0.2839 | 0.1037 | 0.0192 | 6.81E-08 | Yes | No |
| rs4952611 | 2 | 40567743 | T | C | 0.5795 | -0.1401 | 0.018 | 7.38E-15 | Yes | No |
| rs11681462 | 2 | 42352567 | A | C | 0.7885 | -0.1325 | 0.0214 | 5.96E-10 | Yes | No |
| rs76326501 | 2 | 43167878 | A | C | 0.9089 | 0.3618 | 0.0305 | 2.17E-32 | Yes | No |
| rs35590893 | 2 | 43716933 | A | G | 0.2776 | -0.1141 | 0.0193 | 3.23E-09 | Yes | No |
| rs11690961 | 2 | 46363336 | A | C | 0.8832 | -0.1611 | 0.0269 | 2.22E-09 | Yes | No |
| rs6545155 | 2 | 50429861 | T | C | 0.7818 | 0.0844 | 0.0209 | 5.49E-05 | Yes | No |
| rs10189186 | 2 | 53025757 | A | G | 0.5295 | 0.0971 | 0.0173 | 2.19E-08 | Yes | No |
| rs2920899 | 2 | 55279681 | T | G | 0.7859 | 0.0743 | 0.0213 | 0.0004863 | Yes | No |
| rs1975487 | 2 | 55809054 | A | G | 0.4814 | -0.141 | 0.0176 | 1.01E-15 | Yes | No |
| rs6730325 | 2 | 59315828 | A | G | 0.6048 | -0.0231 | 0.0177 | 0.1905 | Yes | No |
| rs72816333 | 2 | 60096560 | A | T | 0.8302 | 0.1432 | 0.023 | 4.66E-10 | Yes | No |
| rs925484 | 2 | 60611437 | C | G | 0.5997 | -0.0233 | 0.0177 | 0.1874 | Yes | No |
| rs7608483 | 2 | 61836235 | A | C | 0.4171 | 0.1171 | 0.0176 | 2.83E-11 | Yes | No |
| rs13014371 | 2 | 64217786 | T | C | 0.5705 | -0.1176 | 0.0175 | 1.68E-11 | Yes | No |
| rs2631669 | 2 | 66104881 | T | C | 0.4705 | -0.0541 | 0.0174 | 0.001831 | Yes | No |
| rs2300481 | 2 | 66782467 | T | C | 0.3861 | 0.0853 | 0.0178 | 1.56E-06 | Yes | No |
| rs6731373 | 2 | 68503044 | A | G | 0.3503 | 0.0718 | 0.0186 | 0.0001116 | Yes | No |
| rs12052761 | 2 | 69065841 | A | G | 0.3938 | -0.1229 | 0.0177 | 4.22E-12 | Yes | No |
| rs3771371 | 2 | 71627539 | T | C | 0.569 | -0.0098 | 0.0174 | 0.5742 | Yes | No |
| rs10193543 | 2 | 72483329 | T | G | 0.8364 | 0.1391 | 0.0235 | 3.20E-09 | Yes | No |
| rs1876487 | 2 | 73114352 | A | C | 0.2948 | -0.1099 | 0.0196 | 1.93E-08 | Yes | Yes |
| rs11689667 | 2 | 85491365 | T | C | 0.5443 | -0.0153 | 0.0174 | 0.3792 | Yes | No |
| rs72847885 | 2 | 86326717 | A | G | 0.663 | 0.1305 | 0.0182 | 8.08E-13 | Yes | No |
| rs2579519 | 2 | 96675166 | T | C | 0.6169 | -0.1818 | 0.018 | 4.24E-24 | Yes | Yes |
| rs4851462 | 2 | 98357163 | T | C | 0.6246 | -0.1199 | 0.0182 | 3.96E-11 | No | No |
| rs150194832 | 2 | 106126880 | C | G | 0.093 | -0.0675 | 0.03 | 0.02458 | Yes | No |
| rs28377357 | 2 | 112769721 | A | G | 0.2938 | -0.1243 | 0.019 | 6.03E-11 | No | No |
| rs62158170 | 2 | 114082175 | A | G | 0.7834 | 0.1645 | 0.0211 | 6.63E-15 | Yes | No |
| rs10864859 | 2 | 121440218 | T | G | 0.9163 | 0.1962 | 0.0325 | 1.52E-09 | Yes | No |
| rs6723509 | 2 | 122000745 | T | C | 0.8612 | 0.1125 | 0.0251 | 7.49E-06 | Yes | No |
| rs13001283 | 2 | 127183454 | A | G | 0.1596 | 0.1522 | 0.0239 | 1.92E-10 | Yes | No |
| rs4954192 | 2 | 135632981 | T | C | 0.3872 | -0.1225 | 0.0179 | 8.15E-12 | Yes | No |
| rs72844590 | 2 | 138421227 | T | G | 0.1508 | 0.113 | 0.0247 | 4.61E-06 | Yes | No |
| rs7606205 | 2 | 144146311 | A | C | 0.7027 | -0.1276 | 0.0191 | 2.36E-11 | Yes | No |
| rs1438896 | 2 | 145646072 | T | C | 0.2981 | 0.195 | 0.0189 | 4.56E-25 | Yes | Yes |
| rs34570306 | 2 | 146272860 | T | C | 0.5275 | -0.1197 | 0.0177 | 1.19E-11 | Yes | No |
| rs62169544 | 2 | 146950908 | A | G | 0.4429 | -0.1207 | 0.0175 | 4.96E-12 | Yes | No |
| rs12990959 | 2 | 148572160 | T | C | 0.6875 | -0.1271 | 0.0187 | 1.11E-11 | Yes | Yes |
| rs4664080 | 2 | 152978341 | A | G | 0.3978 | -0.0129 | 0.0177 | 0.4664 | Yes | No |
| rs3175 | 2 | 153618773 | A | G | 0.6523 | 0.0329 | 0.0186 | 0.07645 | Yes | No |
| rs1220128 | 2 | 158499902 | C | G | 0.852 | 0.1917 | 0.0246 | 6.16E-15 | Yes | Yes |
| rs79523138 | 2 | 161368213 | A | G | 0.8799 | -0.1232 | 0.0271 | 5.30E-06 | Yes | No |
| rs55732192 | 2 | 162278233 | T | G | 0.0944 | -0.1417 | 0.0299 | 2.12E-06 | Yes | No |
| rs1446468 | 2 | 164963486 | T | C | 0.454 | -0.253 | 0.0174 | 1.21E-47 | Yes | No |
| rs6712203 | 2 | 165557318 | T | C | 0.3719 | -0.1117 | 0.018 | 5.22E-10 | Yes | No |
| rs2390258 | 2 | 166250129 | A | G | 0.3074 | -0.0998 | 0.0188 | 1.08E-07 | Yes | Yes |
| rs560887 | 2 | 169763148 | T | C | 0.2986 | 0.0482 | 0.0189 | 0.01069 | Yes | No |
| rs151054210 | 2 | 172381487 | A | G | 0.1829 | 0.0136 | 0.0224 | 0.5423 | Yes | No |
| rs6758859 | 2 | 173965056 | T | C | 0.6355 | 0.1211 | 0.0179 | 1.45E-11 | Yes | Yes |
| rs11694601 | 2 | 174949358 | A | G | 0.5969 | -0.0889 | 0.0177 | 5.33E-07 | Yes | No |
| rs72914576 | 2 | 175529967 | C | G | 0.8105 | -0.0626 | 0.0223 | 0.00491 | Yes | No |
| rs60148403 | 2 | 177989414 | A | T | 0.1955 | -0.0391 | 0.0221 | 0.07618 | Yes | No |
| rs1837164 | 2 | 178716601 | A | T | 0.369 | 0.0677 | 0.0179 | 0.0001524 | Yes | No |
| rs79146658 | 2 | 179786068 | T | C | 0.9145 | -0.3344 | 0.0312 | 7.85E-27 | Yes | No |
| rs1486236 | 2 | 180739450 | A | C | 0.3703 | -0.0751 | 0.0184 | 4.33E-05 | Yes | No |
| rs10184839 | 2 | 181946115 | A | T | 0.2917 | -0.1398 | 0.019 | 2.09E-13 | Yes | Yes |
| rs16823124 | 2 | 183224127 | A | G | 0.3069 | 0.2276 | 0.0187 | 4.06E-34 | Yes | Yes |
| rs12473688 | 2 | 185033470 | A | G | 0.284 | 0.0847 | 0.0191 | 9.69E-06 | Yes | No |
| rs28558491 | 2 | 187816321 | T | C | 0.7335 | -0.1074 | 0.0197 | 4.83E-08 | Yes | No |
| rs11901929 | 2 | 189643316 | A | G | 0.3471 | -0.0433 | 0.0182 | 0.01739 | Yes | No |
| rs7592578 | 2 | 191439591 | T | G | 0.1938 | -0.1998 | 0.0224 | 4.71E-19 | Yes | No |
| rs296797 | 2 | 201102905 | T | C | 0.4048 | 0.0682 | 0.0177 | 0.0001113 | Yes | No |
| rs1469760 | 2 | 204125426 | T | C | 0.5834 | -0.0633 | 0.0176 | 0.000317 | Yes | No |
| rs2162003 | 2 | 205077128 | T | C | 0.6048 | 0.1279 | 0.0183 | 3.20E-12 | No | No |
| rs1263671 | 2 | 207996447 | T | C | 0.8368 | -0.1394 | 0.0238 | 4.69E-09 | Yes | Yes |
| rs55780018 | 2 | 208526140 | T | C | 0.5442 | -0.1334 | 0.0177 | 5.29E-14 | Yes | No |
| rs1047891 | 2 | 211540507 | A | C | 0.319 | -0.1405 | 0.0188 | 8.19E-14 | Yes | No |
| rs12694277 | 2 | 213188795 | T | C | 0.2951 | -0.0866 | 0.0192 | 6.76E-06 | Yes | No |
| rs4674114 | 2 | 217659266 | A | G | 0.2004 | 0.0224 | 0.0216 | 0.2994 | Yes | No |
| rs1063281 | 2 | 218668732 | T | C | 0.6027 | -0.1623 | 0.0179 | 1.21E-19 | Yes | Yes |
| rs1996992 | 2 | 219651349 | T | G | 0.0516 | -0.2967 | 0.0394 | 4.71E-14 | Yes | Yes |
| rs12474050 | 2 | 220362557 | T | C | 0.3431 | 0.1142 | 0.0182 | 3.80E-10 | Yes | No |
| rs2972146 | 2 | 227100698 | T | G | 0.6431 | 0.1326 | 0.018 | 1.73E-13 | Yes | No |
| rs1044822 | 2 | 230629138 | T | C | 0.1488 | -0.1334 | 0.0243 | 4.14E-08 | Yes | No |
| rs12052878 | 2 | 238227594 | A | G | 0.319 | 0.0284 | 0.0186 | 0.1273 | Yes | No |
| rs4507125 | 2 | 239864732 | A | C | 0.7864 | -0.1244 | 0.0211 | 3.60E-09 | Yes | Yes |
| rs139354822 | 2 | 242344695 | T | C | 0.9706 | 0.2612 | 0.0557 | 2.76E-06 | Yes | No |
| rs9865843 | 3 | 7489993 | A | G | 0.5165 | -0.0923 | 0.0177 | 1.73E-07 | Yes | No |
| rs347591 | 3 | 11290122 | T | G | 0.6628 | 0.1347 | 0.0184 | 2.20E-13 | Yes | No |
| rs729639 | 3 | 13826854 | T | C | 0.3452 | 0.0196 | 0.0183 | 0.2836 | Yes | No |
| rs11128722 | 3 | 14958126 | A | G | 0.5704 | -0.1345 | 0.0178 | 3.82E-14 | Yes | No |
| rs189267552 | 3 | 20073193 | A | T | 0.0131 | -0.0965 | 0.0797 | 0.2261 | Yes | No |
| rs4634143 | 3 | 23163749 | T | C | 0.2996 | 0.1161 | 0.0189 | 7.89E-10 | Yes | No |
| rs13082711 | 3 | 27537909 | T | C | 0.7607 | -0.1778 | 0.0203 | 1.70E-18 | Yes | Yes |
| rs72851229 | 3 | 29374219 | C | G | 0.1745 | -0.1364 | 0.0231 | 3.63E-09 | Yes | No |
| rs12638085 | 3 | 30405936 | A | T | 0.3565 | 0.0976 | 0.0182 | 8.55E-08 | Yes | No |
| rs4678915 | 3 | 36964583 | A | G | 0.4313 | 0.0214 | 0.0178 | 0.2287 | Yes | No |
| rs267517 | 3 | 37539090 | A | G | 0.5927 | -0.1052 | 0.0179 | 4.35E-09 | Yes | No |
| rs6801957 | 3 | 38767315 | T | C | 0.4099 | -0.0428 | 0.0176 | 0.01493 | Yes | No |
| rs6788984 | 3 | 41107173 | A | G | 0.8562 | 0.1143 | 0.0248 | 3.92E-06 | Yes | No |
| rs9815354 | 3 | 41912651 | A | G | 0.163 | 0.3216 | 0.0237 | 5.26E-42 | Yes | No |
| rs141979279 | 3 | 44858131 | T | C | 0.9511 | 0.046 | 0.0402 | 0.2522 | Yes | No |
| rs113134141 | 3 | 46861939 | A | G | 0.8983 | -0.1608 | 0.0288 | 2.50E-08 | Yes | No |
| rs6797587 | 3 | 48197614 | A | G | 0.3285 | -0.2378 | 0.0185 | 6.56E-38 | Yes | Yes |
| rs73082337 | 3 | 49009570 | C | G | 0.8792 | 0.1482 | 0.0275 | 7.12E-08 | Yes | No |
| rs36022378 | 3 | 49913705 | T | C | 0.7997 | -0.1765 | 0.0219 | 8.60E-16 | Yes | No |
| rs13303 | 3 | 52558008 | T | C | 0.4368 | 0.0397 | 0.0176 | 0.02382 | Yes | No |
| rs9810888 | 3 | 53635595 | T | G | 0.4981 | -0.1151 | 0.0175 | 4.38E-11 | Yes | No |
| rs9827472 | 3 | 56726646 | T | C | 0.3649 | -0.133 | 0.0181 | 1.75E-13 | No | No |
| rs12486605 | 3 | 57706503 | T | C | 0.5725 | -0.1508 | 0.0176 | 1.01E-17 | Yes | No |
| rs3774702 | 3 | 63856870 | A | G | 0.1768 | 0.147 | 0.0228 | 1.18E-10 | Yes | Yes |
| rs918466 | 3 | 64710253 | A | G | 0.4092 | -0.1402 | 0.0177 | 2.67E-15 | Yes | Yes |
| rs7630745 | 3 | 66427029 | T | C | 0.6586 | 0.007 | 0.0182 | 0.701 | Yes | No |
| rs4499560 | 3 | 70920485 | A | T | 0.3167 | -0.1141 | 0.0187 | 1.05E-09 | Yes | No |
| rs729448 | 3 | 73260545 | A | G | 0.5482 | 0.0077 | 0.0174 | 0.6575 | Yes | No |
| rs9857362 | 3 | 74710462 | A | C | 0.5296 | 0.0948 | 0.0175 | 6.30E-08 | Yes | No |
| rs1375564 | 3 | 85656311 | T | C | 0.6395 | 0.1133 | 0.0181 | 3.71E-10 | Yes | No |
| rs9860290 | 3 | 99839106 | A | G | 0.21 | 0.0356 | 0.0212 | 0.09373 | Yes | No |
| rs11923667 | 3 | 101268080 | A | T | 0.4071 | 0.1175 | 0.0177 | 3.10E-11 | No | No |
| rs28675079 | 3 | 111500002 | A | G | 0.1867 | -0.1444 | 0.0222 | 8.34E-11 | Yes | No |
| rs1882289 | 3 | 114461208 | A | G | 0.8852 | -0.0188 | 0.0271 | 0.487 | Yes | No |
| rs6806529 | 3 | 123049938 | A | C | 0.4341 | 0.0459 | 0.0177 | 0.009416 | Yes | No |
| rs6438857 | 3 | 124557643 | T | C | 0.5779 | 0.1492 | 0.0175 | 1.50E-17 | Yes | No |
| rs62270945 | 3 | 128201889 | T | C | 0.0291 | -0.111 | 0.0545 | 0.04159 | Yes | No |
| rs9875380 | 3 | 132780356 | T | C | 0.4647 | -0.0925 | 0.0173 | 9.29E-08 | Yes | No |
| rs6783086 | 3 | 133959552 | T | C | 0.3944 | 0.1744 | 0.0177 | 6.26E-23 | Yes | No |
| rs590198 | 3 | 135953729 | A | G | 0.5279 | 0.0863 | 0.0173 | 5.83E-07 | Yes | No |
| rs2306374 | 3 | 138119952 | T | C | 0.838 | -0.1774 | 0.0236 | 5.24E-14 | Yes | No |
| rs16851397 | 3 | 141134818 | A | G | 0.9531 | -0.3942 | 0.0415 | 2.04E-21 | Yes | No |
| rs62278541 | 3 | 142631909 | A | G | 0.6474 | -0.0391 | 0.0181 | 0.03051 | Yes | No |
| rs6772704 | 3 | 149524692 | A | C | 0.6837 | 0.014 | 0.0186 | 0.4513 | Yes | No |
| rs73158427 | 3 | 153721493 | A | T | 0.1617 | 0.1801 | 0.0235 | 1.76E-14 | Yes | No |
| rs143112823 | 3 | 154707967 | A | G | 0.0883 | -0.2246 | 0.0317 | 1.42E-12 | Yes | No |
| rs9833313 | 3 | 157576791 | A | T | 0.242 | -0.0707 | 0.0204 | 0.0005172 | Yes | No |
| rs78151625 | 3 | 158316726 | T | C | 0.8342 | -0.1869 | 0.0233 | 1.04E-15 | Yes | No |
| rs419076 | 3 | 169100886 | T | C | 0.4729 | 0.2755 | 0.0173 | 2.66E-57 | Yes | Yes |
| rs4894535 | 3 | 171995605 | T | C | 0.1602 | 0.0142 | 0.0237 | 0.5489 | Yes | No |
| rs73171158 | 3 | 176927949 | T | C | 0.4674 | -0.0594 | 0.0183 | 0.001176 | No | No |
| rs7611674 | 3 | 179169230 | T | G | 0.8038 | 0.1576 | 0.0223 | 1.67E-12 | Yes | No |
| rs262986 | 3 | 183435713 | A | G | 0.4693 | -0.0997 | 0.0175 | 1.15E-08 | Yes | No |
| rs12374077 | 3 | 185317674 | C | G | 0.344 | 0.1748 | 0.0182 | 8.05E-22 | Yes | No |
| rs1706003 | 3 | 194299967 | T | G | 0.4675 | 0.1243 | 0.0181 | 5.77E-12 | No | No |
| rs6777317 | 3 | 197070959 | A | G | 0.2899 | 0.1249 | 0.0195 | 1.51E-10 | No | No |
| rs1250129 | 4 | 1254930 | A | G | 0.1152 | 0.0054 | 0.0272 | 0.8428 | Yes | No |
| rs55829085 | 4 | 2165493 | A | C | 0.9547 | -0.2823 | 0.0425 | 3.03E-11 | Yes | No |
| rs231708 | 4 | 2694773 | C | G | 0.6918 | -0.0652 | 0.0187 | 0.0004932 | Yes | No |
| rs2498323 | 4 | 3451109 | A | G | 0.0982 | 0.0194 | 0.0296 | 0.5113 | Yes | No |
| rs3822239 | 4 | 10095539 | A | T | 0.6966 | 0.0247 | 0.0188 | 0.1897 | Yes | No |
| rs13122790 | 4 | 15356795 | A | G | 0.7315 | -0.0106 | 0.0196 | 0.5874 | Yes | No |
| rs11730129 | 4 | 16032948 | T | C | 0.2184 | 0.0777 | 0.021 | 0.0002225 | Yes | No |
| rs2610990 | 4 | 18008232 | A | G | 0.2642 | -0.1296 | 0.0197 | 4.84E-11 | Yes | No |
| rs28667801 | 4 | 26785356 | A | T | 0.593 | -0.1622 | 0.018 | 1.90E-19 | No | No |
| rs1878825 | 4 | 36091370 | C | G | 0.6424 | -0.1072 | 0.0183 | 4.62E-09 | Yes | Yes |
| rs2291435 | 4 | 38387395 | T | C | 0.534 | -0.1275 | 0.0174 | 2.12E-13 | Yes | No |
| rs12511987 | 4 | 46595623 | T | G | 0.8228 | -0.122 | 0.0229 | 9.83E-08 | Yes | No |
| rs871606 | 4 | 54799245 | T | C | 0.895 | -0.1844 | 0.0284 | 8.56E-11 | Yes | No |
| rs1718845 | 4 | 57943153 | A | G | 0.2982 | -0.0982 | 0.019 | 2.33E-07 | Yes | No |
| rs6551716 | 4 | 63575696 | A | T | 0.856 | 0.1478 | 0.0251 | 3.91E-09 | Yes | Yes |
| rs10008637 | 4 | 77414144 | T | C | 0.5405 | 0.0801 | 0.0173 | 3.79E-06 | Yes | No |
| rs1458038 | 4 | 81164723 | T | C | 0.294 | 0.491 | 0.0191 | 5.46E-146 | Yes | No |
| rs6823199 | 4 | 83925895 | T | C | 0.743 | 0.0521 | 0.0199 | 0.008919 | Yes | No |
| rs2014912 | 4 | 86715670 | T | C | 0.1531 | 0.1488 | 0.0241 | 6.62E-10 | Yes | No |
| rs13149209 | 4 | 89750668 | T | C | 0.7781 | 0.1116 | 0.0211 | 1.19E-07 | Yes | No |
| rs7694000 | 4 | 95324968 | A | T | 0.5387 | -0.0965 | 0.0175 | 3.47E-08 | No | No |
| rs1347345 | 4 | 95938386 | A | G | 0.6175 | -0.0915 | 0.0179 | 3.19E-07 | Yes | No |
| rs17248480 | 4 | 102435265 | A | G | 0.0284 | -0.5177 | 0.0552 | 6.61E-21 | Yes | Yes |
| rs13107325 | 4 | 103188709 | T | C | 0.0742 | -0.6747 | 0.0339 | 3.72E-88 | Yes | No |
| rs223361 | 4 | 103769304 | T | C | 0.6659 | 0.1698 | 0.0184 | 2.70E-20 | Yes | Yes |
| rs4699165 | 4 | 106109381 | A | G | 0.3572 | -0.0313 | 0.0181 | 0.08352 | Yes | No |
| rs13112725 | 4 | 106911742 | C | G | 0.763 | 0.2283 | 0.0205 | 1.01E-28 | Yes | No |
| rs7694643 | 4 | 109017528 | A | G | 0.6443 | -0.1318 | 0.0181 | 3.07E-13 | Yes | No |
| rs6825911 | 4 | 111381638 | T | C | 0.7924 | -0.202 | 0.0215 | 6.94E-21 | Yes | Yes |
| rs4834735 | 4 | 119958809 | T | C | 0.1367 | 0.1511 | 0.0254 | 2.64E-09 | Yes | Yes |
| rs66887589 | 4 | 120509279 | T | C | 0.5221 | -0.161 | 0.0174 | 1.83E-20 | Yes | No |
| rs3097937 | 4 | 124794644 | A | T | 0.8022 | 0.1138 | 0.0218 | 1.82E-07 | Yes | No |
| rs7439567 | 4 | 138464842 | T | C | 0.4105 | 0.1364 | 0.0178 | 1.62E-14 | Yes | No |
| rs72719160 | 4 | 144051276 | A | T | 0.6827 | -0.127 | 0.0186 | 8.34E-12 | Yes | No |
| rs4292285 | 4 | 145271954 | A | T | 0.4021 | -0.1073 | 0.0177 | 1.28E-09 | Yes | No |
| rs4835266 | 4 | 146821725 | T | C | 0.5209 | 0.0561 | 0.0177 | 0.001553 | Yes | No |
| rs10305838 | 4 | 148400256 | T | C | 0.8594 | -0.002 | 0.0248 | 0.9357 | Yes | No |
| rs6823767 | 4 | 151295085 | T | C | 0.7221 | -0.1021 | 0.0195 | 1.74E-07 | Yes | No |
| rs13139571 | 4 | 156645513 | A | C | 0.2366 | -0.2408 | 0.0203 | 2.29E-32 | Yes | Yes |
| rs17035181 | 4 | 157678511 | T | G | 0.8553 | 0.153 | 0.0246 | 5.21E-10 | Yes | No |
| rs869396 | 4 | 169688000 | A | C | 0.4653 | 0.0136 | 0.0175 | 0.4377 | Yes | No |
| rs4957026 | 5 | 361148 | A | G | 0.3392 | 0.0947 | 0.0185 | 3.08E-07 | Yes | No |
| rs10069690 | 5 | 1279790 | T | C | 0.2581 | 0.1615 | 0.021 | 1.42E-14 | Yes | No |
| rs954767 | 5 | 3706050 | A | C | 0.7405 | -0.1496 | 0.0198 | 4.24E-14 | Yes | Yes |
| rs1173771 | 5 | 32815028 | A | G | 0.3985 | -0.3021 | 0.0177 | 1.16E-65 | Yes | No |
| rs74774746 | 5 | 33411769 | C | G | 0.2611 | -0.0962 | 0.0201 | 1.62E-06 | Yes | No |
| rs7710854 | 5 | 43824677 | A | G | 0.8836 | 0.0359 | 0.0274 | 0.19 | Yes | No |
| rs6867399 | 5 | 52135543 | A | C | 0.2685 | 0.0043 | 0.0204 | 0.8317 | Yes | No |
| rs1694068 | 5 | 53283630 | A | T | 0.6143 | 0.1287 | 0.0178 | 4.98E-13 | Yes | No |
| rs13179413 | 5 | 55868097 | T | C | 0.2824 | 0.125 | 0.0198 | 3.04E-10 | Yes | No |
| rs12515541 | 5 | 57095011 | T | G | 0.6072 | 0.1156 | 0.0177 | 6.23E-11 | Yes | No |
| rs1848510 | 5 | 57754005 | A | G | 0.3623 | 0.1256 | 0.0181 | 4.10E-12 | Yes | No |
| rs10062049 | 5 | 61553881 | T | C | 0.1359 | 0.2208 | 0.0255 | 4.50E-18 | Yes | Yes |
| rs6875372 | 5 | 64079015 | A | T | 0.5154 | 0.0652 | 0.0174 | 0.000175 | No | No |
| rs3121685 | 5 | 65662133 | T | C | 0.4843 | -0.0678 | 0.0173 | 9.18E-05 | Yes | No |
| rs4286632 | 5 | 66291370 | A | G | 0.7294 | 0.118 | 0.0196 | 1.88E-09 | Yes | No |
| rs246973 | 5 | 68007803 | T | C | 0.2887 | 0.1167 | 0.0192 | 1.28E-09 | Yes | No |
| rs72761109 | 5 | 71506529 | T | C | 0.3036 | -0.0039 | 0.0188 | 0.8369 | Yes | No |
| rs4443403 | 5 | 72654304 | T | C | 0.903 | -0.0597 | 0.0294 | 0.04221 | Yes | No |
| rs10078021 | 5 | 75038431 | T | G | 0.6277 | -0.1534 | 0.0182 | 3.15E-17 | Yes | No |
| rs10057188 | 5 | 77837789 | A | G | 0.4589 | -0.0492 | 0.0176 | 0.005172 | Yes | No |
| rs10059921 | 5 | 87514515 | T | G | 0.0822 | -0.2184 | 0.0336 | 8.43E-11 | Yes | No |
| rs62380354 | 5 | 89484911 | A | C | 0.8904 | 0.1825 | 0.0291 | 3.68E-10 | Yes | No |
| rs709668 | 5 | 96174186 | A | G | 0.2001 | -0.1522 | 0.0216 | 2.02E-12 | Yes | No |
| rs1871190 | 5 | 97953719 | T | G | 0.3344 | 0.1078 | 0.0186 | 6.63E-09 | Yes | No |
| rs286809 | 5 | 107458637 | A | G | 0.1721 | -0.0485 | 0.0229 | 0.03438 | Yes | No |
| rs79409628 | 5 | 108113740 | T | G | 0.0843 | 0.0467 | 0.0312 | 0.1341 | Yes | No |
| rs10077885 | 5 | 114390121 | A | C | 0.509 | -0.1439 | 0.0177 | 3.89E-16 | Yes | No |
| rs1432457 | 5 | 119781569 | A | G | 0.7146 | -0.0321 | 0.0192 | 0.09399 | Yes | No |
| rs9885577 | 5 | 121194226 | T | C | 0.3703 | 0.0698 | 0.0184 | 0.0001482 | Yes | No |
| rs13359291 | 5 | 122476457 | A | G | 0.1568 | 0.2125 | 0.0239 | 5.28E-19 | Yes | No |
| rs6891344 | 5 | 123136656 | A | G | 0.8186 | 0.2064 | 0.0227 | 8.98E-20 | Yes | No |
| rs62373688 | 5 | 127352807 | A | T | 0.1315 | 0.1089 | 0.026 | 2.79E-05 | Yes | No |
| rs6595838 | 5 | 127868199 | A | G | 0.2994 | 0.1688 | 0.0189 | 4.69E-19 | Yes | No |
| rs12521868 | 5 | 131784393 | T | G | 0.423 | -0.1403 | 0.0176 | 1.83E-15 | Yes | Yes |
| rs55747751 | 5 | 132397351 | A | G | 0.081 | -0.2239 | 0.0331 | 1.39E-11 | Yes | No |
| rs702395 | 5 | 140086677 | T | C | 0.4366 | 0.0826 | 0.0175 | 2.39E-06 | Yes | No |
| rs1650911 | 5 | 141740620 | C | G | 0.7654 | 0.1325 | 0.0211 | 3.19E-10 | Yes | No |
| rs2400509 | 5 | 147696018 | A | G | 0.2583 | -0.016 | 0.0197 | 0.4166 | Yes | No |
| rs9687065 | 5 | 148391140 | A | G | 0.8102 | 0.2199 | 0.0222 | 4.85E-23 | Yes | Yes |
| rs157678 | 5 | 156145654 | A | T | 0.6682 | -0.0415 | 0.0188 | 0.02781 | Yes | No |
| rs11953630 | 5 | 157845402 | T | C | 0.3656 | -0.2367 | 0.0183 | 3.72E-38 | Yes | No |
| rs62385385 | 5 | 158367249 | A | T | 0.3829 | 0.0588 | 0.0178 | 0.0009331 | Yes | No |
| rs114503346 | 5 | 172192350 | T | C | 0.0461 | -0.2678 | 0.0426 | 3.10E-10 | Yes | No |
| rs72812846 | 5 | 173377636 | A | T | 0.2785 | -0.2053 | 0.0197 | 2.15E-25 | Yes | No |
| rs28362590 | 5 | 176731452 | T | G | 0.7537 | 0.1242 | 0.0203 | 8.70E-10 | Yes | No |
| rs12153395 | 5 | 179411477 | A | G | 0.1148 | -0.1161 | 0.0278 | 3.03E-05 | Yes | No |
| rs2745599 | 6 | 1613686 | A | G | 0.5521 | 0.115 | 0.0181 | 2.03E-10 | Yes | No |
| rs1334576 | 6 | 7211818 | A | G | 0.4201 | 0.0421 | 0.0176 | 0.0164 | Yes | No |
| rs9392172 | 6 | 7723962 | C | G | 0.5346 | -0.0099 | 0.0173 | 0.5662 | No | No |
| rs114275780 | 6 | 8224648 | A | T | 0.0591 | 0.0425 | 0.0387 | 0.2723 | Yes | No |
| rs1630736 | 6 | 12295987 | T | C | 0.465 | -0.04 | 0.0177 | 0.02397 | No | No |
| rs9349379 | 6 | 12903957 | A | G | 0.5926 | -0.0053 | 0.0179 | 0.765 | Yes | No |
| rs12216497 | 6 | 19028623 | T | C | 0.5626 | 0.0198 | 0.0174 | 0.2555 | Yes | No |
| rs9368222 | 6 | 20686996 | A | C | 0.268 | 0.1179 | 0.0195 | 1.41E-09 | Yes | No |
| rs6911827 | 6 | 22130601 | T | C | 0.4575 | 0.136 | 0.0176 | 9.68E-15 | Yes | No |
| rs1799945 | 6 | 26091179 | C | G | 0.8503 | -0.3888 | 0.0244 | 3.88E-57 | Yes | No |
| rs409558 | 6 | 31708147 | T | C | 0.8505 | 0.042 | 0.0248 | 0.0906 | Yes | No |
| rs115245297 | 6 | 34244132 | T | C | 0.9551 | -0.3116 | 0.0437 | 1.06E-12 | Yes | No |
| rs3176336 | 6 | 36648816 | A | T | 0.6109 | -0.0554 | 0.018 | 0.002097 | Yes | No |
| rs4714224 | 6 | 39186743 | C | G | 0.2776 | -0.1358 | 0.0196 | 3.78E-12 | Yes | Yes |
| rs1563788 | 6 | 43308363 | T | C | 0.2861 | 0.1299 | 0.0191 | 9.88E-12 | Yes | No |
| rs9472135 | 6 | 43809802 | T | C | 0.6981 | 0.1548 | 0.019 | 4.29E-16 | Yes | No |
| rs78648104 | 6 | 50683009 | T | C | 0.9084 | -0.2413 | 0.0311 | 8.14E-15 | Yes | No |
| rs13205180 | 6 | 51832494 | T | C | 0.4886 | 0.1721 | 0.0174 | 4.38E-23 | Yes | Yes |
| rs631441 | 6 | 53994626 | T | G | 0.6938 | -0.019 | 0.0187 | 0.3111 | Yes | No |
| rs1925153 | 6 | 56102780 | T | C | 0.4465 | 0.0685 | 0.0179 | 0.0001278 | Yes | No |
| rs504691 | 6 | 72206620 | A | C | 0.4002 | -0.1177 | 0.0177 | 3.14E-11 | Yes | Yes |
| rs12195276 | 6 | 73657714 | T | C | 0.7245 | 0.0179 | 0.0195 | 0.3571 | Yes | No |
| rs10943605 | 6 | 79655477 | A | G | 0.4882 | 0.1723 | 0.0173 | 2.93E-23 | Yes | Yes |
| rs7753695 | 6 | 80818531 | T | C | 0.4386 | 0.1031 | 0.0176 | 4.77E-09 | Yes | No |
| rs9449350 | 6 | 82281417 | T | C | 0.6802 | -0.0816 | 0.0185 | 1.04E-05 | Yes | No |
| rs60255247 | 6 | 85283253 | A | C | 0.8876 | -0.0185 | 0.0279 | 0.5074 | Yes | No |
| rs35410524 | 6 | 96885405 | T | C | 0.1878 | 0.1302 | 0.0222 | 4.57E-09 | Yes | No |
| rs72613227 | 6 | 106320771 | A | T | 0.8731 | -0.1884 | 0.0285 | 3.87E-11 | Yes | No |
| rs9486916 | 6 | 109013930 | T | C | 0.1976 | 0.0814 | 0.0221 | 0.0002307 | Yes | No |
| rs3822857 | 6 | 116313931 | C | G | 0.3684 | -0.1238 | 0.018 | 6.09E-12 | Yes | No |
| rs2693560 | 6 | 117523671 | A | G | 0.3669 | -0.1501 | 0.0181 | 1.09E-16 | Yes | No |
| rs2498586 | 6 | 118026126 | T | C | 0.165 | -0.1054 | 0.0236 | 8.22E-06 | Yes | No |
| rs9372498 | 6 | 118572486 | A | T | 0.0809 | 0.2731 | 0.0319 | 1.13E-17 | Yes | No |
| rs9401090 | 6 | 119113317 | T | C | 0.745 | 0.11 | 0.02 | 3.76E-08 | Yes | Yes |
| rs11154027 | 6 | 121781390 | T | C | 0.4617 | -0.0662 | 0.0176 | 0.0001606 | Yes | No |
| rs6925750 | 6 | 122287990 | T | C | 0.8859 | -0.0824 | 0.0272 | 0.002505 | Yes | No |
| rs10782230 | 6 | 126228512 | A | G | 0.4838 | 0.0914 | 0.0173 | 1.25E-07 | Yes | No |
| rs13209747 | 6 | 127115454 | T | C | 0.4409 | 0.3017 | 0.0175 | 6.23E-67 | Yes | Yes |
| rs9885632 | 6 | 131311909 | T | C | 0.7304 | 0.0866 | 0.0195 | 9.03E-06 | Yes | No |
| rs4896104 | 6 | 135119089 | T | C | 0.5588 | 0.0511 | 0.0177 | 0.003842 | Yes | No |
| rs668459 | 6 | 139835689 | T | C | 0.5864 | -0.1132 | 0.0175 | 1.01E-10 | Yes | No |
| rs7763294 | 6 | 140383733 | T | G | 0.3163 | -0.0511 | 0.0186 | 0.005943 | Yes | No |
| rs6941056 | 6 | 143591821 | C | G | 0.5584 | -0.0527 | 0.0175 | 0.002631 | No | No |
| rs7765526 | 6 | 147713764 | A | G | 0.4629 | 0.1145 | 0.0176 | 8.02E-11 | Yes | No |
| rs17080102 | 6 | 151004770 | C | G | 0.0696 | -0.4853 | 0.034 | 3.91E-46 | Yes | Yes |
| rs9479200 | 6 | 152398505 | A | G | 0.8861 | 0.1943 | 0.0275 | 1.45E-12 | Yes | No |
| rs9479509 | 6 | 153427265 | A | G | 0.2954 | -0.1152 | 0.019 | 1.21E-09 | Yes | No |
| rs598682 | 6 | 154418759 | A | G | 0.2512 | -0.1073 | 0.0199 | 7.20E-08 | Yes | Yes |
| rs449789 | 6 | 159699125 | C | G | 0.1392 | -0.0435 | 0.0251 | 0.08238 | Yes | No |
| rs555754 | 6 | 160769423 | A | G | 0.4742 | -0.0167 | 0.0173 | 0.3347 | Yes | No |
| rs9456648 | 6 | 161712235 | T | C | 0.3233 | -0.1166 | 0.0185 | 2.76E-10 | Yes | Yes |
| rs73030266 | 6 | 166179459 | A | T | 0.9329 | 0.304 | 0.0354 | 8.59E-18 | Yes | No |
| rs4342401 | 6 | 169016615 | A | T | 0.5414 | -0.0388 | 0.0174 | 0.02609 | No | No |
| rs1322639 | 6 | 169587103 | A | G | 0.7766 | -0.1584 | 0.0209 | 3.87E-14 | Yes | No |
| rs73033340 | 7 | 1195692 | A | G | 0.9638 | 0.5312 | 0.0525 | 5.06E-24 | No | No |
| rs6959688 | 7 | 1966831 | A | G | 0.5985 | -0.1269 | 0.0178 | 1.02E-12 | Yes | No |
| rs2969070 | 7 | 2512545 | A | G | 0.6309 | -0.1791 | 0.0179 | 1.76E-23 | Yes | Yes |
| rs73049928 | 7 | 4669949 | A | G | 0.8062 | -0.1194 | 0.0224 | 1.05E-07 | Yes | No |
| rs1468520 | 7 | 7290732 | A | G | 0.8329 | -0.1638 | 0.0234 | 2.49E-12 | Yes | Yes |
| rs13240040 | 7 | 14375977 | A | G | 0.6836 | 0.1186 | 0.019 | 3.98E-10 | Yes | Yes |
| rs2107595 | 7 | 19049388 | A | G | 0.1586 | -0.0382 | 0.0238 | 0.1082 | Yes | No |
| rs4507656 | 7 | 22156538 | C | G | 0.6934 | -0.1487 | 0.0199 | 8.69E-14 | No | No |
| rs2069833 | 7 | 22767664 | T | C | 0.5747 | -0.0163 | 0.0175 | 0.3526 | Yes | No |
| rs12979 | 7 | 24738164 | C | G | 0.869 | 0.0913 | 0.0257 | 0.0003883 | Yes | No |
| rs1055144 | 7 | 25871109 | T | C | 0.1929 | -0.042 | 0.0219 | 0.05541 | Yes | No |
| rs6969780 | 7 | 27159136 | C | G | 0.0917 | 0.163 | 0.0301 | 6.36E-08 | Yes | No |
| rs7777128 | 7 | 27337113 | C | G | 0.0798 | 0.2591 | 0.032 | 5.46E-16 | Yes | No |
| rs10274928 | 7 | 28142088 | A | G | 0.484 | 0.0694 | 0.0175 | 7.63E-05 | Yes | No |
| rs917275 | 7 | 28658522 | A | G | 0.6095 | 0.0096 | 0.0179 | 0.5919 | Yes | No |
| rs10233127 | 7 | 30933453 | A | T | 0.1075 | 0.1498 | 0.029 | 2.46E-07 | Yes | No |
| rs342989 | 7 | 35467896 | A | G | 0.229 | 0.1631 | 0.0207 | 3.05E-15 | Yes | Yes |
| rs2052263 | 7 | 36225818 | A | G | 0.1183 | 0.0551 | 0.0275 | 0.04555 | Yes | No |
| rs76206723 | 7 | 40447971 | A | G | 0.1071 | -0.0531 | 0.0282 | 0.05956 | Yes | No |
| rs12701929 | 7 | 41722494 | T | C | 0.2504 | 1.00E-04 | 0.02 | 0.9946 | Yes | No |
| rs1004558 | 7 | 44240407 | T | C | 0.1783 | -0.0168 | 0.0228 | 0.4619 | Yes | No |
| rs73105827 | 7 | 45036785 | T | G | 0.0909 | -0.1878 | 0.0313 | 2.07E-09 | Yes | No |
| rs11977526 | 7 | 46008110 | A | G | 0.4001 | 0.1093 | 0.0179 | 9.96E-10 | Yes | No |
| rs71543920 | 7 | 46554358 | T | G | 0.9395 | 0.0179 | 0.0368 | 0.6267 | Yes | No |
| rs12668436 | 7 | 47548893 | T | C | 0.7537 | -0.0419 | 0.0201 | 0.0369 | Yes | No |
| rs17454517 | 7 | 50915776 | A | G | 0.4936 | 0.1216 | 0.0174 | 2.65E-12 | Yes | No |
| rs6593297 | 7 | 56122058 | A | T | 0.3015 | 0.0492 | 0.0193 | 0.01084 | Yes | No |
| rs2222544 | 7 | 69769369 | T | C | 0.7366 | -0.1001 | 0.0196 | 3.37E-07 | Yes | No |
| rs1091811 | 7 | 73491212 | A | G | 0.169 | 0.0133 | 0.0232 | 0.5674 | Yes | No |
| rs34324971 | 7 | 74107374 | A | G | 0.1851 | 0.1149 | 0.0232 | 7.55E-07 | Yes | No |
| rs6963105 | 7 | 75097488 | A | G | 0.4385 | -0.0768 | 0.0184 | 2.97E-05 | Yes | No |
| rs848445 | 7 | 77572461 | T | C | 0.2854 | -0.0745 | 0.0194 | 0.000124 | Yes | No |
| rs11770630 | 7 | 89805241 | T | C | 0.5355 | -0.1187 | 0.0173 | 7.05E-12 | Yes | No |
| rs10245696 | 7 | 90449362 | A | C | 0.4007 | 0.0176 | 0.0176 | 0.3188 | Yes | No |
| rs2282978 | 7 | 92264410 | T | C | 0.6691 | -0.0064 | 0.0184 | 0.7302 | Yes | No |
| rs1947228 | 7 | 96461649 | T | C | 0.418 | -0.1448 | 0.0177 | 2.59E-16 | Yes | Yes |
| rs12705090 | 7 | 100467700 | T | C | 0.1896 | 0.1596 | 0.0221 | 5.20E-13 | Yes | No |
| rs17477177 | 7 | 106411858 | T | C | 0.7962 | 0.017 | 0.0215 | 0.4301 | Yes | No |
| rs115172170 | 7 | 107019947 | T | C | 0.944 | -0.0479 | 0.0379 | 0.207 | Yes | No |
| rs1997571 | 7 | 116198621 | A | G | 0.5914 | -0.0037 | 0.0176 | 0.8314 | Yes | No |
| rs4728142 | 7 | 128573967 | A | G | 0.4446 | -0.0734 | 0.0175 | 2.70E-05 | Yes | No |
| rs11556924 | 7 | 129663496 | T | C | 0.3827 | -0.181 | 0.0181 | 1.83E-23 | Yes | Yes |
| rs34072724 | 7 | 130432469 | A | G | 0.4903 | -0.1313 | 0.0174 | 4.38E-14 | Yes | No |
| rs13238550 | 7 | 131059056 | A | G | 0.3979 | 0.1357 | 0.0177 | 1.62E-14 | Yes | No |
| rs1722886 | 7 | 134215259 | A | T | 0.5667 | 0.1215 | 0.0175 | 3.75E-12 | No | No |
| rs10267979 | 7 | 136618188 | A | T | 0.3243 | 0.0107 | 0.0185 | 0.5629 | Yes | No |
| rs7810028 | 7 | 139461616 | C | G | 0.8064 | 0.1407 | 0.0219 | 1.32E-10 | Yes | No |
| rs12703989 | 7 | 140238048 | A | G | 0.4999 | 0.103 | 0.0175 | 4.35E-09 | Yes | No |
| rs73727605 | 7 | 149474622 | A | G | 0.0662 | 0.0722 | 0.0357 | 0.04328 | Yes | No |
| rs11771693 | 7 | 150050111 | A | G | 0.6754 | 0.0785 | 0.0187 | 2.59E-05 | Yes | No |
| rs3918226 | 7 | 150690176 | T | C | 0.0813 | 0.6117 | 0.0329 | 5.31E-77 | Yes | No |
| rs891511 | 7 | 150704843 | A | G | 0.3322 | -0.2634 | 0.0191 | 2.11E-43 | Yes | No |
| rs10224002 | 7 | 151415041 | A | G | 0.7139 | -0.203 | 0.0194 | 9.99E-26 | Yes | No |
| rs1870735 | 7 | 155744303 | C | G | 0.4528 | 0.1069 | 0.0178 | 1.94E-09 | No | No |
| rs9638084 | 7 | 156311745 | A | G | 0.3978 | 0.1154 | 0.0178 | 8.51E-11 | Yes | No |
| rs4875958 | 8 | 1721090 | A | G | 0.7123 | 0.1056 | 0.0193 | 4.39E-08 | Yes | No |
| rs2922895 | 8 | 6379932 | C | G | 0.4294 | 0.1317 | 0.0175 | 5.86E-14 | No | No |
| rs61040371 | 8 | 8503700 | T | C | 0.6288 | 0.0902 | 0.018 | 5.25E-07 | Yes | No |
| rs62491354 | 8 | 9730663 | A | G | 0.1337 | 0.1281 | 0.0255 | 4.86E-07 | Yes | No |
| rs1986971 | 8 | 10268736 | A | G | 0.7062 | 0.1047 | 0.0192 | 4.60E-08 | Yes | No |
| rs2898290 | 8 | 11433909 | T | C | 0.4754 | 0.1393 | 0.0175 | 1.46E-15 | Yes | No |
| rs75902664 | 8 | 17427186 | A | G | 0.9739 | -0.3546 | 0.0569 | 4.65E-10 | Yes | No |
| rs1047030 | 8 | 22428708 | A | G | 0.8037 | 0.1264 | 0.0233 | 5.69E-08 | Yes | No |
| rs62503324 | 8 | 23400615 | T | C | 0.2397 | 0.2033 | 0.0204 | 2.11E-23 | Yes | No |
| rs6557876 | 8 | 25900675 | T | C | 0.2503 | -0.1973 | 0.02 | 7.07E-23 | Yes | No |
| rs17321041 | 8 | 26445194 | T | C | 0.0633 | 0.2313 | 0.0363 | 1.78E-10 | Yes | Yes |
| rs2979470 | 8 | 30288272 | T | C | 0.4903 | 0.1016 | 0.0173 | 4.61E-09 | Yes | No |
| rs11991469 | 8 | 32413280 | C | G | 0.5397 | 0.012 | 0.0174 | 0.4896 | No | No |
| rs7845722 | 8 | 33309993 | A | G | 0.6012 | 0.0022 | 0.0177 | 0.9019 | Yes | No |
| rs1906672 | 8 | 38130025 | A | G | 0.2324 | 0.1402 | 0.0205 | 8.48E-12 | Yes | No |
| rs2978456 | 8 | 42324765 | T | C | 0.552 | -0.0095 | 0.0179 | 0.5951 | No | No |
| rs4873492 | 8 | 51947549 | T | C | 0.1725 | 0.1401 | 0.0231 | 1.28E-09 | Yes | No |
| rs6996733 | 8 | 60535824 | T | C | 0.8495 | 0.0935 | 0.0245 | 0.0001337 | Yes | No |
| rs2354862 | 8 | 64501744 | A | C | 0.6402 | 0.1186 | 0.0182 | 6.45E-11 | Yes | No |
| rs13253358 | 8 | 68920135 | T | C | 0.2981 | 0.1061 | 0.0189 | 2.00E-08 | Yes | No |
| rs1350100 | 8 | 76054904 | A | G | 0.4458 | -0.0589 | 0.0175 | 0.0007837 | Yes | No |
| rs1449544 | 8 | 76591880 | A | C | 0.5438 | 0.0131 | 0.0174 | 0.4504 | Yes | No |
| rs72688070 | 8 | 81393697 | T | C | 0.1682 | -0.1363 | 0.0233 | 5.14E-09 | Yes | No |
| rs56345595 | 8 | 82814156 | A | G | 0.5848 | 0.1329 | 0.0177 | 5.20E-14 | Yes | No |
| rs2142141 | 8 | 90940205 | C | G | 0.5331 | -0.1054 | 0.0179 | 3.68E-09 | No | No |
| rs7009170 | 8 | 92149429 | T | C | 0.3186 | -0.0143 | 0.0186 | 0.4401 | Yes | No |
| rs62526122 | 8 | 92769569 | A | G | 0.2917 | 0.1 | 0.0197 | 4.02E-07 | Yes | No |
| rs4582532 | 8 | 95969257 | A | G | 0.5064 | -0.1061 | 0.0174 | 9.88E-10 | Yes | No |
| rs2978098 | 8 | 101676675 | A | C | 0.5465 | 0.1548 | 0.0176 | 1.33E-18 | Yes | No |
| rs142449193 | 8 | 102750597 | T | C | 0.046 | -0.2573 | 0.0426 | 1.51E-09 | Yes | No |
| rs2513877 | 8 | 103883630 | A | G | 0.1917 | -0.1294 | 0.022 | 4.22E-09 | Yes | No |
| rs35783704 | 8 | 105966258 | A | G | 0.104 | -0.2132 | 0.0291 | 2.28E-13 | Yes | No |
| rs28499085 | 8 | 110107161 | A | G | 0.725 | 0.0516 | 0.0195 | 0.00801 | Yes | No |
| rs2205260 | 8 | 116959837 | A | C | 0.1677 | -0.1138 | 0.0232 | 9.67E-07 | Yes | No |
| rs2071518 | 8 | 120435812 | T | C | 0.2619 | -0.1869 | 0.0197 | 1.92E-21 | Yes | No |
| rs62523863 | 8 | 126520544 | A | G | 0.2191 | 0.1485 | 0.0211 | 2.12E-12 | Yes | No |
| rs4598218 | 8 | 129483956 | T | C | 0.6163 | 0.0948 | 0.018 | 1.31E-07 | Yes | No |
| rs894344 | 8 | 135612745 | A | G | 0.5926 | -0.1267 | 0.0176 | 5.79E-13 | Yes | Yes |
| rs4454254 | 8 | 141060027 | A | G | 0.6308 | 0.0249 | 0.0179 | 0.1638 | Yes | No |
| rs10087782 | 8 | 141858620 | T | C | 0.4463 | 0.132 | 0.0174 | 2.99E-14 | Yes | Yes |
| rs34591516 | 8 | 142367087 | T | C | 0.049 | 0.3121 | 0.0409 | 2.26E-14 | Yes | No |
| rs4129585 | 8 | 143312933 | A | C | 0.4414 | 0.0927 | 0.0175 | 1.20E-07 | Yes | No |
| rs62524579 | 8 | 144060955 | A | G | 0.5335 | -0.1656 | 0.0182 | 1.08E-19 | Yes | No |
| rs56233017 | 8 | 144981488 | A | G | 0.0457 | -0.2743 | 0.044 | 4.37E-10 | Yes | No |
| rs520015 | 9 | 211762 | C | G | 0.5129 | 0.0985 | 0.0174 | 1.42E-08 | No | No |
| rs60191654 | 9 | 753648 | A | G | 0.8114 | -0.1191 | 0.0222 | 7.92E-08 | Yes | No |
| rs12216886 | 9 | 2493751 | T | G | 0.8077 | 0.1292 | 0.0221 | 4.76E-09 | Yes | Yes |
| rs28558845 | 9 | 4334791 | C | G | 0.1548 | -0.1237 | 0.0243 | 3.74E-07 | Yes | No |
| rs1332813 | 9 | 9350706 | T | C | 0.3514 | 0.1139 | 0.0181 | 3.34E-10 | Yes | No |
| rs35287509 | 9 | 10594635 | T | C | 0.6622 | -0.1082 | 0.0184 | 4.17E-09 | Yes | No |
| rs11789875 | 9 | 16872323 | A | G | 0.1515 | 0.0112 | 0.0243 | 0.6433 | Yes | No |
| rs4977492 | 9 | 19057551 | T | C | 0.6621 | 0.0058 | 0.0184 | 0.7525 | Yes | No |
| rs4364717 | 9 | 21801530 | A | G | 0.5475 | -0.1006 | 0.0174 | 7.56E-09 | Yes | Yes |
| rs9886665 | 9 | 22942770 | T | C | 0.2668 | 0.1003 | 0.0198 | 4.21E-07 | Yes | No |
| rs4553000 | 9 | 34223553 | T | C | 0.514 | -0.0594 | 0.0173 | 0.000591 | Yes | No |
| rs76452347 | 9 | 35906471 | T | C | 0.2053 | -0.2246 | 0.0229 | 9.37E-23 | No | No |
| rs11139596 | 9 | 85128518 | T | G | 0.7757 | 0.0333 | 0.0213 | 0.1182 | Yes | No |
| rs11141731 | 9 | 89888472 | T | C | 0.228 | -0.1258 | 0.0207 | 1.31E-09 | Yes | No |
| rs7045409 | 9 | 95201540 | A | T | 0.3669 | -0.0637 | 0.0181 | 0.0004253 | Yes | No |
| rs10988442 | 9 | 101739709 | A | G | 0.6195 | -0.0304 | 0.018 | 0.0909 | Yes | No |
| rs7020564 | 9 | 109670016 | A | T | 0.7009 | -0.1111 | 0.0192 | 6.73E-09 | Yes | Yes |
| rs7043304 | 9 | 112358150 | T | C | 0.8548 | 0.176 | 0.0249 | 1.63E-12 | Yes | Yes |
| rs111245230 | 9 | 113169775 | T | C | 0.9654 | -0.37 | 0.0481 | 1.45E-14 | Yes | No |
| rs13290326 | 9 | 116696625 | T | C | 0.5019 | -0.0641 | 0.0173 | 0.0002183 | Yes | No |
| rs10982910 | 9 | 118534500 | T | G | 0.9039 | 0.0718 | 0.0298 | 0.01596 | Yes | No |
| rs1861881 | 9 | 119312256 | T | G | 0.3159 | 0.115 | 0.0186 | 6.49E-10 | Yes | Yes |
| rs1953126 | 9 | 123640500 | T | C | 0.3534 | -0.0088 | 0.0182 | 0.6287 | Yes | No |
| rs10818775 | 9 | 125755571 | T | C | 0.1213 | -0.0296 | 0.0267 | 0.2679 | Yes | No |
| rs7861040 | 9 | 127044135 | C | G | 0.3674 | -0.0776 | 0.0179 | 1.50E-05 | Yes | No |
| rs72765298 | 9 | 127900996 | T | C | 0.8746 | -0.0228 | 0.0266 | 0.3904 | Yes | No |
| rs7023828 | 9 | 128498594 | T | C | 0.418 | -0.1259 | 0.0176 | 9.79E-13 | Yes | No |
| rs1891730 | 9 | 130309028 | T | C | 0.6163 | -0.0839 | 0.0181 | 3.61E-06 | Yes | No |
| rs7869756 | 9 | 131210410 | A | G | 0.8106 | 0.091 | 0.0225 | 5.29E-05 | Yes | No |
| rs184457 | 9 | 131940019 | A | G | 0.3087 | -0.0798 | 0.0188 | 2.18E-05 | Yes | No |
| rs687621 | 9 | 136137065 | A | G | 0.6733 | 0.1834 | 0.0186 | 5.30E-23 | Yes | No |
| rs6271 | 9 | 136522274 | T | C | 0.0737 | -0.4313 | 0.0352 | 1.72E-34 | No | No |
| rs11145807 | 9 | 139520789 | A | G | 0.4058 | 0.155 | 0.0184 | 4.10E-17 | No | No |
| rs56352451 | 10 | 5804865 | T | C | 0.1323 | 0.0981 | 0.0255 | 0.0001168 | Yes | No |
| rs36006409 | 10 | 10876943 | T | G | 0.7932 | -0.0303 | 0.0215 | 0.1588 | Yes | No |
| rs10906391 | 10 | 13523937 | T | C | 0.3171 | 0.1286 | 0.0188 | 7.57E-12 | Yes | No |
| rs4373814 | 10 | 18419972 | C | G | 0.4269 | 0.1981 | 0.0175 | 1.28E-29 | No | No |
| rs1813353 | 10 | 18707448 | T | C | 0.663 | 0.3013 | 0.0183 | 5.97E-61 | Yes | Yes |
| rs11010905 | 10 | 19934813 | A | T | 0.4855 | -0.0602 | 0.0173 | 0.0005097 | No | No |
| rs72795925 | 10 | 20531420 | T | C | 0.2206 | -0.0428 | 0.0209 | 0.04048 | Yes | No |
| rs10732433 | 10 | 21037294 | T | C | 0.4205 | -0.0324 | 0.0175 | 0.06419 | Yes | No |
| rs3802517 | 10 | 28233469 | A | T | 0.4615 | 0.1286 | 0.0173 | 9.29E-14 | No | No |
| rs1265842 | 10 | 28924901 | T | C | 0.4834 | 0.1113 | 0.0174 | 1.70E-10 | Yes | No |
| rs9337951 | 10 | 30317073 | A | G | 0.3417 | -0.1073 | 0.0191 | 1.89E-08 | No | No |
| rs11008355 | 10 | 31412561 | C | G | 0.2389 | 0.0893 | 0.0203 | 1.10E-05 | Yes | No |
| rs10826995 | 10 | 32082658 | T | C | 0.7157 | -0.0328 | 0.0192 | 0.08807 | Yes | No |
| rs76164690 | 10 | 32590362 | T | G | 0.8587 | -0.154 | 0.025 | 7.19E-10 | Yes | No |
| rs2246438 | 10 | 45273079 | A | G | 0.2757 | -0.1119 | 0.0194 | 7.81E-09 | Yes | No |
| rs34130368 | 10 | 48411796 | T | G | 0.1172 | -0.2027 | 0.0284 | 8.77E-13 | No | No |
| rs10761530 | 10 | 62390726 | T | C | 0.4981 | 0.117 | 0.0172 | 1.14E-11 | Yes | No |
| rs1530440 | 10 | 63524591 | T | C | 0.1877 | -0.3877 | 0.0221 | 9.10E-69 | Yes | No |
| rs7090758 | 10 | 65335315 | T | C | 0.5282 | -0.1533 | 0.0173 | 6.89E-19 | Yes | No |
| rs7914287 | 10 | 69350563 | T | C | 0.7801 | -0.0442 | 0.0211 | 0.03633 | Yes | No |
| rs10823136 | 10 | 69855363 | T | C | 0.9223 | -0.1062 | 0.0336 | 0.001568 | Yes | No |
| rs10998362 | 10 | 70404159 | T | C | 0.3169 | -0.0278 | 0.0192 | 0.1474 | Yes | No |
| rs12572586 | 10 | 74751579 | T | C | 0.9375 | -0.1724 | 0.0366 | 2.54E-06 | Yes | No |
| rs10887914 | 10 | 82215288 | T | C | 0.462 | 0.0456 | 0.0173 | 0.008412 | Yes | No |
| rs77413490 | 10 | 89681688 | T | G | 0.0424 | 0.1757 | 0.0438 | 5.92E-05 | Yes | No |
| rs11187142 | 10 | 94468685 | T | C | 0.1049 | 0.1237 | 0.0284 | 1.34E-05 | Yes | No |
| rs932764 | 10 | 95895940 | A | G | 0.5696 | -0.1809 | 0.0175 | 5.94E-25 | Yes | No |
| rs4494250 | 10 | 96563757 | A | G | 0.3627 | 0.1917 | 0.0181 | 3.06E-26 | Yes | No |
| rs603424 | 10 | 102075479 | A | G | 0.1772 | 0.1776 | 0.023 | 1.24E-14 | Yes | No |
| rs112184198 | 10 | 102604514 | A | G | 0.1044 | -0.3566 | 0.0285 | 8.23E-36 | Yes | No |
| rs72847884 | 10 | 103115345 | A | G | 0.9534 | 0.2664 | 0.0423 | 3.04E-10 | Yes | No |
| rs11191156 | 10 | 103702763 | A | G | 0.6482 | -8.00E-04 | 0.0182 | 0.9644 | Yes | No |
| rs11191548 | 10 | 104846178 | T | C | 0.9191 | 0.5062 | 0.0318 | 4.53E-57 | Yes | No |
| rs4387287 | 10 | 105677897 | A | C | 0.1642 | 0.1575 | 0.0234 | 1.77E-11 | Yes | No |
| rs191784289 | 10 | 106894942 | T | C | 0.0123 | 0.7062 | 0.0815 | 4.66E-18 | Yes | No |
| rs111777102 | 10 | 111965826 | T | C | 0.0657 | 0.214 | 0.0354 | 1.56E-09 | Yes | No |
| rs34872471 | 10 | 114754071 | T | C | 0.7077 | -0.0116 | 0.0191 | 0.5433 | Yes | No |
| rs2782980 | 10 | 115781527 | T | C | 0.2832 | -0.298 | 0.0194 | 3.25E-53 | Yes | No |
| rs11197813 | 10 | 118523933 | A | G | 0.699 | -0.0911 | 0.0189 | 1.47E-06 | Yes | No |
| rs72842207 | 10 | 121433675 | T | C | 0.2149 | -0.2112 | 0.0211 | 1.10E-23 | Yes | No |
| rs11592107 | 10 | 122968964 | A | G | 0.3094 | 0.1203 | 0.0187 | 1.23E-10 | Yes | No |
| rs72834453 | 10 | 124235226 | T | G | 0.8762 | -0.1555 | 0.0267 | 5.87E-09 | Yes | No |
| rs4411245 | 10 | 126712781 | A | G | 0.294 | 0.0929 | 0.019 | 9.74E-07 | Yes | No |
| rs7096563 | 10 | 133770229 | A | G | 0.3547 | 0.1122 | 0.0184 | 1.00E-09 | Yes | No |
| rs1133400 | 10 | 134459388 | A | G | 0.7852 | -0.1318 | 0.0215 | 8.30E-10 | Yes | No |
| rs7126805 | 11 | 828916 | A | G | 0.7278 | 0.0066 | 0.02 | 0.7423 | Yes | No |
| rs661348 | 11 | 1905292 | T | C | 0.5757 | -0.1966 | 0.0181 | 1.33E-27 | Yes | No |
| rs17224476 | 11 | 4673788 | A | G | 0.1116 | 0.1597 | 0.0276 | 7.37E-09 | Yes | Yes |
| rs2929184 | 11 | 6289118 | A | G | 0.7711 | 0.1092 | 0.0214 | 3.29E-07 | Yes | No |
| rs110419 | 11 | 8252853 | A | G | 0.4781 | 0.112 | 0.0171 | 6.16E-11 | Yes | No |
| rs10743086 | 11 | 8774923 | A | G | 0.2071 | -0.1317 | 0.0214 | 7.46E-10 | Yes | Yes |
| rs360153 | 11 | 9762274 | T | C | 0.4172 | -0.2198 | 0.0175 | 4.37E-36 | Yes | No |
| rs7129220 | 11 | 10350538 | A | G | 0.1191 | 0.2636 | 0.0271 | 2.02E-22 | Yes | No |
| rs900145 | 11 | 13293905 | T | C | 0.7038 | 0.1493 | 0.0189 | 2.62E-15 | Yes | Yes |
| rs4757391 | 11 | 16302939 | T | C | 0.7977 | -0.3041 | 0.0215 | 1.69E-45 | Yes | Yes |
| rs381815 | 11 | 16902268 | T | C | 0.2754 | 0.1733 | 0.0194 | 4.00E-19 | Yes | No |
| rs5219 | 11 | 17409572 | T | C | 0.3654 | 0.1317 | 0.0179 | 2.10E-13 | Yes | No |
| rs10766533 | 11 | 19224677 | A | T | 0.7112 | 0.082 | 0.0193 | 2.19E-05 | Yes | No |
| rs11026586 | 11 | 22515533 | A | G | 0.0702 | 0.2898 | 0.0343 | 2.68E-17 | Yes | No |
| rs11030119 | 11 | 27728102 | A | G | 0.302 | -0.1679 | 0.0189 | 7.31E-19 | Yes | No |
| rs871004 | 11 | 28512458 | A | G | 0.3479 | 0.1188 | 0.0182 | 6.05E-11 | Yes | No |
| rs11031051 | 11 | 30355707 | A | C | 0.6909 | -0.0529 | 0.0188 | 0.0049 | Yes | No |
| rs919045 | 11 | 31111810 | T | C | 0.6289 | 0.1193 | 0.0179 | 2.75E-11 | Yes | No |
| rs4922591 | 11 | 32374199 | T | C | 0.3853 | -0.0355 | 0.018 | 0.04865 | Yes | No |
| rs190194639 | 11 | 34068037 | T | C | 0.0764 | 0.1479 | 0.0331 | 7.84E-06 | Yes | No |
| rs7480089 | 11 | 45207851 | A | G | 0.1176 | 0.0033 | 0.0271 | 0.9023 | Yes | No |
| rs1585453 | 11 | 46884713 | A | T | 0.887 | -0.238 | 0.0281 | 2.35E-17 | Yes | No |
| rs7103648 | 11 | 47461783 | A | G | 0.613 | -0.235 | 0.0178 | 7.18E-40 | Yes | Yes |
| rs11537751 | 11 | 47587452 | T | C | 0.0551 | 0.2117 | 0.0385 | 3.75E-08 | Yes | No |
| rs2688716 | 11 | 54835623 | T | G | 0.2891 | -0.1333 | 0.02 | 2.38E-11 | No | No |
| rs75905900 | 11 | 55113534 | A | C | 0.8653 | 0.2015 | 0.0257 | 4.10E-15 | Yes | No |
| rs11607056 | 11 | 57496820 | T | C | 0.3281 | -0.0624 | 0.0184 | 0.0006895 | Yes | No |
| rs11229457 | 11 | 58207203 | T | C | 0.2125 | -0.1504 | 0.0212 | 1.25E-12 | Yes | No |
| rs751984 | 11 | 61278246 | T | C | 0.8826 | 0.3937 | 0.0275 | 1.38E-46 | Yes | Yes |
| rs4980515 | 11 | 63744609 | T | C | 0.4987 | 0.0687 | 0.0174 | 7.61E-05 | Yes | No |
| rs3741378 | 11 | 65408937 | T | C | 0.1354 | -0.2272 | 0.0255 | 5.79E-19 | Yes | No |
| rs67976715 | 11 | 68023742 | C | G | 0.2342 | 0.1333 | 0.0206 | 9.78E-11 | Yes | No |
| rs67330701 | 11 | 69079707 | T | C | 0.0933 | -0.2798 | 0.0322 | 3.22E-18 | Yes | No |
| rs875106 | 11 | 70005641 | A | G | 0.5219 | -0.1329 | 0.0173 | 1.71E-14 | Yes | Yes |
| rs504217 | 11 | 72006086 | T | C | 0.0736 | 0.2745 | 0.0335 | 2.51E-16 | Yes | No |
| rs2298807 | 11 | 73068571 | T | C | 0.7842 | 0.1233 | 0.0211 | 4.89E-09 | Yes | Yes |
| rs4420291 | 11 | 74374950 | A | G | 0.5056 | 0.0971 | 0.0174 | 2.17E-08 | Yes | No |
| rs7927515 | 11 | 76125330 | A | C | 0.3459 | 0.1198 | 0.0183 | 5.92E-11 | Yes | No |
| rs59986178 | 11 | 77359909 | C | G | 0.1045 | 0.176 | 0.0299 | 3.92E-09 | Yes | No |
| rs2450128 | 11 | 77940075 | A | G | 0.1544 | -0.1505 | 0.024 | 3.53E-10 | Yes | No |
| rs2289125 | 11 | 89224453 | A | C | 0.2196 | 0.1181 | 0.0215 | 4.11E-08 | Yes | No |
| rs10830963 | 11 | 92708710 | C | G | 0.7237 | 0.0092 | 0.0199 | 0.6446 | Yes | No |
| rs11021221 | 11 | 95308854 | A | T | 0.1668 | -0.1877 | 0.0233 | 6.93E-16 | Yes | Yes |
| rs633185 | 11 | 100593538 | C | G | 0.7133 | 0.376 | 0.0192 | 2.29E-85 | Yes | Yes |
| rs61892344 | 11 | 101100768 | T | C | 0.1741 | -0.1172 | 0.0229 | 2.92E-07 | Yes | No |
| rs12807220 | 11 | 102077200 | A | G | 0.3596 | 0.0731 | 0.0182 | 5.88E-05 | Yes | No |
| rs4754196 | 11 | 107096777 | A | G | 0.5204 | -0.1535 | 0.0174 | 1.09E-18 | Yes | No |
| rs12362593 | 11 | 111586091 | C | G | 0.73 | -0.1275 | 0.0199 | 1.57E-10 | Yes | No |
| rs17119370 | 11 | 116097136 | A | T | 0.6933 | 0.1221 | 0.0189 | 1.11E-10 | Yes | No |
| rs1076485 | 11 | 116772441 | T | C | 0.13 | 0.1576 | 0.0262 | 1.83E-09 | Yes | No |
| rs8258 | 11 | 117283676 | T | C | 0.3746 | -0.063 | 0.0179 | 0.0004321 | Yes | No |
| rs12574332 | 11 | 122521123 | T | C | 0.1227 | 0.2072 | 0.0266 | 6.14E-15 | Yes | No |
| rs11222386 | 11 | 130779068 | C | G | 0.1971 | -0.0132 | 0.0218 | 0.5458 | Yes | No |
| rs78998485 | 12 | 434755 | C | G | 0.7438 | -0.0848 | 0.0199 | 2.14E-05 | Yes | No |
| rs11571376 | 12 | 1059556 | C | G | 0.706 | -0.0737 | 0.0192 | 0.0001205 | Yes | No |
| rs55935819 | 12 | 2521579 | A | G | 0.3636 | 0.1271 | 0.0181 | 1.96E-12 | Yes | No |
| rs117233107 | 12 | 4328521 | A | G | 0.0158 | -0.0846 | 0.0783 | 0.2801 | Yes | No |
| rs75507123 | 12 | 5417856 | T | G | 0.1274 | -0.1434 | 0.0261 | 3.94E-08 | Yes | No |
| rs7132012 | 12 | 8832203 | A | G | 0.6747 | 0.1563 | 0.0185 | 3.17E-17 | Yes | Yes |
| rs2024385 | 12 | 12888438 | A | T | 0.4241 | -0.1113 | 0.0177 | 2.88E-10 | No | No |
| rs28621435 | 12 | 13860990 | A | G | 0.1148 | -0.1333 | 0.0278 | 1.65E-06 | Yes | No |
| rs7313556 | 12 | 15297359 | A | G | 0.3487 | 0.0878 | 0.0181 | 1.24E-06 | Yes | No |
| rs61912333 | 12 | 19554817 | C | G | 0.4963 | 0.1191 | 0.0176 | 1.13E-11 | No | No |
| rs12579720 | 12 | 20173764 | C | G | 0.2435 | -0.2865 | 0.0203 | 3.46E-45 | Yes | Yes |
| rs73080726 | 12 | 20754154 | T | C | 0.1015 | -0.0272 | 0.0289 | 0.3468 | Yes | No |
| rs704191 | 12 | 22015022 | T | C | 0.4634 | -0.0155 | 0.0174 | 0.373 | Yes | No |
| rs7976167 | 12 | 24210599 | T | C | 0.6891 | 0.0868 | 0.0187 | 3.55E-06 | Yes | No |
| rs17287293 | 12 | 24770878 | A | G | 0.8505 | 0.1249 | 0.0242 | 2.48E-07 | Yes | No |
| rs6487543 | 12 | 26438189 | A | G | 0.7705 | 0.1325 | 0.0212 | 4.21E-10 | Yes | No |
| rs1098708 | 12 | 27321112 | A | G | 0.5413 | -0.0956 | 0.0175 | 4.63E-08 | Yes | No |
| rs10842991 | 12 | 27962103 | T | C | 0.1973 | 0.0921 | 0.022 | 2.87E-05 | Yes | No |
| rs7965392 | 12 | 42540280 | A | G | 0.387 | 0.1118 | 0.0179 | 4.16E-10 | Yes | No |
| rs11168245 | 12 | 48204499 | C | G | 0.7644 | 0.1758 | 0.0205 | 1.09E-17 | No | No |
| rs2261608 | 12 | 48721634 | A | T | 0.3512 | 0.0016 | 0.0181 | 0.9313 | Yes | No |
| rs1126930 | 12 | 49399132 | C | G | 0.0351 | 0.2633 | 0.0491 | 8.26E-08 | Yes | No |
| rs7977389 | 12 | 49981722 | T | C | 0.8937 | 0.0713 | 0.0282 | 0.01155 | Yes | No |
| rs7302981 | 12 | 50537815 | A | G | 0.3783 | 0.2652 | 0.0177 | 1.69E-50 | Yes | Yes |
| rs61926181 | 12 | 50767037 | A | G | 0.0385 | -0.4813 | 0.0484 | 2.76E-23 | Yes | No |
| rs73099903 | 12 | 53440779 | T | C | 0.0829 | 0.2204 | 0.032 | 5.57E-12 | Yes | No |
| rs7297416 | 12 | 54443090 | A | C | 0.6958 | 0.1597 | 0.019 | 4.33E-17 | Yes | No |
| rs7137749 | 12 | 57098040 | T | C | 0.3675 | 0.1412 | 0.0181 | 7.23E-15 | Yes | No |
| rs10437954 | 12 | 58003922 | A | G | 0.9068 | -0.2062 | 0.0308 | 2.14E-11 | Yes | No |
| rs4143175 | 12 | 67782397 | T | C | 0.2411 | 0.1139 | 0.0203 | 2.11E-08 | Yes | No |
| rs513177 | 12 | 69950545 | T | C | 0.1404 | 0.1694 | 0.0249 | 9.93E-12 | No | No |
| rs7963801 | 12 | 79685226 | T | C | 0.4226 | -0.1083 | 0.0179 | 1.49E-09 | No | No |
| rs17249754 | 12 | 90060586 | A | G | 0.1685 | -0.3826 | 0.0232 | 5.89E-61 | Yes | No |
| rs10858966 | 12 | 90567026 | C | G | 0.2987 | 0.151 | 0.0191 | 2.77E-15 | Yes | Yes |
| rs76785029 | 12 | 94882905 | T | C | 0.079 | 0.1561 | 0.0334 | 2.96E-06 | Yes | No |
| rs7977311 | 12 | 95487226 | T | C | 0.1153 | -0.0153 | 0.0271 | 0.5727 | Yes | No |
| rs11108209 | 12 | 96109855 | T | C | 0.9068 | -0.1901 | 0.03 | 2.40E-10 | Yes | Yes |
| rs7134060 | 12 | 96717095 | A | G | 0.4467 | -0.1058 | 0.0174 | 1.13E-09 | Yes | No |
| rs10778174 | 12 | 102838996 | A | G | 0.248 | -0.0922 | 0.0201 | 4.37E-06 | Yes | No |
| rs11112548 | 12 | 105871914 | A | T | 0.9556 | 0.2742 | 0.0443 | 5.80E-10 | Yes | No |
| rs12184466 | 12 | 111281636 | T | C | 0.1999 | 0.2533 | 0.0249 | 2.83E-24 | No | No |
| rs3184504 | 12 | 111884608 | T | C | 0.4808 | 0.4999 | 0.0175 | 8.04E-180 | Yes | Yes |
| rs10850411 | 12 | 115387796 | T | C | 0.6966 | 0.1765 | 0.0188 | 6.24E-21 | Yes | No |
| rs35444 | 12 | 115552437 | A | G | 0.6141 | 0.2671 | 0.0179 | 1.91E-50 | Yes | Yes |
| rs11067763 | 12 | 116198341 | A | G | 0.8986 | 0.2177 | 0.0288 | 4.43E-14 | Yes | Yes |
| rs11615689 | 12 | 116699675 | T | C | 0.8198 | 0.008 | 0.0225 | 0.7233 | Yes | No |
| rs3898618 | 12 | 120813921 | T | C | 0.9445 | -0.268 | 0.0383 | 2.49E-12 | Yes | No |
| rs28498002 | 12 | 122599796 | T | C | 0.4753 | -0.1566 | 0.0178 | 1.44E-18 | Yes | No |
| rs1060105 | 12 | 123806219 | T | C | 0.2031 | -0.1894 | 0.0216 | 2.11E-18 | Yes | Yes |
| rs1271309 | 12 | 124820705 | A | G | 0.1621 | -0.1979 | 0.024 | 1.45E-16 | Yes | No |
| rs117206641 | 12 | 133086888 | T | C | 0.1107 | 0.1454 | 0.0287 | 3.98E-07 | Yes | No |
| rs2480171 | 13 | 21559858 | T | C | 0.1214 | 0.1114 | 0.027 | 3.74E-05 | Yes | No |
| rs606950 | 13 | 22298923 | A | G | 0.6158 | 0.1383 | 0.0179 | 1.26E-14 | Yes | No |
| rs55641580 | 13 | 25257917 | T | C | 0.126 | 0.1745 | 0.0265 | 4.79E-11 | Yes | No |
| rs1331012 | 13 | 27115424 | T | G | 0.2706 | 0.0935 | 0.0195 | 1.62E-06 | Yes | No |
| rs63418562 | 13 | 30146201 | T | C | 0.7569 | -0.1944 | 0.0204 | 1.34E-21 | No | No |
| rs9532243 | 13 | 32191408 | A | C | 0.4843 | 0.1339 | 0.0173 | 1.09E-14 | Yes | No |
| rs9549297 | 13 | 41397482 | A | G | 0.8203 | -0.1483 | 0.0229 | 9.27E-11 | Yes | No |
| rs4274337 | 13 | 41967193 | A | G | 0.169 | -0.1464 | 0.0234 | 4.07E-10 | Yes | No |
| rs73187288 | 13 | 42738672 | A | C | 0.8909 | -0.1344 | 0.0279 | 1.43E-06 | Yes | No |
| rs912434 | 13 | 47189928 | T | G | 0.759 | 0.0956 | 0.0202 | 2.27E-06 | Yes | No |
| rs12583615 | 13 | 50564085 | A | G | 0.132 | 0.1576 | 0.0258 | 9.46E-10 | Yes | No |
| rs9526707 | 13 | 51489186 | A | G | 0.3222 | -0.1217 | 0.0186 | 6.59E-11 | Yes | No |
| rs75961402 | 13 | 56398286 | A | G | 0.1535 | 0.1254 | 0.0241 | 2.01E-07 | Yes | No |
| rs9563529 | 13 | 58316637 | T | G | 0.2043 | 0.1222 | 0.0215 | 1.38E-08 | Yes | No |
| rs3861113 | 13 | 72364382 | A | C | 0.0825 | 0.2126 | 0.0322 | 3.95E-11 | Yes | Yes |
| rs78474310 | 13 | 73826901 | A | G | 0.9551 | -0.2428 | 0.0423 | 9.74E-09 | Yes | No |
| rs4304924 | 13 | 79238925 | A | G | 0.5677 | 0.0269 | 0.0176 | 0.1262 | Yes | No |
| rs7988232 | 13 | 79808655 | A | G | 0.4109 | 0.0939 | 0.0178 | 1.24E-07 | Yes | No |
| rs1215469 | 13 | 80707408 | A | C | 0.2295 | -0.1383 | 0.0211 | 5.23E-11 | Yes | No |
| rs55684003 | 13 | 97988689 | A | G | 0.6959 | 0.122 | 0.0189 | 1.01E-10 | Yes | No |
| rs3742182 | 13 | 111375132 | T | C | 0.8103 | -0.0261 | 0.0221 | 0.2373 | Yes | No |
| rs9549328 | 13 | 113636156 | T | C | 0.2303 | 0.0868 | 0.0209 | 3.18E-05 | Yes | No |
| rs7331680 | 13 | 115000650 | T | G | 0.1488 | 0.1767 | 0.0244 | 4.80E-13 | Yes | No |
| rs17880989 | 14 | 23313633 | A | G | 0.0259 | 0.4014 | 0.0591 | 1.11E-11 | No | No |
| rs452036 | 14 | 23865885 | A | G | 0.3546 | 0.0873 | 0.0182 | 1.64E-06 | Yes | No |
| rs17115145 | 14 | 30122409 | T | C | 0.397 | 0.0924 | 0.0177 | 1.86E-07 | Yes | No |
| rs4424827 | 14 | 35110857 | T | C | 0.5669 | -0.0981 | 0.0175 | 2.11E-08 | Yes | No |
| rs8904 | 14 | 35871217 | A | G | 0.3674 | 0.1 | 0.0181 | 3.36E-08 | Yes | No |
| rs34983854 | 14 | 39858442 | A | G | 0.6022 | -0.071 | 0.0177 | 6.06E-05 | Yes | No |
| rs72683923 | 14 | 50735947 | T | C | 0.9788 | 0.5325 | 0.0635 | 5.02E-17 | No | No |
| rs9888615 | 14 | 53377540 | T | C | 0.2906 | -0.112 | 0.0192 | 4.96E-09 | Yes | No |
| rs210381 | 14 | 54107791 | A | G | 0.5655 | -0.0081 | 0.0175 | 0.6453 | Yes | No |
| rs7144602 | 14 | 55285588 | T | G | 0.6484 | -0.0348 | 0.0183 | 0.05715 | Yes | No |
| rs11628933 | 14 | 60700903 | C | G | 0.2336 | -0.1222 | 0.0206 | 3.12E-09 | Yes | Yes |
| rs731681 | 14 | 68010224 | C | G | 0.5584 | -0.1071 | 0.0174 | 7.98E-10 | No | No |
| rs57786342 | 14 | 69260028 | A | G | 0.2055 | 0.1423 | 0.0216 | 4.37E-11 | Yes | No |
| rs11623535 | 14 | 72462381 | A | G | 0.7442 | 0.1026 | 0.0198 | 2.20E-07 | Yes | No |
| rs4903064 | 14 | 73279420 | T | C | 0.7645 | 0.1543 | 0.0206 | 7.84E-14 | Yes | Yes |
| rs11159091 | 14 | 75074316 | A | G | 0.4615 | 0.0631 | 0.0175 | 0.0003164 | Yes | No |
| rs11627326 | 14 | 85785251 | C | G | 0.2869 | -0.0604 | 0.0192 | 0.001708 | Yes | No |
| rs4904503 | 14 | 89565130 | T | C | 0.298 | -0.073 | 0.019 | 0.0001261 | Yes | No |
| rs11160085 | 14 | 93112102 | T | C | 0.7034 | 0.024 | 0.0193 | 0.2133 | Yes | No |
| rs8013933 | 14 | 94465789 | T | C | 0.6909 | 0.0421 | 0.0189 | 0.02538 | Yes | No |
| rs9323988 | 14 | 98587630 | T | C | 0.6158 | -0.0558 | 0.0178 | 0.001736 | Yes | No |
| rs1475130 | 14 | 100225144 | T | C | 0.3466 | -0.0012 | 0.0183 | 0.948 | Yes | No |
| rs28470843 | 14 | 100742658 | T | C | 0.5973 | -0.0266 | 0.0178 | 0.1351 | Yes | No |
| rs11626434 | 14 | 101998443 | C | G | 0.361 | 0.0219 | 0.0185 | 0.2343 | Yes | No |
| rs8014182 | 14 | 103859962 | T | C | 0.1319 | -0.1942 | 0.0257 | 3.94E-14 | Yes | Yes |
| rs34161718 | 14 | 104620193 | T | C | 0.2417 | -0.1319 | 0.0216 | 1.07E-09 | No | No |
| rs10873612 | 15 | 26105602 | T | C | 0.5961 | -0.1096 | 0.0179 | 9.51E-10 | Yes | No |
| rs11629850 | 15 | 40317075 | A | G | 0.529 | 0.1279 | 0.0174 | 1.78E-13 | Yes | No |
| rs2925345 | 15 | 41311799 | T | C | 0.4676 | 0.189 | 0.0174 | 1.60E-27 | Yes | Yes |
| rs4924570 | 15 | 41974660 | T | C | 0.6287 | -0.1692 | 0.0181 | 9.62E-21 | Yes | Yes |
| rs1036477 | 15 | 48914926 | A | G | 0.8973 | -0.1015 | 0.0285 | 0.0003731 | Yes | No |
| rs3098186 | 15 | 50810621 | T | C | 0.5159 | -0.068 | 0.0175 | 0.0001014 | Yes | No |
| rs3191402 | 15 | 59429160 | A | G | 0.4073 | -0.0127 | 0.0179 | 0.4789 | No | No |
| rs956006 | 15 | 62808539 | T | C | 0.3307 | 0.0752 | 0.0186 | 5.35E-05 | Yes | No |
| rs832890 | 15 | 65166309 | T | C | 0.4655 | -0.026 | 0.0174 | 0.1349 | Yes | No |
| rs7178615 | 15 | 66869072 | A | G | 0.3756 | -0.1371 | 0.018 | 2.90E-14 | Yes | No |
| rs2289261 | 15 | 67457485 | C | G | 0.6524 | 0.0553 | 0.0183 | 0.002497 | Yes | No |
| rs62004794 | 15 | 68454523 | A | G | 0.4394 | -0.0962 | 0.0174 | 3.38E-08 | Yes | No |
| rs11853359 | 15 | 71621524 | A | G | 0.3324 | -0.166 | 0.0183 | 1.30E-19 | Yes | Yes |
| rs61653296 | 15 | 74557817 | A | G | 0.8008 | -0.1405 | 0.0218 | 1.13E-10 | Yes | No |
| rs1378942 | 15 | 75077367 | A | C | 0.6688 | -0.388 | 0.0185 | 6.03E-98 | Yes | Yes |
| rs11634028 | 15 | 76276150 | A | T | 0.2149 | 0.1114 | 0.0216 | 2.52E-07 | Yes | Yes |
| rs62011052 | 15 | 79156983 | T | C | 0.8494 | 0.1467 | 0.0244 | 1.76E-09 | Yes | No |
| rs2759308 | 15 | 81016227 | A | G | 0.4752 | 0.1409 | 0.0175 | 7.11E-16 | Yes | No |
| rs2034618 | 15 | 83799632 | T | C | 0.2224 | -0.1157 | 0.0209 | 3.36E-08 | Yes | No |
| rs899927 | 15 | 84561879 | T | C | 0.3183 | -0.0282 | 0.0188 | 0.1339 | Yes | No |
| rs7180952 | 15 | 85162551 | T | C | 0.5393 | -0.1008 | 0.0176 | 9.82E-09 | Yes | No |
| rs3743157 | 15 | 85680532 | A | C | 0.1671 | 0.1284 | 0.0233 | 3.72E-08 | Yes | No |
| rs11632436 | 15 | 86295286 | C | G | 0.5032 | 0.0996 | 0.0174 | 1.09E-08 | No | No |
| rs28611491 | 15 | 90641809 | T | C | 0.0808 | 0.0864 | 0.0332 | 0.00937 | Yes | No |
| rs2521501 | 15 | 91437388 | A | T | 0.6747 | -0.3693 | 0.0191 | 1.84E-83 | No | No |
| rs873122 | 15 | 92702020 | C | G | 0.7196 | 0.121 | 0.0196 | 6.52E-10 | Yes | Yes |
| rs11632112 | 15 | 93468276 | C | G | 0.2405 | -0.0089 | 0.0204 | 0.6609 | Yes | No |
| rs12906962 | 15 | 95312071 | T | C | 0.6767 | -0.2378 | 0.0188 | 8.73E-37 | Yes | Yes |
| rs4984496 | 15 | 96635898 | T | G | 0.3353 | 0.1763 | 0.0187 | 4.93E-21 | Yes | No |
| rs34756251 | 15 | 100192540 | T | C | 0.1715 | -0.1386 | 0.023 | 1.75E-09 | Yes | No |
| rs9932866 | 16 | 706067 | A | G | 0.3667 | 0.1154 | 0.0183 | 2.90E-10 | Yes | No |
| rs11248862 | 16 | 1344291 | A | G | 0.126 | 0.0663 | 0.0266 | 0.01274 | Yes | No |
| rs28590346 | 16 | 2080653 | A | T | 0.6596 | -0.1914 | 0.0191 | 9.84E-24 | Yes | No |
| rs2379829 | 16 | 3538873 | C | G | 0.7307 | -0.148 | 0.0198 | 6.84E-14 | Yes | No |
| rs4785955 | 16 | 4297651 | T | G | 0.2178 | -0.0342 | 0.0216 | 0.113 | Yes | No |
| rs12921187 | 16 | 4943019 | T | G | 0.4283 | -0.175 | 0.0175 | 1.65E-23 | Yes | Yes |
| rs35450617 | 16 | 6889675 | T | G | 0.6997 | -0.079 | 0.0192 | 4.00E-05 | Yes | No |
| rs11642631 | 16 | 11198835 | T | C | 0.5624 | 0.0069 | 0.0175 | 0.695 | Yes | No |
| rs57327054 | 16 | 14487036 | T | C | 0.3081 | -0.1173 | 0.0191 | 8.07E-10 | Yes | No |
| rs3915425 | 16 | 15912544 | T | C | 0.6822 | 0.0125 | 0.0188 | 0.505 | Yes | No |
| rs4782211 | 16 | 19152219 | A | G | 0.271 | 0.1076 | 0.0208 | 2.45E-07 | No | No |
| rs13333226 | 16 | 20365654 | A | G | 0.8159 | 0.2965 | 0.0223 | 2.94E-40 | Yes | Yes |
| rs11639856 | 16 | 24788645 | A | T | 0.1936 | -0.1003 | 0.0219 | 4.59E-06 | Yes | No |
| rs6565174 | 16 | 30111904 | A | C | 0.1126 | -0.195 | 0.0278 | 2.42E-12 | Yes | Yes |
| rs72799341 | 16 | 30936743 | A | G | 0.2398 | 0.1599 | 0.0204 | 4.92E-15 | Yes | No |
| rs10468291 | 16 | 49768046 | A | C | 0.569 | -0.1166 | 0.0176 | 3.70E-11 | Yes | Yes |
| rs34941092 | 16 | 50550137 | A | G | 0.1498 | -0.1538 | 0.0245 | 3.42E-10 | Yes | No |
| rs9932220 | 16 | 51758116 | A | G | 0.2177 | -0.1591 | 0.021 | 3.76E-14 | Yes | Yes |
| rs37060 | 16 | 58566304 | A | G | 0.2466 | -0.0341 | 0.0201 | 0.08935 | Yes | No |
| rs28633979 | 16 | 65282820 | A | C | 0.4305 | 0.0198 | 0.0175 | 0.2574 | Yes | No |
| rs45474499 | 16 | 66914492 | T | C | 0.0473 | 0.3562 | 0.0415 | 8.50E-18 | Yes | No |
| rs7185555 | 16 | 69131281 | C | G | 0.1533 | -0.154 | 0.0243 | 2.34E-10 | Yes | No |
| rs33063 | 16 | 69640217 | A | G | 0.149 | -0.0914 | 0.0244 | 0.0001827 | Yes | No |
| rs62053102 | 16 | 71654365 | A | T | 0.9525 | 0.0322 | 0.0431 | 0.4554 | Yes | No |
| rs1012089 | 16 | 74171973 | C | G | 0.475 | -0.0994 | 0.0174 | 1.19E-08 | No | No |
| rs35261357 | 16 | 75444572 | T | C | 0.5862 | 0.1216 | 0.0177 | 7.14E-12 | Yes | No |
| rs56844452 | 16 | 80864776 | T | C | 0.0707 | -0.1193 | 0.0344 | 0.0005228 | Yes | No |
| rs8059962 | 16 | 81574197 | T | C | 0.4208 | -0.1397 | 0.0177 | 3.38E-15 | Yes | No |
| rs7500448 | 16 | 83045790 | A | G | 0.7471 | -0.1303 | 0.0202 | 1.14E-10 | Yes | No |
| rs7187540 | 16 | 85318302 | A | C | 0.336 | -0.0914 | 0.0196 | 2.95E-06 | No | No |
| rs3851018 | 16 | 86437811 | C | G | 0.5694 | 0.0744 | 0.0178 | 3.06E-05 | No | No |
| rs6540125 | 16 | 87993889 | T | G | 0.3406 | 0.0888 | 0.0183 | 1.28E-06 | Yes | No |
| rs1126464 | 16 | 89704365 | C | G | 0.2428 | 0.2071 | 0.0208 | 1.89E-23 | Yes | Yes |
| rs12941318 | 17 | 1333598 | T | C | 0.4952 | -0.0532 | 0.0178 | 0.002772 | Yes | No |
| rs4480845 | 17 | 1958609 | T | C | 0.3646 | 0.0943 | 0.0183 | 2.41E-07 | Yes | No |
| rs7215084 | 17 | 3880148 | T | C | 0.5127 | 0.1116 | 0.0173 | 1.17E-10 | Yes | Yes |
| rs28427409 | 17 | 6473882 | T | C | 0.4171 | -0.0066 | 0.0177 | 0.7071 | Yes | No |
| rs78378222 | 17 | 7571752 | T | G | 0.986 | -0.6122 | 0.0798 | 1.71E-14 | Yes | No |
| rs8069739 | 17 | 8078765 | T | C | 0.3213 | -0.0987 | 0.0186 | 1.04E-07 | Yes | No |
| rs4925159 | 17 | 18185510 | A | G | 0.4253 | 0.1202 | 0.0176 | 8.26E-12 | Yes | No |
| rs7502046 | 17 | 19196440 | T | C | 0.8108 | 0.1555 | 0.0224 | 3.95E-12 | Yes | No |
| rs138285687 | 17 | 27195674 | T | C | 0.0434 | -0.0146 | 0.0436 | 0.7374 | No | No |
| rs11080134 | 17 | 29161503 | A | G | 0.6425 | -0.0955 | 0.0184 | 2.19E-07 | Yes | No |
| rs1551355 | 17 | 30032420 | T | C | 0.2334 | 0.1185 | 0.0206 | 8.37E-09 | Yes | No |
| rs9899540 | 17 | 30777924 | A | T | 0.3987 | 0.0679 | 0.0182 | 0.0001874 | Yes | No |
| rs3135967 | 17 | 33313729 | A | G | 0.5242 | -0.0364 | 0.0174 | 0.03596 | Yes | No |
| rs79089478 | 17 | 40317241 | T | C | 0.9726 | -0.058 | 0.0545 | 0.2871 | Yes | No |
| rs56228409 | 17 | 40919596 | A | C | 0.8457 | 0.0452 | 0.0252 | 0.07296 | Yes | No |
| rs9904409 | 17 | 42680402 | A | G | 0.0979 | 0.0796 | 0.0293 | 0.006556 | Yes | No |
| rs12946454 | 17 | 43208121 | A | T | 0.7343 | -0.1614 | 0.0197 | 2.43E-16 | Yes | No |
| rs115231027 | 17 | 44199290 | T | C | 0.8207 | -0.0904 | 0.0239 | 0.0001561 | No | No |
| rs17608766 | 17 | 45013271 | T | C | 0.8552 | -0.1707 | 0.0249 | 7.63E-12 | Yes | No |
| rs62076103 | 17 | 45888374 | A | G | 0.9311 | -0.1437 | 0.0348 | 3.62E-05 | Yes | No |
| rs7406910 | 17 | 46688256 | T | C | 0.0885 | -0.1426 | 0.0308 | 3.51E-06 | Yes | No |
| rs12940887 | 17 | 47402807 | T | C | 0.3666 | 0.2279 | 0.018 | 9.63E-37 | Yes | Yes |
| rs12325702 | 17 | 55446364 | A | T | 0.4435 | -0.0287 | 0.0177 | 0.1048 | No | No |
| rs34430710 | 17 | 56876627 | A | T | 0.6769 | -0.1291 | 0.0186 | 3.81E-12 | Yes | No |
| rs2645466 | 17 | 57853214 | A | C | 0.705 | -0.0275 | 0.019 | 0.1481 | Yes | No |
| rs1036902 | 17 | 58950791 | T | C | 0.8478 | -0.1257 | 0.0244 | 2.72E-07 | Yes | No |
| rs2240736 | 17 | 59485393 | T | C | 0.7347 | 0.189 | 0.0198 | 1.59E-21 | Yes | No |
| rs740698 | 17 | 60767151 | T | C | 0.5642 | 0.0178 | 0.0178 | 0.3156 | Yes | No |
| rs4308 | 17 | 61559625 | A | G | 0.3772 | 0.1753 | 0.0181 | 3.74E-22 | Yes | No |
| rs6504213 | 17 | 62381714 | T | C | 0.4175 | -0.1335 | 0.018 | 1.17E-13 | Yes | No |
| rs112260610 | 17 | 64252393 | T | C | 0.1407 | 0.1088 | 0.0251 | 1.41E-05 | Yes | No |
| rs2467099 | 17 | 73949045 | T | C | 0.2233 | -0.1428 | 0.0209 | 7.58E-12 | Yes | No |
| rs35504735 | 17 | 74686809 | A | G | 0.5578 | -0.0926 | 0.0174 | 1.06E-07 | Yes | No |
| rs57927100 | 17 | 75317300 | C | G | 0.7366 | 0.206 | 0.02 | 7.34E-25 | Yes | No |
| rs9302885 | 17 | 76799898 | A | G | 0.4454 | 0.0976 | 0.0174 | 2.13E-08 | Yes | No |
| rs112280096 | 17 | 79367409 | A | C | 0.362 | -0.1186 | 0.0192 | 6.95E-10 | No | No |
| rs34413141 | 18 | 777282 | A | T | 0.1821 | -0.1808 | 0.0227 | 1.49E-15 | Yes | No |
| rs11665020 | 18 | 10879503 | C | G | 0.322 | -0.1423 | 0.0187 | 2.78E-14 | Yes | No |
| rs963920 | 18 | 12711052 | T | G | 0.6751 | 0.0126 | 0.0185 | 0.4959 | Yes | No |
| rs4800420 | 18 | 20158965 | A | G | 0.2911 | 0.1192 | 0.0192 | 5.18E-10 | Yes | Yes |
| rs1154214 | 18 | 24546824 | T | G | 0.3963 | -0.1064 | 0.0177 | 1.77E-09 | Yes | No |
| rs10164193 | 18 | 31161426 | T | G | 0.9223 | -0.2196 | 0.0327 | 1.87E-11 | Yes | No |
| rs61735998 | 18 | 34289285 | T | G | 0.0242 | 0.0894 | 0.0592 | 0.1313 | Yes | No |
| rs12958173 | 18 | 42141977 | A | C | 0.2967 | 0.1704 | 0.019 | 2.99E-19 | Yes | No |
| rs7236548 | 18 | 43097750 | A | C | 0.1847 | -0.0285 | 0.0224 | 0.2031 | Yes | No |
| rs745821 | 18 | 48142854 | T | G | 0.7551 | 0.1544 | 0.0203 | 2.71E-14 | Yes | No |
| rs36010659 | 18 | 48283949 | T | C | 0.8585 | 0.0476 | 0.0249 | 0.05604 | Yes | No |
| rs11876341 | 18 | 48799991 | A | G | 0.692 | -0.1202 | 0.0193 | 4.35E-10 | Yes | No |
| rs34163044 | 18 | 51851616 | A | C | 0.42 | 0.1486 | 0.0179 | 9.63E-17 | Yes | No |
| rs72930904 | 18 | 52607301 | T | C | 0.16 | -0.14 | 0.0237 | 3.24E-09 | Yes | No |
| rs12605156 | 18 | 53498114 | A | T | 0.8084 | 0.1418 | 0.0221 | 1.51E-10 | Yes | Yes |
| rs10048404 | 18 | 54578482 | T | C | 0.3694 | -0.1096 | 0.0183 | 2.01E-09 | No | No |
| rs7235890 | 18 | 55732115 | T | G | 0.8956 | -0.1692 | 0.0288 | 4.12E-09 | Yes | No |
| rs6567160 | 18 | 57829135 | T | C | 0.7663 | 0.0852 | 0.0206 | 3.58E-05 | Yes | No |
| rs12172847 | 18 | 60223017 | A | G | 0.3233 | -0.0141 | 0.0185 | 0.4456 | Yes | No |
| rs12454712 | 18 | 60845884 | T | C | 0.6223 | 0.1047 | 0.0188 | 2.73E-08 | No | No |
| rs10460108 | 18 | 73034151 | A | G | 0.48 | 0.1038 | 0.0174 | 2.35E-09 | Yes | No |
| rs1047922 | 18 | 74070562 | T | C | 0.8482 | 0.0215 | 0.0252 | 0.3926 | No | No |
| rs7250835 | 19 | 670234 | T | C | 0.1646 | 0.0333 | 0.0247 | 0.1784 | Yes | No |
| rs3760994 | 19 | 1435771 | A | G | 0.492 | -0.042 | 0.0183 | 0.02183 | No | No |
| rs740406 | 19 | 2232221 | A | G | 0.9403 | -0.0836 | 0.0379 | 0.02741 | Yes | No |
| rs2656523 | 19 | 4085896 | C | G | 0.7736 | -0.0122 | 0.0213 | 0.5679 | Yes | No |
| rs2613765 | 19 | 5066330 | A | G | 0.4745 | -0.0976 | 0.0174 | 1.98E-08 | Yes | No |
| rs7248104 | 19 | 7224431 | A | G | 0.4129 | -0.0514 | 0.0177 | 0.003604 | Yes | No |
| rs4247374 | 19 | 7252756 | T | C | 0.1397 | -0.3616 | 0.0262 | 3.35E-43 | No | No |
| rs2009733 | 19 | 8398714 | A | G | 0.4995 | 0.1217 | 0.0176 | 5.10E-12 | Yes | No |
| rs10409243 | 19 | 10332988 | T | C | 0.5911 | -0.1199 | 0.018 | 2.89E-11 | Yes | No |
| rs1529744 | 19 | 10841472 | T | C | 0.3185 | -0.04 | 0.0187 | 0.03217 | Yes | No |
| rs167479 | 19 | 11526765 | T | G | 0.4722 | -0.362 | 0.0188 | 1.67E-82 | No | No |
| rs17638167 | 19 | 11584818 | T | C | 0.0437 | -0.256 | 0.0429 | 2.35E-09 | Yes | No |
| rs10418305 | 19 | 15278808 | C | G | 0.0989 | 0.1341 | 0.0292 | 4.36E-06 | Yes | No |
| rs3745318 | 19 | 16436262 | T | C | 0.2496 | 0.1396 | 0.0206 | 1.30E-11 | No | No |
| rs1077795 | 19 | 17222584 | A | G | 0.7389 | 0.1987 | 0.0199 | 1.62E-23 | Yes | No |
| rs8111708 | 19 | 18558876 | A | G | 0.6523 | -0.0807 | 0.0183 | 9.85E-06 | Yes | No |
| rs2304130 | 19 | 19789528 | A | G | 0.9156 | -0.2396 | 0.0318 | 4.48E-14 | Yes | No |
| rs6511291 | 19 | 21950402 | T | C | 0.436 | -0.1158 | 0.0177 | 6.89E-11 | Yes | Yes |
| rs62104477 | 19 | 30294991 | T | G | 0.3302 | 0.1703 | 0.0185 | 4.22E-20 | Yes | No |
| rs8105753 | 19 | 31927547 | A | C | 0.632 | 0.109 | 0.0183 | 2.71E-09 | No | No |
| rs1821295 | 19 | 32590773 | T | C | 0.6985 | -0.138 | 0.0189 | 3.13E-13 | Yes | No |
| rs7256564 | 19 | 33889593 | A | G | 0.3124 | 0.0787 | 0.0187 | 2.56E-05 | Yes | No |
| rs12983238 | 19 | 39438532 | A | G | 0.307 | -0.1266 | 0.0201 | 3.23E-10 | Yes | No |
| rs1800470 | 19 | 41858921 | A | G | 0.6242 | 0.0428 | 0.0181 | 0.01781 | Yes | No |
| rs7412 | 19 | 45412079 | T | C | 0.0817 | -0.0635 | 0.0331 | 0.05489 | Yes | No |
| rs34783010 | 19 | 46180414 | T | G | 0.1992 | 0.0212 | 0.022 | 0.3351 | Yes | No |
| rs73046792 | 19 | 49605705 | A | G | 0.1592 | -0.1518 | 0.0245 | 5.87E-10 | Yes | No |
| rs138877676 | 19 | 50935809 | T | G | 0.0193 | -0.2026 | 0.0712 | 0.004407 | Yes | No |
| rs2143635 | 20 | 2793063 | T | C | 0.8995 | 0.0718 | 0.0296 | 0.01531 | Yes | No |
| rs1764975 | 20 | 4101290 | A | T | 0.7956 | 0.1609 | 0.0219 | 1.88E-13 | Yes | No |
| rs11087740 | 20 | 6657554 | T | C | 0.5123 | 7.00E-04 | 0.0175 | 0.9667 | Yes | No |
| rs6108168 | 20 | 8626271 | A | C | 0.2546 | -0.1901 | 0.0199 | 1.10E-21 | Yes | No |
| rs680515 | 20 | 10458688 | A | G | 0.3897 | 0.1723 | 0.0179 | 5.61E-22 | Yes | No |
| rs1327235 | 20 | 10969030 | A | G | 0.5286 | -0.3018 | 0.0173 | 4.76E-68 | Yes | No |
| rs1232482 | 20 | 11886643 | T | C | 0.4019 | -0.1214 | 0.0176 | 6.10E-12 | Yes | No |
| rs2618647 | 20 | 17882452 | A | G | 0.5068 | -0.1216 | 0.0174 | 2.70E-12 | Yes | No |
| rs6081613 | 20 | 19465907 | A | G | 0.2748 | -0.1111 | 0.0194 | 1.00E-08 | Yes | No |
| rs6060114 | 20 | 30169673 | T | C | 0.8417 | 0.1689 | 0.0238 | 1.41E-12 | Yes | No |
| rs6141767 | 20 | 31225069 | C | G | 0.1568 | 0.1048 | 0.0241 | 1.41E-05 | Yes | No |
| rs13042148 | 20 | 32298286 | T | C | 0.1537 | -0.1674 | 0.0244 | 7.24E-12 | Yes | No |
| rs6141479 | 20 | 33121942 | C | G | 0.1961 | -0.0469 | 0.0226 | 0.03812 | Yes | No |
| rs4811601 | 20 | 36849007 | T | C | 0.4589 | -0.0669 | 0.0177 | 0.0001523 | Yes | No |
| rs4810332 | 20 | 40268334 | A | T | 0.3795 | -0.1715 | 0.0183 | 6.51E-21 | Yes | No |
| rs6031435 | 20 | 42797358 | A | G | 0.5399 | -0.1128 | 0.0175 | 1.10E-10 | Yes | No |
| rs6095241 | 20 | 47308798 | A | G | 0.4396 | -0.1358 | 0.0174 | 6.22E-15 | Yes | Yes |
| rs237485 | 20 | 48004238 | A | G | 0.6976 | 0.1124 | 0.0191 | 3.63E-09 | Yes | Yes |
| rs6021247 | 20 | 50108980 | A | G | 0.5289 | 0.1297 | 0.0174 | 8.21E-14 | Yes | No |
| rs6015450 | 20 | 57751117 | A | G | 0.8762 | -0.4911 | 0.0266 | 5.37E-76 | Yes | Yes |
| rs35213536 | 20 | 62694319 | T | G | 0.2467 | 0.2044 | 0.0205 | 2.54E-23 | Yes | No |
| rs1882961 | 21 | 16556367 | T | C | 0.3088 | 0.1272 | 0.0188 | 1.40E-11 | Yes | No |
| rs11909120 | 21 | 30131872 | A | T | 0.8565 | 0.121 | 0.0248 | 1.06E-06 | Yes | No |
| rs11701033 | 21 | 33788341 | C | G | 0.818 | -0.1072 | 0.0227 | 2.24E-06 | Yes | No |
| rs9976596 | 21 | 35596842 | T | C | 0.8433 | -0.112 | 0.0244 | 4.59E-06 | Yes | No |
| rs62229372 | 21 | 37692507 | T | C | 0.1276 | 0.1413 | 0.027 | 1.66E-07 | Yes | No |
| rs2277788 | 21 | 40817702 | C | G | 0.1059 | 0.0359 | 0.0282 | 0.2023 | Yes | No |
| rs12627651 | 21 | 44760603 | A | G | 0.2866 | 0.215 | 0.0197 | 7.86E-28 | Yes | Yes |
| rs9306160 | 21 | 45107562 | T | C | 0.4113 | -0.1435 | 0.0178 | 8.67E-16 | Yes | No |
| rs35796750 | 21 | 47422412 | T | C | 0.5496 | -0.0054 | 0.0176 | 0.7597 | Yes | No |
| rs11701512 | 21 | 47962811 | A | G | 0.1809 | 0.023 | 0.0225 | 0.3076 | Yes | No |
| rs12628032 | 22 | 19967980 | T | C | 0.309 | 0.0229 | 0.019 | 0.2266 | Yes | No |
| rs134041 | 22 | 28056338 | T | C | 0.436 | 0.1223 | 0.0175 | 3.05E-12 | Yes | Yes |
| rs9608690 | 22 | 28921347 | A | G | 0.0685 | -0.1321 | 0.0346 | 0.0001339 | Yes | No |
| rs4823006 | 22 | 29451671 | A | G | 0.5549 | 0.1396 | 0.0174 | 1.17E-15 | Yes | Yes |
| rs737721 | 22 | 30172254 | C | G | 0.9482 | -0.1144 | 0.0397 | 0.003903 | Yes | No |
| rs5753103 | 22 | 30768777 | A | G | 0.4515 | -0.0082 | 0.0175 | 0.639 | Yes | No |
| rs9609429 | 22 | 32517431 | T | C | 0.7236 | 0.1203 | 0.0195 | 6.32E-10 | Yes | Yes |
| rs5750482 | 22 | 38117943 | T | C | 0.3799 | 0.0312 | 0.0179 | 0.08182 | Yes | No |
| rs470113 | 22 | 40729614 | A | G | 0.8184 | 0.0476 | 0.0225 | 0.03445 | Yes | No |
| rs73161324 | 22 | 42038786 | T | C | 0.056 | -0.1327 | 0.0403 | 0.001004 | Yes | No |
| rs77692990 | 22 | 50219952 | T | C | 0.0798 | -0.1088 | 0.0326 | 0.0008343 | Yes | No |
| rs28578714 | 22 | 50727921 | T | C | 0.6064 | 0.0814 | 0.0188 | 1.46E-05 | Yes | No |

**Supplemental Table 9.** Number of genetic variants used in all analyses

|  | **Genetic variants**<br>**in original**<br>**GWAS (*n*, %)** | **Used in main**<br>**MR analyses**<br>**(*n*, %)** | **Used in MR**<br>**+ Steiger filt.**<br>**(*n*, %)** |
| --- | --- | --- | --- |
| Fasting glucose | 138 (100%) | 121 (87.7%) | 67 (48.6%) |
| Glycated hemoglobin | 146 (100%) | 124 (84.9%) | 47 (32.2%) |
| Fasting insulin | 62 (100%) | 56 (90.3%) | 30 (48.4%) |
| LDL cholesterol | 389 (100%) | 311 (79.9%) | 72 (18.5%) |
| HDL cholesterol | 371 (100%) | 311 (83.8%) | 34 (9.2%) |
| Triglycerides | 384 (100%) | 322 (83.9%) | 31 (8.1%) |
| Systolic blood pressure | 883 (100%) | 817 (92.5%) | 83 (9.4%) |
| Diastolic blood pressure | 885 (100%) | 818 (92.4%) | 114 (12.9%) |

**Supplemental Table 10.**
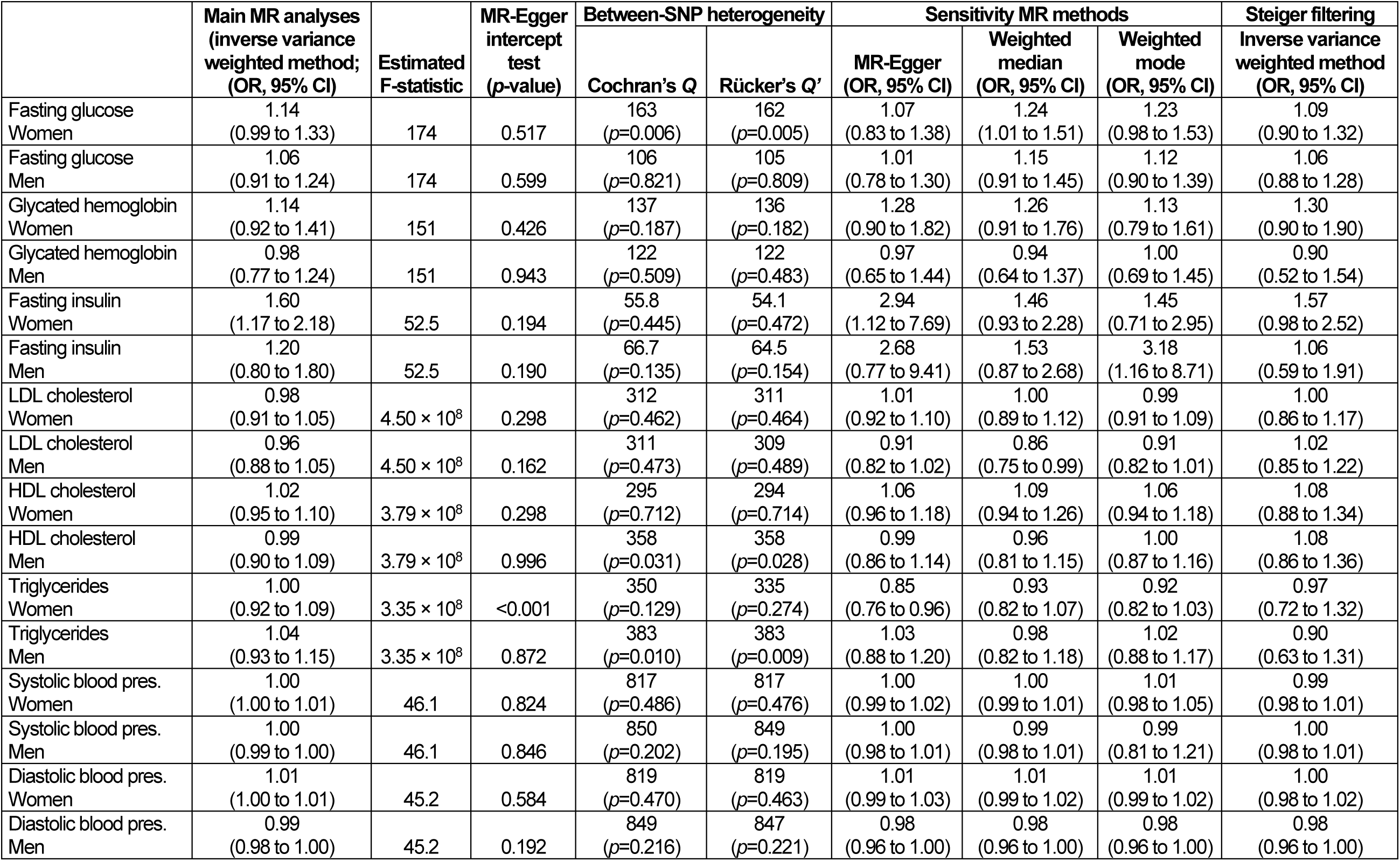
Sensitivity analyses on the verification of Mendelian randomization assumptions.

### SUPPLEMENTAL FIGURES

**Supplemental Figure 1.**
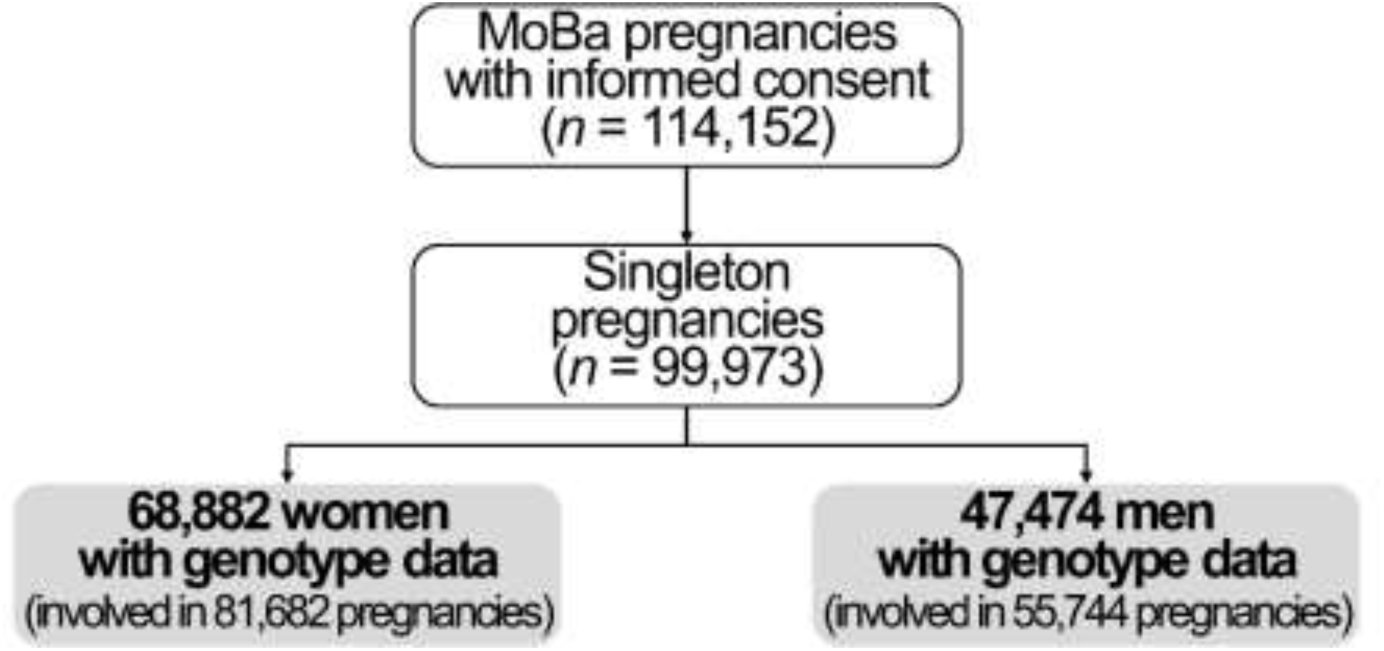
Study flow chart.

